# Development, implementation, and validation of an open-source Federated Learning platform to accelerate innovation and boost personalized medicine in rare and ultra-rare haematological diseases: an initiative by GenoMed4All Consortium

**DOI:** 10.1101/2025.08.07.25333044

**Authors:** Gianluca Asti, Patricia A. Apellaniz, Luciana Carota, Francesco Casadei, Davide Piscia, Mattia Delleani, Imanol Isasa, Daniela Martinez Duarte, Cesare Rollo, Catalina Gonzalez Martin, Borja Arroyo Galende, Nono Saha Cyrille Merleau, Michel Van Deventer, Betzabel Cajiao Garcia, Elisabetta Sauta, Marilena Bicchieri, Anna Rizzo, Nathan Lea, Diana Lopez, Alessia Campagna, Giulia Maggioni, Luca Lanino, Alessandro Buizza, Eleonora Iascone, Alessandro Bruseghini, Matteo Zampini, Alejandro Almodóvar, Silvia Uribe, Victor Savevski, Torsten Haferlach, Wolfgang Kern, Pierre Fenaux, Lin-Pierre Zhao, Mar Manu Pereira, Uwe Platzbecker, Maria Diez-Campelo, Anders Krogh, Raffaella Colombatti, Konstantinos Marias, Eduard J. Van Beers, Petros Kountouris, Elisa Rossi, Teresa Garcia Lezana, Carolina Terragna, Tiziana Sanavia, Piero Fariselli, Enrico Giampieri, Juan Parras, Santiago Zazo, Rudolf Mayer, Andoni Beristain, Saverio D’Amico, Matteo Giovanni Della Porta, Gastone Castellani, Federico Álvarez Garcia, GenoMed4All Consortium

## Abstract

**Background:** Rare haematological diseases (RHD) pose significant clinical challenges due to their heterogeneity, limited patient populations, and fragmented datasets. To overcome these limitations, improve access to, and use of real-world multimodal data for scientific and clinical purposes, the GenoMed4All Consortium developed an open-source Federated Learning (FL) platform. This platform enables collaborative, privacy-preserving AI model training without the need to centralize sensitive patient information.

**Methods:** The FL platform was deployed within EuroBloodNet, the European Reference Network for RHD, across multiple use cases, including myelodysplastic syndromes (MDS), acute myeloid leukemia (AML), chronic myelomonocytic leukemia (CMML), and multiple myeloma (MM). Multimodal datasets (including clinical, genomic information together with histopathological and radiological extracted features) were utilized. Predictive models (DeepSurv and SAVAE) and generative Artificial intelligence (AI) algorithms (CTGAN, Bayesian Networks, and VAE-BGM) were trained using a federated approach. A dedicated data harmonization pipeline based on the FHIR standard ensured consistency across participating centers.

**Findings:** Federated models achieved performance comparable to centralized approaches, with highest benefit for institutions with smaller datasets. The platform enabled integration of multimodal data demonstrating flexibility across diverse data types and clinical endpoints. The inclusion of multimodal information improved predictive accuracy over currently available prognostic schemes. Generative models successfully created synthetic datasets that preserved both clinical and statistical fidelity while ensuring patient privacy; this allows extraction of insights from real-world data that can be used beyond the boundaries of FL, as a source for accelerating the conduction of clinical trials. A preliminary implementation within the EuroBloodNet clinical network demonstrated feasibility for broader scale-up.

**Interpretation:** This study validates FL as a robust, privacy-compliant approach to enable AI-driven precision medicine in RHD. The platform facilitates real-world data integration and model scalability, providing a foundation for multicenter collaboration, regulatory-grade evidence generation, and innovative trial designs in rare diseases.

**Funding:** European Union’s Horizon 2020 research and innovation programme.

## Introduction

Haematological diseases comprise up to 100 rare and ultra-rare entities, including both neoplastic and non-neoplastic disorders. Within these, patient subgroups often experience poor outcomes and face unmet clinical needs.^1,2^

In many instances, rare haematological diseases (RHD) arise from gene mutations that may result from inherited predispositions, environmental exposures, or spontaneous errors during hematopoietic cell replication. The discovery of recurrent genetic lesions has improved personalized medicine.^3^ Despite these scientific advances, RHD continue to face significant challenges. The limited number of clinical trials and historically fragmented care infrastructures have hindered generation of high-level evidence and development of effective diagnostic and treatment guidelines^4,5^. As a result, patients often face diagnostic delays, restricted access to referral centers, reduced opportunities for trial participation, and suboptimal treatment – factors that collectively increase healthcare and social burdens of RHD.^6^

To address these gaps, in 2017 the European Commission established 24 European Reference Networks (ERN) aimed at improving care for patients with rare diseases. EuroBloodNET is the ERN dedicated to RHD and includes 90 referral centers across Europe.^7^ Development of infrastructures that can support and improve the collection and use of data in the health-care community represents a research priority for RHD. Currently, only 3% of healthcare data is used for research purposes due to strict privacy regulations, lack of harmonization, fragmented infrastructures, and the unstructured nature of much clinical information; these barriers are even greater in rare diseases.^8^ In response, EuroBloodNET endorsed the establishment of the GenoMed4All consortium, aimed at developing innovative technologies to improve personalized medicine for RHD by enhancing access to and integration of multimodal data.^9^

A key strategy of the GenoMed4All project was to build and validate a platform that facilitates multi-layer data access and integration, based on the principle of Federated Learning (FL). FL allows multiple centers to collaboratively train shared predictive models without centralizing raw data. In this architecture, each institution (“worker” node) trains algorithms locally on its own dataset and transmits model updates, rather than patient-level data, to a coordinating (“manager”) node for aggregation. This design minimizes privacy risks, aligns with data-protection regulations, and retains high fidelity to local data environments.^10–12^

The primary clinical objectives of the GenoMed4All FL platform for RHD were: i) increase access to and harmonization of healthcare data via a common data model (CDM); ii) compare the reliability of federated versus centralized modelling approach; iii) integrate multiple data layers to increase personalized prediction of clinically relevant endpoints; iv) demonstrate proof-of-concept to accelerate the conduction of clinical trials, by leveraging high-quality real-world data. Complementing these goals, key technological and dissemination objectives included: i) developing a fully open-source, scalable FL framework adaptable to different clinical domains and data modalities; ii) implementing and clinically validating the platform across EuroBloodNET referral centers. Through this integrated strategy, GenoMed4All aims to deliver actionable insights and equitable access to precision-medicine solutions for patients with RHD.

## Methods

### Study populations and procedures

The Ethics Committees of the project’s partners approved the study. Written informed consent was obtained from each participant. This study was registered at ClinicalTrials.gov (ClinicalTrials.gov Identifier:NCT04889729)

### Federated Learning Infrastructure

The development of the GenoMed4All platform was guided by four principles: (i) Interoperability – enabling seamless information sharing and collaboration across different systems; (ii) Open-source design – the platform was built entirely with open-source components; (iii) Scalability and flexibility – its modular architecture supports large-scale deployment and allows adaptation to evolving functional requirements; (iv) Standardization – internationally recognized standards were applied to ensure data harmonization. (**Figure 1**)

**Figure 1.**
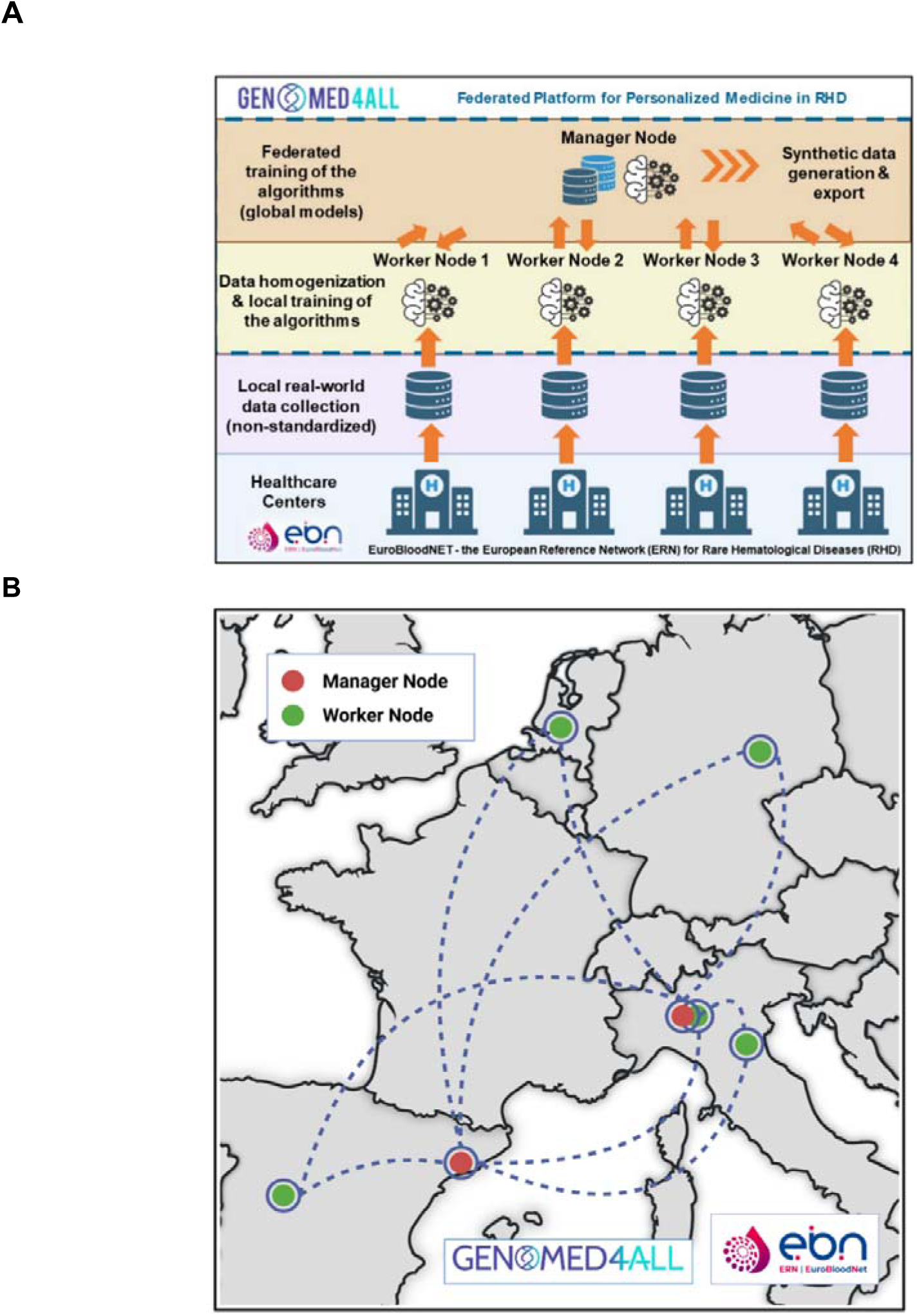
**A)** Genomed4All FL approach for AI Training. The centralized model is shared to each Worker Node for a training round on the local dataset. The coefficients are then collected and aggregated in the Manager Node, to create a more robust artificial intelligence. **B)** Current map of the Genomed4All platform deployment within EuroBloodNET clinical network. The Manager Nodes for oncological and non-oncological use cases are deployed at Humanitas AI Center, Milan Italy and at Vall d’Hebron Research Institute, Barcelona Spain, respectively. Five Worker Nodes are currently installed and operative at Universidad Politécnica de Madrid, Spain; University of Bologna, Italy; University Hospital of Leipzig, Germany; University Medical Center Utrecht, the Netherlands; and IRCCS Humanitas Research Hospital, Milan, Italy.

### Research Governance and GDPR Requirements

In developing its framework, GenoMed4All adopted the principles outlined in the General Data Protection Regulation (GDPR), specifically the ‘Data Protection by Design and by Default’ approach.^13^ A comprehensive Data Protection Impact Assessment (DPIA) was developed to ensure GDPR compliance and map data flows across the project. The DPIA enabled early integration of data protection, security, and management measures, while aligning partner consent requirements from data collection to infrastructure deployment.

### FL Platform Architecture

The GenoMed4All FL platform architecture comprises two main components. The first is the “Manager Node”, a central server responsible for orchestration, partner communication, data storage on PostgreSQL^14^, federated model management with MLflow^14^, and user interface. The second component consists of “Worker Nodes”, deployed at data provider sites to process local datasets and perform model training. These nodes utilize FastAPI^14^, PostgreSQL^14^, and SQLAlchemy,^14^ ensuring consistent functionality and ease of deployment across heterogeneous IT environments. Communication between nodes occurs via Application Programming Interfaces (API), with FastAPI endpoints^14^. Edge node registration with the central server involves secure public-key exchanges, authentication tokens, and health checks to maintain cluster stability. GenoMed4All adopted Flower^14,15^ an open-source, model-agnostic FL Python framework that is scalable, user-friendly, and fully compatible with Docker containers.

### Security Framework

Security of the FL platform was a primary focus since the early designing stages. Patients’ data remain under the control of custodians and are never directly shared. The platform uses Transport Layer Security (TLS) with symmetric and asymmetric encryption to protect data in transit and at rest. Role-Based Access Control (RBAC) enforces strict permissions at multiple levels. Communication is handled via FastAPI and HTTP(s) protocols, with authentication managed through OpenID Connect and Keycloak, and server-to-server authentication secured via asymmetric cryptography. Fine-grained and broad authorization policies ensure appropriate access to sensitive and general datasets.

### Data and Model Lifecycle Management

The FL platform links training plans to unique dataset identifiers. These datasets—typically in CSV format—are generated through an Extract, Transform, and Load (ETL) process structured according to a Common data Model (CDM). Metadata are attached to enhance traceability and usability; MLflow^14^ supports comprehensive model lifecycle management. Its key functionalities include: i) Experiment Tracking: Logging hyperparameters, metrics, artifacts, and versioning; ii) Model Packaging: Standardizing model storage and deployment; iii) Model Registry: Managing versioning, transitions, and annotations for reproducibility and traceability.

### Data Homogenization Platform

The Data Homogenization Platform is designed to standardize heterogeneous multi-omics datasets in FL environments according to HL7 FHIR^14^ (Fast Healthcare Interoperability Resources) standard. An ETL pipeline transforms source data into structured FHIR resources, following use-case-specific implementation guides. A feature extraction module converts these locally-saved FHIR resources into machine learning–ready (ML-ready) datasets, with associated quality reports. The extracted datasets are registered in the FL Platform Dataset Manager, which tracks their location for usage in local training. All platform operations are accessible through a web-based user interface or RESTful APIs for external integrations.

### Federated Training Execution

FL workflows automate task orchestration, error handling, and resource allocation. The system supports iterative training and aggregation stages, parallelization of jobs, and automatic termination of tasks exceeding time thresholds. Responsiveness is emphasized, allowing users to intervene in ongoing jobs when necessary.

### Generation of Synthetic Data into a federated environment

We explored the generation of Synthetic Data (SD) using innovative generative artificial intelligence (AI) applied within a FL framework. These models learn the joint statistical distribution of real data and produce new instances, statistically indistinguishable at the aggregate level while entirely artificial at the individual patient level. The resulting SD provide a secure alternative for public dissemination or collaborative development outside the protected federated environment, supporting reproducibility and transparency without compromising patient confidentiality.^16^ Here we report preliminary findings from experiments that generated SD within a simulated FL setting.

### Clinical use cases and data collection

Several RHDs clinical use cases were included, leveraging high-quality, multimodal healthcare data collected by the EuroBloodNET clinical network. The selection criteria for clinical use cases required that the haematological disease have a rare or ultra-rare prevalence in the general population and exhibit high clinical and molecular heterogeneity. This choice aimed to assess the performance of the FL platform under the most challenging clinical scenarios.

The study population consisted of several retrospective cohorts, including: 4,427 patients with myelodysplastic syndromes (MDS); 1,547 with acute myeloid leukemia (AML); 1,737 with chronic myelomonocytic leukemia (CMML); 1,005 with multiple myeloma (MM).

Inclusion criteria for all cases were: age ≥18 years; diagnosis of a haematological malignancy according to the WHO 2016 classification; and availability of data on demographics, clinical and hematologic parameters, chromosomal abnormalities, genomic features (i.e., gene mutations from targeted next-generation sequencing for MDS, AML, and CMML; and copy number variations for MM), treatment (including response assessment), and survival outcomes. In selected patient subgroups, additional multimodal data were integrated: i) multidimensional embedding vectors extracted from whole slide images (WSI) of bone marrow biopsies (Hematoxylin and Eosin staining) using the Prov-Gigapath foundation model^17^ (MDS, n=860); Positron Emission Tomography (PET scan)-derived radiomic features were added to MM patients (n=1,005). Radiomic features were extracted from PET images using the skeleton as Regions Of Interest (ROI). The skeleton segmentation was obtained from the CT image and feature extraction was carried out using the PyRadiomics Python package. (**Table 1**)

**Table 1.** Overview of the Experimental setup across different RHD. Federated models were trained on multi-institutional datasets for predicting overall survival or generating synthetic data (SD) across five use cases: MDS, AML, CMML, and MM. Models used clinical, genomic, and imaging features, with three to five Worker Nodes per experiment. Federated models included DeepSurv, SAVAE, and generative approaches (CTGAN, Bayesian Network, VAE-BGM).

| Use case (RHD) | N of patients | Type of data included in the model | Federated model trained | Clinical Endpoint | N of worker nodes |
| --- | --- | --- | --- | --- | --- |
| Myelodysplastic Syndromes (MDS) | 4427 | Age, sex, clinical, and haematological features, chromosomal abnormalities on hematopoietic progenitors; gene mutations by targeted NGS including 54 recurrently mutated genes; treatment (including assessment of clinical response); probability of survival | Deepsurv | Probability of overall survival | 5 |
| MDS | 860 | Age, sex, clinical, and haematological features, chromosomal abnormalities on hematopoietic progenitors; gene mutations by targeted NGS including 54 recurrently mutated genes; multidimensional embedding vectors extracted from of bone marrow biopsies Whole Slide Images (WSI); treatment (including assessment of clinical response); probability of survival | SAVAE | Probability of overall survival | 3 |
| Acute Myeloid Leukemias (AML) | 1547 | Age, sex, clinical, and haematological features, chromosomal abnormalities on hematopoietic progenitors; gene mutations by targeted NGS including 54 recurrently mutated genes; treatment (including assessment of clinical response); probability of survival | SAVAE | Probability of overall survival | 3 |
| Chronic Myelomonocytic Leukemias (CMML) | 1737 | Age, sex, clinical, and haematological features, chromosomal abnormalities on hematopoietic progenitors; gene mutations by targeted NGS including 54 recurrently mutated genes; treatment (including assessment of clinical response); probability of survival | Deepsurv | Probability of overall survival | 3 |
| Multiple Myeloma (MM) | 1005 | Age, sex, clinical, and haematological features, chromosomal abnormalities; copy number variations; PET extracted features; treatment (including assessment of clinical response); probability of survival | SAVAE | Probability of overall survival | 3 |
| MDS | 4427 | Age, sex, clinical, and haematological features, chromosomal abnormalities on hematopoietic progenitors; gene mutations by targeted NGS including 54 recurrently mutated genes; treatment (including assessment of clinical response); probability of survival | Conditional Tabular Generative Adversarial Network (CTGAN), a probabilistic Bayesian Network (BN), and a hybrid Variational Autoencoder with a Bayesian Gaussian Mixture Model (VAE-BGM). | Statistical fidelity and privacy preservability (compared with real data) | 3 |

Each dataset was randomly split into a training set (90%) and a test set (10%). The training data were further unevenly distributed across the Worker Nodes, using either a three-node configuration (data distribution: 60%, 30%, 10%) or a five-node configuration (data distribution: 45%, 20%, 15%, 10%, 10%). Two experimental scenarios were tested for each use case: i) best-case scenario: all Worker Nodes had complete datasets with 100% of the information; ii) worst-case scenario: nodes contained incomplete information or partial datasets, thus simulating a real-world context of fragmented and heterogeneous data across different institutions.

Clinical validation of the FL approach was performed by evaluating its ability to train federated prognostic models and to generate high-quality, privacy-preserving SD from Worker Nodes using dedicated algorithms.

### Federated Algorithms

The GenoMed4All platform employs the FedAvg algorithm to coordinate FL.^10^ The Manager Node initializes and orchestrates the training. At the start of each training round, a subset of available Worker Nodes is selected. Each selected node receives a copy of the current global model and performs local training using only its own dataset. Upon completion, the nodes transmit their updated model coefficients back to the Manager Node, which aggregates these parameters, weighting them by the number of patients at each institution. Weighted aggregation ensures proportional contribution, fostering fairness within the FL process. This cycle repeats over a predefined number of rounds, converging towards a robust global model that benefits from the diversity of distributed datasets while maintaining data privacy.

Clinical analyses were conducted using an FL framework combining two flexible, state-of-the-art models for survival analysis: DeepSurv^18^ and the Survival Analysis Variational Autoencoder (SAVAE).^19^ DeepSurv extends the Cox proportional hazards model with deep neural networks to capture complex features relationships in high-dimensional data.^25^ SAVAE is a Bayesian variational autoencoder and by optimizing the Evidence Lower Bound (ELBO) with the reparameterization trick, it is capable of effectively handling censoring and improving survival prediction accuracy.^19^

We aimed to train generative AI models using an FL strategy for the creation of high-quality privacy-preserving SD. To evaluate various models and training strategies, the dataset of 4,427 MDS patients was partitioned across three nodes. Three generative models were tested: i) a deep-learning-based Conditional Tabular Generative Adversarial Network (CTGAN)^16^ ii) a probabilistic Bayesian Network (BN);^20^ iii) a hybrid Variational Autoencoder with Bayesian Gaussian Mixture Model (VAE-BGM).^21^ These models were trained using different FL strategies, including FedAvg,^10^ a two-step BN training procedure (structure learning followed by parameter learning), and a SD sharing approach.

To evaluate and compare the performance of survival prediction models, we used Harrell’s Concordance Index (CI), a standard metric in survival analysis.

The validation of SD generation was conducted using the SAFE framework, which evaluates synthetic datasets across three key dimensions—statistical fidelity, clinical utility, and privacy—in comparison with real-world data. Statistical fidelity was assessed through distributional comparisons and formal statistical tests; clinical utility was evaluated by examining genomic pairwise associations, correlation structures, and survival analyses; and privacy was quantified using metrics such as the Nearest Neighbor Distance Ratio (NNDR).^22^

## Results

### Cinical Validation of FL platform across different Use Cases

#### Predictive AI Training in Federated Learning

The FL platform delivered consistent, high-quality training/validation performance across all case studies and data modalities, as summarized in **Table 2**.

**Table 2.** Performances of survival models across different experimental setup. The table shows the CI performances in the four clinical use cases: CMML, MDS (with and without medical images embeddings), AML, and MM. Each cell also highlights the percentage of data available in each Worker Node or setup. The results are presented under various data-sharing scenarios: centralized (Full Dataset), FL across five nodes, three nodes, and isolated nodes (both the largest and smallest). For each use case, results are reported for both a Best Case (complete dataset) and a Worst Case (limited subset of features) scenario. Additionally, a column shows the CI of conventional prognostic scores applied to the study populations, as a reference. Across all conditions, federated models consistently outperform isolated training, and in many cases their performances overcome those of conventional scores, highlighting the added prognostic value of molecular and multimodal data integration.

| Use Case | Scenario | Full Dataset | Node 1 with FL | Node 2 with FL | Node 3 with FL | Node 4 with FL | Node 5 with FL | Isolated Biggest Node | Isolated Smallest Node | Prognostic Models currently adopted in clinical practice |
| --- | --- | --- | --- | --- | --- | --- | --- | --- | --- | --- |
| CMML (n=1,737) | Best Case | 0.69 (100%) | 0.712 (60%) | 0.692 (30%) | 0.688 (10%) | .. | .. | 0.705 (60%) | 0.636 (10%) | CPSS-mol score *: 0.62 |
| CMML (n=1,737) | Worst Case | 0.69 (100%) | 0.712 (60%) | 0.692 (30%) | 0.691 (10%) | .. | .. | 0.705 (60%) | 0.613 (10%) |  |
| MDS (n=4427) | Best Case | 0.756 (100%) | 0.744 (45%) | 0.749 (20%) | 0.747 (15%) | 0.756 (10%) | 0.746 (10%) | 0.744 (45%) | 0.733 (10%) | IPSS-M** score: 0.70 |
| MDS (n=4427) | Worst Case | 0.752 (100%) | 0.743 (45%) | 0.746 (20%) | 0.743 (15%) | 0.751 (10%) | 0.745 (10%) | 0.737 (45%) | 0.725 (10%) |  |
| AML (n=1547) | Best Case | 0.67 (100%) | 0.66 (60%) | 0.66 (30%) | 0.65 (10%) | .. | .. | 0.67 (60%) | 0.62 (10%) | ELN prognostic stratification***: 0.64 |
| AML (n=1547) | Worst Case | 0.66 (100%) | 0.64 (60%) | 0.64 (30%) | 0.64 (10%) | .. | .. | 0.67 (60%) | 0.64 (10%) |  |
| MDS with WSI embedding (n=860) | Best Case | 0.797 (100%) | 0.767 (60%) | 0.785 (30%) | 0.785 (10%) | .. | .. | 0.752 (60%) | 0.753 (10%) | IPSS-M score: ** 0.70 |
| MDS with WSI embedding (n=860) | Worst Case | 0.802 (100%) | 0.816 (60%) | 0.710 (30%) | 0.778 (10%) | .. | .. | 0.69 (60%) | 0.649 (10%) |  |
| MM (n=1005) | Best Case | 0.802 (100%) | 0.79 (60%) | 0.78 (30%) | 0.80 (10%) | .. | .. | 0.78 (60%) | 0.71 (10%) | Revised MM International Staging System (R-ISS)****: 0.61 |
| MM (n=1005) | Worst Case | 0.76 (100%) | 0.74 (60%) | 0.77 (30%) | 0.77 (10%) | .. | .. | 0.73 (60%) | 0.66 (10%) |  |

\* Elena C, Galli A, Such E, et al. Integrating clinical features and genetic lesions in the risk assessment of patients with chronic myelomonocytic leukemia. Blood 2016; 128 (10): 1408–17.
\*\* Bernard E, Tuechler H, Greenberg PL, et al. Molecular International Prognostic Scoring System for Myelodysplastic Syndromes. NEJM Evid. 2022; 1(7):EVIDoa2200008.
\*\*\* Döhner H, Wei AH, Appelbaum FR, et al. Diagnosis and management of AML in adults: 2022 recommendations from an international expert panel on behalf of the ELN. Blood. 2022;140(12):1345-1377
\*\*\*\* Rajkumar SV, Dimopoulos MA, Palumbo A, et al. International Myeloma Working Group updated criteria for the diagnosis of multiple myeloma. Lancet Oncol. 2014; 15(12):e538-48.

As a representative example, we present the clinical validation results for CMML —an ultra-rare hematologic malignancy with marked clinical and molecular heterogeneity. Within the FL environment, the DeepSurv model^18^ achieved overall survival prediction performance comparable to that obtained using traditional centralized training methods. Notably, all “Worker Nodes” benefited from the federated approach, regardless of their local dataset size. (**Table 2 and Figure 2**) On the other hand, models trained locally on smaller datasets showed lower predictive performance, directly proportional to the number of patients available in the Best-Case Scenario (i.e., with complete access to all patient-level data). A similar pattern emerged in the Worst-Case Scenario, where the limited per-patient information negatively affected outcomes when training was performed in isolation. In both scenarios, however, FL enabled a more stable and robust training process, resulting in improved model performance—an effect that was particularly pronounced for nodes with smaller datasets or more limited data. (**Table 2**) These findings demonstrate that institutions with modest patient cohorts benefit substantially from the collaborative FL setting, especially when paired with nodes contributing larger datasets.

**Figure 2:**
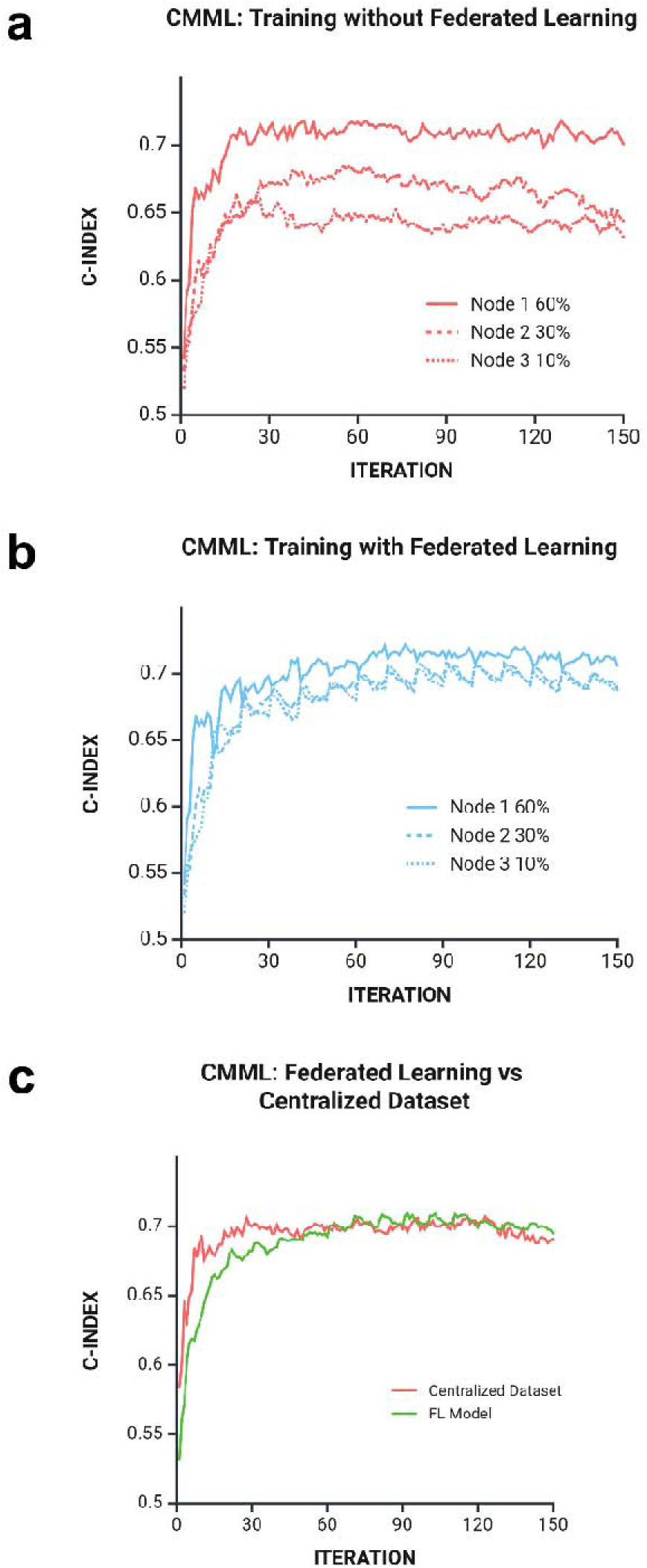
**A)** Training of DeepSurv model without FL in 1,737 patients with Chronic Myelomonocytic Leukemia (CMML). The institutions with smaller datasets (i.e., Node 2 and 3 with 10% and 30% of data of the whole patient population, respectively) have worse performances **B)** Training of DeepSurv model within GenoMed4all FL platform environment. All the institutions benefit from the FL environment in terms of model performances, especially the ones with smaller datasets (Node 2 and 3) **C)** Training performances of a Centralized vs, Federated DeppSurv models, showing comparable performance

Comparable results were observed across the other use cases, where two federated models—SAVAE and DeepSurv^18,19^—were trained to predict patient outcomes. (**Table 2**)

In an FL environment, the number of participating nodes and the degree of data heterogeneity are critical determinants of model quality: the inclusion of more institutions increases data diversity, thereby producing more robust and generalizable models. This was confirmed in experiments using the MDS dataset distributed across five Worker Nodes. The DeepSurv model^18^ achieved high CI values overall, though some performance variability was noted at nodes with smaller patient cohorts. These findings align with the results of other use cases, underscoring that FL delivers stable training performance comparable to centralized AI models, with particularly strong benefits for institutions contributing limited data—a crucial factor in rare disease research. (**Table 2**)

Moreover, the platform demonstrated solid performance in use cases involving MDS and MM datasets enriched with histopathological and radiological embeddings. These results confirm the feasibility of federating not only structured, human-interpretable features derived from medical images but also complex representations such as embeddings generated by vision transformer models. Interestingly, incorporating extensive molecular inputs and/or embeddings from additional data layers within a multi-omics evaluation framework increased the model’s predictive performance beyond the thresholds of currently available scoring systems. (**Table 2**)

Overall, the Genomed4All FL platform proved to be flexible, scalable, and effective, supporting a wide range of datasets, predictive tasks, and ML models across multiple clinical scenarios involving RHDs.

### Synthetic Data Generation in a FL environment

Three different generative algorithms were evaluated for SD creation: CTGAN, BN, and VAE-BGM.^16,20,21^ The effectiveness of the FL approach in generating SD was assessed by comparing its results with those obtained from a fully centralized dataset. Model performance was analyzed under two scenarios: i) independent model training on each node and ii) training on a fully centralized dataset, which served as an “upper-bound” benchmark representing the ideal condition. When applicable, SD were also generated at each FL round to capture the central node’s learning progression throughout the training process. (**Table 2** and **Figure 3**)

**Figure 3.**
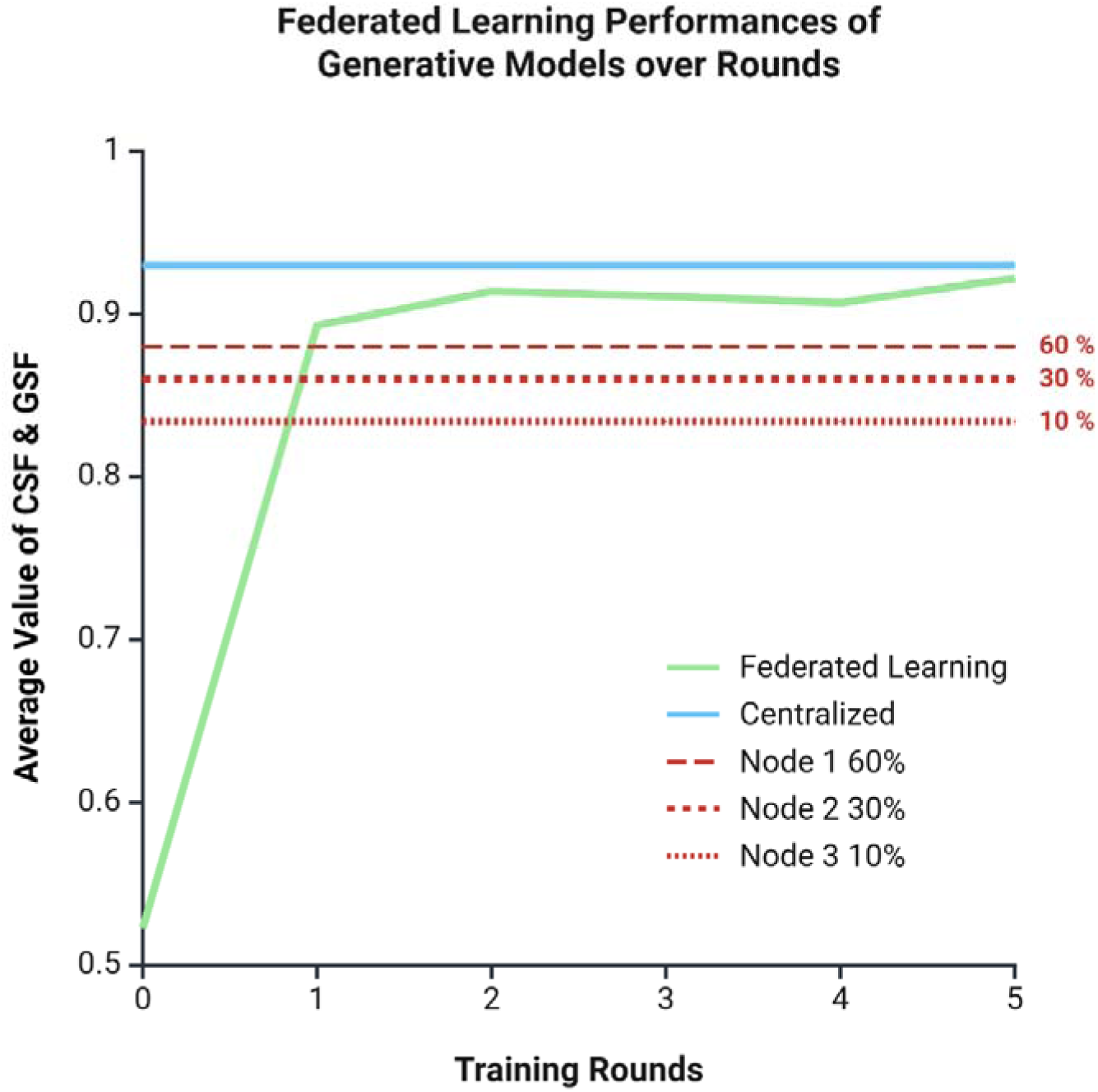
Validation performance of the VAE-BGM model for generating Synthetic Data (SD) trained under different paradigms across five FL rounds. The green line represents the average performance of Clinical Similarity Fidelity (CSF) and Genetic Statistical Fidelity (GSF) using the FL approach, showing a strong improvement from round zero to round one, followed by a gradual increase and convergence toward the performance of the centralized dataset. The centralized training, depicted by the solid blue line, maintains consistently high validation performance. The dashed and dotted red lines represent the performance of individual local nodes (Node 1, Node 2, and Node 3) when trained independently. These local models show consistently lower performance compared to both the federated and centralized approaches, highlighting the benefit of collaborative learning. Overall, the figure demonstrates that FL rapidly closes the performance gap with centralized training and significantly outperforms isolated local training, highlighting its effectiveness in distributed generative modelling.

The results demonstrate that the FL approach achieves performance comparable to the upper-bound scenario while outperforming models trained in isolation on individual nodes (**Figure 3**). Metrics such as Clinical Similarity Fidelity (CSF), Genetic Statistical Fidelity (GSF), and NNDR^22^—particularly those obtained using the VAE-BGM model^21^—showed that SD generated via FL closely matched those produced by centralized training, confirming the effectiveness of FL in producing high-quality synthetic patients.

From a clinical perspective, analyses of genomic abnormality frequencies, gene lesion co-occurrence, and mutual exclusivity, as well as predictions of overall survival probabilities, further validated these findings by demonstrating strong consistency between synthetic and real datasets across multiple evaluation metrics.

The central node exhibited progressive improvements in CSF and GSF scores over successive training rounds, underscoring the role of FL in improving data synthesis over time. (**Figure 3**) Finally, comparisons of overall survival analyses derived from real-world data, single-node models, and the FL framework revealed that p-values differed significantly when data were generated at individual nodes but not when using the FL approach. This finding further confirms that FL preserves the statistical integrity and robustness of the generated data, enabling consistent inference across distributed sources without compromising result reliability.

### Implementation of Genomed4ALL FL platform within EuroBloodNet clinical network

A specific implementation strategy was developed to facilitate the deployment of the FL platform within the EuroBloodNET network of healthcare providers. This initiative was formally presented in collaboration with the European Hematology Association (EHA) at the EHA Annual Meeting held in Madrid in June 2024. Two Manager Nodes were established: one at IRCCS Humanitas Research Hospital (Milan, Italy) for oncological rare haematological diseases (RHD) and another at Vall d’Hebron Hospital (Barcelona, Spain) for non-oncological RHD (in this context, GenoMed4All created the first harmonized database in the EU for Sickle Cell Disease and successfully conducted a pilot federated analysis across three centers). To date, five centers completed the installation and the validation of the FL platform Worker Nodes: Universidad Politécnica de Madrid, Spain; University of Bologna, Italy; University Hospital of Leipzig, Germany; University Medical Center Utrecht, the Netherlands; and IRCCS Humanitas Research Hospital, Milan, Italy. (**Figure 1**) We anticipate that more than 50 centers could be integrated within the next 24 months.

## Discussion

In this study, conducted by the GenoMed4All consortium and EuroBloodNet, we present the first implementation of a FL platform within a clinical reference network for RHD. This initiative aims to advance personalized medicine for these conditions characterized by significant unmet needs.

One of the main barriers to innovation in rare diseases is the limited availability and accessibility of high-quality real-world data. ^4,8^ FL was developed to overcome the constraints of centralized learning, where data from multiple sites are transferred to a central repository. ^10–12^ Centralized models face challenges related to privacy, data ownership, intellectual property and infrastructure costs. In contrast, FL enables algorithm training by sharing only model parameters, with all data remaining locally stored. This enhances privacy protection and removes the need for direct data sharing, while maintaining performance comparable to centralized models. FL thus enables broader collaboration among healthcare providers, increasing the diversity and amount of data available for training ML models while distributing the computational and storage burden/costs across the network.^10–12^

Aligned with these principles, the GenoMed4All FL platform demonstrated the ability to use multimodal RHD data from multiple centers to train predictive algorithms without formal data exchange, ensuring privacy compliance. The platform was developed entirely with open-source tools, supporting customization, scalability, and cross-domain adaptability—essential features for rare diseases, which involve small patient populations with unique clinical and therapeutic profiles.^1–4^The platform also includes a homogenization layer to map heterogeneous datasets into a Common Data Model, improving interoperability, facilitating large-scale analysis, and reducing management costs.^21^ Performance of the FL system was comparable to centralized architectures and effectively supported smaller centers by leveraging data-rich partners within the network.

Clinical validation across multiple centers and use cases (also including ultra rare conditions) confirmed the platform’s versatility, handling diverse data types including omics and AI-extracted features from medical images.^17^ In RHD, high-dimensional data such as histopathology and radiology images are increasingly relevant for diagnosis and prognosis but remain underutilized.^17^ Recent integration of AI in image analysis allows detection of subtle patterns and extraction of quantitative features to support clinical decisions.^23^ Federated models combining granular genomic and image-derived data significantly increased the predictive accuracy of conventional clinical predictions, outperforming the currently available tools

Furthermore, this study provides proof of concept for using FL to support innovative trial designs in RHD, in which randomized controlled trials are often unfeasible due to small patient populations.^4,5^ In this context, real-world data can be used to generate synthetic control arms, reducing costs and trial durations while ensuring investigational treatment access to all participants.^16,24,25^ Regulatory bodies such as the FDA and EMA have recently endorsed the use of real-world data, registries, and synthetic datasets to support drug approval and accelerate innovation.^26–28^ In this context, we explored the use of generative AI algorithms within a federated setting to produce synthetic patients based on real-world data in RHD. Synthetic data generation technologies accurately replicate the statistical properties and complex interactions/associations found in real data, address limitations of conventional anonymization processes, and enable effective data augmentation.^16,24,25^ Importantly, since are not real data, that they can be easily accessed and shared. Our preliminary results showed that synthetic data generation within a federated environment was efficient, privacy-preserving and clinically reliable, validating the concept that FL combined with generative AI can improve access to high-quality real-world data. Two EU-funded projects—SYNTHEMA and SYNTHIA—are currently expanding these efforts, developing platforms for synthetic data generation in hematology using FL.^29,30^

An implementation effort is ongoing for the broader deployment of the FL platform. The goal is to establish a federated architecture within a range of EuroBloodNet centers, to achieve the following aims: i) improving access to clinical and omics data to foster novel insights and knowledge in RHD; ii) supporting prospective initiatives to validate predictive models and enable implementation of next-generation predictive tools and clinical guidelines; iii) Supporting the EMA’s Qualification of Registry Initiative, enabling certification of a scalable amount of real-world data from EuroBloodNet for regulatory use, and leveraging synthetic data to accelerate clinical research; and iv) building trust in AI-based tools through educational programs for clinicians and patients, addressing key barriers to the adoption of digital technologies in medicine.

## Data Availability

All data produced in the present study are available upon reasonable request to the authors

## References

1. Nguengang Wakap S, Lambert DM, Olry A, et al. Estimating cumulative point prevalence of rare diseases: analysis of the Orphanet database. Eur J Hum Genet. 2020; 28:165–173.

2. The Lancet Global Health. The landscape for rare diseases in 2024. Lancet Glob Health. 2024 Mar;12(3):e341.

3. Boycott KM, Hartley T, Biesecker LG, et al. A diagnosis for all rare genetic diseases: the horizon and the next frontiers. Cell. 2019; 177:32–37

4. Taruscio D, Gahl WA. Rare diseases: challenges and opportunities for research and public health. Nat Rev Dis Primers. 2024 Feb 29;10(1):13.

5. United Nations. Addressing the challenges of persons living with a rare disease and their families. https://documents.un.org/doc/undoc/gen/n23/420/62/pdf/n2342062.pdf Date: 2021.

6. Burns R, Leal J, Sullivan R, Luengo-Fernandez R. Economic burden of malignant blood disorders across Europe: A population-based cost analysis. Lancet Haematol. 2016 Aug;3(8):e362–70.

7. EuroBloodNet. EuroBloodNet. [visited 23 july 2025]. ERN-EuroBloodNet, the European Reference Network on Rare Hematological Diseases. Available at: https://eurobloodnet.eu/

8. Legido-Quigley C, Wewer Albrechtsen NJ, Bæk Blond M, et al. Data sharing restrictions are hampering precision health in the European Union. Nat Med 31, 360–361 (2025).

9. GenoMed4All. [visited 23 july 2025]. GenoMed4All - Genomics For Next Generation Healthcare. Available at: https://genomed4all.eu/

10. McMahan HB, Moore E, Ramage D, Hampson S, Arcas, BA y. Communication-Efficient Learning of Deep Networks from Decentralized Data. arXiv; 2023. Available at: http://arxiv.org/abs/1602.05629

11. Sheller MJ, Edwards B, Reina GA, et al. Federated learning in medicine: facilitating multi-institutional collaborations without sharing patient data. Sci Rep. 2020;10(1):12598.

12. Rieke N, Hancox J, Li W, et al. The future of digital health with federated learning. npj Digit Med. 2020;3(1):1–7.

13. Regulation (EU) 2016/679 of the European Parliament and of the Council of 27 April 2016 on the protection of natural persons with regard to the processing of personal data and on the free movement of such data, and repealing Directive 95/46/EC (General Data Protection Regulation) (Text with EEA relevance). 2016. Available at: http://data.europa.eu/eli/reg/2016/679/2016-05-04/eng

14. Technical References: PostgreSQL. 2025 [visited 23 july 2025]. PostgreSQL. Available at: https://www.postgresql.org/; MLflow. [visited 23 july 2025]. Available at: http://mlflow.org/; FastAPI. [visited 23 july 2025]. Available at: https://fastapi.tiangolo.com/; SQLAlchemy. [visited 23 july 2025]. Available at: https://www.sqlalchemy.org; MongoDB. [visited 23 july 2025]. MongoDB: The World’s Leading Modern Database. Available at: https://www.mongodb.com/; Authors TF. Flower: A Friendly Federated AI Framework. [visited 23 july 2025]. Available at: https://flower.ai/; Keycloak. [visited 23 july 2025]. Available at: https://www.keycloak.org/; FHIR v5.0.0. [visited 23 july 2025]. Available at: https://www.hl7.org/fhir/

15. Beutel DJ, Topal T, Mathur A, et al. Flower: A Friendly Federated Learning Research Framework. arXiv; 2022. Available at: http://arxiv.org/abs/2007.14390

16. D’Amico S, Dall’Olio D, Sala C, et al. Synthetic Data Generation by Artificial Intelligence to Accelerate Research and Precision Medicine in Hematology. JCO Clin Cancer Inform. 2023 Jun;7:e2300021.

17. Xu H, Usuyama N, Bagga J, et al. A whole-slide foundation model for digital pathology from real-world data. Nature. 2024; 630:181–188

18. Katzman J, Shaham U, Bates J, Cloninger, A, Jiang, T, Kluger, Y. DeepSurv: Personalized Treatment Recommender System Using A Cox Proportional Hazards Deep Neural Network. BMC Med Res Methodol. 2018;18(1):24.

19. Apellániz PA, Parras J, Zazo S. SAVAE: Leveraging the variational Bayes autoencoder for survival analysis. arXiv; 2023. Available at: http://arxiv.org/abs/2312.14651

20. Kaur D, Sobiesk M, Patil S, et al. Application of Bayesian networks to generate synthetic health data. J Am Med Inform Assoc. 2020;28(4):801–11.

21. Apellániz PA, Parras J, Zazo S. An improved tabular data generator with VAE-GMM integration. arXiv; 2024. Available at: http://arxiv.org/abs/2404.08434

22. Zazzetti E, Jacobs F, D’Amico S, et al. Longitudinal Synthetic Data Generation by Artificial Intelligence to Accelerate Clinical and Translational Research in Breast Cancer. JCO CCI 2025 in press

23. Barragán-Montero A, Javaid U, Valdés G, et al. Artificial intelligence and machine learning for medical imaging: A technology review. Phys Med. 2021;83:242–256. doi: 10.1016/j.ejmp.2021.04.016. Epub 2021 May 9. PMID: 33979715; PMCID: PMC8184621.

24. Thorlund K, Dron L, Park JJH, Mills EJ. Synthetic and external controls in clinical trials - a primer for researchers. Clin Epidemiol. 2020;12:457–467.

25. Serrano C, Rothschild S, Villacampa G, et al. Rethinking placebos: embracing synthetic control arms in clinical trials for rare tumors. Nat Med. 2023;29(11):2689–269

26. EMA Cross-Committee Task Force on Patient Registries. Discussion paper: use of patient disease registries for regulatory purposes – methodological and operational considerations. Available at: https://www.ema.europa.eu/en/human-regulatory/post-authorisation/patient-registries#-useof-patient-disease-registries-for-regulatorypurposes-(open-consultation)-section

27. U.S. Food and Drug Administration. FDA Guidance for Industry: Submitting Documents Using Real-World Data and Real-World Evidence to FDA for Drugs and Biologics. May 2019. Available at: https://www.fda.gov/media/124795/download

28. Yoshino T, Shi Q, Misumi T, et al. A synthetic control arm for refractory metastatic colorectal cancer: the no placebo initiative. Nat Med. 2023;29(10):2389–2390.

29. SYNTHEMA | Synthetic Haematological Data – SYNTHEMA | Synthetic Haematological Data [Internet]. [visited 23 july 2025]. Avaliable at: https://synthema.eu/

30. Synthia [Internet]. [visited 23 july 2025]. Avaliable at: https://www.ihi-synthia.eu/

