## Supplementary Appendix for "Development, implementation, and validation of an open-source Federated Learning platform to accelerate innovation and boost personalized medicine in rare and ultra-rare haematological diseases: an initiative by GenoMed4All Consortium"

### Supplementary

### Supplementary Information 1| FedAvg

The FedAvg algorithm is initialized on the central Manager Node. At each step $t$ a subset $K$ of available Worker Nodes is selected for training. Each one of them downloads the centralized model and performs a local round of training: at the end of it, the model's weights are shared back with the Manager Node. The Worker Nodes' coefficients are aggregated using the following formula, where their contribution to the centralized model is weighted based on the number of samples available in each single node.

$$w^{(t+1)} = \sum_{k=1}^{K} \frac{n_k}{n} w_k^{(t+1)}$$

where:

- $K$ : number of participating clients
- $n = \sum_{k=1}^{K} n_k$ : total data points from all participating clients
- $w_k^{(t+1)}$ : the model weights trained locally on client $k$

Nodes with a higher number of patients will make a larger contribution than institutions with a lower number of samples. Once the centralized model is updated in this way, the process is repeated for the number of steps $T$ required by the user.
Although this privacy-preserving algorithm is sensitive to high data heterogeneity across institutions, it serves as a strong and scalable benchmark for future versions of the Federated Learning (FL) platform that will incorporate more advanced aggregation techniques such as FedProx or the SIGMA approach introduced by Hassan et al.

### Supplementary Information 2| DeepSurv and SAVAE Models for patients' prognosis

#### DEEPSURV MODEL

The DeepSurv model is a deep learning-based evolution of the Cox proportional hazards for survival analysis. The AI is composed of a neural network architecture that analyzes the complex and non-linear relationship between input features and the risk of an event occurring (e.g., death, relapse). The last layer of the architecture returns a single value of the log-risk function, similar to the linear predictor in the Cox model.

DeepSurv is a deep feedforward neural network designed to predict the effect of a patient's covariates on their hazard rate, parameterized by the network weights $\theta$. The input to the network is the patient's baseline data $x$. The hidden layers consist of fully connected layers followed by dropout for regularization. The network output, $\hat{h}_\theta(x)$, is a single node with a linear activation function that estimates the log-risk function of the Cox proportional hazards model.

The network is trained by minimizing the average negative log partial likelihood with $L_2$ regularization:

$$\ell(\theta) = -\frac{1}{N_{E=1}} \sum_{i:E_i=1} (\hat{h}_\theta(x_i) - \log \sum_{j\in\mathcal{R}(T_i)} e^{\hat{h}_\theta(x_j)}) + \lambda||\theta||_2^2$$

where $N_{E=1}$ is the number of patients with an observed event, $\lambda$ is the regularization parameter, $T_i$ is the observed time for patient $i$, and $\mathcal{R}(T_i)$ denotes the risk set at time $T_i$.

Optimization of the network parameters is performed using gradient descent, specifically the Adaptive Moment Estimation (Adam) optimizer with Nesterov momentum. Additional modern deep learning techniques are applied to enhance training stability and performance, including input standardization, the use of Scaled Exponential Linear Units (SELU) as the activation function, and learning rate scheduling. Hyperparameters are tuned via random hyperparameter search.

The neural network component of the model improves its capabilities to handle high-dimensional data and understand complex non-linear relationships between covariates and survival outcomes.

**SAVAE MODEL**

Besides the DeepSurv model, another AI was trained in these use cases: the Survival Analysis Variational Autoencoder (SAVAE) model.
This approach consists of a Bayesian VAE framework, where a latent variable is sampled from a prior distribution and used to generate data via a conditional generative model. Optimization is required to approximate the true posterior distribution, leveraging deep neural networks and the reparameterization trick to compute the Evidence Lower Bound (ELBO) efficiently.

The advantage of this model is extending the traditional VAE framework to model covariates and time-to-event outcomes while explicitly handling censoring jointly..

SAVAE models a dataset consisting of covariates $x$, observed times $t$, and event indicators $\delta$, by introducing a latent variable $z$ that captures the underlying factors governing survival outcomes. The generative process first samples $z \sim p(z)$, then reconstructs covariates $x \sim p_\theta(x|z)$ and models the time-to-event distribution $p_\theta(t|z)$.

The training objective is to maximize the ELBO on the marginal log-likelihood of the observed data. However, optimizing the ELBO with respect to the parameters $\phi$ of the approximate posterior $q_\phi(z|x)$ is challenging due to the presence of $\phi$ inside the expectation. To overcome this, SAVAE employs the reparameterization trick, which allows the gradient of the ELBO to be estimated through a differentiable transformation of an auxiliary noise variable.

Instead of sampling $z \sim q_\phi(z|x)$ directly, SAVAE samples $\epsilon \sim p(\epsilon)$, typically a standard normal distribution, and computes

$$z = g_\phi(x, \epsilon)$$

where $g_\phi$ is a deterministic function parameterized by the encoder network. Under this reparameterization, the ELBO can be approximated as:

$$\hat{\mathcal{L}}(x, \theta, \phi) = \frac{1}{N} \sum_{i=1}^{N} \left( -\mathrm{KL}(q_\phi(z|x_i) \| p(z)) + \log p_\theta(x_i | g_\phi(x_i, \epsilon_i)) \right)$$

where $\theta$ denotes the parameters of the decoder network, and the Kullback-Leibler (KL) divergence term regularizes the approximate posterior towards the prior.

This formulation enables the use of standard gradient-based optimization techniques, with gradients computed efficiently via backpropagation, and implemented in modern deep learning frameworks. The VAE architecture used by SAVAE resembles an autoencoder, where the encoder network $q_\phi(z|x)$ outputs the parameters of the latent distribution, and the decoder $p_\theta(x|z)$ reconstructs the input data. The latent space is structured such that $z$ captures both the features relevant for reconstructing the covariates and the factors influencing survival times.

For the survival modeling component, the event times $t$ are generated by a survival distribution (e.g., Weibull or exponential) parameterized by $z$, allowing the model to predict individual survival curves and handle right-censoring directly in the likelihood formulation.

### Supplementary Information 3| Synthetic Data Generation

#### Conditional Tabular GAN in Federated Learning

*Methodology:* Generative Adversarial Networks (GANs) are a class of deep learning (DL) architectures consisting of two neural networks: a generator, and a discriminator. These networks are trained simultaneously in an adversarial process, where the generator learns to approximate the original data distribution, while the discriminator attempts to distinguish between real samples and those generated by the generator(1).

The Conditional Tabular GAN (CTGAN) is an evolution of the original GAN framework designed for synthetic tabular data generation. It incorporates mode-specific normalization techniques and allows for conditional data generation (e.g., generating samples for individuals aged 20 to 30 years). As reported by Kiran et al., CTGAN is among the most effective models for synthesizing realistic tabular data(2), which is the main reason for considering it as a de facto baseline. Therefore, a study was conducted to evaluate whether its performance remains consistent when deployed in a FL setting. To do that, an initial baseline model was trained with the completeness of the data, while node-specific baselines were then created using the data partitions. The federated model was evaluated using checkpoints every 50 federation rounds.

*Data Preprocessing:* For the input dataset, categorical variables were one-hot encoded by default to ensure a consistent mapping across nodes. For numerical variables, data distributions were approximated using a Gaussian mixture model with multiple components, aiming to better represent multimodal distributions and enhance model convergence, and stability. This transformation was restricted to a maximum of four components and a weight threshold of 0.005 to maintain consistency in the shape of the weight matrices in a federated framework. All transformations were performed on the same scheme to avoid aggregation problems between nodes.

*Experimental Setting:* The implementation of CTGAN from the Synthetic Data Vault (SDV) library(3) was used as the baseline model, with modifications to accommodate the requirements of FL using the Flower Framework(4). The model's hyperparameters were configured with a batch size of 500 samples and a pac value of 10 samples, while the layer sizes, learning rates, and weight decay parameters were retained at their default settings. The number of federation rounds was set to 500, as empirical trials showed no improvements with additional training. Additionally, local models at each node were trained for 30 epochs, since in this case, there was no significant improvement detected in empirical analyses using higher quantities.

#### Variational Autoencoder with Bayesian Gaussian Mixture

*Methodology:* This study implemented and trained the Variational Autoencoder with a Bayesian Gaussian Mixture (VAE-BGM) model as proposed in (5) for synthetic data generation (SDG). The model extended the VAE framework by incorporating a Bayesian Gaussian Mixture (BGM) model in the latent space post-training, enhancing its ability to capture complex and multimodal data distributions. This approach is particularly effective for Electronic Health Records (EHRs) and clinical tabular datasets, where variables exhibit intricate dependencies and heterogeneity.

Unlike GANs(1), which are prone to mode collapse, VAEs provide a structured approach to learning latent representations. The standard VAE (6) assumes a Gaussian prior, which may not be sufficient for capturing real-world medical data complexities. The VAE-BGM method addresses this by applying a Gaussian mixture model (GMM) to the latent representations, refining the learned distribution, and preserving distinct patient subpopulations.

The VAE-BGM architecture consists of two main components:

1. Variational Autoencoder: Learns a probabilistic mapping between the input and a lower-dimensional latent space, where the encoder maps the input $x$ to a latent representation $z$, modeled as a multivariate Gaussian distribution. The decoder reconstructs the original input from $z$, ensuring synthetic samples maintain statistical fidelity. The ELBO loss function optimizes reconstruction while regularizing the latent space with a KL divergence term.

2. Bayesian Gaussian Mixture: After training, a BGM is fitted to the learned latent space, modeling it as a mixture of Gaussians to provide a more structured representation. This step does not modify VAE training but enhances its ability to capture multimodal distributions.

The VAE-BGM approach improves synthetic patient data generation by capturing complex subpopulations, reducing overfitting, and ensuring higher-quality synthetic samples. Additionally, it enhances privacy preservation, as synthetic data are not direct reconstructions of real patient records. This method offers a scalable and robust solution for generating high-fidelity synthetic datasets, supporting privacy-preserving medical research, and secure data-sharing.

*Data Preprocessing:* The preprocessing pipeline involved distribution-based normalization, survival data correction, and encoding of categorical and numerical variables to ensure consistency across federated nodes. Each feature was transformed based on its detected distribution: Gaussian features were standardized, log-normal features were log-transformed, and categorical variables were assigned integer-based representations. Survival data underwent preprocessing to remove inconsistencies, calculating the differences between *OverallSurvival* and both *LeukemiaFreeSurvival* and *AMLOverallSurvival*, replacing the original columns while ensuring no negative values. Additionally, zeros in survival-related variables were adjusted by adding 1 to prevent issues during log transformations.

*Experimental Setting:* We simulated a FL environment with three nodes, each with different amounts of training data. Each node trained its VAE-BGM model for up to 200 epochs, with an Early Stopper monitoring validation loss. The training included five federated steps, where information was shared at most every 200 epochs. To evaluate aggregation strategies, we tested two approaches: FedAvg(7), where nodes shared model weights and performed a weighted aggregation based on dataset size, and Synthetic Patient Data Sharing (SDS)(8), where nodes exchanged synthetically generated patient records to enhance data diversity without compromising privacy. The VAE-BGM architecture consisted of an encoder with ReLU activations, a decoder with dropout (20%), and a latent space with 15 Gaussian distributions and 256 hidden neurons. The BGM component used a Dirichlet process prior with 15 mixture components and individual covariance matrices. Training was conducted with a batch size of 64, an 80%-20% train-validation split, and early stopping. By comparing FedAvg and SDS, we assessed the benefits of parameter aggregation vs. knowledge transfer via synthetic data in heterogeneous and privacy-sensitive FL scenarios.

**Federated Bayesian Networks for Synthetic Data Generation**

*Methodology:* A BN is a probabilistic graphical model that provides a compact representation for describing the joint probability distribution of a set of random variables in a dataset(9). It consists of two main components: a directed acyclic graph (DAG) and a set of parameters. The graph represents each variable as a node and conditional dependencies between variables as directed edges, where preceding nodes are referred to as parents and posterior nodes as children. Parameters correspond to the local conditional probability distributions (CPDs) associated with each node in the graph, given its parents. By leveraging the structure and parameters of a BN, synthetic data can be generated by sampling variables in topological order according to their respective CPDs.

BNs have proven particularly useful for generating high-quality synthetic tabular data (10). They offer the advantage of interpretability, the ability to model complex distributions. They can be integrated with other privacy-preserving technologies such as differential privacy (11), making them strong candidates for generating synthetic data in the healthcare domain. This work implements a BN in a federated setting extending the approaches presented in (11) and (12) in the centralized setting. The design comprises three main parts: the preprocessing step, the structure learning step, and the parameter learning step.

*Preprocessing:* The preprocessing step involves discretizing numerical attributes and encoding categorical attributes. In our approach, clients share min and max values for each numerical attribute, which the server uses to compute the global min and max, allowing each client to locally discretize its attribute into bins. For categorical data, the server collects all categories for each attribute and shares them with the clients to ensure consistent integer encoding across nodes. While common categories pose no privacy risk, the server could exploit rare or sensitive attributes, as well as the statistics (min and max values) to infer private information. This can be mitigated with SMPC protocols (13,14). Alternatively, in a production-ready setting clients could agree on a standard data model before starting the federation.

*Structure learning:* The structure learning step involves learning the DAG structure of the BN model in a distributed manner. In our approach, we draw inspiration from (15), which applied Grammatical Evolution in a federated setting using individual migration to enhance diversity and extend the genetic algorithm from (12) to support federated Bayesian network structure learning. Each client runs the GA algorithm locally, exchanging the top-performing individual (DAG structure) with the server at predefined intervals. The server aggregates and redistributes these individuals in several federated rounds, allowing clients to refine their selection process. Mutation and crossover operations locally further integrate information from multiple clients, improving structure learning. In the last iteration, clients rank their best individuals, and the server selects the DAG structure with the highest average fitness score as the global solution for the structure learning step.

*Parameter learning:* The parameter learning step involves estimating the CPDs based on the global DAG structure. To achieve this, each client must share *sufficient statistics* to compute the CPDs (i.e., the counts for all possible combinations of nodes and their parents). However, sharing this data may expose sensitive information about their private datasets to the server. This can be mitigated using additional privacy-preserving techniques during the aggregation process. One such technique is the Distributed Laplace Perturbation Algorithm (DLPA) (16), which leverages the threshold Paillier cryptosystem to protect intermediate results and exploits the Laplace distribution's property of infinite divisibility to ensure differential privacy in the output.

*Experiment setup:* For the experimental setup, we used the following parameters for synthetic data generation (SDG) in a federated setting. We set up three federated nodes, each with a different number of samples, and trained the BN model over 40 federated rounds. Each client performed 10 local rounds to train the GA algorithm. The final two federated rounds were dedicated to structure selection and parameter estimation. The genetic algorithm parameters were adopted from the original paper (12).

### Supplementary Information 4| Synthetic Validation Framework (SAFE)

Assessing the quality of the generated data is a notoriously challenging task. A Synthetic vAlidation FramEwork (SAFE)(17) powered by Train was implemented to assess the models and quality of the synthetic patients with respect to their fidelity, correlation, utility, and privacy.
The evaluation encompassed multiple data domains, including demographics, clinical features, genomics (analyzed as categorical variables), and clinical outcomes (overall survival and leukemia-free survival probabilities).

To assess data fidelity, we compared distributional properties, correlations, and principal component structures between real and synthetic datasets across all data types. Descriptive statistics and pairwise association analyses were conducted to quantify similarity. We introduced two composite metrics: Clinical Synthetic Fidelity (CSF) and Genomic Synthetic Fidelity (GSF), calculated as the average performance across multiple statistical tests. A fidelity score ≥85% was considered optimal for both metrics.

To evaluate the ability of synthetic data to capture real clinical-genomic structures, we applied hierarchical Dirichlet clustering to stratify real and synthetic patients and identify meaningful genomic subgroups. Kaplan–Meier survival curves, Cox proportional hazards models(18), and L1-penalized Cox regression were used to identify features associated with survival outcomes. Model discrimination was quantified using Harrell's Concordance Index(19).

Privacy preservation was assessed by measuring the extent of overlap and similarity between synthetic and original data. We calculated the Identical Match Share (IMS) to quantify exact matches between synthetic and real records. To further evaluate re-identification risk, we computed the Distance to Closest Record (DCR) and the Nearest Neighbor Distance Ratio (NNDR). NNDR, defined as the ratio of distances to the closest and second-closest real neighbors for each synthetic record, provides a normalized measure of proximity. An optimal NNDR range was defined as 0.60–0.85; values near 0.50 suggest insufficient similarity (potential fidelity loss), whereas values approaching 1.00 indicate excessive similarity (potential privacy risk).

Finally, we employed Shapley Additive Explanations (SHAP) to interpret the predictive models and assess the explainability of AI-based outputs. SHAP values provided individualized explanations of feature importance, grounded in cooperative game theory.

### Supplementary Information 5| Deliverable 1.2: First Interim Periodic Report

#### 1. Explanation of the Work Carried Out by the Beneficiaries and Overview of the Progress

GenoMed4All is a European initiative aimed at transforming the diagnosis and treatment of haematological diseases through the application of artificial intelligence and secure, cross-border data sharing. The project leverages FL to enable the pooling and analysis of genomic, clinical, and multi-omics health data while maintaining privacy and complying with data protection regulations. Built on existing infrastructures, including HPC resources, clinical registries, and data repositories, GenoMed4All aspires to accelerate personalized medicine by integrating data from 10 initial clinical partners, eventually expanding to 66 repositories across 15 EU Member States, along with 20 additional sources.

The initiative demonstrates the potential of transparent and trustworthy AI tools, employing advanced machine learning techniques such as deep learning, deep generative models and FL, across three clinical use cases involving both oncological and non-oncological haematological diseases. The project is structured in three phases over 48 months: initial requirements gathering and technical design; development, pilot testing, and refinement of the platform; and final validation and optimization.

The work completed thus far reflects solid progress across all nine Work Packages (WPs). WP1 has successfully overseen coordination and risk management despite COVID-19 constraints. WP2 has established an ethical and data protection framework in line with GDPR and EU AI guidelines. WP3 completed the definition of technical, clinical, and user requirements. WP4 delivered a prototype of the FL infrastructure. WP5 focused on data standardization, anonymization, and curation processes. WP6 initiated the development of AI algorithms and validation protocols. WP7 began applying these tools to real-world clinical data in defined use cases. WP8 initiated activities around genomics standardization, impact evaluation, and user training. WP9 established a recognizable digital presence and advanced stakeholder engagement and dissemination activities.

The project has met its Phase 1 objectives and is progressing well into Phase 2, laying a strong foundation for its long-term goal of enhancing personalized care in haematology through AI-powered insights. During the initial phase of the GenoMed4All project, five key milestones were successfully achieved, marking critical progress in the establishment of the project's digital infrastructure, ethical and legal frameworks, data quality assessment processes, user co-design methods, and the deployment of a functional FL model. These achievements have laid the groundwork for the project's broader mission of leveraging AI and cross-border data sharing to transform the management of haematological diseases.

In terms of strategic objectives, considerable advancement has been made across several areas. The project began integrating clinical repositories across Europe through a hybrid FL approach, with 10 clinical partners onboarded and expansion efforts underway. The foundational architecture of the GenoMed4All platform is being implemented to support data pooling and HPC usage, with multiple “-omics” data types identified and standardization efforts initiated. While full validation of some KPIs remains scheduled for later phases, core infrastructure elements and open standards integration are progressing as planned.

AI algorithm development has also seen promising early results. For example, statistical tools aimed at improving the predictive accuracy of clinical outcomes in diseases such as MDS have already surpassed benchmarks, with similar work underway for additional use cases. Other AI-driven objectives, such as multi-stage modeling for personalized treatment and the integration of biological age estimation, are scheduled for subsequent project periods.

Regarding data handling, mechanisms for anonymization, curation, and cybersecurity are in active development, with measurable security indicators to be validated in future phases. Standards in genomics and cooperation with European Digital Innovation Hubs are progressing through collaborative efforts among technical WPs, setting the stage for secure and interoperable data use across clinical and research environments.

Ethical and legal frameworks are well-established, with data assets fully inventoried and quality assessments completed, ensuring compliance with GDPR and the ethical integrity of all data sources.

The project’s outreach and exploitation strategy is also underway, targeting both academic and commercial audiences. Preliminary impact is evident through an expanding digital presence, the dissemination of printed materials, event participation, and the early pipeline of scientific publications. Plans for achieving market introduction of AI-based clinical tools and securing technology readiness levels are aligned with the final project phases.

Overall, GenoMed4All has made substantial progress in establishing its infrastructure, validating initial use cases, and initiating the development of advanced AI methodologies, with many of its foundational goals achieved or actively in progress. The next phases will focus on full-scale deployment, performance validation, and broader dissemination and impact across the European healthcare ecosystem.

*Explanation of the work carried out per WP*

During the initial reporting period, the GENOMED4ALL project successfully executed key activities under WP1 (Management and Coordination) and WP2 (Ethical, Legal, and Data Protection Framework).

WP1, led by UPM, focused on general project coordination, financial and administrative oversight, and communication with the Project Officer. UPM organized the project’s kick-off and three plenary meetings, maintained internal coordination through monthly WP leader meetings, submitted the Deliverable D1.2 Interim Periodic Report, and supported the preparation and submission of a project amendment. DW contributed by defining and implementing quality assurance and risk management procedures, including the development of the D1.1 Quality Plan and coordination of deliverable reviews. ICH, as the technical and scientific coordinator, ensured technical coherence, monitored progress across WPs, and facilitated the resolution of technical issues through regular follow-ups. No deviations from the Description of Action were reported.

WP2, under the leadership of i~HD, established the ethical and data protection foundation of the project. This included forming an independent Ethical Advisory Board, completing a project-wide Data Protection Impact Assessment, mapping data flows, and drafting the Joint Controller and Associate Member Agreements. Deliverables such as D2.1 and D2.2 were submitted with minor delays to accommodate thorough review and expert input. The work also covered GDPR compliance, ethical assessments across the use cases, and preparation of consent documentation. DW, UNIBO, and other partners played vital roles in aligning AI development with ethical standards and forthcoming regulatory frameworks. All tasks proceeded with no significant deviations, and planning for the upcoming period includes further review of security measures, completion of outstanding agreements, and continued oversight by the Ethical Advisory Board.

WP3, led by Data Wizard (DW), from Month 1 to Month 12, successfully coordinated the co-design of user, technical, and clinical requirements for the GenoMed4All platform. The objective was to establish a harmonized foundation for data integration, AI implementation, and clinical alignment across three disease use cases, SCD, MDS, and MM.

The WP involved extensive collaboration among clinical, technical, and AI stakeholders. Partners contributed to the identification and linkage of relevant genomic and health data repositories,the definition of end-user and beneficiary requirements, and development of a common data model. DW led the effort by moderating interdisciplinary meetings, supporting deliverables, and organizing two platform workshops: a two-session workshop and a virtual patient engagement workshop, ensuring active involvement from clinicians, AI developers, and patient representatives.

Key activities included technical requirement gathering, clinical data harmonization, and the assessment of suitable data standards, resulting in the selection of FHIR for data modeling. Several partners such as UPM, DED, and FORTH contributed to the structuring of the data model, incorporating considerations for AI performance metrics, explainability, and compatibility across technologies. Clinical partners like VHIR, UMCU, and PU facilitated the definition of clinical aims, metadata models, and GDPR-compliant data handling, especially for the SCD use case. UNIBO supported the AI implementation strategy across use cases, while partners like CRG and CING provided valuable input on data standard comparisons and repository analysis. Three key deliverables were produced on schedule: D3.1, a roadmap for data repository linkage; D3.2, end-user requirements for AI implementation in clinical settings; and D3.3, the finalized data model. These deliverables collectively outline the project's approach to data standardization, integration, and architecture for FL. No deviations from the Description of Action were reported, and all planned milestones were achieved.
In summary, WP3 established the foundational framework for the GenoMed4All platform by aligning clinical needs, technical specifications, and user requirements through a collaborative, structured process that bridged disciplines and ensured stakeholder engagement.

During the reporting period, significant progress was achieved in WP4, focused on designing and developing the FL infrastructure for GenoMed4All. Led by UPM, this work laid the foundation for a hybrid FL platform by establishing the system's functional and non-functional requirements, performing a comprehensive technology analysis, and selecting core tools such as Flower for FL, MLFlow for model management, and Keycloak for security. A prototype encompassing essential features, secure data access, model management, and task definition, was developed, with key modules dockerized to support streamlined deployment. Collaborative contributions from DXC, CRG, FORTH, and CINECA supported efforts in PoC development, algorithm implementation, HPC integration, and platform deployment. Deliverables D4.1 and D4.2 were submitted, and milestone MS05 was achieved. The next six months will focus on completing a fully functional version of the platform.

WP5, under the leadership of DED, concentrated on data engineering, anonymization, and the creation of privacy-preserving tools. Progress included establishing a common data model and implementing data homogenization strategies for diverse omics and clinical data types. FORTH led the anonymization task, identifying suitable tools such as ARX and TCIA and initiating guideline development. Pre-processing pipelines tailored to each use case were developed under MLL's guidance, while DXC led efforts to standardize and define data workflows and ETL pipelines for integration into the federated infrastructure. Key deliverables D5.1 and D5.2 were submitted on schedule, and planning is underway for large-scale data ingestion into the common data model. No significant deviations from the plan were reported in either WP.

During the current reporting period, significant progress has been achieved under WP6, which focuses on the development and implementation of AI algorithms for personalized medicine in haematological diseases. Led by UNIBO and running from Month 6 to Month 48, the WP saw substantial contributions from multiple partners. UPM took a leading role, both in coordinating contributions and advancing FL methodologies. They delivered D6.2 and introduced a pipeline for federated implementations, theoretical insights into ADMM and PVI algorithms, and developed an original survival analysis model based on variational autoencoders (SAVAE). Their prototype also incorporates collaborative learning strategies using synthetic "virtual patients" for training and imputation purposes. UPM also devised a validation framework to assess the viability and benefit of federated models based on synthetic benchmark scenarios.

ICH coordinated comparative analysis between statistical and deep learning methods for genomic-based classification in rare hematologic cancers, aiming for a high-impact publication and developing a public repository for model code. VHIR, APHP, and CING contributed to defining AI strategies aligned with the SCD and MDS clinical use cases. UCPH developed an encoder-free generative model for structure discovery and missing data imputation in Genomed4All datasets, while UNITO focused on data harmonization and AI pipeline preparation, contributing to multiple deliverables and model development for SCD, MM, and MDS. FORTH contributed to imaging data analysis, producing guidance for radiological segmentation of bone marrow lesions.

In WP7, led by ICH, the project focused on use case validation and practical testing of federated AI models. UPM spearheaded the creation of a robust validation protocol based on simulated benchmark scenarios and virtual patient generation. ICH directed the design and outcome definition of oncological use cases and initiated strategic collaborations with European registries like EurobloodNet. UMCU, as the leader of the SCD non-oncological use case, was instrumental in preparing data collection approvals and harmonization efforts, particularly in metabolomics and imaging analysis. Clinical partners such as APHP, MLL, and IBSAL provided curated datasets and contributed to the definition of clinical outcomes and AI algorithm validation.

The work culminated in the timely submission of key deliverables including D6.1, D6.2, D6.3 under WP6, and D7.1 under WP7. These efforts collectively serve to validate AI-driven approaches for clinical decision support in haematological malignancies and pave the way for FL adoption. The next reporting period will focus on finalizing the SCD analysis and further refining models for MDS and MM, with continued integration and validation efforts across clinical and technical partners.

WP8, led by CRG and spanning from M01 to M48, focuses on genomics data standardization, value-based healthcare, open science data, and education/training. Notable contributions include the submission of a comprehensive genomics data standardization plan (Deliverable D8.1), introductory workshops on data pipelines and federated platforms, and early-stage research on value-based healthcare outcomes. Significant coordination has taken place across various work packages to ensure harmonization of genomics data, planning for data reuse, and alignment with European Open Science Cloud (EOSC) initiatives. Collaborative efforts also include preparing a dissemination strategy featuring webinars targeted at clinicians and patients. Looking ahead, the WP will continue developing D8.2, providing support for data standardization, harmonization, and contributing to healthcare outcome measurement frameworks.

The four core tasks of WP8 are progressing as planned:

- **T8.1** (Genomics Data Standardization) delivered a key report, organized workshops, and supports data sharing via the Beacon federated discovery platform.
- **T8.2** (Value-Based Healthcare Analysis) is in its early stages with planning meetings underway and D8.4 due by M48.
- **T8.3** (Open Science Data Collaboration), led by CINECA, has initiated reuse planning with an MDS dataset as a pilot.
- **T8.4** (Training and Education), led by VHIR, has drafted a GenoMed4All educational program in collaboration with ERN-EuroBloodNet, with webinars under development.

WP9, coordinated by AUS, oversees impact, outreach, and stakeholder collaboration. The WP has established a strong communications and dissemination infrastructure, including branding, website, social media presence, and outreach content. To date, WP9 has reached over 1,300 followers, published 24 blog entries and 10 Zenodo entries, and contributed to 11 upcoming scientific publications. One workshop has been hosted, and the team has participated in multiple webinars and public events.

Under T9.1 (Dissemination and Communication), the consortium has developed all foundational branding and communication tools, published educational content, and supported promotional actions. T9.2 (Exploitation), led by DW, is developing a roadmap and identifying Key Exploitable Results (KERs) through internal analysis and integration with platform design efforts. T9.3 (Stakeholder Collaboration), led by AUS, has successfully engaged new stakeholders, hosted collaborative events, and expanded the project's network to include initiatives such as ERN-EuroBloodNet, 1+ Million Genomes, and sister EU projects.

Across both WPs, there have been no deviations from the Description of Action (DoA), and no corrective actions are required. Planning for the next reporting periods includes further deliverables, targeted dissemination efforts, value-based healthcare surveys, and continued stakeholder engagement.

*Impact*

During the initial phase of GenoMed4All, considerable effort has been dedicated to defining the project's intended impact and implementing key actions to enhance its visibility and reach. This has resulted in the creation of over 430 social media posts, 24 blog articles, 10 Zenodo publications, with 10 more underway, and participation in various dissemination activities, including six featured events, one hosted workshop, and attendance at three webinars. The project has built a growing online presence, with more than 1,300 followers on Twitter and LinkedIn, and has distributed over 250 hard copies of promotional materials. Community expansion, particularly in the haematological domain, has been driven by a targeted engagement strategy aimed at increasing clinical partner participation. Comprehensive details, including the full impact master plan and an initial evaluation of impact, exploitation, and sustainability, are available in D9.1 and D9.3.

#### 2. Update of the Plan for Exploitation and Dissemination of Results

The initial strategy for communication, dissemination, and exploitation of the GenoMed4All project was outlined in Deliverable D9.1, GenoMed4All Impact Master Plan, submitted in Month 6. This plan will be further refined and expanded in Deliverables D9.2 and D9.3, due in September and November, respectively. As part of its dissemination efforts, GenoMed4All established a dedicated repository on Zenodo to meet Open Access requirements, ensuring broad accessibility to publications, deliverables, brand materials, and press releases.

To date, the project has produced several notable scientific outputs. These include peer-reviewed articles, clinical studies, and conference posters addressing key topics such as protein stability in haploinsufficient genes, improved classification and prognosis of MDS through AI, sex-informed approaches to patient stratification, and clonal hematopoiesis in elderly populations. Additional work focuses on AI-driven diagnostics and mass spectrometry applications in SCD, as well as radiomics-based monitoring techniques. Two more papers are currently in progress, one exploring multimodal health data modeling in FL contexts, and the other comparing modern and classical machine learning methods in haematological diseases.

The project has been active in prominent dissemination events, participating in high-profile conferences and meetings such as the European Hematology Association Congress, the 4th Global Congress on Sickle Cell Disease, and the European Conference on Rare Diseases. GenoMed4All has also hosted and taken part in various webinars and workshops focused on genomics, patient engagement, AI in personalised medicine, and data sharing in health research. These activities have reinforced the project’s visibility and engagement across clinical, research, and patient communities.

### Supplementary Information 6| Deliverable 1.3: Second Interim Periodic Report

#### 1. Introduction

GenoMed4All is a European initiative aimed at revolutionizing the management of haematological diseases through the integration of AI and standardized, interoperable cross-border data sharing. The project facilitates the secure and privacy-conscious pooling of genomic, clinical, and other “-omics” health data via a FL platform, promoting advancements in personalized medicine. Leveraging existing infrastructures such as high-performance computing resources, hospital registries, and data processing tools, GenoMed4All focuses on both common and rare haematological conditions, initially integrating ten clinical partners with plans to expand to 66 clinical repositories across 15 EU Member States, as well as additional repositories outside the EuroBloodNet network.

The project employs advanced and explainable AI techniques, including deep learning, variational autoencoders, and generative models, combined with statistical and machine learning methods to analyze complex "-omics" datasets. This approach aims to generate reliable, clinically meaningful insights across three use cases covering oncological and non-oncological haematological diseases, ensuring full compliance with data protection and ethical standards. GenoMed4All spans 48 months, organized into three iterative phases: the first focusing on user requirements, system architecture, initial AI model development, and platform implementation; the second on platform completion and pilot testing; and the third on final validation, analysis, and refinement. Milestones achieved in the initial phases demonstrate significant progress, including platform development, FL implementation, data quality assessment, and co-design with end users.

Key objectives include linking clinical repositories across Europe through a hybrid FL approach, developing the infrastructure to support multi-omics data integration, enhancing predictive accuracy of AI tools, and ensuring robust data security and anonymization. The project has already connected initial clinical partners and identified over ten types of omics data for integration, while ongoing efforts focus on scaling the platform, improving AI model performance, and standardizing data formats. GenoMed4All also emphasizes ethical compliance, data protection, and stakeholder engagement, with a growing outreach program that includes numerous publications, events, and community-building activities. The project is on track to deliver innovative AI-driven decision support systems and reach significant technology readiness levels by its conclusion.

*Explanation of the work carried out per WP*

WP1, led by UPM, has overseen project management and coordination from month 1 to 48, ensuring continuous administrative, financial, and strategic monitoring alongside regular partner communication and progress tracking through monthly meetings. UPM managed consortium-wide coordination and reporting, while DW led quality assurance and risk management throughout the project, and ICH provided technical and scientific oversight, supporting task execution, fostering collaboration, and advancing key technical milestones such as platform deployment and validation.

WP2, under the leadership of i~HD, focused on establishing the ethical, legal, and data protection frameworks, including AI trust, by ensuring compliance with regulatory requirements, updating data protection impact assessments, executing Joint Controller Agreements, and facilitating ethics approvals across sites. The Ethical Advisory Board provided ongoing oversight, and the team addressed delays in AI trust deliverables by conducting internal assessments aligned with the EU ALTAI framework, maintaining project compliance without impacting resources.

WP3, led by DW, concentrated on co-designing user, technical, and clinical requirements during the first 12 months. Several partners, including APHP, IBSAL, UNIBO, and UCPH, contributed to defining clinical use case requirements, performing literature reviews, and designing user interfaces for medical data modeling to ensure alignment between technical development and clinical needs.

WP4, led by UPM from month 6 to 48, focused on the development and deployment of the FL (FL) infrastructure and data sharing mechanisms. Key efforts included agile sprint planning, platform development and security enhancement, and deployment across test and production environments at Cineca and Humanitas. The team introduced a validation protocol, published relevant findings, and expanded HPC resource utilization for AI training and pipeline adjustments, providing consortium-wide HPC training. The FL platform evolved with improved model management and monitoring features, fully validated across environments, and supported by detailed technical guides. The shift in HPC use reflected adaptive project needs without impacting deliverables or timelines. Overall, WP4 successfully deployed a functional, secure FL platform aligned with user requirements and project goals.

WP5, spanning from month 6 to 30 and led by DED, focused on data engineering, anonymization algorithms, curation, and privacy preservation tools within the GenoMed4All project. Multiple partners contributed to distinct but complementary activities including dataset documentation, understanding file formats, developing a data homogenization platform, defining a common data model (CMD) based on FHIR standards, and supporting anonymization pipelines. Efforts included standardizing clinical terminologies, preprocessing genetic and imaging data, and integrating these processes within a high-performance computing (HPC) environment hosted by CINECA.
Task T5.1 involved monitoring anonymization requests and supporting genomic data anonymization, culminating in D5.3, which detailed the anonymization methods for structured, imaging, and genomic data without any deviations or corrective actions required.
Task T5.2, led by MLL, centered on uniform genomic data preprocessing, developing and validating pipelines for the use cases, optimizing their deployment on the CINECA HPC infrastructure, and providing dockerized tools to integrate anonymization into data ingestion. D5.4 was submitted on schedule, describing the MDS bioinformatics pipeline and providing educational resources. While the storage of MM and SCD pipelines on the HPC system is ongoing and extends beyond WP5's timeline, these activities will be completed under subsequent WPs.
Task T5.3, involving DXC, UPM, and FORTH, defined the architecture of the data homogenization platform, established a common data model using FHIR and OHDSI Athena vocabularies, and developed the data ingestion and extraction processes to facilitate machine learning applications. This task included delivering tools for data harmonization, integration with FL platforms, and a user-friendly interface. MDS was used as a primary use case to define CMD specifications, and initial work on SCD CMD began. D5.5, the data homogenization framework, was completed without issues.

Overall, WP5 successfully advanced data standardization, preprocessing, anonymization, and integration capabilities critical to GenoMed4All, delivering key frameworks and tools on time, while smoothly transitioning ongoing tasks to subsequent project phases.

Between months 19 and 36, WP6 focused on advancing AI algorithms tailored for personalized medicine in hematological diseases, under the leadership of UNIBO with contributions from multiple partners. Significant progress included designing a validation protocol for federated deployment of AI algorithms addressing the specific use cases (MM, MDS, SCD). UPM developed and validated a synthetic patient data generator, enabling robust training of machine learning models despite data scarcity, and implemented survival and relapse prediction models evaluated in simulated federated environments.
ICH contributed clinical insights and methodological frameworks for multimodal AI analysis, including molecular classification of chronic myelomonocytic leukemia and natural language processing for clinical data extraction. VHIR led the development of AI-based risk score models in SCD using genomic, metabolomic, and clinical data with advanced clustering and machine learning techniques. UNIBO implemented centralized and federated AI algorithms, developed Clinical Decision Support Systems (CDSS) across the three diseases, and conducted FL simulations validating performance gains over isolated models. UNITO focused on federated AI model development for MDS and MM, including survival and relapse prediction, while also supporting CDSS development and explainability analysis.

Additional partners contributed to model validation, federated algorithm implementation, and synthetic data generation, with FORTH advancing a federated XGBoost algorithm and survival analysis on synthetic MDS data. UCPH developed a flexible multimodal deep generative neural network tested on large medical datasets. Throughout the period, all AI software was deployed in the project repository, datasets were preprocessed and harmonized, and FL validation protocols were established, showcasing the feasibility and advantages of federated approaches for sensitive clinical data. Deliverables included software releases, clinical decision systems, radiomics models, and FL results, collectively supporting the development of personalized AI-driven clinical tools in hematology.

WP7, led by ICH and spanning months 10 to 48, focuses on use cases and validation across multiple hematological diseases, with significant contributions from consortium partners. Activities include data collection, standardization, algorithm development, and clinical trials execution, supported by expertise from clinical, technological, and research institutions.
ICH has overseen the entire data collection and dissemination process, fostering collaboration with European

registries and external partners, and has been pivotal in installing and refining the federated platform and homogenization tools. They have also driven scientific visibility through international presentations and publications and contributed to education and open science tasks. VHIR and UMCU lead the SCD use case, successfully enrolling over 1,000 patients across Europe and standardizing clinical and multi-omic data, despite some administrative and logistical challenges. UNIBO developed federated and centralized algorithms across three use cases contributing significantly to clinical decision support system development and imaging analysis.

Collaborative efforts on MDS include creating a harmonized classification system using multi-modal data integration and AI-driven methods, validated in large international cohorts. Similarly, chronic myelomonocytic leukemia (CMML) has been molecularly classified through genomic and clinical feature analysis, providing a foundation for improved prognostic tools. The MM use case expanded its patient dataset for enhanced analysis, including harmonized genomic and PET/CT imaging data, identifying potential prognostic imaging markers.

Across all use cases, standardized datasets have been successfully released for algorithm development and clinical validation. Key deliverables, including methodology for data collection and trial progress reports, have been completed, with significant milestones met such as patient accrual targets in SCD and initial trial phases. The consortium's efforts have yielded impactful scientific publications, advancing AI applications in hematology and supporting the clinical utility of developed algorithms.

The activities across tasks T7.1 to T7.5 have progressed with a focus on the design, validation, data collection, execution, and evaluation of clinical use cases within the GENOMED4ALL project. Task T7.1, led by ICH with partners APHP, LU, NKUA, and UNIBO, successfully delivered the use case design and study protocols by month 18, updating the MM use case based on clinical discussions and validating AI algorithms tailored to address specific clinical needs. Task T7.2, led by UMCU and VHIR, developed and refined a pilot plan for the non-oncological SCD use case, aiming to integrate multi-omics data and clinical features to predict patient outcomes through AI-driven models. Task T7.3, led by CING, established a harmonized and standardized methodology for data collection across trials, leveraging platforms like RADeep and ENROL to ensure consistent and high-quality clinical and omics data from multiple European centers, achieving full enrollment for the SCD use case. Task T7.4, led by VHIR, initiated trial execution to test the federated platform and AI algorithms, with the first hospital node (ICH) successfully deploying the system and ongoing training and setup at other sites. Key clinical AI applications include automated MRI lesion segmentation and development of predictive risk scores for SCD complications, with support from high-performance computing resources. Finally, task T7.5, co-led by ICH and VHIR, is responsible for validating and reviewing trial outcomes and gathering stakeholder feedback; however, the final deliverable was delayed due to partial platform deployment and pending integration of federated algorithms. Corrective actions are underway, including the distribution of updated installation guidelines and preparation for broader platform deployment, with an expected completion and report release within the next four months. Overall, the project demonstrates steady progress in protocol refinement, data standardization, AI validation, and platform deployment, while addressing initial delays through structured corrective measures.

WP8 focuses on genomics data standardization, value-based healthcare analysis, open science data collaboration, and training/education over a 48 months period, led by CRG. The team includes multiple partners contributing expertise in data modeling, genomics standardization, healthcare parameters, and educational program development. WP8 activities during months 18 to 36 centered on bibliographic reviews, data analysis, and surveys to understand use cases and their data needs, aiming to draft standardization guidelines and best practices. Key achievements include initial evaluation of genomic data pipelines and workflows, defining parameters for value-based healthcare models, agreement on metadata publication protocols via the EOSC portal, and launching the first wave of the GenoMed4All & ERN-EuroBloodNet educational program on AI in hematology. Training sessions were well-attended and recorded for broader dissemination. The open science task established workflows for dataset publication and access requests, ensuring partner consensus. All tasks are progressing without deviations, with deliverables and reports planned for completion by month 48. Overall, WP8 provides critical coordination, expertise, and dissemination efforts to support genomic data integration and education within the project framework.

WP9, led by AUS and running from month 1 to month 48, focuses on impact, outreach, and collaboration within the GenoMed4All project. The WP is progressing as planned, with increased dissemination efforts since month

19, including peer-reviewed publications, targeted communication campaigns, and a dedicated educational program. Key activities involve managing digital channels, producing promotional materials, supporting publications, and internal communication tools. Consortium partners actively contribute through data sharing, dissemination at conferences, social media outreach, and collaboration on exploitation planning. WP9 emphasizes community building by developing an Associated Members Onboarding Handbook and a positioning manifesto to onboard external partners and ensure sustainability. The project has achieved significant outreach metrics, including substantial social media engagement, website traffic, numerous publications, and event participation. Exploitation activities include prioritizing KERs and conducting workshops to explore exploitation routes, supported by a business plan submitted in month 24. Stakeholder collaboration is reinforced through coordination with associated projects and networks, organizing and attending various workshops, webinars, and scientific meetings. Overall, WP9 aims to maximize the project's visibility, impact, and long-term sustainability through strategic dissemination, engagement, and exploitation efforts. There have been no deviations from the plan, and corrective actions have not been necessary.

**2. Update of the Plan for Exploitation and Dissemination of Results**

The Communication, Dissemination, and Exploitation strategy was updated and detailed in the interim report (D9.2) at month 21, subsequently revised to reflect progress through month 36, with a final comprehensive report (D9.4) forthcoming. A dedicated Zenodo repository ensures compliance with European Commission Open Access mandates, hosting 15 public outputs including publications, deliverables, and press materials. GenoMed4All has produced numerous peer-reviewed publications spanning clinical studies, articles, and conference presentations across prestigious platforms and journals, covering topics from AI applications in hematology to synthetic data generation. The project has been actively featured in multiple high-profile events, including ERN-EuroBloodNet meetings, international congresses such as ASH and EHA, and specialized workshops globally, engaging thousands of participants. Additionally, GenoMed4All has organized and contributed to various webinars and workshops focusing on genomics, AI, data sharing, and patient engagement, reaching wide audiences in both academic and patient communities. These sustained dissemination efforts have effectively promoted the project’s scientific advancements and fostered collaboration within the hematology and AI research communities.

### Supplementary Information 7| Deliverable 2.1: Data processing, data management plan and GDPR compliance report

**1. Introduction**

This deliverable outlines the coordinated strategy developed under WP 2 to guide GenoMed4All partners in achieving local GDPR compliance and promoting best practices for data protection across the consortium. Recognizing the complexity of aligning data governance across multiple European jurisdictions, the plan emphasizes a dual approach: enabling individual partners to meet their specific national requirements while supporting collective understanding and coordination at the consortium level.

Compliance depends on mapping data flows accurately, understanding local legal obligations, and ensuring that each partner’s role (as data provider, recipient, or controller) is clearly defined. Given the diversity of regulatory interpretations across EU member states, the deliverable acknowledges that a one-size-fits-all model is neither feasible nor appropriate. WP2 proposes a flexible yet harmonized structure built on GDPR-compliant tools and frameworks, such as the Data Protection Impact Assessment (DPIA), to identify and mitigate risks in high-impact data processing activities. GenoMed4All itself does not act as a legal data controller but serves as a central facilitator to support transparency, risk assessment, and regulatory clarity.

To operationalize this approach, the consortium has developed and deployed a range of practical instruments, including a standardized DPIA template, partner-specific compliance questionnaires, and an information asset register. These tools collectively support the identification of data responsibilities, compliance gaps, and legal bases for processing, while also serving as a foundation for broader ethical and data-sharing governance.

Initial implementation began with the SCD group as a pilot to test this model in practice. Insights gained from this pilot have informed engagement strategies with other disease areas, such as MM and MDS, and highlighted the importance of distinguishing between consortium-level coordination and site-level compliance. A data-sharing agreement template, informed by questionnaires and consortium discussions, has also been drafted to formalize the legal and ethical parameters for intra-consortium and external data exchanges.
Ultimately, this deliverable presents a comprehensive, evolving framework for ensuring GDPR compliance across a diverse research partnership, reinforcing GenoMed4All's commitment to lawful, ethical, and secure data handling throughout the project's lifecycle.

### 2. Results to Date and Forthcoming Work

To date, the GenoMed4All project has progressed significantly in establishing its data protection compliance framework, with key components currently in development and refinement. DPIA serves as a foundational, evolving document, shaped by responses to distributed questionnaires and insights from GDPR workshops. This DPIA is designed not only to guide compliance within the project but also to assist individual partners in assessing their own workflows and obligations. Concurrently, the Information Asset Register is being constructed to document each partner's data responsibilities, ensuring traceability, governance, and regulatory alignment across the consortium.

In terms of engagement, the project has adopted an incremental approach, piloting the process through the SCD VHIR site to evaluate the necessary steps for broader partner involvement. This includes technical engagement, dissemination of questionnaires, updates to the DPIA and asset register, and collaborative workshops, ultimately culminating in agreed approaches to pseudonymisation and anonymisation in compliance with local and EU standards.

The Data Management Plan (DMP), structured around Horizon 2020 guidelines, outlines the nature, purpose, and handling of de-identified clinical and omics data. This data is central to the development of machine learning models aimed at risk stratification across diseases such as SCD, MM and MDS. Both existing and newly generated data will be used, with the possibility of re-use by third parties under a governance model ensuring compliance with FAIR principles and ethical standards.

The DMP also addresses future data sharing, emphasizing that open access will be pursued where robust anonymisation is possible and legally permitted. This includes plans for the creation of a central anonymous repository, subject to licensing agreements and oversight by a Data Access Committee. Interoperability is a core objective, with HL7 FHIR standards being adopted to ensure consistency across multiple pilot sites.

In addition to outlining technical and ethical aspects of data handling, the DMP provides a preliminary resource and cost framework, noting that the full financial requirements for long-term preservation and access will be assessed as the project matures. Security protocols, including encrypted data transfer, strict access controls, and audit mechanisms, are being implemented to protect sensitive information.

Finally, the ethical dimension remains a focal point, with data sharing contingent on compliance with national and institutional review board requirements. The project does not involve direct patient questionnaires; instead, informed consent and data use are managed at the institutional level.

Overall, the project's current efforts focus on establishing a compliant, transparent, and interoperable data infrastructure, with a clear trajectory toward secure and ethically responsible data sharing, supporting both the project's aims and the broader research community.

### 3. Conclusions

In conclusion, this deliverable outlines the approach adopted by WP2 to understand, map, and guide data flows, management, and compliance requirements, thereby supporting partners in achieving their respective local regulatory obligations. The key outputs include the development of a data management plan aligned with the Horizon 2020 Template, facilitation of partners' approval processes, and monitoring of each partner's progress toward compliance. Additionally, the work provides essential support in areas such as information security and data flow design, leveraging insights gained from questionnaires, stakeholder engagements, and impact assessments to address uncertainties and offer clear guidance.

This work also contributes to ensuring that responsibilities around data processing are clearly defined and respected, preventing any regulatory oversight between jurisdictions. The collaborative approach taken enables the consortium to proactively resolve challenges and align on data protection and processing standards. Going forward, the focus will be on continued analysis of questionnaires, updates to the DPIA via GDPR Soundboards, finalisation of the Consortium-Wide Data Sharing Agreement, specification of governance agreements for external data contributors, and development of licensing frameworks for anonymous data access by third parties.

### Supplementary Information 8| Deliverable 2.2: 1st Report of the Ethics Advisory Board

#### 1. Introduction

D2.2 presents the inaugural report from the GenoMed4All Ethical Advisory Board (EAB), which consists of three independent experts specializing in advanced health data techniques to enhance outcomes and promote digital innovation. The EAB's purpose is to offer an independent evaluation of the project's ethical integrity, focusing particularly on data protection and the ethical development of AI. It aims to ensure that key ethical principles, such as non-malfeasance, equity, autonomy, and the public good, are upheld, while also verifying regulatory compliance and robust risk management practices related to data use.

Operating independently, the EAB reviews project processes, decisions, and approaches, meeting biannually and receiving regular updates, with the authority to convene extraordinary sessions if ethical concerns or risks arise. Its opinions are final and binding, with reports made publicly accessible to both project partners and the wider community.

This report summarizes the outcomes of the first EAB meeting held in January 2022, where members reviewed compliance documents, including the DPIA, DMP, and consent procedures. It details the EAB's assessment of ethical challenges identified by the project team and those uncovered by the board itself. The report concludes with the EAB's guidance and recommended next steps for addressing ethical considerations and ensuring ongoing oversight.

#### 2. Constitution of the EAB and first meeting preparation

The EAB of GenoMed4All serves as an independent oversight body, offering expert guidance on ethics, regulatory compliance, and risk management in data-driven healthcare innovation. Composed of three external experts with specialized backgrounds in privacy law, ethics, and public sector digital governance, the EAB works in close coordination with the Project Leadership and WP 2 team. These internal members provide contextual information, logistical support, and preparatory documentation to ensure the EAB is well-informed and positioned to evaluate the project's ethical standing.

The EAB was formally convened on January 19, 2022, with its constitution designed to ensure an impartial and knowledgeable review of the project's progress and regulatory adherence. During this first meeting, chaired by Nikolaus Forgó following his appointment, the EAB agreed to meet biannually, with provision for extraordinary meetings as needed. The composition of the Board reflects a deliberate inclusion of interdisciplinary expertise necessary to assess the project's approach to data ethics and protection.

Ahead of the meeting, external EAB members received a comprehensive briefing package one month in advance. This included key documents such as the DPIA, the DMP, consent forms, and participant materials, all curated to familiarize members with the project's ethical framework. These materials provided insights into both retrospective and prospective study designs for Genomed4All's use cases. The documentation also detailed existing ethical approvals, consent reconciliation efforts, and legal obligations under GDPR, ensuring the EAB had all relevant information to deliver a thorough and informed assessment.

**3. Ethical Challenges Under Consideration**

WP2 identified several core ethical challenges facing the GenoMed4All project, which were presented and discussed during the initial meeting of the EAB, alongside additional observations raised by EAB members. These challenges stem from coordination across clinical sites, university partners, and data stakeholders, particularly in the context of ML and AI.

One key question was whether all data-contributing sites should conduct self-assessments to determine the need for updated or additional research ethics approvals, especially in light of the specific ethical dimensions introduced by ML and AI. A second consideration was whether GenoMed4All should establish a unified set of ethical principles based on the variety of approvals received by participating projects. This would help ensure consistent standards across WPs and guide external collaborators in aligning with the consortium's ethical framework, particularly on issues like data protection, intellectual property, and liability.

Another significant challenge was the representativeness of the datasets, especially with respect to ethnicity and gender. Given the project's focus on rare diseases, clinical input will be essential to ensure algorithmic development accounts for diversity and does not inadvertently introduce bias through unbalanced data. The issue of data availability was also highlighted, particularly for SCD, where broad inclusion criteria may introduce heterogeneity that complicates algorithm training and validation. Ethical considerations must guide the selection of data for different phases of algorithm development to preserve both scientific integrity and fairness.

Separately, EAB members, while noting the project's ambitious scope and broad participation, expressed interest in improving the accessibility of project information. One member requested more introductory materials, prompting WP2 to suggest that the EAB review the project's website to assess whether it effectively communicates the project's objectives and activities.

**4. Outcome of the meeting**

The EAB agreed that Nikolaus Forgó would chair future meetings, with support from i~HD in preparing materials and agendas, and from UPM in organizing logistics, whether virtual or in-person. The Board also confirmed it would meet twice a year, with provisions for extraordinary meetings if necessary.

During its first meeting, the EAB expressed general satisfaction with the project's progress on ethical and regulatory matters, while recognizing the complexity of its data handling framework. In response to questions about the status of ethical approvals, WP2 clarified that for retrospective studies, some sites were covered under existing approvals, while others were in the process of seeking new or amended approvals. For prospective studies, new consents and ethical clearance would be required. The EAB endorsed this approach but emphasized the need for a formal and comprehensive method of tracking compliance and ethical oversight across all sites and study types.

To this end, the EAB recommended a detailed mapping exercise to document the ethical and data protection status of each participating site. This should incorporate insights from the DPIA, the DMP, data dictionaries, and the full data processing pipeline (whether centralized or decentralized). This mapping would provide clarity on compliance requirements and help sites perform self-assessments, even accounting for language differences or local approval timelines. In addition, the EAB supported the establishment of unified ethical and legal principles for the project. This would aid internal coordination and foster alignment with external collaborators on key issues such as data protection, equity, and responsible AI development.

The Board also addressed concerns about representation and bias in datasets, particularly regarding disease severity and demographic diversity. Given the complexity of these issues, the EAB recommended the use of European Commission toolkits for AI risk assessment to help the project identify, evaluate, and either mitigate or transparently acknowledge such risks in its outcomes.

**5. Conclusions**

The inaugural meeting of the EAB led to the identification of key actions aimed at strengthening the ethical and regulatory integrity of the GenoMed4All project. The immediate priority is the initiation of a comprehensive mapping exercise, leveraging existing materials to systematically document ethical and data protection compliance across all sites. This work will also inform the formulation of a set of project-wide ethical principles, which will support the integration of AI risk assessment into the current Data Protection by Design and Default strategy implemented by WP2.

In parallel, the project will need to address the establishment of contractual agreements among both internal and external partners. This process may be complicated by varying ethical approval statuses, and in such instances, WP2 and its collaborators may need to offer guidance or propose phased approaches to ensure all partners can proceed with confidence and clarity.

The EAB will be updated on the progress of these activities through a formal report within the next three to four months.

### Supplementary Information 9| Deliverable 2.3: Genomed4All Data Protection Safeguards, Code of Practice and Data Sharing Agreements

**1. Introduction**

D2.3 outlines the data protection compliance strategy for the GenoMed4All project, detailing the contractual and documentary framework that underpins its regulatory and data protection safeguards. Developed in alignment with the project's Data Protection by Design and Default methodology, the deliverable complements earlier reports (D2.1, D2.2) by offering a cohesive view of data protection, research governance, and ethical oversight, supported by the independent EAB.

Since the project's inception, a continuous process of stakeholder engagement and technical assessment has guided the establishment of data protection measures. The DPIA and the DMP have served as the foundation for defining roles and responsibilities, shaping codes of practice, and drafting necessary data sharing and processing agreements. This collaborative process has ensured that all partners understand their obligations and the applicable regulatory frameworks.

As a result, a comprehensive set of governance tools has been created, including a Code of Practice for all partners, Joint Controller Agreements tailored to each use case, Data Processing Agreements for controller partners and the software provider, security specifications, and an Associate Member Agreement for third-party collaborators. These materials collectively ensure that data protection and security standards are upheld throughout the project’s lifecycle.

The structure of the deliverable provides a detailed account of these safeguards, with specific sections dedicated to their development, content, and application, along with concluding guidance on their future oversight.

**2. Data Protection Safeguards Overview**

Section 3 of D2.3 outlines the comprehensive approach GenoMed4All has taken to define and implement data protection safeguards. Central to this effort was a clear delineation of the nature of data processing and an accurate assignment of regulatory roles and responsibilities across the consortium. These determinations directly informed the drafting of contracts, codes of practice, and supporting documentation, including consent forms and participant information materials.

Given the scale and complexity of the consortium, GenoMed4All initially explored a dual model of data sharing but later opted for a FL framework. This approach allows for local data processing while centrally validating only the trained algorithms, thereby strengthening privacy and ethical compliance. The shift to this model required careful adaptation of legal and governance structures in line with the GDPR.

All partners, except the technical provider DEDALUS, were designated as Joint Controllers under the GDPR, with DEDALUS assuming the role of sole Data Processor. DEDALUS, in turn, appointed CINECA as Sub-Processor to deliver the necessary infrastructure. The consortium's data protection framework is built around several key artefacts, including a General Code of Practice that sets out individual and shared responsibilities, Joint Controller Agreements (JCAs) based on a neutral European Commission-endorsed template, and a centralised Data Processing Agreement (DPA) between UPM and DEDALUS on behalf of all partners.

In addition, the project developed detailed Codes of Practice for security management and drafted an Associate Member Agreement to govern third-party participation. This ensures that external contributors are subject to the same responsibilities and standards as core consortium members. Together, these measures provide a robust, compliant, and coordinated framework for safeguarding personal data and maintaining regulatory integrity throughout the project's execution.

#### 3. Code of Practice: Project-Wide Practice and Processes

The Code of Practice for GenoMed4All is designed to align with the roles established in the JCAs, specifically distinguishing between Data Sources, Data Recipients, and infrastructure providers acting as Processors or Sub-Processors. Within the consortium, DEDALUS serves as the sole Data Processor and contracts with CINECA as its Sub-Processor. This Code is intended to be read in conjunction with the JCAs, the project-wide and local DPIAs, and any other relevant codes or risk assessments established across the project.

The Code reaffirms that each consortium partner retains full responsibility for their individual data protection obligations under GDPR, as GenoMed4All itself is not a legal entity and cannot assume these roles. All partners must comply with their designated responsibilities, adhere to the JCAs, and ensure alignment with infrastructure and security specifications, as well as all applicable DPIAs. Each Data Source and Recipient is required to conduct a DPIA for their activities or provide a justified exception, and must inform the consortium, via i~HD, of any changes or necessary amendments to the generic DPIA.

Furthermore, all partners are obligated to meet local legal and ethical requirements for data processing, including securing necessary ethics committee approvals, with GenoMed4All maintaining a central record but not bearing responsibility for compliance. Staff training is also emphasized; partners must ensure their teams are properly trained and aware of both project-specific and institutional data protection requirements.

In the event of a data breach, all parties must follow their institutional reporting protocols and inform GenoMed4All accordingly, although the project will not initiate breach responses on behalf of institutions. Finally, all participants, particularly those involved in algorithm development, must limit their use of personal data to what is strictly necessary for the completion of their tasks.

#### 4. Joint Controller Agreement Main Text

This document outlines the JCA governing data protection responsibilities among the participating parties in the GenoMed4All programme. It establishes the framework by which these parties – collectively referred to as

"Joint Controllers" – jointly determine and manage the processing of personal data under Regulation (EU) 2016/679 (GDPR). The agreement supplements the Consortium and Grant Agreements that underpin GenoMed4All, a collaborative research initiative focused on SCD and the training of AI models for clinical support. The main body of the agreement standardizes terms and responsibilities across the consortium, regardless of individual party roles.

The JCA explicitly defines key terms, outlines the scope of collaboration, and details each party's obligations with regard to transparency, data protection, and GDPR compliance. It mandates clear division of responsibilities for ensuring data security, responding to data subject requests, and managing data breaches. Each Joint Controller must maintain adequate technical and organizational safeguards, cooperate in audits, and ensure third-country transfers comply with GDPR via approved mechanisms such as Standard Contractual Clauses.

The agreement also regulates the use of processors, specifically naming DH HEALTHCARE PROVIDER SOFTWARE SPAIN S.L.U. (DEDALUS) and its subcontractor CINECA, and defines how additional processors may be engaged. Joint Controllers are liable for direct damages arising from GDPR breaches and must adhere to defined procedures for audits, amendments, and the accession of new parties. The agreement remains in effect throughout the cooperation and any subsequent period in which obligations persist.

In conclusion, this JCA serves to ensure a unified and compliant approach to the processing of personal data across the GenoMed4All collaboration, safeguarding data subjects' rights and ensuring accountability among all participants.

### 5. Security Management Configuration

The JCA outlines a mandatory code of practice concerning information security and confidentiality that all parties must strictly observe. It requires that, where feasible, data be anonymised or pseudonymised in alignment with local approvals (while recognising that full anonymisation may not be possible for genetic data). Each party must share their data handling methodologies upon request, ensuring that data is sufficiently de-identified before sharing. Robust organisational and technical safeguards must be in place to protect data confidentiality, including the use of industry-standard encryption for any data transmission and regular monitoring, testing, and auditing of systems to support continuous improvement.

Personnel involved in the cooperation must be properly trained in data protection and made aware of the sensitivity of shared data. A comprehensive training program should be maintained and tracked to ensure compliance.

From a systems standpoint, only up-to-date and supported hardware and software may be used, and suppliers must adhere to recognised industry standards. Parties are also required to have contingency plans in place, including the ability to restore access in the event of an incident, routine data backups stored securely, and effective malware and virus protection measures that are regularly updated.

Access to data must be strictly controlled and granted only as necessary to fulfil the aims of the cooperation, by approved consents. Access must be restricted to authorised personnel only, including those responsible for maintaining systems. At a minimum, password-protected access must be enforced, and user privileges within systems must be actively managed.

### 6. Conclusions

Implementing effective data protection safeguards within GenoMed4All requires navigating a complex framework of contractual agreements, codes of practice, and technical specifications. Like many large-scale initiatives, GenoMed4All faces heightened challenges due to the consortium's size and the nature of the high-risk data processing involved. To address this, a strict adherence to the principles of Data Protection by

Design and Default has been essential, ensuring that partner relationships, processing activities, and system architectures are clearly defined to support the development of appropriate safeguards.

These protections must also align with broader regulatory obligations, including varying interpretations of the GDPR across Member States and the ethical and governance standards imposed by local institutions. Consequently, the project has integrated oversight from individual partner regulatory teams, EAB, and site-specific IRBs to form a cohesive and compliant approach.

While no data protection or governance system can claim perfection, GenoMed4All has established a robust framework for compliance. The effectiveness of this framework lies in its ongoing application, particularly through continuous collaboration with partners and proactive risk assessment mechanisms such as the DPIA. These tools must be reviewed regularly, ideally every six months, or sooner in response to significant changes or breaches.

The adoption of a FL model introduces an added layer of protection but does not eliminate the need for the established safeguards. As JCAs are finalized, the emphasis must remain on a regulatory compliance model that is not only rigorous but also educative, empowering researchers, data providers, and data subjects through clear, informed engagement.

### Supplementary Information 10| Deliverable 2.4: 2nd Report of the Ethics Advisory Board

#### 1. Introduction

D2.4 presents the second report from the GenoMed4All EAB, which comprises three independent experts specializing in advanced health data technologies to enhance outcomes and drive digital innovation. The EAB's role is to provide an impartial assessment of the project's ethical rigor, focusing on data protection and ethical AI development across consortium partners. This report builds on the initial findings presented in D2.2.

Since the EAB's first meeting in January 2022, GenoMed4All has prioritized establishing a comprehensive data sharing agreement among partners, reflecting careful review of each partner's role. Regulatory changes at the Member State level influenced governance adaptations, leading the project to adopt a FL approach for both project phases to address security and ethical concerns related to centralized cross-border data sharing. Additionally, the project has aligned its strategies with evolving frameworks such as the Data Governance and AI Acts and the European Health Data Space.

Progress has been made on key action points identified in the prior EAB meeting, including mapping ethical approvals and data processes across partners, developing core ethical principles to strengthen internal and external collaborations, and applying European Commission toolkits to assess AI-related risks specific to GenoMed4All's context.

The report outlines personnel updates and regulatory compliance developments since the last meeting, provides updates on ongoing action items, and concludes with proposed next steps and scheduling for the subsequent EAB meeting.

#### 2. Matters Arising Since the First EAB Meeting

This section outlines several project-specific and broader regulatory developments impacting GenoMed4All. Since the first EAB meeting, there have been personnel changes, including new appointments and the planned replacement of an EAB member. The current EAB comprises both internal project leads and external experts. Due to scheduling conflicts, updates were communicated individually rather than through a plenary meeting.

In terms of GDPR compliance, the project has clarified partner roles, designating most as Joint Controllers, with infrastructure and software providers acting as Data Processors or Sub-Processors. JCAs, aligned with European

Commission templates, have been developed for each use case and are nearing signature, with adjustments reflecting the project's shift to a FL approach.

Technically, the consortium transitioned from sharing anonymized or pseudonymized data toward FL, thereby enhancing participant protection. This approach keeps data at provider sites, with algorithm training occurring locally, which addresses several regulatory and ethical concerns while enabling continuous algorithm development and validation throughout 2023.

At the European regulatory level, recent and forthcoming legislation (including the Data Governance Act, Data Act, AI Act, and European Health Data Space) introduces new frameworks for data sharing, AI safety, and health data interoperability. These regulations, together with evolving GDPR interpretations and stricter provisions for health data sharing, have influenced GenoMed4All's governance choices, notably the adoption of FL. This shift has necessitated more detailed JCA negotiations and reframing of ethical principles to align with the fully FL model.

#### 3. Update on Action Items from Last EAB

This section reviews the three primary action items from the January 2022 Ethical Advisory Board meeting, noting that priorities have shifted toward addressing GDPR compliance through the development of JCAs, influenced by changes to the FL approach and evolving regulations. The first action item involved creating a comprehensive ethics and governance mapping, which serves to document each partner's responsibilities and data activities for regulatory oversight, particularly in anticipation of the forthcoming AI Act's certification requirements for safety-critical health systems. The second action item focused on engaging partners to develop core ethical principles to support agreements within the consortium and with external parties. This engagement, driven by GDPR compliance and ethical approval considerations, led to the decision to avoid data sharing by conducting algorithm training locally at data provider sites, minimizing risks to participant rights and addressing regulatory concerns. From these discussions, several ethical recommendations emerged, emphasizing minimization of data transfers, cautious use of synthetic data, and strict adherence to consent and independent review board oversight when identifiable data is involved. The final action item involves applying European Commission risk assessment toolkits related to AI development; however, given the shift to FL, these assessments are planned for implementation once pilot work is underway, likely by mid-2023, though the toolkits remain in draft form.

#### 4. Conclusions

Since the January 2022 EAB meeting, GenoMed4All has made significant progress in defining its data flows and strengthening its commitment to ethical standards in response to evolving regulatory and societal expectations. The project has adapted by avoiding the central sharing of personal or de-identified data, instead conducting algorithm training locally at data provider sites and sharing only the trained models for validation and testing. This strategy aligns with regulatory requirements and participant consents, enhancing data protection without compromising research quality. Three core ethical principles have been proposed for review by project partners, with the next EAB meeting scheduled for June 2023.

### Supplementary Information 11| Deliverable 2.5: Genomed4All Recommendations on the Ethical Principles, Quality Processes and Stakeholder Engagements for AI Development

#### 1. Introduction

D2.5 outlines the ethical principles and stakeholder engagement framework for GenoMed4All and similar AI-driven initiatives focused on managing and assessing risks in rare hematological diseases. These principles are grounded in the European Commission's Ethics Guidelines for Trustworthy AI, which define seven key requirements to ensure AI trustworthiness, supported by the Assessment List for Trustworthy AI (ALTAI) as a measurement tool. The deliverable presents GenoMed4All's evaluation against the ALTAI, reflecting on the project's operation since 2021 and the ethical framework developed as a result.

The report details the approach taken to integrate the ALTAI requirements into the project, highlighting key stakeholders involved, including clinical researchers, infrastructure operators, algorithm developers, project managers, regulators, and patients. Each group's roles and priorities are discussed, emphasizing their contributions to maintaining transparency, robustness, privacy, accountability, and fairness throughout the project. Particular attention is given to ensuring that AI interventions align with regulatory expectations, ethical standards, and the needs of end-users.

The deliverable also assesses GenoMed4All's adherence to the seven ALTAI requirements, human agency and oversight, technical robustness and safety, privacy and data governance, transparency, diversity and fairness, environmental and societal well-being, and accountability, interpreting their relevance and application within the project. It concludes with recommendations for the wider adoption of these principles, reflections on the challenges of AI development in health research, and considerations for compliance with forthcoming AI regulations.

### 2. Exploring the Key Requirements, Ethical Principles and ALTAI Assessment

This section examines the seven key requirements of the ALTAI framework, as outlined in the Ethics Guidelines for Trustworthy Artificial Intelligence, and how GenoMed4All has addressed each in its development. It details the implications for the project and prepares for the ALTAI toolkit responses as well as the ethical principles derived.

Human agency and oversight are fundamental, ensuring that research participants provide informed, voluntary consent and maintain control over the use of their data, while clinical professionals retain interpretative authority over AI-generated insights. GenoMed4All emphasizes transparency and clinical oversight by avoiding opaque "black box" models, allowing algorithm outputs to be scrutinized and questioned. This principle is critical as the project scales to integrate large omics datasets via FL, requiring developers and infrastructure operators to maintain a clear understanding of data usage and algorithm training. Maintaining human control throughout data acquisition, processing, and interpretation is essential for ethical compliance and regulatory acceptance, with ongoing challenges to preserve this balance as the AI tools enter practice and research.

Technical robustness and safety are addressed through the development of a complex, interoperable FL infrastructure, prioritizing data security by minimizing centralization. GenoMed4All has documented its architecture and risk mitigation strategies to ensure safe algorithm training. Given the sensitivity of health data, accuracy and quality assurance at multiple points, data provision, algorithm training, and processing, are paramount. The system's design balances robustness with operational efficiency, considering redundancy and backup capabilities, and sets the foundation for meeting ALTAI's standards for safety and reliability.

Privacy and governance underpin the project's approach, guided by GDPR and data protection by design principles. GenoMed4All employs a FL model, where data remains within providers' control, limiting exposure and reducing privacy risks. This model facilitates localized security management and governance while enabling algorithm development through remote access to statistical models. JCAs clearly define partner responsibilities, ensuring all parties understand their roles in lawful data processing. The governance framework emphasizes early stakeholder engagement to streamline contract negotiation and uphold ethical standards throughout data handling.

Transparency is vital for ethical and legal compliance, especially under the emerging AI Act, which mandates clear communication about data use and AI processes. GenoMed4All stresses the importance of informed consent overseen by independent ethics committees, though it acknowledges variability across committees and jurisdictions. The project has developed and shared template materials to promote consistent, understandable transparency for participants and stakeholders, supporting both research ethics and regulatory compliance.

Diversity, non-discrimination, and fairness are critical given the project's focus on rare diseases with limited data availability. GenoMed4All aims to mitigate inequities by leveraging FL and synthetic data to enhance model training and access. Attention is paid to ensuring that ethical concerns do not inadvertently restrict access to emerging interventions for disadvantaged patient groups. The project also recognizes the potential for inconsistencies in ethics review to affect fairness, highlighting the need for careful advocacy and consistent oversight.

Environmental and societal well-being considerations primarily focus on the resource demands of high-performance computing infrastructure, acknowledging the challenges posed by energy consumption and carbon impact. While GenoMed4All has not yet quantified its environmental footprint, it anticipates adopting safety-critical standards similar to those used in clinical trials upon deployment. Societally, the project supports the augmentation rather than replacement of human expertise, reinforcing the value of human oversight in AI-assisted healthcare.

Finally, accountability is established through a governance framework aligned with GDPR and medical device regulatory requirements, providing clear traceability and third-party auditing. This foundation enables GenoMed4All to demonstrate compliance with ALTAI's accountability criteria and supports the project's overall ethical and regulatory integrity.

### 3. ALTAI Assessment Framework Responses

Section 3 presents a comprehensive summary of the ALTAI Assessment results, detailing their implications for the GenoMed4All project, supported by supplementary PDFs and embedded figures. The ALTAI Toolkit serves as a self-assessment framework, guiding the identification of areas requiring further development and validation, particularly in preparation for clinical trials.

In addressing Requirement 1 (Human Agency and Oversight), GenoMed4All has taken measures to prevent overreliance on AI and ensure meaningful human intervention, although limitations exist due to the absence of product trialing within the project's scope. The deliverable recommends clearly defining AI deployment in trials to evaluate behavioral impacts and support future compliance with health technology and AI regulations.

Requirement 2 (Technical Robustness and Safety) revealed significant risks, but the collaborative risk assessment and the establishment of a Joint Controller Agreement have laid a foundation for managing security and compliance. The report advises commissioning audits, including penetration testing and resilience reviews, to validate security measures and ensure data quality and transparency in algorithm development.

For Requirement 3 (Privacy and Governance), the project demonstrates strong adherence to privacy rights and data protection principles, supported by independent ethical reviews and governance aligned with GDPR. Consent management and data lifecycle protection are embedded within established frameworks and standards.

Requirement 4 (Transparency) emphasizes the importance of traceability, explainability, and clear communication with both users and data subjects. Transparency is closely linked with ethical oversight to ensure meaningful participant engagement and informed consent throughout AI tool development.

Regarding Requirement 5 (Diversity, Non-Discrimination, and Fairness), GenoMed4All acknowledges the challenges posed by limited data availability for rare diseases. The assessment highlights the need for further evaluation within clinical trials to better understand and mitigate risks related to bias and fairness, with a strong focus on representative data and stakeholder involvement.

Requirement 6 (Environmental and Societal Well-Being) presents difficulties in collectively assessing environmental impacts due to the project's multi-partner structure, suggesting the need for a dedicated follow-up effort. Societal impacts have been scoped within the project's health innovation framework, but further analysis during interventional trials is recommended.

Finally, Requirement 7 (Accountability) is underpinned by robust ethical review processes and risk management embedded across project WPs. The framework relies on established mechanisms such as GDPR's Data Protection by Design, research governance, and regulatory compliance to ensure clear accountability for the development and operation of AI components.

Overall, the ALTAI assessment provides a strategic roadmap to guide GenoMed4All's ongoing development, emphasizing the integration of ethical, technical, and regulatory considerations to advance AI-driven health solutions responsibly and compliantly.

### 4. Ethical Principles, Quality Processes, Stakeholder Engagements and Discussion

This section outlines the recommended ethical principles aligned with the seven key ALTAI requirements, followed by a review of stakeholder engagement strategies and quality assurance recommendations, concluding with a discussion on the limitations of the approach and implications of the forthcoming AI Act. Ethical guidance emphasizes early identification and clear understanding of stakeholder agency and oversight expectations, transparent and explainable technical robustness, stringent privacy and governance commitments, and meaningful transparency that supports informed consent and equitable participation. The principles further address fairness and non-discrimination by highlighting the need to carefully balance risks in vulnerable populations, as well as environmental and societal well-being considerations through responsible procurement and trial design. Accountability is underscored by the necessity for AI components to meet established standards equivalent to those in data use and research conduct.

Stakeholder engagement is tailored according to roles, stressing continuous involvement of clinical researchers in development and validation, ensuring infrastructure operators maintain robust and secure systems, and promoting transparency and traceability among technical experts. Programme managers must oversee progress and ethical compliance, regulators require ongoing updates on development and oversight, and patients and the wider citizenry must be meaningfully informed and engaged given their direct interest and contributions.

The discussion acknowledges the value of ALTAI as a comprehensive, consultative ethical framework that supports risk management and operationalization of AI ethics within GenoMed4All. However, it also notes the primary limitation of relying on self-assessment rather than independent certification, which the AI Act is expected to mandate for high-risk AI systems. Consequently, this report is positioned as a foundational step toward compliance, recommending that GenoMed4All pursue formal assurance frameworks and incorporate these evaluations into forthcoming pre-clinical trials. Only through such real-world testing can concerns related to agency, oversight, transparency, and fairness be conclusively addressed, ensuring the ethical deployment and sustained governance of AI-driven healthcare solutions.

### 5. Conclusions

The application of AI in health data innovation remains in its early stages, particularly regarding its impact and practical use in clinical interventions. Amid ongoing efforts across the EU and EEA to empower citizens with greater control over their personal data, including health information, there are growing concerns that such control is increasingly being transferred to complex algorithmic systems and the entities controlling these technologies. In this context, projects like GenoMed4All are critically important. They uphold rigorous standards of research integrity, participant engagement, and clinical decision-making while providing valuable insights into ensuring that AI-driven solutions comply with both current and forthcoming regulatory frameworks. This deliverable captures GenoMed4All's experiences in governing early AI interventions to inform future innovation efforts and to support equitable and effective healthcare advancements. The next phase involves operationalizing these insights through carefully designed trials that address the ethical and practical challenges identified, following the ALTAI assessment framework.

### Supplementary Information 12| Deliverable 2.6: 3rd Report of the Ethics Advisory Board

#### 1. Introduction

D2.6 represents the third report from the GenoMed4All EAB, which comprises three independent experts specializing in advanced health data technologies aimed at improving outcomes and driving digital innovation. The EAB's primary role is to provide an independent assessment of the project's ethical integrity, with particular emphasis on data protection and the ethical development of AI across consortium partners. This report reflects on recent updates and developments regarding ethical and regulatory requirements for GenoMed4All, aligned with the EAB's mandate outlined in D2.2. Since their November 2022 meetings, GenoMed4All has finalized and executed three JCAs covering the project's use cases, with all partners' approval, as detailed in D2.3. The EAB has also monitored evolving regulatory frameworks, including the Data Governance Act, AI Act, and European Health Data Space (EHDS) regulations. Progress on establishing core ethical principles, quality assurance processes, and stakeholder engagement recommendations, as outlined in D2.5, has been shared with and reviewed by the EAB. The report is structured to present recent project developments, updates on previous action items, and planned next steps, concluding with scheduling for the forthcoming EAB meeting.

#### 2. Developments Since the November 2022 EAB Meetings and Activities for 2024

This section outlines key project-related and broader European regulatory developments impacting GenoMed4All. The EAB underwent changed in its membership, with Julie Power replacing Mahsa Shabani and Jorge Alfonso Kurano joining as Programme Manager. The EAB now includes a mix of internal project leads and independent experts, who convened for their third meeting in November 2023 to review updates. The JCAs for the project's three use cases were finalized, signed, and executed in early 2023, clearly defining data controller and processor roles. These agreements, alongside DPIA, will be routinely monitored to ensure ongoing compliance. Ethical principles, quality processes, and stakeholder engagement strategies have been developed in alignment with these agreements and the European Commission's Ethics Guidelines for Trustworthy AI. The EAB endorsed the use of the ALTAI to evaluate adherence to key requirements such as human oversight, privacy, transparency, fairness, and accountability. Due to delays in the assessment toolkit, the project team proposed leveraging existing principles and stakeholder input to draft detailed ethical guidelines, with a review scheduled for February 2024. On the regulatory front, the enactment of the Data Governance Act, revisions to the AI Act, and developments around the EHDS signal significant compliance requirements for high-risk AI interventions, including certification, data integrity, and cataloguing. GenoMed4All's current ethical framework and ALTAI-based approach position it well to meet these emerging regulatory standards.

#### 3. Conclusions

Throughout 2023, GenoMed4All has advanced amid evolving regulatory frameworks and shifting societal expectations concerning AI use and the reuse of health and personal data. The adoption of a FL model, alongside the finalization of JCAs, has strengthened the project's foundation as its architecture is completed and data processing begins. This report to the EAB outlines the project's current progress and establishes the framework for ethical principles, quality processes, and stakeholder engagement. It also details the approach taken to navigate regulatory changes, ensuring that GenoMed4All remains both compliant and trustworthy to the public. The EAB is scheduled to review the draft of D2.5 in mid-February 2024 and will convene again in the fourth quarter of 2024.

### Supplementary Information 13| Deliverable 2.7: Validated Recommendations on the Trustworthy Development of AI for Personalized Medicine

#### 1. Introduction

This deliverable assesses AI development and validation within GenoMed4All, focusing on WP6's creation of AI tools for analyzing rare hematological disease data and WP7's verification of data sharing and AI use across clinical trials. Its purpose is to provide recommendations and guidance to consortium partners and the wider research and healthcare communities for building conformity assessments that support adherence to the European Commission's Ethics Guidelines for Trustworthy AI, as well as related regulatory frameworks such as the ALTAI, the AI Act, and the Medical Device Regulation (MDR). The current healthcare AI landscape remains uncertain, with regulatory enforcement expected to increase gradually over the next few years while standards are still under development. This evolving environment demands adaptability in validation approaches by both regulators and innovators. Accordingly, the deliverable also offers supplementary recommendations based on international standards to guide AI applications in clinical research. The experiences and validations from GenoMed4All serve as a model for broader healthcare AI transformation, demonstrating the benefits of AI-driven innovation in practical medical contexts. The ethical assessment methodology ensures that AI systems are lawful, ethical, and robust, relying on an ALTAI self-assessment initially reported in D2.5 and further refined for compliance with the AI Act. This methodology builds on GDPR-compliant data governance and ethical standards, acknowledging limitations such as inherent biases due to patient population constraints, which are addressed through explainability and ongoing verification of algorithmic outputs. The deliverable comprehensively reviews the AI technologies developed, the ethical frameworks applied, the alignment with ALTAI and AI Act requirements, and evaluates GenoMed4All's compliance and remaining challenges, providing a rigorous foundation for trustworthy AI both within the project and for future initiatives.

### 2. AI technologies in GenoMed4All

GenoMed4All advances AI applications in healthcare by integrating clinical, genomic, and imaging data through a secure, privacy-preserving FL platform to develop and validate AI tools for personalized medicine in hematological diseases. The project aims to demonstrate the potential of trustworthy and explainable AI by combining deep learning, generative models, and advanced statistical methods to generate new insights and support clinical decision-making while ensuring full compliance with data protection laws and ethical standards.

The project focuses on three hematological conditions. SCD, a hereditary chronic disorder, presents diverse phenotypes that the project addresses by developing AI models to classify patients by inflammatory and hemolytic risk, predict complications such as microalbuminuria, and identify genetic markers associated with disease progression. For MM, a malignant plasma cell cancer, GenoMed4All integrates extensive genomic and clinical data to improve patient stratification, predict early relapse risk, and personalize treatment, addressing the limitations of existing prognostic scores. In MDS, a heterogeneous blood cancer primarily affecting older adults, the project applies AI to refine disease classification by linking clinical, morphological, and genomic data and to develop prognostic models predicting survival and leukemic progression. Various machine learning and statistical methods have been employed to enhance disease understanding and outcome prediction, emphasizing model robustness and explainability.

Overall, GenoMed4All represents a significant effort to harness AI for precision medicine in hematology, addressing unmet clinical needs through innovative data integration and analysis while upholding ethical and regulatory standards.

GenoMed4All has developed a comprehensive suite of AI-driven algorithms to address critical clinical unmet needs. These include advanced pipelines for preprocessing and preparing diverse data types such as clinical records, whole genome sequencing (WGS), PET/CT, whole-body low-dose CT (WBLDCT), and MRI imaging, ensuring standardized, high-quality inputs for subsequent analysis. The project also implemented sophisticated clustering techniques for patient stratification and early relapse prediction, leveraging hierarchical Bayesian models, dimensionality reduction, and unsupervised machine learning based on genomic and radiomic profiles.

Further, differential gene expression and pathway enrichment analyses were performed to elucidate biological mechanisms underpinning disease progression. Survival analysis models were developed using multimodal data integration, including variational autoencoders combined with deep survival models, which have been validated on large patient cohorts and outperform traditional methods. Radiomics-based survival prediction and genomic cluster survival assessments complement these efforts, enhancing prognostic precision.

Data integration is a key focus, with Bayesian frameworks such as Multi-Omics Factor Analysis (MOFA) and Hierarchical Dirichlet Mixture Models (HDMM) enabling the fusion of clinical, genomic, and imaging data to uncover latent biological patterns and improve patient stratification. Additionally, machine learning methods were developed to predict cytogenetic profiles from imaging features, addressing challenges of limited mutation prevalence. A multimodal generative model was designed to synthesize heterogeneous medical data, overcoming the constraints of single-modality AI approaches.

In the context of SCD, fully automated pipelines employing deep learning ensembles facilitate the segmentation and identification of silent cerebral infarctions from MRI scans, incorporating transfer learning and specialized loss functions to handle small lesion volumes effectively.

Collectively, these algorithms form an integrated platform that supports personalized medicine by enabling robust data processing, clustering, comparative genomics, survival prediction, multimodal data integration, synthetic data generation, and precise image analysis, thus advancing clinical decision-making in hematological malignancies and SCD.

### 3. Ethical and Legal Frameworks for AI in the EU

As AI increasingly impacts healthcare and interventional medicine, ethical considerations are critical to ensuring trust, safety, and regulatory compliance. The European Union has proactively established comprehensive ethical frameworks to guide the responsible development and deployment of AI in healthcare and clinical research. Central to these frameworks are principles such as human agency, transparency, fairness, and accountability, supported by regulatory tools like the GDPR, the Ethics Guidelines for Trustworthy AI, the EU AI Act, and CEN-CENELEC standards.

The GDPR, effective since 2016, provides a foundational legal framework for data privacy, emphasizing lawful, fair, and transparent processing of personal data, purpose limitation, data minimization, accuracy, storage limitation, security, and accountability. Within the GenoMed4All project, GDPR compliance was ensured through careful assessment of data flows, legal responsibilities, and formalized agreements among partners, including the conduct of a Data Protection Impact Assessment.

The Ethics Guidelines for Trustworthy AI, developed by the European Commission's High-Level Expert Group, frame AI governance around the pillars of lawfulness, ethics, and technical robustness. These guidelines define seven key requirements, ranging from human agency and oversight to accountability and societal well-being, that aim to ensure AI systems are designed and implemented to enhance human capabilities, protect rights, and operate reliably and fairly. GenoMed4All aligned its principles with these requirements, using tools such as ALTAI to self-evaluate compliance and integrate ethical considerations into system development.

The AI Act, set to come fully into force in August 2024, is the world's most comprehensive AI regulation. It introduces a risk-based classification framework, identifying high-risk AI systems (such as clinical decision support tools) that require strict conformity assessments prior to deployment. The Act mandates human oversight, transparency, data quality, and post-market monitoring, emphasizing patient safety and fundamental rights protection. Though GenoMed4All's research phase is exempt, compliance with the AI Act will be essential as the project moves toward market application. The Act also supports innovation through regulatory sandboxes, allowing controlled testing environments under relaxed rules.

Finally, the CEN-CENELEC standards for AI contribute technical harmonization aligned with EU values and legislative frameworks, complementing international standards such as ISO/IEC 42001. While adherence to these standards alone does not guarantee AI Act compliance, they provide valuable guidance to streamline conformity efforts and align AI technologies with European regulatory expectations.

Together, these ethical guidelines, regulations, and standards form a cohesive governance ecosystem that promotes the responsible, transparent, and safe use of AI in healthcare, reinforcing public trust and safeguarding fundamental rights amid technological advancement.

### 4. Ethical AI Requirements for the Genomed4All

The integration of AI within GenoMed4All has been guided by a comprehensive set of frameworks and standards that balance innovation with ethical responsibility. This section highlights the core ethical requirements drawn from the ALTAI and relevant international standards, focusing on transparency, accountability, and safety to ensure that AI applications remain compliant, equitable, and centered on patient well-being. These guidelines offer a clear roadmap for navigating the evolving regulatory environment and implementing AI solutions that meet the highest ethical and operational standards. Key principles emphasize the importance of recognizing and respecting human agency and oversight among all stakeholders, ensuring detailed documentation and technical robustness to safeguard system reliability, and establishing strong privacy and data governance practices with clear roles and responsibilities. Transparency is prioritized through independent ethical review and meaningful communication with patients and participants, fostering trust and informed engagement. Additionally, the guidelines stress the need to address diversity and fairness, avoiding discrimination while maintaining equitable access. Consideration of environmental and societal impacts guides the responsible development and deployment of AI tools, particularly in clinical contexts. Finally, the framework underscores the necessity of accountability mechanisms that hold AI components to rigorous standards equivalent to those governing data use and research conduct. Together, these principles underpin GenoMed4All's commitment to ethical, safe, and patient-focused AI innovation.

This section builds on the ALTAI self-assessment recommendations by providing supplementary guidance aligned with the latest provisions of the AI Act 2024, reflecting an enhanced understanding of its ethical and regulatory demands. The AI Act imposes rigorous compliance requirements, particularly for high-risk AI systems, encouraging developers to adopt a comprehensive suite of European and international standards to ensure responsible and trustworthy AI deployment. Emphasizing human oversight and agency, the Act mandates that AI systems support human decision-making, allow interpretability and user control, and incorporate mechanisms for verification and protection of fundamental rights. Technical robustness, safety, accuracy, and cybersecurity are prioritized through requirements for resilient system design, rigorous testing, fail-safe mechanisms, and defenses against adversarial threats. Privacy and data governance principles demand well-managed, representative, and bias-mitigated datasets, with stringent safeguards for sensitive data. Transparency and user information obligations require clear explanations of AI logic, comprehensive documentation, and guidance on human oversight, ensuring that users understand system capabilities, limitations, and operational requirements. Finally, the Act stresses the importance of diversity, non-discrimination, and fairness, calling for inclusive design, early bias prevention, and ethical practices to avoid exploitation or unfair competitive behaviors. Collectively, these enhanced recommendations offer a robust framework to ensure AI systems within GenoMed4All meet the highest standards of trustworthiness, safety, and ethical integrity under the evolving regulatory landscape.

The sections emphasize that AI systems must be designed and managed with ethical, sustainable, and democratic values, prioritizing societal well-being and environmental responsibility while considering future generations. High-risk AI systems require robust accountability and risk management frameworks, including continuous risk identification, mitigation, testing, and transparency, with special attention to vulnerable groups. Rigorous technical documentation, comprehensive record keeping, and conformity assessments are essential to demonstrate compliance and maintain trust. To support these requirements, a broad set of international standards covering human oversight, robustness, safety, privacy, transparency, fairness, environmental impact, and governance are recommended. These standards provide detailed guidance on lifecycle processes, risk management, data quality, cybersecurity, and inclusivity, ensuring AI systems meet legal obligations such as the AI Act, GDPR, and sector-specific regulations, ultimately fostering trustworthy, safe, and socially responsible AI deployment.

The GenoMed4All project has carefully aligned the roles, interests, and responsibilities of its diverse stakeholders with the regulatory and ethical requirements of the EU AI Act, ensuring comprehensive compliance, ethical AI deployment, and enhanced trustworthiness throughout its activities. Clinical researchers and practitioners play a critical role in maintaining human oversight, interpretability, and transparency, ensuring

AI supports rather than replaces clinical judgment for sound, ethical decision-making. Infrastructure operators are responsible for the technical robustness, cybersecurity, and resilience of AI systems, guaranteeing operational reliability. Designers, analysts, and developers focus on transparency, bias mitigation, and validation to ensure accurate, reliable AI outputs. Programme managers and internal oversight teams enforce accountability through risk management, conformity assessments, continuous monitoring, and thorough documentation, maintaining ethical and regulatory alignment. Regulators and commissioners provide ongoing supervision to ensure compliance through conformity assessments and post-market surveillance, reinforcing accountability. Engagement with patients, research participants, and the public is fundamental, promoting transparency, safeguarding rights, and addressing the societal impact of AI in healthcare. Together, these coordinated efforts establish a solid framework ensuring GenoMed4All's AI systems are not only technically sound and compliant but also ethically responsible and socially accountable. Detailed, stakeholder-specific recommendations based on the AI Act and ALTAI framework guide transparency, fairness, and technical rigor, reinforcing the project's commitment to responsible and trustworthy AI use in healthcare.

### 5. Validation of the Recommendations in Genomed4All: Results and Findings

This section validates the supplementary recommendations in light of GenoMed4All's practical experience, demonstrating alignment with the ALTAI framework and the principles set out in the EU AI Act. While GenoMed4All has not deployed its AI algorithms in live clinical settings, all development and testing were conducted in secure, in vitro environments, with a strong emphasis on transparency, accountability, and ethical integrity. The project's activities reflect a robust commitment to safeguarding human oversight, interpretability, and fundamental rights throughout the AI lifecycle.

Crucially, GenoMed4All anticipates that future validation of its algorithms will benefit from AI Regulatory Sandboxes, which provide controlled environments for testing compliance with emerging standards. Although these were not available during the project's development phase, their eventual use is recommended for further validation and certification of the algorithms, ensuring safety, reliability, and alignment with evolving regulatory expectations.

Human oversight remains central in the GenoMed4All framework, with AI outputs designed to support clinical judgment. Transparent output design, dual-review processes, and a clear delineation of algorithmic steps ensure that clinicians and researchers can interpret, trace, and override results as needed. Even in a research setting, all outputs are subject to human scrutiny, allowing for identification and correction of anomalies or biases in algorithmic processing.

Interpretability and control have been addressed through a stepwise development model that enables clear mapping of AI outputs to underlying data and decisions. This framework allows users to intervene at critical points, halting system activity where necessary. In future live deployments, clinicians will receive both AI-generated recommendations and contextual explanations, equipping them to make informed, accountable decisions with full control over outcomes.

Verification of outputs has been embedded through multidisciplinary collaboration, where results were reviewed and validated by qualified professionals. This approach sets a precedent for dual-verification processes in clinical deployment, ensuring that any AI-influenced decision is subject to human review and scrutiny before action is taken. Such safeguards reinforce professional accountability and build trust in the system's reliability.

Finally, GenoMed4All maintains a firm commitment to protecting fundamental rights. Ethical oversight, patient privacy, data governance, and informed consent were embedded in the project's design, with transparent communication practices around data use and algorithmic development. These safeguards ensure that AI is not misused, that over-reliance is avoided, and that patients retain autonomy and agency. By integrating these principles into every stage of the AI lifecycle, GenoMed4All provides a strong foundation for future clinical use that is both ethically responsible and legally compliant.

The GenoMed4All project demonstrates a comprehensive and proactive approach to ensuring the technical robustness, safety, accuracy, and cybersecurity of its AI systems. Though not yet deployed in live clinical settings, the project's in vitro development and validation processes were structured around transparency, fault tolerance, and rigorous quality controls, aiming to build dependable systems capable of maintaining performance across varied conditions and input types. This foundation includes detailed risk mitigation measures, secure data governance practices, and attention to the integrity and resilience of AI outputs, all designed to support future real-world implementation.

Dependability was established through consistent, structured testing across diverse datasets, validating the AI's ability to operate reliably despite data variability or adversarial challenges. GenoMed4All's phased development identified key use case steps and integrated mechanisms to detect anomalies, adapt to changes, and ensure safe, traceable outputs. Future deployment would require that these validations continue within Regulatory Sandboxes and operational environments, where clinicians could monitor outputs, flag unexpected behaviour, and feed insights back into the system for iterative improvement.

The technical design of the platform was built on clear lifecycle stages, covering multi-modal data preparation, clustering, analysis, and output generation. These were guided by benchmarks and measurement methodologies to ensure consistent accuracy and security. Real-world deployment would necessitate ongoing auditing against clinical and technical standards, supported by cybersecurity protocols, real-time performance monitoring, version tracking, and clear impact assessments for any system changes. This would ensure the integrity and reliability of outputs throughout the system's lifecycle.

To address errors and bias, GenoMed4All's development included assessments of performance across underrepresented cohorts and edge cases. The system was designed to be resilient, with early mechanisms for identifying and correcting biased or erroneous outputs. Future implementations would involve real-time bias detection tools, clinician-led feedback loops, and technical safeguards such as fail-safes, model rollbacks, and human-in-the-loop processes, all of which support a fair, adaptive, and safe operational framework.

Protection against external threats was treated with equal priority. GenoMed4All's data and system security protocols were shaped by a multi-institutional, cross-jurisdictional risk management framework rooted in EU data protection principles. These controls emphasized encryption, access control, auditability, and traceability. Full deployment would require continued adherence to these practices, along with robust model integrity checks, adversarial stress testing, and secure training environments. The project also contributes valuable insight into the ongoing development and refinement of AI Regulatory Sandboxes, offering a reference model for secure, compliant, and resilient AI design.

On the matter of data governance, GenoMed4All implemented stringent controls to ensure ethical, legal, and transparent handling of sensitive health and genomic data. Detailed documentation of data provenance, processing steps, and application use cases provides traceability and accountability. Compliance with GDPR was fully integrated into the project's processes, supported by formal data protection tools and participant-facing materials such as informed consent forms and transparency notices. Critical concerns such as data gaps, biases, and representational fairness were not only identified but actively addressed in the system's design. These efforts form a strong basis for future live deployment, where the same governance structures and documentation should inform risk and quality management practices in line with the EU AI Act and local regulatory standards.

The GenoMed4All project has demonstrated a strong commitment to transparency, interpretability, and the provision of clear, accessible information to users throughout the lifecycle of its AI system. Through comprehensive documentation, user-centred design, and adherence to European data protection and research governance standards, the project ensures that both clinical users and stakeholders can understand, monitor, and responsibly engage with AI-generated outputs. These efforts include detailed technical explanations, clear delineation of system purposes and limitations, and explainability features that help bridge the gap between complex algorithms and user comprehension.

While the system has not yet been deployed in live clinical settings, GenoMed4All has proactively addressed ongoing oversight and accountability. Logging mechanisms, structured data storage, and traceable result generation processes have been embedded into the development framework. These provisions lay the foundation for regulatory compliance, responsible innovation, and user trust, although further validation will be essential to optimize these methods for practical, real-world application.

Transparency has been treated as a foundational principle, with the entire AI development process, from data sourcing and training to result generation, carefully documented and auditable. The system has been designed to support traceability of outputs and articulate its limitations clearly, ensuring that users are informed of the contexts in which the AI may underperform or be less reliable. This transparency is essential for fostering ethical, safe, and responsible use, particularly in clinical environments where decision-making must be rigorous and justified.

Explainability has also been central to the project. GenoMed4All ensures that users can interpret AI-generated outputs through insights into the logic, data inputs, and decision-making processes. Even prior to live deployment, the system has been developed to highlight contributing data elements such as specific genomic or clinical markers. In practice, this would be supported by explanatory tools, metadata, and visual aids to enable clinicians to align AI reasoning with their medical judgment, enhancing both utility and trust.

Finally, transparency is supported by a design philosophy rooted in user-centred principles. Interfaces are built to be intuitive and accessible for a range of users, from clinicians to researchers, while still providing the necessary depth for expert scrutiny. System architecture accommodates both traceability and interpretability, with onboarding and training materials tailored to various user groups to promote confident and informed engagement. Through this approach, GenoMed4All not only advances the technical reliability of its AI systems but also ensures they are comprehensible, trustworthy, and viable in real-world clinical settings.

The GenoMed4All project has placed a strong emphasis on ensuring its AI system is supported by clear, accessible, and comprehensive documentation that facilitates responsible deployment and effective user engagement. From the early stages, the project developed internal technical records and training materials to support understanding of how the system operates, including its purpose, capabilities, and limitations. Though not yet in clinical deployment, GenoMed4All has prepared for future real-world implementation by establishing guidelines that explain the system's functionality, accuracy levels, provider credentials, and potential risks. This groundwork will allow for the provision of structured user instructions, integrated into the system interface and available in multiple formats, that guide clinicians and researchers in interpreting AI-generated results and acting on them with confidence and oversight.

Human oversight is a critical part of the GenoMed4All approach. The system has been designed with mechanisms that enable clinicians to monitor AI outputs, identify inconsistencies, and intervene when necessary. This includes planned tools such as explanation panels, confidence scores, and alert indicators, all aimed at supporting real-time interpretation and decision-making. Clear guidance on how to use these tools, when to question outputs, and how to escalate concerns will be integral to ensuring the system supports safe clinical practice while maintaining human authority over healthcare decisions.

Performance management has also been built into the project, with detailed records of the datasets used in training, including their demographic and clinical diversity. By monitoring the system's performance across different patient groups and making these metrics available to users, GenoMed4All ensures that its outputs are reliable, equitable, and subject to continuous improvement. Feedback channels are also envisaged, enabling clinicians to report any anomalies or concerns, feeding directly into iterative model refinement.

Operationally, the system is designed with transparency and sustainability in mind. Early work has identified software and hardware requirements, system maintenance protocols, and logging and audit mechanisms. These technical specifications also support readiness for integration into future Regulatory Sandbox environments and clinical deployments.

Beyond functionality, GenoMed4All is committed to upholding principles of diversity, fairness, and inclusion. From the outset, the project has worked to mitigate bias in its datasets by ensuring broad demographic representation, particularly across the rare disease populations it addresses. This commitment is reinforced by collaborations with synthetic data projects such as SYNTHEMA and SYNTHIA to compensate for gaps in available data. These efforts will be critical in assessing and managing bias in real-world applications, particularly when subject to further scrutiny in Regulatory Sandboxes.

The system's inclusive design is intended to ensure accessibility for users of all backgrounds and technical abilities. By engaging clinicians and domain experts during development, GenoMed4All has prioritised a user-friendly interface and interpretability features suited to a range of needs. Future deployment will be guided by universal design principles, accessibility testing, and multilingual support, enabling broad adoption and equitable access.

Ethical governance has been a foundational element of the project, with structures in place to prevent exploitation, ensure data privacy, and maintain transparency. The project has adhered to strong legal and ethical standards, including oversight from external ethics boards and internal reviews. These practices safeguard against unfair competition, intentional bias, or market distortion and will remain central as the system moves toward real-world use.

Together, these elements position GenoMed4All as a model for trustworthy, transparent, and equitable AI in clinical research. Its focus on thorough documentation, robust human oversight, continuous performance evaluation, and ethical integrity supports not only regulatory compliance but also long-term user trust and societal benefit.

The GenoMed4All project demonstrates a strong commitment to environmental and societal well-being, embedding ethical, ecological, and civic responsibilities into the development of its AI technologies. While not yet deployed in clinical practice, the project's foundation is built on principles that promote sustainability, social inclusion, fairness, and democratic integrity. This ethos has been supported by its technical partners and infrastructure teams, whose work ensures that innovation aligns with broader societal values.

A central aspect of the project is stakeholder engagement. GenoMed4All actively incorporates feedback from clinicians, researchers, and ethics bodies across Europe, with input guided by an Independent EAB. This engagement ensures that the AI systems are not only technically robust but also socially responsible, addressing concerns around healthcare inequality, data sovereignty, and the equitable distribution of genomic technologies. Looking forward, continued dialogue with patients, civil society organisations, and environmental experts will help anticipate and mitigate potential societal and environmental risks.

Social and psychological impacts have also been thoughtfully considered. Although the AI has not interacted directly with patients during development, the project recognises the importance of preserving human agency and relationships in healthcare. The system is designed to support, not replace, clinical decision-making, maintaining the trust and communication essential to patient care. Future deployment will involve ongoing evaluations of user experience and mental well-being to ensure that the technology enhances rather than disrupts these dynamics.

Sustainability is another core pillar. GenoMed4All's FL model minimises energy use and supports local data stewardship, contributing to more environmentally sustainable healthcare practices. The project aligns itself with global goals such as the Sustainable Development Goals by promoting scalable, equitable, and efficient healthcare solutions. As high-performance computing becomes more scrutinised for its environmental impact, GenoMed4All's transparent documentation of infrastructure use positions it well for compliance with emerging regulatory standards.

The project also affirms its commitment to democratic integrity. While not directly involved in political systems, its work in healthcare demands transparency, fairness, and accountability. By preventing disinformation, ensuring oversight, and communicating responsibly, GenoMed4All fosters trust in both its technology and the institutions it supports. The project's governance reflects pluralistic values, reinforcing democratic participation and institutional resilience.

Finally, GenoMed4All is oriented toward future generations. Its ethical design, commitment to health equity, and scalable infrastructure contribute to a sustainable healthcare ecosystem. With adaptability, continuous evaluation, and environmental awareness embedded in its framework, the project aims to avoid long-term risks such as technological lock-in or erosion of clinical judgment. This foresight ensures that GenoMed4All's innovations benefit not only current populations but also those to come, supporting enduring human and societal progress.

The GenoMed4All project exemplifies a rigorous and forward-looking approach to accountability and risk management in the development of high-risk AI systems, particularly within healthcare. From early design through to anticipated deployment, the project integrates robust technical and procedural safeguards that ensure continuous risk identification, evaluation, and mitigation. This approach is deeply grounded in ethical standards, GDPR compliance, and research governance, reinforced by transparent documentation, user-focused practices, and readiness for third-party audits.

GenoMed4All's accountability framework is operationalised through structured development practices and is designed to remain adaptable to evolving regulatory landscapes, including the AI Act and requirements for Trustworthy AI. The project has proactively embedded principles of "risk-by-design," conducting early and ongoing risk assessments, particularly around clinical utility, data representativeness, and potential misuse.

As the system progresses toward market readiness, it incorporates a layered risk management strategy with real-time monitoring, iterative evaluations, and stakeholder feedback mechanisms. Residual risks are documented and assessed regarding system benefits, particularly in clinical contexts, with clear pathways for mitigating or managing these through regulatory sandboxes and pre-authorisation evaluation frameworks. Regular testing is embedded throughout development, with plans for simulation in real-world clinical settings, ensuring consistent performance, usability, and compliance.

Special emphasis is placed on the inclusion and protection of vulnerable groups. From early design through to planned deployment, GenoMed4All considers the specific risks faced by minors, the elderly, and marginalized populations. This is reflected in targeted risk assessments, testing protocols, and stakeholder engagement.

Crucially, GenoMed4All aligns its risk management strategy with broader legal and ethical requirements, including GDPR, the MDR, and the emerging European AI Act. By embedding its processes within this regulatory ecosystem, the project supports long-term sustainability, accountability, and trust, positioning itself at the forefront of responsible AI innovation in healthcare.

The GenoMed4All project demonstrates a strong commitment to transparency, accountability, and regulatory preparedness through its comprehensive approach to technical documentation, record keeping, and conformity assessment. Although the system is not yet deployed clinically, its development has been guided by anticipated requirements under the EU AI Act and related frameworks, establishing a robust foundation for future certification and lawful deployment in healthcare settings.

Detailed technical documentation has been developed throughout the project, capturing the AI system's purpose, architecture, data flows, design specifications, validation methods, and risk mitigation strategies. This evolving documentation ensures the system remains traceable, auditable, and compliant with sector-specific regulations, including the Medical Device Regulation. It also supports real-world readiness by accounting for interoperability and clarifying system boundaries.

The project has also prioritized automated record-keeping. Audit trails are designed to log key system events, including usage times, data references, inputs, outputs, and human interventions. These logs are vital for post-market monitoring, risk tracking, and incident resolution, and will be maintained in accordance with data protection regulations, ensuring security and transparency.

In preparation for formal regulatory evaluation, GenoMed4All has embedded processes consistent with the conformity assessment requirements of the AI Act. Internal controls, thorough documentation, and risk management practices have been aligned to support either internal reviews or third-party assessments, depending on the final classification and scope of the AI system. Furthermore, the project is structured to accommodate reassessment in response to substantial modifications, technical advancements, or regulatory updates.

GenoMed4All's integration of these processes not only supports future clinical deployment but also exemplifies a responsible and forward-thinking model for AI development in healthcare. By aligning with emerging legal standards and embedding compliance mechanisms throughout its lifecycle, the project ensures its outputs are technically sound, ethically grounded, and prepared for safe, lawful, and trustworthy real-world use.

### 6. Conclusions

This deliverable presents a comprehensive evaluation of AI development and validation within the GenoMed4All project, focusing on the integration, analysis, and interpretation of data related to rare haematological diseases across multiple WPs. It marks a significant advancement toward establishing a conformity assessment framework aligned with the European Commission's Ethics Guidelines for Trustworthy AI, the AI Act and the MDR.

The project has prioritised the development of lawful, ethical, and technically robust AI systems by employing tools such as the ALTAI and a GDPR-compliant data governance model. These efforts have helped shape a forward-looking compliance strategy, anticipating the roles of forthcoming regulatory bodies and the rollout of European standards expected in 2025. As regulatory expectations evolve, GenoMed4All has adapted accordingly, refining its recommendations to match the AI Act's requirements as they are currently understood.

This Deliverable acknowledges the challenges posed by rapid technological advancement and the dynamic nature of regulatory frameworks. GenoMed4All has responded proactively, offering valuable insights for future healthcare AI projects by validating core principles such as transparency, accountability, fairness, and data governance. These validations contribute both to meeting existing regulations and to shaping best practices for AI in healthcare.

While substantial progress has been made, challenges remain, particularly in bias mitigation and the integration of AI technologies within existing clinical systems. Continued oversight, validation, and refinement of models will be essential to maintain both trust and effectiveness. The use of Regulatory Sandboxes will play a key role in supporting real-world testing and iterative development as part of future compliance strategies.

This deliverable not only serves as a strategic guide for the continued development and potential market authorisation of GenoMed4All's AI tools but also provides a foundation for future research and innovation in clinical applications. It reinforces the need for sustained regulatory assurance, even when AI systems are used to support research and discovery. The outcomes of this work will also inform the responsible deployment of similar AI-driven interventions across clinical domains, particularly in rare disease management and predictive modelling.

Ultimately, this deliverable reflects GenoMed4All's commitment to advancing trustworthy, reliable, and transparent AI in healthcare. It establishes a clear pathway for compliance while contributing to a broader vision of improved outcomes for patients, caregivers, and clinicians across Europe's healthcare innovation ecosystem.

### Supplementary Information 14| Deliverable 3.1: Roadmap for Linkage of Relevant Data Repositories

#### 1. Executive Summary

The GenoMed4All project introduces a groundbreaking approach to linking multimodal health data from repositories across the EU through a privacy-preserving FL framework. This initiative confronts complex challenges spanning legal, ethical, academic, technical, and data governance domains. Successfully addressing these issues could establish the first large-scale, GDPR-compliant federated AI model training effort with tangible benefits for clinical care and patients across Europe. Given the low prevalence of patients with Rare Diseases at individual institutions, the project aligns with the EU's strategy of fostering international collaboration through ERN.

This deliverable outlines the available data repositories within and beyond the GenoMed4All consortium and provides a detailed roadmap for their integration. It highlights the methodologies used, including expert-led literature reviews, targeted use case meetings, and surveys conducted within the Eurobloodnet ERN, all aimed at assessing data availability and sharing potential under ethical and regulatory constraints. A recurring obstacle has been the specialized nature of existing datasets, which often lack the comprehensive elements needed for AI research in the defined use cases.

To support the development of AI models, the project integrates heterogeneous data from clinical studies, hospital records, public repositories, patient registries, and EU health platforms. Integration strategies vary, including roles for consortium members as principal investigators and the incorporation of platforms like RD-Connect GPAP as FL nodes. Initial AI development requires centralizing a sufficient dataset subset, demanding strict oversight of data sharing practices, ethics approvals, and secure technical implementation. Accordingly, all data are treated as pseudonymized personal data, ensuring full GDPR compliance.

This document also defines the technical specifications for the centralized storage platform, covering cybersecurity, access control, governance, and auditability. Drawing from experience in linking and managing data tailored to Hematological Disease research, the deliverable offers a transferable framework for extending these methods to other disease areas and data types. Collectively, the outlined strategies and insights form a robust foundation for enabling secure, large-scale data pooling for healthcare AI across the EU.

#### 2. Introduction

GenoMed4All introduces a groundbreaking approach to linking distributed, multimodal healthcare data across Europe through a privacy-preserving framework. By the project's conclusion, this FL platform will enable healthcare providers to train AI models on an unprecedented scale without compromising data ownership or patient privacy. Developing such a complex infrastructure demands a comprehensive strategy that balances technical, legal, and ethical requirements. This report outlines the methodology, outcomes, and key challenges encountered while mapping the European healthcare data landscape and establishing initial requirements for integrating these diverse sources.

The project centers on secondary-use data derived from structured sources such as EHRs, patient registries, and clinical trials, as these are most suitable for AI modeling. Accessing this data remains a major hurdle due to both regulatory constraints and the lack of standardized, interoperable systems. To address this, the consortium conducted a detailed survey among members and collaborators to identify viable data sources.

These include registries such as ENROL and RaDEEP, biobanks holding genomic materials, EHRs from clinical institutions, ongoing natural history studies, public repositories like RD-Connect GPAP, and clinical studies covering both interventional and observational methodologies. Additionally, real-world data generated in routine clinical practice plays a central role in GenoMed4All, supporting the generation of robust evidence complementary to traditional clinical trials. The data types used in the project are equally varied, encompassing demographic and clinical data, laboratory test results and complex laboratory assays. A fundamental component of GenoMed4All is -omics data, including genomics, transcriptomics, proteomics, metabolomics, and radiomics.

These high-dimensional datasets pose substantial storage and processing challenges, reinforcing the need for specialized infrastructure and methodologies.

Overall, this work lays the foundation for the secure, GDPR-compliant integration of heterogeneous health data across Europe, enabling advanced AI-driven clinical research while safeguarding patient privacy and data sovereignty.

**3. Data Repositories for Rare Hematological Diseases**

*Data Repositories within the Consortium*

The GenoMed4All project is fundamentally driven by the integration of high-quality, trusted datasets, with a focus on collaborative data pooling across the EU. This section outlines both the internal data assets of the consortium and the ongoing strategy to map broader European data repositories. Initial efforts have centered on surveying existing data within consortium members, using an iterative and collaborative approach between clinical and AI experts to define disease-specific research objectives and align on data requirements. For each disease area, dedicated datasets are being curated and developed, tailored to specific clinical questions.

In the SCD domain, two datasets have been made accessible consortium-wide: one focused on brain MRI radiomics data to predict future Silent Cerebral Infarcts, and another aggregating demographic, genomic, laboratory, and radiological data from across multiple institutions. Data integration is complicated by heterogeneity in infrastructure and data formats, but efforts are underway to standardize and incorporate these into the RaDeep registry for AI-driven analysis.

For MDS, a large-scale dataset is under development comprising demographic, clinical, and multi-omics data from over 6,000 patients. This includes targeted NGS, RNA sequencing, whole-exome/genome sequencing, and data on inherited predispositions. A preliminary dataset of 2,043 patients is already in use for AI training, supporting the development of predictive models for disease evolution and outcomes.

The MM use case features a comprehensive multi-modal dataset combining clinical, laboratory, genomic, and radiological data from over 1,000 patients across Bologna and Athens. Genomic data includes copy number alterations and mutational profiling, while imaging data from FDG-PET/CT and MRI support radiogenomic analyses. Clinical datasets provide detailed treatment and outcome information, enabling the development of predictive models for residual disease and patient prognosis.

Across all disease areas, the GenoMed4All project is building the infrastructure and analytical frameworks necessary to enable robust, AI-driven insights into disease mechanisms and patient outcomes through harmonized and enriched data sources.

*EU Data Platforms, Registries, and Public Repositories*

The GenoMed4All platform is designed to interconnect not only individual datasets but also large-scale data aggregators across Europe, including rare disease registries and public repositories that meet the project's requirements. Key among these is ENROL, the European Rare Blood Disorders Platform developed within ERN-EuroBloodNet. ENROL serves as an umbrella registry for both new and existing databases, offering structured demographic, clinical, and laboratory data on rare hematological diseases. Of particular relevance to GenoMed4All is the RADeep platform within ENROL, which focuses on rare anemia disorders. Both ENROL and RADeep are in advanced development and are expected to expand significantly in patient coverage.

In addition, the European Genome-phenome Archive (EGA) plays a central role as a secure, long-term repository for consented genomic and phenotypic data, currently housing over 2 million data files from global biomedical research initiatives. The CoMMpass study also serves as a critical data source, providing comprehensive clinical and genomic information on more than 750 patients with MM, tracked from diagnosis through disease progression.

Further enriching the data landscape is the RD-Connect Genome-Phenome Analysis Platform (GPAP), part of the European Joint Programme on Rare Diseases and Solve-RD. RD-Connect GPAP integrates genomic and phenotypic data from approximately 18,000 participants and supports family-based analyses and cohort studies through a user-friendly interface with access to a range of analytical tools and external genomics resources.

Together, these platforms significantly enhance GenoMed4All's capacity to leverage diverse, high-quality data for advanced research into rare hematological disorders, enabling broader discovery and development of AI-driven insights.

*Discovering Data Repositories outside the Consortium*

As part of task T3.1, a key objective was to identify publicly accessible datasets that could complement the internal datasets already available to the GenoMed4All consortium. These additional datasets needed to align with the clinical aims of each disease use case and include demographic, clinical, laboratory, and genomic data suitable for AI-based analyses. To achieve this, a coordinated literature review and database search were conducted across all use cases, alongside a targeted survey distributed to EuroBloodNet members to assess institutional readiness and willingness to contribute data.

The literature search, focused exclusively on PubMed and limited to English-language full-text articles from the past decade, yielded limited usable datasets. For SCD, although numerous references to relevant genetic markers and inflammation-related outcomes were identified, no complete, non-aggregated public datasets matching the project's needs were found. Access to potentially useful data from portals such as dbGAP and EGA requires permission, which may be pursued in later stages. However, this process clarified the specific genetic markers of interest and identified research that may support SNP selection for future studies.

In the case of MDS, a handful of relevant datasets were identified across platforms like cBioPortal, Zenodo, dbGAP, and EGA. These include clinical, cytogenetic, and genomic data, though most were centered on acute myeloid leukemia, with limited coverage of MDS-specific parameters. For MM, a comprehensive literature and data review was conducted, yielding several relevant publications and datasets. These span state-of-the-art clinical, imaging, and genomic research, though data access remains fragmented.

To complement these findings, a data availability survey was launched in April 2021 to evaluate the capacity of member institutions to share relevant data. While 34 responses were collected, no single center could provide a complete dataset that fully matched the project's SCD clinical aims, largely due to the novelty of technologies like Oxygenscan. Only two centers could provide data on GWAS and CRP, indicating some potential for focused genetic-inflammation studies. MRI-related data availability was more encouraging, with several centers offering radiological reports and anonymized image data.

In conclusion, although no comprehensive, fully aligned public datasets were found for immediate use, this phase has established a foundation for targeted data collection, clarified clinical data requirements, and highlighted promising collaborations within the network to fill current data gaps progressively.

*Summary of Readily Available Datasets for AI Model Development*

By consolidating internal consortium data, findings from the literature review, and responses from the EuroBloodNet survey, a set of datasets has been identified as readily available for the initial development and piloting of AI models within the GenoMed4All project. These datasets, either already accessible or soon to be shared, will serve as foundational resources for the creation of federated AI methodologies across the three targeted disease areas. Each dataset aligns with specific clinical objectives, such as patient stratification, survival prediction, risk profiling, and radiomics-based outcome prediction. Their timely availability marks a critical first step toward implementing GenoMed4All's broader vision, enabling scalable, AI-driven insights and setting the precedent for future integration of additional data sources, modalities, and clinical applications.

### 4. Data Linkage Roadmap

A robust ML strategy requires a thorough understanding of the underlying data. Accordingly, the GenoMed4All approach is anchored in its three defined disease use cases while also establishing a set of overarching requirements to support both the lawful integration of current consortium data repositories and the future inclusion of new ones. Building on a detailed assessment of the existing data landscape, this roadmap outlines a framework for linking and federating rare disease data sources. It encompasses the integration of data ( centralized, hybrid, or fully federated) tailored to the project's clinical use cases. It emphasizes comprehensive data curation, standardization, and harmonization to align with FAIR principles, ensuring metadata availability, interoperable vocabularies, and appropriate licensing for accessibility and reuse. The roadmap also ensures full compliance with GDPR and relevant national laws, addressing data security, governance, and legal accountability. Finally, it advocates for linking internal datasets with external EU platforms and public repositories to enrich the data ecosystem and unlock the full potential of federated AI development.

*Data Integration*

The GenoMed4All platform is designed to accommodate diverse capacities and infrastructures across clinical data providers, ensuring flexibility in how data is contributed. At its core, the platform supports a fully FL model, where dedicated software is installed locally at healthcare providers to handle data extraction, preprocessing, and communication with the FL server. Responsibility for deploying this software may lie either with the provider or another consortium partner, based on preference and capability.

Recognizing that not all providers may have the technical infrastructure to support on-site FL, alternative models are also supported. One such option involves leveraging trusted public repositories, such as the RD-Connect platform, which is already equipped to host rare disease data and function as a FL node. A third option allows for data to be shared with a centralized repository through a secure interface, with the responsibility for ethical approval and pseudonymization resting with the data provider. To maintain AI model stability while ensuring data remains current, GenoMed4All will permit data updates at fixed intervals, with immediate correction allowed for errors. Batch processing techniques will be used to handle new data efficiently.

Regardless of the chosen integration pathway, a critical requirement emerged: all data must be temporarily centralized before AI model development begins. This ensures that researchers have the necessary access to explore and manipulate the data at high frequency during the development phase. Accordingly, the centralized repository will initially function as a secure exchange platform and later as a surrogate federated node.

Detailed case studies illustrate the practical application of these integration strategies. In simpler scenarios, such as the MDS use case, one partner, like Humanitas, will act as the data integrator. In more complex cases like MM, where no central aggregator exists, each provider is responsible for the local integration of clinical, genomic, and radiomic data. The SCD use case follows a hybrid model, where the RaDeep registry oversees primary integration. Still, data may also flow directly from providers or be supplemented by biobank samples and natural history studies. These examples underscore the flexible yet rigorous framework of GenoMed4All in enabling collaborative, secure, and legally compliant AI-driven research across a federated data ecosystem.

*Data Curation, Standardization and Harmonization*

Data quality and harmonization are critical foundations for establishing a distributed data repository and enabling FL within the GenoMed4All platform. Harmonizing clinical data at the source ensures interoperability among providers, while FL requires additional transformation layers to adapt the data to AI model specifications. Achieving this level of harmonization demands significant preparatory work, particularly in aligning equipment and procedures, as illustrated by the Oxygenscan method used in the SCD dataset. Without early standardization, later data aggregation becomes unfeasible.

The harmonization process encompasses both semantic and syntactic dimensions. Semantic standardization focuses on aligning the meaning of clinical terms and parameters, directly impacting data quality. Syntactic standardization addresses the structure and format of the data, enabling technical interoperability and improving machine readability. GenoMed4All clinical teams have invested in aligning semantic definitions across their datasets, while a dedicated effort is underway to define a Common Data Model (CDM) and standard formats, which will be detailed in upcoming deliverables.

Data integration into the platform involves multiple transformation steps. Data from healthcare providers will first be mapped to the CDM, tailored to the original data format, and then further processed to meet the input requirements of specific AI models. In federated scenarios, these steps occur locally on the edge node; in centralized integration, they are managed by the GenoMed4All platform itself. Additional transformations are needed when transferring data, such as encryption using formats like Crypt4GH.

To support research while preserving data privacy, the platform will implement robust metadata models compliant with FAIR principles, allowing users to explore the characteristics of available datasets without exposing individual-level data. Only aggregate information will be available through data exploration tools, with advanced functionalities such as cohort selection reserved for future development.

In parallel, a focused standardization initiative for genomic data is being carried out, ensuring compatibility across sources and facilitating integration with phenotypic and metadata. This involves the use of established interoperability standards such as FHIR, Phenopackets, and Beacon v2, alongside rigorous pseudonymization protocols, to ensure both privacy and usability of sensitive genomic information for AI-driven analysis.

*Data Privacy and Security*

Following extensive deliberations at the EU level, the GDPR entered into force on May 24, 2016, and became fully applicable as of May 25, 2018. This regulation superseded all previous national data protection laws implemented under Directive 95/46/EC, establishing a harmonized framework for the protection of personal data and the free movement of such data across EU Member States. The GDPR seeks to safeguard individuals' rights by ensuring robust personal data protection while simultaneously enabling data mobility to support the Digital Single Market and the broader EU internal market.

In the context of the GenoMed4All project, GDPR compliance is central to the handling of sensitive clinical and genomic data. The technical approach focuses on ensuring security and privacy through appropriate measures, primarily pseudonymization, to protect individual identities without compromising the utility of data for AI development. Full anonymization, while ideal, is impractical for rare disease datasets due to their small size and high granularity, which pose significant re-identification risks. Therefore, all data within the centralized repository and those used for FL will be pseudonymized. Clinical centers will be responsible for the pseudonymization process, guided by consortium standards and potentially using a shared tool to ensure consistency across disease domains and institutions.

From a technical perspective, GenoMed4All's centralized storage infrastructure must adhere to GDPR-compliant data protection standards. It will be hosted within the EU, configured as a secure sFTP server with encrypted storage, two-factor authentication, and strict access control. Direct file content access will be prohibited, and all interactions will be logged and audited to support traceability and accountability. Upload, update, and clearly defined access rules will govern deletion permissions, while mechanisms will be implemented to distinguish between temporary (soft) and permanent (hard) deletions. Data governance in the platform also includes role-based access controls, ensuring that only authorized users can interact with datasets according to their designated roles. AI developers will require case-by-case authorization for data access, while administrators will act in a purely supportive capacity, managing platform functionality without engaging directly with data content unless explicitly authorized.

Under the GDPR framework, GenoMed4All partners will assume the role of Joint Controllers for the centralized repository's processing operations. Data providers remain the Controllers for their data until it is transferred to the platform. Organizations delivering technical services without processing interest will act as Processors. External entities receiving data will either be considered Processors or Controllers, depending on their role and interest in data use. Pseudonymized data shared with Controllers will fall under Data Sharing Agreements, while fully anonymized data will be subject to Data Licensing Agreements.

Overall, GenoMed4All's approach embodies a comprehensive and rigorously governed data protection model, ensuring regulatory compliance, technical security, and ethical integrity throughout its federated and centralized data handling activities.

*Linking to EU Data Platforms, Registries, and Public Repositories*

The GenoMed4All platform is designed to facilitate the integration and linkage of external data sources such as public repositories, patient registries supported by ERNs, and other platforms, with the primary purpose of feeding data into the federated training of AI models. External data aggregators can participate either as FL edge nodes or by directly transferring their data to GenoMed4All's centralized storage, with all activities subject to GDPR compliance. While patient registries generally provide highly structured data, challenges remain in ensuring that their data parameters and granularity align with the specific requirements of ML algorithms, necessitating case-by-case evaluation.

GenoMed4All will demonstrate this integration through examples such as ENROL and RADeep within the SCD use case, with preliminary assessments indicating that many registries will need to expand the scope of collected data beyond epidemiological metrics to support meaningful AI analysis. Public datasets, typically accessed through Data Access Committees, usually do not function as federated nodes but can be incorporated via consortium members acting as Principal Investigators who secure access and either make the data available through federated nodes or upload it to the centralized repository. For instance, the CoMMpass dataset will be utilized in the MM use case through the UNIBO FL node.

A key objective of GenoMed4All is to link existing repositories, exemplified by RD-Connect GPAP, which will adopt the necessary technical and legal frameworks to function as a federated node. Principal Investigators within RD-Connect will have the option to contribute data for federated training and leverage GenoMed4All's AI models, while researchers already part of GenoMed4All may access RD-Connect GPAP upon completing the appropriate registration. This interconnected approach aims to streamline data sharing, enhance AI model development, and foster collaboration within the broader research community.

**5. Expanding the Portfolio of Datasets**

The GenoMed4All platform's FL approach offers complete scalability for incorporating additional data; however, integrating new diseases or data types demands significant collaboration between data scientists and clinical experts. Any new clinical data provider must submit a research proposal to a Data Access Committee, composed of experts across clinical, AI, and engineering domains, which reviews and approves access based on a clearly defined research question and intended use of platform resources. When integrating data into an existing ML model, providers must meet all GDPR and institutional approvals, after which protocols trigger model retraining. For new ML models, a precise research question must be defined, with co-design efforts serving as a foundation for expanding the platform to new diseases and data modalities.

Several challenges have emerged during dataset integration, notably data quality concerns stemming from variability in patient populations, treatment protocols, and the lack of a standardized vocabulary, which may bias analyses and limit predictive accuracy. Addressing these issues requires close collaboration among clinical data managers, researchers, and statisticians, a practice actively fostered within GenoMed4All through regular workshops. Feature incompleteness is another significant challenge, as many secondary datasets lack the breadth of laboratory, clinical, and genomic parameters needed for comprehensive ML analyses. This often results from datasets being designed for specific purposes, novel assessment techniques, or cost constraints, limiting their generalizability for new research questions. While aggregating parameters from multiple repositories could be a potential solution, the necessity of complete patient-level data for personalized medicine renders this approach difficult.

Finally, the success of GenoMed4All depends on timely acquisition of sufficient, high-quality data and requires stakeholders to develop a shared understanding of the distinct challenges posed by ML compared to traditional statistical methods. Notably, clinical experts' familiarity with classical inference-based approaches contrasts with the predictive focus of ML, affecting research question formulation and data extraction expectations. Efforts to bridge this gap and establish common terminology and guidelines will support the platform's ongoing expansion to new use cases and data types.

#### 6. Conclusions

This deliverable provides a comprehensive overview of the health data landscape relevant to the GenoMed4All platform, grounded in the initial disease use cases and offering a framework for extending to new datasets, diseases, and data types. It outlines the foundational requirements necessary to develop a GDPR-compliant and ethically sound platform for data pooling and FL. The healthcare sector's inherent heterogeneity in data processing is reflected in the diverse sources and types of data generated throughout patient care. A key outcome of this work is the exhaustive cataloging of available data repositories both within and beyond the consortium, detailing their sources, data types, and related research questions. Addressing the cyclical challenge of defining research questions versus assessing data availability was achieved through an iterative dialogue between clinical and AI experts, a process expected to recur with future dataset integrations.

Following this, the team gathered requirements for data processing and analysis through collaborative workshops, establishing initial logical and technical standards to ensure compliance with GDPR and ethical data management, while supporting personalized medicine research. The focus was primarily on pathways for clinical data providers, such as hospitals and research institutions, to integrate their data and the necessary transformations to prepare the data for AI analysis. Recognizing that federated AI model development requires an initial phase of centralized data handling, particular attention was given to the legal, privacy, technical, governance, audit, and access control requirements for centralized storage. These specifications will guide subsequent implementation efforts in the platform's architecture. The deliverable concludes by outlining key challenges and lessons learned to inform future data integrations. Overall, this work marks a significant advance in the GenoMed4All project and offers valuable insights applicable to the broader adoption of FL in healthcare within the EU.

### Supplementary Information 15| Deliverable 3.2: End User Requirements

#### 1. Executive Summary

GenoMed4All is an interdisciplinary research initiative leveraging AI to advance personalized medicine by addressing clinical challenges in diagnosis, risk assessment, patient stratification, and treatment within pilot areas such as SCD, MM and MDS. It operates on a large-scale, distributed, and privacy-preserving platform that integrates clinical, omics, and imaging data through FL. The project's methodology emphasizes continuous collaboration between clinicians and AI developers, enabling the translation of clinical questions into tailored AI-driven solutions based on data integration, harmonization, and exploratory analysis.
This deliverable outlines the end-user requirements developed over the first nine months through interdisciplinary meetings involving AI experts, clinicians, and patients. The process involved identifying key stakeholders, understanding clinical pathways and unmet needs, translating those needs into precise research questions, and defining data sharing, privacy, and ethical standards compliant with GDPR. The primary actors include software and AI developers responsible for implementing and refining AI algorithms and federated infrastructure; clinical researchers and data scientists managing data curation and providing domain expertise; clinicians who will use the platform for decision support and potentially contribute data; and patients, who are the ultimate beneficiaries and whose perspectives inform ethical and legal considerations.

The requirements address both federated AI development, emphasizing data standardization, coding conventions, robust IT support at each data source, and data transformation pipelines, and clinical needs, including the definition of relevant clinical questions, necessary data elements, harmonized vocabularies, and protocols for data security and ethical use. Challenges encountered during requirements gathering included managing heterogeneity across disease pilots, bridging technical and disciplinary gaps, and fostering clear communication between clinical and AI teams. To address these, the consortium established knowledge-sharing activities and ongoing engagement with patient organizations. Ultimately, this work establishes a comprehensive framework guiding the design and implementation of GenoMed4All technologies, ensuring alignment with clinical priorities and regulatory standards while facilitating the development of effective, personalized AI-based healthcare solutions.

**2. Introduction and Research Design**

GenoMed4All adopts an interdisciplinary approach that harnesses AI models and algorithms to advance personalized medicine by addressing clinical challenges in diagnosis, risk assessment, patient stratification, and treatment. This is achieved through a large-scale, distributed repository of integrated clinical, omics, and imaging data managed in a federated, privacy-preserving manner. The project's research methodology is grounded in continuous collaboration between clinicians and AI developers, where clinicians pose experimental questions and guide the development, validation, and refinement of AI tools by providing curated data, clinical insights, and feedback on predictive accuracy. This iterative process ensures that model outputs are translated into meaningful clinical indicators usable by healthcare providers.

The initial research design and requirements gathering phase, spanning approximately one year and culminating in D3.3, involved several key activities: identifying principal stakeholders, understanding clinical care pathways and patient journeys, translating unmet clinical needs into well-defined research questions, exploring AI and FL solutions, and defining data sharing, GDPR compliance, privacy, and ethical requirements. This document summarizes the methodology and principal findings of this process, outlining the main actors and users of the GenoMed4All platform, describing user journeys, and detailing the approach taken to capture stakeholder requirements. It also elaborates on how specific disease use cases were incorporated into the general requirements framework and highlights patient engagement activities, underscoring the project's commitment to ongoing collaboration with patient associations and future participatory initiatives.

*Main Actors*

The GenoMed4All project identifies four primary user groups essential to its success. Software and AI developers are responsible for implementing and validating the FL infrastructure, which connects diverse European data registries across the clinical pilot areas, enabling distributed AI computation while ensuring data integration, harmonization, and interoperability. They also oversee the development, training, testing, and iterative refinement of AI algorithms in collaboration with clinical researchers and data scientists. Clinical researchers and data scientists serve as both primary users and contributors, curating and annotating data, posing clinical questions, and providing ongoing feedback throughout the AI development cycle to advance research and clinical care. Clinical care providers are the end users of the platform, utilizing it for AI-supported clinical decision-making in diagnosis, risk assessment, patient stratification, and treatment planning. Their input during development is crucial for optimizing platform usability and efficiency, and they will help establish guidelines for integrating AI into clinical practice. Finally, patients stand as the principal beneficiaries, with their engagement ensuring that the platform's implementation addresses ethical, legal, and practical concerns related to big data usage, AI reliability, and patient care outcomes.

*Use Case Scenarios*

The GenoMed4All project has outlined several use cases to illustrate how end users will interact with the final platform. These use cases, developed by domain experts within the Consortium, draw on previously defined actors, data integration options, and anticipated analysis needs. The first use case involves a clinical research and care institution specializing in one of the project's disease areas seeking to join the platform. Such a center contributes clinical and research data and ensures compliance with FL specifications by providing the necessary computational resources. Upon subscription to relevant AI models, the center's data is integrated according to established standards, triggering federated re-training of these models. This process incorporates the new data into model development while maintaining quality and data integrity, with the center's contribution duly recognized. The second use case focuses on AI developers who identify datasets of interest and decide to develop new AI models. They begin by accessing synthetic data for preliminary training and subsequently launch FL training across clinical sites hosting real patient data. Throughout this cycle, developers monitor model performance and validate against potential data shifts. The third use case highlights clinical care providers benefiting from the platform through access to regularly updated AI models that support clinical decision-making. These providers contribute local data to federated training and receive updated global models, enabling them to query clinical decision support systems for personalized medicine insights. Data required for these predictions is sourced directly from hospital information systems and processed locally to deliver actionable results. This structured approach ensures secure, collaborative development and application of AI tools across the GenoMed4All network.

*Research Design Methodology*

At the onset of the GenoMed4All project, it became clear that regular, focused discussions were essential to address clinical needs and potential AI strategies. Organizing these meetings proved challenging due to the significant heterogeneity among disease pilots, varying levels of AI familiarity among clinical stakeholders, the need to maintain engagement despite technical complexity, and the interdisciplinary nature requiring effective communication among healthcare professionals, AI experts, ethicists, and data protection specialists. To manage these challenges, the project divided WP3 into sub-teams aligned with each disease pilot, holding regular, focused meetings that included AI experts to foster collaboration. A central coordinator ensured consistency across groups by moderating discussions and facilitating communication. A major challenge identified was the development of a shared vocabulary and mutual understanding between clinical and AI experts, as differences in perspectives around data-driven approaches, inference, medical data standards, modeling techniques, data availability, and workflows often led to misunderstandings. To bridge these gaps, knowledge-sharing initiatives were implemented, including workshops on machine learning fundamentals tailored to healthcare, scientific communications via a project blog, and clear explanations of AI workflows. Throughout the meetings, considerable effort was devoted to collaboratively eliciting requirements, particularly regarding data gathering and protection. The lawful and ethical sharing of data was highlighted as a complex process involving research ethics approvals, data protection oversight, detailed data protocols, and precisely defined research questions, all of which demand close interdisciplinary collaboration. Given the complexity of data availability and regulatory constraints, an iterative, collaborative approach including legal, ethical, clinical, AI, and technical stakeholders was emphasized. Beyond routine meetings, broader workshops were held to refine user journeys and consolidate stakeholder needs for both the centralized and federated phases of the platform development, employing moderated breakout sessions to ensure active participation across disciplines.

### 3. Requirements

The development of federated AI algorithms requires a careful approach to collaborative machine learning that respects privacy constraints, particularly in sensitive fields such as healthcare. FL enables multiple organizations to train models without sharing raw data, relying instead on local data processing pipelines standardized through unified coding practices, programming languages, and software frameworks. The selection of these technical tools must balance the needs of AI researchers for rapid prototyping with the platform engineers' demands for stability, scalability, and security. Open-source federated frameworks like PyTorch, TensorFlow, FATE, and FLOWER are considered viable options, alongside the development of federated versions of key algorithms. Standardization is critical for successful implementation, necessitating consistent data definitions, transformation pipelines, and designated IT support at each participating center to ensure high-quality and homogeneous inputs. While distributed hyperparameter tuning is excluded from the platform's scope, optimal parameters will be pre-determined by the consortium.

Clinically, the project has defined precise requirements, including detailed clinical questions, essential variables, and harmonized vocabularies to facilitate clear communication between clinicians and AI developers. Case

Report Forms (CRFs) tailored to each disease use case ensure consistent data collection and coding, supporting multi-center harmonization and regulatory compliance. A common protocol addressing objectives, methodology, data protection, and ethics has been created to streamline IRB submissions and align data sharing practices across countries.

Data extraction remains resource-intensive, often involving manual efforts by clinical data managers and requiring adaptable software solutions to accommodate heterogeneous clinical data sources, such as EHRs and biobanks. Automated extraction tools will be supported wherever feasible, always complying with GDPR standards.

Regarding software regulation, although obtaining CE marking for medical device software is desirable, the consortium recognizes that the project's research focus excludes clinical decision support as a formal requirement. Cybersecurity is a paramount concern, with software adhering to institutional security policies and high standards to protect sensitive medical data.

Finally, the AI models must meet stringent clinical requirements, including personalization at the patient level, robustness across independent datasets, rigorous validation procedures, fairness to avoid bias against protected groups, interpretability to foster clinical trust, and overall fitness for clinical use. The platform should also facilitate patient follow-up by enabling secure patient re-identification and incorporation of longitudinal data, enhancing the models' accuracy and relevance over time.

**4. Clinical aims: A Case Study of the Disease Use Cases within GenoMed4All**

*Sickle Cell Disease*

SCD is a prevalent and severe hematological disorder caused by a mutation in the beta-globin gene, resulting in abnormal sickle-shaped red blood cells that impair microvascular blood flow. This leads to acute painful crises, chronic hemolytic anemia, tissue ischemia, inflammation, and progressive organ damage, contributing to significant morbidity and early mortality. The most common form, homozygous HbSS, produces pathological hemoglobin S, while other compound heterozygous forms also cause disease. Diagnosis can occur prenatally, at birth through screening, or later based on clinical suspicion. Management requires a multidisciplinary team for lifelong monitoring of both acute and chronic complications, with treatments primarily including blood transfusions and hydroxycarbamide, while stem cell transplantation offers a potential cure but is limited by donor availability. Emerging therapies such as gene therapy and novel pharmacological agents targeting various pathophysiological pathways are under development.

Despite advances, clinical challenges remain due to the disease's variable phenotypic expression, complicating risk prediction and management. Within the GenoMed4All project, AI is being leveraged to stratify patients based on inflammatory and hemolytic risk profiles, predict complications like microalbuminuria, and integrate multi-omics and clinical data to personalize care. AI-driven efforts include identifying genetic variants linked to inflammation and blood rheology, developing predictive models based on the point of sickling, and analyzing radiomic features from brain MRI to detect silent cerebral infarctions. These initiatives aim to address unmet clinical needs by improving early detection and prevention of complications through tailored interventions.

*Multiple Myeloma*

MM is a complex, evolving hematological disease influenced by numerous factors, notably therapy and genomic heterogeneity, which together shape the patient's clinical trajectory. Understanding the interplay between these factors is crucial for effective disease monitoring, preventing relapse, and optimizing treatment regimens to enhance therapeutic outcomes. Recent advances in MM treatment, including monoclonal antibodies and CAR-T therapies, have significantly improved response rates and minimal residual disease negativity. Despite these advances, the disease course remains unpredictable, characterized by alternating periods of remission and relapse, with responses typically shortening and relapses becoming more aggressive over time, ultimately leading to therapy resistance. Current treatment approaches largely follow a uniform strategy, failing to account for patient heterogeneity, despite extensive genomic insights gained in recent years. Presently, age is the primary factor used for patient stratification, supplemented recently by frailty assessments, but these remain insufficient for precise risk prediction. Numerous genomic-based risk scores have been proposed, yet their accuracy varies and consensus on their reliability is lacking. This underscores the need for more robust integrative methods capable of capturing the complex interactions influencing prognosis. The GenoMed4All initiative aims to address this by applying AI to large retrospective datasets encompassing biological, genomic, clinical, and imaging data. The goal is to develop a prognostic tool that accurately identifies high-risk patients at diagnosis, enabling more personalized treatment strategies and improved management of therapeutic resources.

*Myelodysplastic Syndromes*

MDS are a rare and heterogeneous group of myeloid blood cancers primarily affecting the elderly, characterized by dysfunctional hematopoietic stem cells in the bone marrow. The clinical presentation varies widely, ranging from indolent cases with near-normal life expectancy to aggressive forms resembling AML. Early symptoms are often subtle, leading to delayed diagnosis, which is typically prompted by signs such as fatigue, infections, or bleeding. Diagnosis involves a combination of blood tests revealing cytopenias, bone marrow biopsy to assess malignant blasts, cytogenetic analysis to identify chromosomal abnormalities, peripheral blood smears, and gene mutational analysis using NGS. Treatment options remain limited, with allogeneic HSCT being the only curative approach for eligible patients under 65-70 years, although its success is hampered by significant morbidity and mortality. Other therapies, such as lenalidomide and hypomethylating agents, offer symptomatic relief or survival benefits in select cases. The World Health Organization classifies MDS into several subtypes based on clinical and morphological criteria, with cytogenetics playing a crucial role in prognosis and treatment decisions. However, existing prognostic tools like the Revised International Prognostic Scoring System (IPSS-R) have limitations, particularly in capturing individual patient risk, as they rely heavily on morphological assessment and cytogenetics, which may not reflect the full spectrum of genomic abnormalities. Recent advances have identified recurrent mutations in genes involved in DNA methylation, chromatin modification, and splicing, which significantly influence disease progression, survival, and response to therapy. Notably, mutations in TP53 and others predict relapse risk post-transplant, while specific mutations such as SF3B1 correlate with certain phenotypes and treatment responses. The GENOMED4ALL project aims to address two critical needs in MDS: improving disease classification by integrating genomic data to better define genotype-phenotype correlations, and developing innovative prognostic models that incorporate clinical and molecular features to more accurately predict survival and leukemic transformation. To achieve these goals, advanced AI techniques, including statistical learning and ML methods, will be applied to large, well-characterized patient cohorts with comprehensive clinical, genomic, and outcome data. This approach seeks to enhance the accuracy, interpretability, and clinical utility of predictive models, ultimately enabling more personalized and effective management of MDS patients.

**5. Involvement of Patients**

The GenoMed4All project emphasizes a collaborative dialogue between clinical and AI domains, recognizing the essential role of patients and their organizations from the earliest stages. This engagement is critical not only for transparent communication of clinical outcomes and societal benefits but also to understand and address patients' legitimate concerns, expectations, and requirements throughout the project. To facilitate this, the Consortium has established regular interactions with patient associations at both European and national levels, sharing the project's mission, methodologies, clinical challenges, and results to gather feedback and incorporate patient perspectives into the project's development.

A key milestone was the first patient engagement e-workshop held in September 2021, which brought together clinical and AI experts from GenoMed4All with representatives from European patient groups related to MDS, SCD MM, and broader rare disease networks. The workshop was carefully organized with broad dissemination efforts and attracted around 45 participants, including speakers and patient representatives. The agenda included presentations on the project's objectives, clinical pilots, AI applications in hematological diseases, and the transformative potential of AI in advancing personalized medicine. Patient representatives shared their missions, raised important questions regarding AI data security, interoperability with other European repositories, and the risks of algorithmic errors.

The workshop succeeded in fostering mutual understanding and interest in the project's goals and the evolving role of AI in personalized healthcare. Moving forward, GenoMed4All plans to deepen patient involvement, integrating their insights into prioritizing research and patient care issues. The project remains committed to a patient-centered approach, ensuring continuous collaboration and transparency with patient associations. It will also invest in training and educational initiatives about AI's role in hematology, conducted in multiple languages and in partnership with relevant European networks, to empower patients and stakeholders alike.

### Supplementary Information 16| Deliverable 3.3: Data model

#### 1. Executive Summary

This deliverable outlines the progress made in designing the data model that underpins data representation and processing within the GenoMed4All FL platform. It translates identified requirements into detailed specifications that guide the implementation of data flows across multiple WPs. Given limited data availability, the design process relied heavily on available metadata and clinical expert input, resulting in a set of well-founded recommendations and guidelines for future development. The platform will adopt a select set of terminologies and standards to standardize internal data representation and provide tools for mapping raw clinical data. Development will occur in two phases: initially centralized, followed by a federated phase, both utilizing the same data model to ensure consistency and efficiency. Key stakeholders include clinical data providers, AI developers, and platform engineers, all benefiting from the model through streamlined integration, standardized data formats, and support for FL, while promoting FAIR principles, scalability, and data quality.

The design evolved iteratively through surveys, workshops, and consultations, establishing foundational principles such as patient-centricity, ease of data retrieval and transformation, connectivity, and data normalization. These informed requirements span data structure, transformations, storage, security, quality, and extensibility. A notable requirement was that source data formats remain transparent to AI model training, necessitating a preprocessing pipeline for non-numerical features. A parallel review of genomic data concluded that variant calling, rather than raw sequencing data, would define the genomic granularity initially supported.

The evaluation of candidate data models (OMOP, HL7 FHIR, and Phenopackets) led to the selection of HL7 FHIR due to its widespread adoption, interoperability, community support, and Consortium expertise. The finalized data model adopts a patient-centric structure based on FHIR resources, linking patients to various data elements such as laboratory results, genomic information, and clinical manifestations, facilitating efficient dataset assembly for AI training. Standard mappings to FHIR resources are provided for key data types, and data homogenization across providers will be implemented via ETL tools. Data storage will utilize a document-oriented NoSQL server to optimize retrieval speed. The deliverable also presents an overview of the platform's software components and data flows, detailing the transformation processes from raw clinical data to AI training inputs.

#### 2. Introduction

Most clinical and healthcare data providers in the EU store information in proprietary, often heterogeneous formats, which hinders interoperability, data sharing, and reproducible scientific collaboration. To address this challenge, standardized data models are developed to formalize the structure and relationships of data elements in a machine-readable format, encompassing both syntactic rules for data representation and semantic definitions that reduce ambiguity in interpretation. These models typically define data types, constraints, relationships, and metadata to ensure consistent meaning and use across datasets. Various healthcare-specific data models and standards exist, including i2b2, OMOP, PCORnet, FHIR, and rare disease-focused models like Phenopackets, complemented by standardized terminologies such as SNOMED-CT, LOINC, ICD, Orphanet's ORDO, and genomics ontologies like HGNC and Ensembl.

The data model for the GenoMed4All platform was developed through a comprehensive, multidisciplinary process involving six key steps: exploratory analysis, requirements elicitation, dataset and metadata collection, comparative model analysis, consensus building, and final design with necessary extensions. Due to privacy constraints, the design relied on metadata rather than actual patient data, leveraging close collaboration with clinical partners to gather structural and semantic information. This approach allowed a preliminary model design while acknowledging potential future challenges as real data became available.

This deliverable represents the conclusion of WP 3's efforts and serves as a foundation for subsequent WPs, especially WPs 4, 5, 6, and 8, which will handle the implementation of the FL architecture, interoperability platform, and genomics standardization. Contributions from multiple WPs ensured that requirements were gathered from all relevant perspectives: clinical data providers defined input data standards; AI developers specified output needs for model training; platform engineers addressed architectural and technical constraints; and genomics experts guided omics data modeling.

While based on metadata agreements viewed as preliminary contracts between data providers and implementers, the data model is presented as a set of well-motivated recommendations rather than a finalized specification. This approach provides necessary flexibility to accommodate unforeseen complexities once actual data ingestion begins, with final decisions on the data model's implementation deferred to the teams responsible for subsequent development stages.

**3. Data Model**

GenoMed4All aims to create an interoperability platform to enable federated training of AI models specifically for hematological rare diseases by connecting dispersed health data repositories across the EU. Achieving this requires a foundation of homogenized and interoperable data, guided by principles including system and data interoperability to facilitate international collaboration, the use of open-source components, adherence to international standards for data representation supported by data dictionaries and standardized ontologies, and prioritization of scalability, replicability, configurability, and modularity. In designing a scalable and interoperable data model, three levels of harmonization were identified: allowing data sources to follow arbitrary standards with thorough documentation; enforcing specific international terminologies and standards rigidly at the source; and developing a custom standard tailored for federated AI training, also enforced at the source. The first approach was deemed impractical due to scalability concerns, while the third was not feasible because clinical data providers lack the resources to reshape their data to fit a proprietary format. Consequently, GenoMed4All will adopt a limited set of selected terminologies and standards for internal data representation and supply software tools to map raw clinical data for both input and output, minimizing the burden on data providers and ensuring consistency within the platform.

*Genomed4All Context and Motivation*

The data flows within the GenoMed4All platform are structured around the roles of its stakeholders and the platform's development stages. Clinical data providers are responsible for generating, collecting, and sharing raw data, while AI experts and data scientists utilize this data to train predictive models. The platform's development is divided into two phases: an initial centralized phase and a subsequent federated phase. During the centralized phase, data is securely shared and stored centrally, allowing AI algorithm development to proceed with data represented in a standardized CDM to enhance interoperability, discoverability, and usability. Given that raw clinical data varies in structure and format, multiple rounds of data transformation and mapping are required to harmonize disparate datasets into a unified format, facilitating the training of multiple AI models across various disease use cases. In the federated phase, clinical data remains locally stored at each provider's site, with only AI models exchanged over the network to ensure privacy and security. In this context, the CDM is essential for guaranteeing that data contributions from different centers remain standardized and interoperable, necessitating local transformation into the CDM before AI model training. Scalability and semantic harmonization are critical to enabling the seamless integration of additional data sources, requiring each dataset to be accompanied by a detailed data vocabulary and adherence to established interoperability standards and healthcare ontologies. The platform's design supports multiple dimensions of scalability, including sample size, AI analysis types, disease areas, and diverse clinical infrastructures. Organizing data into datasets aligned with specific clinical objectives allows for multi-use by various AI models, promoting maximal data reuse and avoiding data silos. Furthermore, ensuring high data quality is vital for effective AI training, and automated quality assessment tools depend on data being represented consistently through the CDM. Overall, adopting a unified common data model across diseases delivers significant benefits by simplifying data mapping, fostering semantic consistency, enabling cross-border interoperability, supporting FAIR data principles, facilitating FL, enhancing scalability, lowering barriers for new developers, maximizing data reuse, and enabling automated quality controls.

*Genomed4All Data Types and Metadata*

The initial phase of designing the GenoMed4All data model focused on thoroughly understanding the types of data to be represented, balancing the need to accommodate the specifics of existing datasets to create a scalable, generalizable platform. This required extensive collaboration among clinical teams, AI experts, and engineers to anticipate future research questions and translate them into concrete data requirements. Clinical teams provided detailed information on the datasets relevant to their disease areas, specifying data types and preferred standardized terminologies or ontologies to ensure semantic consistency. Particular attention was given to genomic data, where three levels of granularity were identified: high-level mutation presence within genes, detailed variant calling information, and raw sequencing data. However, the project's initial phases focus on variant calling data, as raw sequencing data is not yet available. For each case the respective datasets were outlined, including gene mutation screening, cytogenetics, somatic mutation data, copy number alterations, SNP arrays, and metabolomics, each contributing to the AI-driven analyses. The approach ensures that the data model is both detailed enough to capture critical molecular and clinical parameters while remaining adaptable for future expansions and varied research objectives.

*Requirements and Assumptions*

Task T3.3 aimed to establish foundational assumptions and requirements for the GenoMed4All data model through collaborative efforts within the data modeling task force and broader stakeholder validation. The process began with identifying key assumptions, including acceptance that raw source data would not be altered prior to ingestion, and that data formats may vary and not always conform to interoperability standards. The model needed to accommodate heterogeneous clinical data types from multiple sources linked to individual patients, with infrequent updates reflecting longitudinal follow-up or corrections. The data model was designed to facilitate efficient extraction and transformation of relevant datasets for AI algorithm training, emphasizing a patient-centric approach, normalization to minimize data duplication, and seamless integration between data sources and AI consumption points.

The model supports both centralized storage and FL environments while maintaining strict data protection through pseudonymization and limiting data access during federated phases. Data quality and harmonization were prioritized, ensuring that the data model at least preserves all information present in source data, enriched only after clinical validation, and aligns with existing terminologies and ontologies.

Given that AI training requires numerical tabular data, a robust, transparent preprocessing pipeline was mandated to convert non-numerical data such as categorical terms, genomics, and free text, into machine-readable formats. This pipeline must be standardized, reproducible, and collaboratively defined across consortium partners.

Specific requirements for -omics data representation were developed based on detailed surveys of clinical partners regarding sequencing protocols, data types, and research goals. The model captures essential metadata such as sequencing details, pipelines used, reference genomes, and variant definitions, while accommodating varying granularity in genomic data availability, from detailed variant-level information to gene-level mutation presence. Established ontologies like HGNC, HGVS, and ISCN were selected for semantic consistency, with plans to supplement them to enhance stability and reproducibility, ensuring the data model can flexibly support diverse genomic datasets and evolving research needs.

*State of the Art*

The review highlights three major healthcare data standards relevant to the GenoMed4All project. FHIR, developed by HL7, facilitates efficient data exchange through modular resources designed for rapid transmission of discrete information. While FHIR supports semantic interoperability and streamlined data sharing, its tree-like structure, optimized for exchange rather than analysis, poses challenges for data storage and complex analytics. In particular, integrating genomics data into EHRs is a technical hurdle due to varied collection and coding methods, though extensions like SMART on FHIR Genomics enable standardized clinical genetics data representation to support personalized medicine.

The Observational Medical Outcomes Partnership Common Data Model (OMOP-CDM) is a widely adopted framework designed to unify diverse observational health data for robust analytical use, including clinical, economic, and metadata domains. While initially limited in cancer-specific detail, extensions such as the OMOP Oncology Module and integration of specialized ontologies have enhanced its capability to comprehensively represent cancer diagnoses, treatments, and episodes. Additionally, the genomic data gap was addressed by the Genome Common Data Model (G-CDM), which expands OMOP-CDM to include next-generation sequencing data and links clinical and genetic information using standardized vocabularies, thereby enabling secure multi-center genomic research through aggregate data sharing.

Finally, Phenopackets addresses the critical need for a standardized format to share phenotypic information alongside genotype data, which traditional genomic formats often omit. By fostering interoperability between clinical and research communities, Phenopackets facilitates more accurate genotype-phenotype matching, enhancing diagnosis, treatment, and research opportunities. Collectively, these standards provide complementary frameworks for integrating clinical, genomic, and phenotypic data, each with distinct strengths and limitations relevant to the project's objectives.

*Exploratory Mapping of Data Model Requirements to OMOP, FHIR and Phenopacket*

The following section details a mapping exercise conducted to evaluate the adequacy of previously discussed data models in addressing the comprehensive data requirements of GenoMed4All. Drawing from prior requirement analyses, several evaluation dimensions were established to assess each model's expressivity and capability to represent all relevant data types. The results, summarized across multiple tables, indicate that all three data models generally meet the clinical and genomics data needs of the project. However, Phenopackets show some limitations in representing less common data types such as imaging and other -omics data. Further evaluation considered factors such as ease of data interaction and the capacity to incorporate governance and auditing information for quality assurance, providing a thorough basis for selecting the most suitable data model framework.

*GenoMed4All Choice of CDM*

Based on the mapping exercise, both FHIR and OMOP-CDM adequately support the clinical, genomic, and other relevant data requirements of the project, as well as the necessary processes for querying and transforming data for AI applications. The GenoMed4All consortium has chosen HL7 FHIR as the primary data model for data selection and ingestion from hospitals, given its comprehensive adherence to project requirements and its widespread adoption in EHRs. The increasing implementation of FHIR by hospitals and EHR vendors further supports its selection, facilitating future cross-institutional collaborations and interoperability. Additionally, several partners possess established expertise with FHIR and its associated tools. Nonetheless, the team remains open to utilizing OMOP-CDM should FHIR prove insufficient for representing currently unknown data types, potentially adopting OMOP-CDM as the main model while maintaining FHIR for data exchange or integrating missing components. Consistent with the original proposal, efforts continue to implement Phenopackets within the FHIR framework to enhance genomic data interoperability and standardization, potentially through integration with Phenopackets-based platforms or by leveraging its protobuf interface for data science applications.

*GenoMed4All Data Model Specifications*

The HL7 FHIR standard defines a comprehensive set of resources to model administrative and clinical concepts, including patients, providers, organizations, devices, and a broad range of clinical data such as problems, medications, diagnostics, and care plans. Within GenoMed4All, a focused subset of these resources will be employed to model patient clinical, genomic, and other -omics data following a patient-centric approach. Each patient is represented by the Patient resource at the center, with linked resources for laboratory tests, genomics data, clinical manifestations, and more, connected through FHIR references to facilitate efficient querying via the FHIR API. Key resources include Patient for demographics, Observation for clinical and genomic data (such as hematological values or genomic findings), Condition for diseases and clinical events, Procedure for medical interventions, ServiceRequest for planned diagnostics or treatments, and DiagnosticReport for summarizing diagnostic information, particularly genomics data grouped from variant call files. These resources are stored and exchanged in JSON format consistent with the project's architectural principles.

For genomics and cytogenetics data, FHIR suggests using DiagnosticReport to encapsulate overall genomic report metadata and group related observations, which are the primary means to represent genomic information. Observations are categorized into genomic findings (descriptions of genomic characteristics like variants, genotypes, and haplotypes), genomic interpretations (laboratory conclusions such as positive or negative results), and genomic implications (assertions about the effects of genomic results on the patient). Due to limited data volume from clinical providers, only relevant information will be stored in the common data model, without covering all resource specifications. Examples illustrate how genomics data, such as genotype and variant findings, are structured within Observation resources, demonstrating FHIR's flexibility and specificity for integrating complex clinical and genomic information within GenoMed4All's data framework.

**4. Overview of Functional Architecture and Data Flows**

The functional architecture, which will be detailed further in D5.1, is based on a distributed edge-node model closely mirroring the central node in terms of components and interactions. The CDM serves as the reference framework for standardizing data from multiple providers, with data integration implemented via an ETL tool. Data is stored in a document-oriented NoSQL database, such as Elasticsearch or MongoDB, enabling rapid access. For privacy and security reasons, Genomed4All does not directly access healthcare data; instead, data is provided in standardized formats defined by each provider. The ETL process translates this input into the FHIR-based CDM, storing it within an integrated record accessible through a FHIR server and API. The architecture and data flows are illustrated through simplified and detailed diagrams, highlighting the software components deployed at FL edge nodes. Data undergoes multiple transformation stages from raw clinical input to AI model training data, with these processes being consistent across both federated and centralized training approaches. During centralized training, transformations occur within the platform, while federated training requires all transformations to happen locally at each edge node, ensuring data privacy and consistency. Detailed workflows clarify each transformation and data transfer step alongside the involved software components.

**5. Conclusions**

This deliverable outlines the methodology and key outcomes of the data modeling task T3.3, which was developed through a collaborative and iterative process starting from foundational assumptions and principles, culminating in a comprehensive set of requirements. These requirements guided the evaluation of various data models, resulting in the selection of the HL7 FHIR standard. The document also explains how the data model integrates into the overall platform architecture, emphasizing data flows and transformations from raw clinical data to inputs for AI model training. Recognizing the limited availability of diverse datasets at the time, the deliverable presents its specifications as best recommendations for future implementation, while allowing for alternative approaches if necessary. Key requirements include using a CMD for both centralized and federated phases, ensuring transparency in the data transformation pipeline for both data providers and AI developers, and focusing on variant-calling genomic data rather than raw sequence reads during initial project phases. Following extensive literature review and evaluation, the chosen FHIR-based data model adopts a patient-centric approach, linking patients to multiple resource types such as laboratory values, clinical manifestations, and genomics data. This design facilitates efficient data retrieval, simplifies data ingestion and transformation, and aligns well with the platform architecture. This work completes the project's requirements-gathering phase and, together with related deliverables, establishes the foundation for the technical implementation of data flows within the GenoMed4All platform, providing detailed guidelines based on stakeholder input from clinical, AI, and platform development perspectives. Combined with other key documents, this deliverable offers a clear understanding of the platform's capabilities and architecture, serving as the logical basis for implementation by subsequent WPs.

### Supplementary Information 17| Deliverable 4.1: Hybrid Federated Learning Models

#### 1. Executive Summary

The GenoMed4ALL platform seeks to unify clinical, genomic, and other "-omics" data under a privacy-preserving, distributed learning architecture. The sensitivity of the data involved necessitates a technical infrastructure that supports robust anonymization and local computation to comply with legal and ethical constraints. D4.1 presents a dual implementation strategy that begins with a centralized training phase to develop initial model baselines, followed by a FL phase that enables distributed training across institutional nodes without requiring raw data transfer. The deliverable includes benchmarking results, a review of existing FL frameworks, and the rationale for selecting Flower as the base platform for further development.

#### 2. Objectives

The overarching objective of this work is to define a hybrid FL infrastructure capable of integrating and analyzing health-related data distributed across multiple institutions. The approach merges federated and centralized machine learning strategies to allow participation from both large, technically equipped centers and smaller institutions with limited computational capabilities. The hybrid design ensures that institutions can contribute to AI model development without compromising patient privacy or contravening data governance regulations. The deliverable also sets out to create a set of technical and functional requirements that will guide the implementation of the GenoMed4ALL FL system, ultimately enabling the development and sharing of AI models to support clinical decision-making in rare hematological conditions.

#### 3. Methods Overview

The methods employed in this deliverable rely on the structured integration of clinical and genomic data from project partners. The selected ML models include XGBoost, known for its performance on tabular data, and multi-layer perceptrons, which were used for initial experiments in both centralized and FL contexts. The FL experiments were built using Flower, an open-source framework chosen for its flexibility, extensibility, and support for popular machine learning libraries such as TensorFlow and PyTorch. The hybrid learning strategy begins with centralized training using pseudonymized features derived from real patient data. Once the baseline model is established, the architecture transitions to an FL mode that allows edge nodes to train locally and share only model updates, thereby preserving data privacy. A range of tools was also developed to support secure communication, training orchestration, and performance monitoring across the federated network.

#### 4. Key Findings

The benchmarking experiments confirmed that the centralized models performed strongly, particularly with XGBoost, delivering high accuracy on the structured datasets available. FL experiments, though in the early stages, proved technically feasible and capable of replicating the performance trends seen in centralized setups, albeit with added communication overhead and operational complexity. The report demonstrates that Flower, while suitable for initial deployment, requires further modular development to meet the full set of requirements specified by clinical and AI stakeholders. In particular, additional modules are needed for managing node registration, secure aggregation, and task scheduling. The findings also highlight the importance of aligning data formats and procedures across institutions to support federated model convergence and generalizability.

**5. Conclusions and Future Work**

This deliverable establishes a technical and conceptual foundation for FL in the GenoMed4ALL project. It validates the hybrid approach as a viable strategy for enabling collaborative model development across geographically and institutionally diverse healthcare settings. The work confirms that FL can provide strong data privacy guarantees while still enabling the development of high-performing AI models. Nonetheless, the current implementation represents an early-stage prototype, and significant work remains. Future activities will focus on refining the FL pipeline, expanding support for more diverse and complex datasets, integrating additional privacy-preserving technologies such as differential privacy, and ensuring the robustness and scalability of the system. The eventual goal is to deploy the federated platform across multiple clinical sites, enabling real-time clinical decision support for rare hematological diseases based on shared, yet private, AI model training.

### Supplementary Information 18| Deliverable 4.2 - Federated Data Sharing Platform Architecture and Technologies

The GenoMed4All federated data sharing platform, developed under the Horizon 2020 programme (Grant Agreement No. 101017549), delivers a secure, extensible infrastructure enabling FL across distributed clinical and genomic datasets for hematological diseases. In recognition of legal and ethical constraints on health data sharing, the platform is designed to support data locality while enabling collaborative AI model training across decentralized nodes.

**1. Architecture Overview**

The GenoMed4All platform architecture follows a modular and layered design, comprising a Central Node and multiple Edge Nodes that work together to enable secure federated machine learning workflows. The Central Node functions as the orchestrator, handling user interactions, model and dataset registries, job scheduling, and security enforcement. It is developed using FastAPI, a modern Python web framework that supports asynchronous, high-performance RESTful APIs and integrates with OpenAPI for automatic documentation through Swagger and Redoc.

Metadata persistence in the Central Node is managed using PostgreSQL, interfaced via SQLAlchemy ORM to ensure reliable and scalable data handling. User authentication is implemented using Keycloak, an open-source identity and access management system that supports the OpenID Connect protocol, allowing federated identity management. For API security and role-based authorization, the platform employs Kong as a gateway, ensuring endpoint protection and policy enforcement.

MLflow is integrated into the architecture to support model tracking, versioning, and lifecycle management. It allows researchers to register models, monitor training metrics, and store artifacts in a structured and reproducible manner. FL orchestration is handled by Flower, a Python-based framework that manages decentralized training rounds, model aggregation, and coordination between the Central Node and Edge Nodes.

The user interface is implemented as a Single Page Application (SPA) using Vue.js. This allows users, clinicians and data scientists to define training jobs, select models and datasets, monitor job progress, and access training results through an intuitive web interface.

Each data provider hosts an Edge Node, which is also built using FastAPI and PostgreSQL. Edge Nodes perform local training tasks on private data and communicate securely with the Central Node. These nodes are authenticated using asymmetric cryptography and designed for lightweight deployment to accommodate diverse IT environments. The modular design ensures that both nodes maintain consistent APIs and integration pathways, facilitating seamless orchestration of federated tasks.

**2. Data Homogenization and Dataset Management**

Data harmonization is a foundational step in federated training. GenoMed4All adopts a Common Data Model (CDM) to standardize clinical and genomic datasets provided by various institutions. The data homogenization pipeline, developed in WP5, ensures that disparate data sources are transformed and loaded into a consistent structure suitable for federated processing.

Edge nodes use this ETL (Extract, Transform, Load) process to convert local data into structured datasets, typically CSV format, tagged with relevant metadata (e.g., disease, date range, data provider, version). These datasets are then registered with the central node, which maintains a searchable registry of available datasets via API endpoints.

In the future, the platform plans to support additional formats such as Parquet, and to integrate dataset quality metrics and semantic tags to enhance discoverability and usability for model training.

**3. Platform Workflow and Training Execution**

The platform workflow begins with user authentication, which is managed through Keycloak using the OpenID Connect protocol to ensure secure and federated access. Once authenticated, users can register preprocessed datasets through the FastAPI-based endpoints exposed by the platform. These datasets, previously harmonized through a standardized ETL pipeline, are stored along with associated metadata. Researchers then upload Python-based machine learning models to the MLflow registry, which serves as the central repository for versioning, tracking, and managing models. Through the platform's web-based user interface, users create training plans by selecting a model, associating it with one or more registered datasets, and defining the necessary training parameters. Once a training plan is submitted, the FL process is coordinated by the Flower framework. Flower handles communication and orchestration between the central server and the participating edge nodes, allowing training to be executed across decentralized datasets without compromising data privacy. Throughout this process, MLflow tracks key training metrics, hyperparameters, and artifacts generated during training. The results can be accessed via MLflow's integrated interface or exported for visualization in TensorBoard, enabling researchers to analyze and compare experiments effectively.

**4. Security and Development Methodology**

Security is enforced through layered mechanisms: Keycloak provides federated identity management, Kong controls API access, and nodes use asymmetric cryptography for mutual verification. TLS tunneling and audit logging are planned for future iterations.

Development follows agile methodology using GitLab for issue tracking and code versioning. Continuous Integration (CI) via Jenkins is planned, with an emphasis on test coverage for backend services. Stakeholder feedback is continuously integrated into development cycles to ensure alignment with evolving requirements.

**5. Conclusion**

The GenoMed4All platform offers a robust, scalable solution for secure federated machine learning on sensitive health data. Its modular, standards-based architecture ensures interoperability, data sovereignty, and alignment with FAIR data principles. The system plays a central role in harmonizing and integrating inputs from multiple WPs across the GenoMed4All project ecosystem.

### Supplementary Information 19| Deliverable 4.3 – Computing distribution, including HPC AI algorithms computing

#### 1. Introduction and Background

Genomed4All aims to advance personalized medicine in hematological diseases through the integration of genomics, clinical records, and other omics data using innovative AI methods. From its inception, the project foresaw large computational needs and proposed to meet these through the use of High-Performance Computing (HPC) hosted at CINECA. The intention was to centralize FL tasks, where a powerful central aggregator node would handle model updates from multiple participating hospitals or institutions.

However, as the project progressed, the need for centralized HPC resources was reassessed. FL inherently distributes computational effort across nodes and does not necessitate heavy central processing. Additionally, privacy regulations and logistical barriers made centralized data processing infeasible. As a result, the role of HPC shifted to supporting specific, high-demand use cases in an on-demand fashion, making resources like Marconi100, Galileo100, and ADA Cloud available to all consortium members.

#### 2. Computing Distribution Requirements and Project Redesign

The initial architecture proposed three scenarios for incorporating HPC. The first involved HPC acting as a centralized orchestrator for FL, where model parameters would be exchanged via secure messaging and aggregation would occur at CINECA. The second scenario envisioned HPC also supporting client nodes, particularly when institutions lacked local infrastructure. This model was technically and ethically complex due to data-sharing limitations. A hybrid solution combined both models: centralized training with shareable data followed by federated updates from non-sharing institutions.

However, practical experience during the development of tasks D4.1 and D4.2 indicated that model complexity and data volume did not justify the use of centralized HPC. Models could be trained effectively on standard hardware. The relaxed time constraints for model training further undermined the need for HPC. Legal and ethical concerns around sensitive data hosting, even with mechanisms such as JCAs and encrypted VMs, added barriers. Consequently, the strategy was redesigned: HPC would no longer be the central training node but would instead be made available to partners for specific tasks.

#### 3. Use Cases for HPC Integration

Several practical use cases emerged where HPC was beneficial despite the shift in project strategy.

Data Sharing Platform: A cloud infrastructure managed by CINECA was deployed to enable secure data sharing and storage. Technical partners created self-managed virtual machines with encrypted volumes (LUKS). Strict security protocols governed access, and partners managed platform configurations. This infrastructure underpinned the secure exchange and handling of genomics data while adhering to GDPR and ethics requirements.

MRI Scans Semantic Segmentation: For SCD, the semantic segmentation of MRI images was computationally intensive, particularly during training. Marconi100's GPU-enabled environment was used to fine-tune a U-Net model through transfer learning from a multiple sclerosis segmentation task. The ensemble of fine-tuned models demonstrated superior DICE scores and segmentation accuracy compared to baseline models. These improvements were essential for improving clinical image analysis pipelines.

Validation Protocol Implementation: A validation framework was proposed to test whether an AI model could be effectively federated without significant performance degradation. The initial design involved a sophisticated architecture hosted on HPC, simulating various FL scenarios through SLURM-managed clusters. However, the complexity of opening ports and coordinating node communication raised security concerns. Instead, the solution was reimplemented using ADA Cloud virtual machines, offering similar functionality through REST

APIs, Celery, RabbitMQ, and gRPC-based services while remaining simpler and more maintainable.

Genomic Data Pipeline Testing (Support for D5.4): Hospitals lacking sufficient local computational power could process their genomic data using HPC infrastructure, particularly Galileo100. Containerized genomic pipelines were executed remotely, ensuring reproducibility and enabling participation from all institutions regardless of local resources. This support was critical for meeting D5.4 objectives.

**4. HPC Training and Capacity Building**

Given the diversified use of HPC, comprehensive training was necessary. CINECA organized two-day sessions attended by up to 30 consortium members. The first session covered foundational HPC knowledge, including system architecture, file storage environments, logging in, and loading modules. A focus was placed on the SLURM workload manager and differences between CPU and GPU computing.

The second session, aimed at users with programming experience, showcased optimized file formats (e.g., Parquet, Feather), Python multiprocessing, GPU-accelerated libraries (e.g., CuPy, cuDF), and Snakemake for pipeline management. The sessions emphasized practical demonstrations and real-time HPC usage. Recordings and materials were distributed to all partners. Further training sessions were proposed to deepen knowledge in bioinformatics workflows and distributed computing with Dask and Spark.

**5. HPC Infrastructure Overview**

CINECA offers several HPC systems:

- Marconi100, a GPU-optimized cluster ideal for AI and ML tasks, providing 32 Pflop/s performance with 980 nodes.
- Galileo100, a data-oriented cluster with high RAM and storage, better suited for bioinformatics and genomics.
- Leonardo, one of the world’s top supercomputers, was under final setup during the project and will offer 309 Pflop/s when operational.
- ADA Cloud, a secure OpenStack-based platform for managing virtualized HPC/cloud resources, critical for sensitive data applications.

These systems collectively enable flexible access to computing capabilities across a wide range of use cases.

**6. Conclusions and Future Contributions**

The shift in HPC’s role within Genomed4All, from a centralized computing backbone to a distributed, on-demand resource, reflects both the evolving understanding of FL’s needs and the complexity of real-world constraints. Although full-scale centralized HPC use was abandoned, the infrastructure continues to support critical project components such as image analysis and genomic data processing.

Key takeaways include recognizing that FL’s distributed nature reduces the need for centralized computational power, and that GDPR and system security limitations can hinder centralized data processing. There remains a significant knowledge gap between biomedical research and HPC usage. Closing this gap will require further training, simplification of access procedures, and the development of tools tailored specifically for life sciences. By maintaining flexibility and offering specialized support, HPC remains an integral part of the project’s infrastructure and continues to enable high-impact outcomes.

### Supplementary Information 20| Deliverable 5.1 - Homogenization Requirements and Specifications

#### 1. Overview

The GENOMED4ALL project is a large-scale European initiative focused on improving diagnosis and treatment of hematological diseases through the application of artificial intelligence. A major challenge lies in the heterogeneity of biomedical data, which includes structured, semi-structured, and unstructured formats, encompassing EHRs, genomic sequences, medical imaging, laboratory tests, and patient-reported outcomes. This document details the requirements and technical specifications for data homogenisation as part of the broader effort to create a secure, scalable, and AI-ready data infrastructure. By establishing a unified approach based on HL7 FHIR standards and incorporating both centralized and federated data architectures, GENOMED4ALL seeks to enable effective, ethical, and interoperable data use across clinical and research institutions in Europe.

#### 2. Available Data Types and Sources

Data collected within GENOMED4ALL originates from a diverse range of repositories and healthcare systems, each contributing different modalities of information. Demographic data, such as age and gender, are included with appropriate anonymization to avoid reidentification, while more sensitive personal identifiers are redacted. Clinical data capture diagnostic details, therapeutic interventions, and patient trajectories through various stages of disease. Laboratory data, particularly from blood analyses, provide critical indicators of hematological status and are often structured and standardized, making them ideal for modeling. A key component is the integration of omics data, including genomics, transcriptomics, proteomics, and metabolomics. These data offer insights into the molecular underpinnings of disease and are essential for precision medicine approaches. Radiomics data, derived from imaging modalities, and complex tests like the Oxygenscan are also incorporated. The wide variability in data types, formats, and quality necessitates a comprehensive and adaptable strategy for homogenisation, one that ensures semantic alignment and analytical readiness.

#### 3. Methodological Foundations and Standards

The project adopts a semantic integration framework based on the use of established procedures and common data models. Ontologies such as SNOMED CT, MeSH, UMLS, the Disease Ontology, and the Gene Ontology provide a controlled vocabulary to unify disparate datasets. Standards like HL7 FHIR are used to define the structure of health data, enabling interoperability and traceability. FHIR's modular design facilitates granular data representation while its native API support ensures seamless integration with existing health information systems. The OMOP Common Data Model is also recognized for its ability to support observational studies and has been extended in GENOMED4ALL to accommodate oncology and genomic data. Integration architectures are categorized into centralized, federated, and hybrid models. Centralized models offer efficiency in querying and data management but raise concerns about privacy and regulatory compliance. Federated models preserve institutional autonomy and data sovereignty by keeping data locally stored, transmitting only anonymized outputs or trained models. Hybrid models combine elements of both, offering a balanced approach to performance, security, and interoperability.

#### 4. Platform Requirements and Specifications

A robust data infrastructure must satisfy a range of functional and non-functional requirements. Privacy and security are prioritized through mandatory data anonymization, pseudonymization, role-based access control, authentication and authorization protocols, and audit logging. Compliance with the GDPR is enforced across all components. The platform must support a unified common data model that captures the full scope of clinical and molecular data without loss of fidelity. Terminology services must enable mappings between native data schemas and standardized ontologies. ETL tools are needed to facilitate the transformation of raw data into the common data model, including the extraction of relevant variables, normalization of formats, and application of coding systems. The system should accommodate updates to data, particularly in the context of longitudinal studies. Data storage must be flexible enough to support both central and local repositories, depending on the deployment phase. Querying tools and APIs should allow the extraction of clean, well-curated, and numerically encoded datasets for downstream AI processing, with support for metadata and data quality assessment.

**5. End-to-End Data Homogenisation Workflow**

The data homogenisation workflow begins with data provision from healthcare institutions, which export raw data in predefined formats in compliance with ethical and legal constraints. ETL processes then transform this data into the FHIR-based common data model. Transformed data are ingested into integrated repositories using ElasticSearch or MongoDB backends and exposed via a FHIR server. Next, a data curation pipeline performs quality checks and structural validations, ensuring that the data is suitable for advanced analytics. A unified Data API facilitates the generation of AI-ready inputs by transforming the harmonized datasets into tabular formats such as CSV, suitable for machine learning. These files are then used to train and validate AI models. In the FL phase, each institution operates as an edge node, training models on local data and transmitting parameters to a central server. The server merges these inputs to create a global model, which is redistributed to all nodes, thereby enhancing generalizability while preserving patient confidentiality.

**6. Specific Considerations for Data Formats**

The homogenisation process accommodates various data types. For structured data like relational databases and CSVs, mapping tables are defined to transform source fields into corresponding FHIR resource attributes. Non-structured data, such as free-text reports or legacy formats, require closer collaboration with data providers to extract relevant features using natural language processing or manual annotation. Imaging data are handled in two categories. DICOM images are transformed into FHIR Media or ImagingStudy resources, with patient metadata extracted separately and linked accordingly. Non-DICOM images are integrated only if they can be securely accessed and semantically matched to the appropriate clinical context. For genomic data in Variant Call Format (VCF), the vcf2fhir tool is used to generate structured FHIR resources including DiagnosticReports and linked Observations. These representations are then stored in the data repository and made accessible to AI pipelines for preprocessing and analysis.

**6. Conclusion**

This deliverable defines the roadmap for building a harmonized, scalable, and privacy-compliant data infrastructure that supports artificial intelligence research within the GENOMED4ALL project. By addressing the full lifecycle of biomedical data, from acquisition and transformation to querying and federated model training, it ensures that the platform remains adaptive and interoperable. The project recognizes that the homogenisation process is iterative, requiring ongoing refinement as new datasets, standards, and analytical needs emerge. Future deliverables will continue to evolve these frameworks, with contributions from all relevant WPs to ensure coherence, efficiency, and scientific rigor.

### Supplementary Information 21| Deliverable 5.2 - Data Pre-Processing Architecture and Components

**1. Overview**

D5.2 outlines a structured, detailed account of genomic data pre-processing strategies employed across the three major hematological disease areas of the use cases. It defines harmonized pipelines, pre-processing stages, and quality control protocols required to standardize heterogeneous data into machine-readable formats suitable for artificial intelligence-driven analyses. The deliverable emphasizes reproducibility, traceability, and biological validity in converting raw sequencing output into high-confidence variant or copy number information. Each pipeline is customized to the disease-specific challenges, addressing clinical relevance and genomic complexity. The document also details the data flow from initial sample handling, through sequencing and computational analysis, to the generation of tabular datasets, which serve as the foundation for AI training within the GENOMED4ALL platform.

### 2. Myelodysplastic Syndromes Workflow

The MDS pipeline starts with the extraction of genomic DNA from either bone marrow or peripheral blood, using automated extraction systems to reduce human error and improve reproducibility. DNA quality and concentration are assessed using fluorometric tools and capillary electrophoresis systems. A custom-targeted sequencing panel tailored for MDS-related genes is used to generate libraries suitable for sequencing on Illumina platforms. After sequencing, the raw data in BCL format is converted to FASTQ files using demultiplexing tools like bcl2FASTQ. Adapter sequences are trimmed with Trimmomatic and quality is assessed using FASTQC.

Reads are aligned to the hg38 reference genome. Depending on whether Unique Molecular Identifiers (UMIs) are used, alignment and uBAM generation follow slightly different paths. With UMIs, metadata is added using the fgbio tools. GATK's best practices are employed to produce analysis-ready BAM files, involving duplicate marking, sorting, and Base Quality Score Recalibration (BQSR). Variant calling is executed in two parallel paths: GATK Mutect2 is used to detect single-nucleotide variants (SNVs) and small indels, while Pindel identifies larger indels such as FLT3-ITD or CALR deletions. Filtering mechanisms address contamination, strand bias, and sequencing artifacts.

Annotation of variants is performed using ANNOVAR, with references to population and disease-specific databases such as gnomAD, ClinVar, COSMIC, and 1000 Genomes. A consortium-defined whitelist of pathogenic variants is also applied. Semi-automated filters remove artifacts, low-confidence calls, and possible germline variants. The resulting high-confidence somatic mutations are curated manually and classified by oncogenic potential. The final output is a structured tabular file for each patient, detailing the presence of mutations in MDS-specific genes, mutation counts, and variant allele frequencies (VAFs), which can be merged with additional clinical metadata for cohort-level analysis.

### 3. Sickle Cell Disease Workflow

The SCD pipeline is based on Genome-Wide Association Studies (GWAS), aimed at identifying genetic modifiers that influence the severity and clinical presentation of the disease. While the pathogenic mutation in the HBB gene is well known, phenotypic variability is believed to be modulated by other genetic loci. Genotyping arrays are used to generate SNP data, which is then subjected to rigorous quality control to remove individuals with discordant sex, extreme heterozygosity, low call rates, or cryptic relatedness. SNPs are filtered by call rate, Hardy-Weinberg equilibrium, and allele frequency thresholds.

Ethnic background is assessed through Principal Component Analysis (PCA), ensuring stratified analysis within population subgroups to avoid confounding. Genotype imputation is conducted using the TopMed imputation server, referencing comprehensive haplotype panels to enhance SNP density. Association studies are then conducted across multiple clinical and laboratory endpoints, including inflammation markers, silent infarctions, renal and pulmonary complications. Separate regression models are applied to each phenotype, yielding per-endpoint summary statistics and genotype matrices.

Final outputs include per-patient genotype profiles for significant SNPs and statistical results for each association. These are provided in CSV/tabular formats, ready for downstream machine learning modeling and integration with other patient metadata. The modularity and scalability of the workflow make it suitable for integration with other datasets or for longitudinal data tracking.

#### 4. Multiple Myeloma Workflow

In MM, the objective is to detect and quantify copy number aberrations (CNAs) from bone marrow-derived CD138+ plasma cells using SNP arrays. After sample enrichment and DNA extraction, SNP genotyping is performed on Affymetrix platforms (SNP6.0 or Cytoscan HD). Raw intensity data are processed using the RawCopy package, generating logR and B-allele frequency tracks. Segmentation is done using the PSCBS algorithm, applying stringent significance thresholds.

ASCAT is then used to correct for tumor purity and cellularity, producing genome-wide CNA profiles for each sample. These profiles are converted to integer copy number values using established formulas, applying thresholds to detect gains, losses, and homozygous deletions. Events are filtered based on cancer cell fraction and segment length. Samples with apparent genome doubling are re-normalized to diploid for simplicity.

GISTIC 2.0 is used to identify focal and broad alterations across the genome. Focal lesion analysis targets known driver regions such as TP53, MYC, and RB1. Only CNAs affecting 50% or more of tumor cells are retained to ensure analytical robustness. The result is a binary-coded table indicating the presence of specific CNAs at predefined loci, which can be merged with additional data and used for AI model training or patient stratification studies.

#### 5. Conclusion

D5.2 serves as the foundational document for the genomic data pre-processing architecture in GENOMED4ALL. The described pipelines ensure that diverse genomic data from multiple disease contexts are harmonized and standardized. This consistency is vital for the application of machine learning models across datasets originating from different platforms and clinical environments. Furthermore, the output formats are compatible with FL frameworks, enabling decentralized AI training while preserving data sovereignty. The pipelines will be shared with partners and the broader research community following related publications, contributing to open science and reproducibility. Ultimately, these workflows operationalize the integration of high-throughput genomics into clinical research, supporting personalized medicine initiatives within the scope of hematological disorders.

### Supplementary Information 22| Deliverable 5.4 - Framework for Data Pre-processing, Deployment, and Validation

#### 1. Aim and Current Progress

This document provides a comprehensive summary of the GenoMed4ALL pipeline for the analysis of genomic data from MDS. The pipeline encompasses data pre-processing, deployment on computing infrastructure, and rigorous validation procedures. Developed under the Horizon 2020 framework, this pipeline supports standardized and reproducible analysis across consortium partners, facilitating AI-based clinical support systems.

The GenoMed4ALL pipeline aims to transform raw genomic data obtained from patients diagnosed with MDS into clean, interpretable formats suitable for downstream analysis and clinical usage. The MDS pipeline is complete and functional, and its source code is prepared for distribution. Pipelines for MM and SCD are in development and will follow a similar deployment strategy.

During the project, all pipeline codes are hosted on a dedicated CINECA server. This infrastructure supports containerized versions of each pipeline, ensuring consistent environments and simplifying local execution by partner institutions.

#### 2. Use Case: MDS Genomic Analysis

The pipeline focuses on detecting somatic mutations in DNA extracted from either bone marrow or peripheral blood samples of patients with MDS. Leveraging deep sequencing (2000–3000X coverage), the process comprises demultiplexing, quality filtering, alignment, variant detection, annotation, and filtering. Outputs include detailed information on mutation type, position, frequency, and pathogenicity.

**3. Data Pre-processing Workflow**

The workflow comprises the following steps:

1. BCL to FASTQ Conversion: Raw data from Illumina sequencers in BCL format is demultiplexed using the 'bcl2fastq' tool, resulting in paired-end FASTQ files.

2. Quality Control: FASTQ files undergo quality assessment using FastQC. Trimming of adapters and low-quality bases is handled by Trimmomatic scripts, resulting in trimmed and quality-filtered FASTQ files.

3. Alignment: Reads are aligned to the hg38 human reference genome using alignment tools. Output is stored as BAM files after intermediate conversion to uBAM.

4. Variant Calling: Tools like Mutect2 and Pindel detect SNPs and structural variants. Mutect2 filters variants based on mapping artifacts using FilterMutectCalls. Outputs are saved in standardized VCF format.

5. Annotation: ANNOVAR annotates each variant with predicted impact, population frequency, and pathogenicity scores using data from SIFT, PolyPhen2, ClinVar, COSMIC, and gnomAD. Annotated results are stored in XLS format.

6. Variant Filtering: The final step integrates annotated VCFs into a single XLS file and filters variants using a prioritization algorithm. High-confidence oncogenic variants are flagged for clinical review.

**4. Deployment Strategy**

Each pipeline is encapsulated in a container including necessary dependencies. Users can execute the entire workflow using a single command-line script (`MDS_bioinf_pipeline.sh`). This guarantees reproducibility and simplifies setup. Execution options include remote analysis on the CINECA server or local deployment via Docker.

**5. Final Output and Clinical Application**

The final XLS file includes critical fields: variant coordinates, gene name, functional type, depth, VAF, and pathogenicity tags. Variants are classified using a prioritization system based on clinical relevance. AI tools ingest this data for predictive modeling and manual validation by clinicians remains essential before patient-level interpretation.

Each sample's intermediate and final files are stored in structured directories. Core gene mutation data and associated VAFs are key features for AI-based risk stratification and diagnostics.

### Supplementary Information 23| Deliverable 5.5 - Data Homogenization Framework

**1. Introduction**

The GENOMED4ALL project addresses a critical barrier in FL for healthcare: data heterogeneity. In a landscape where sensitive medical data such as EHRs, genomic profiles, and radiological images, must remain within institutional boundaries, FL offers a promising privacy-preserving alternative for developing machine learning models. Yet, the variation in structure, semantics, and quality of datasets across participating sites can severely impact model performance and interoperability.

To mitigate this challenge, the GENOMED4ALL Data Homogenization Platform provides a robust preprocessing framework that standardizes datasets into a unified format, based on HL7's Fast Healthcare Interoperability Resources (FHIR). This approach ensures semantic alignment and syntactic consistency across all partner institutions. By transforming diverse clinical, genomic, and radiological data into FHIR-compliant resources, the platform establishes a common data model that serves as a foundation for downstream machine learning and federated training tasks.

**2. Platform Architecture**

The platform architecture is organized into four key layers: the Data Integration Layer, the FHIR Modeling and Storage Layer, the User Layer, and the Feature Extraction Layer. Each layer plays a critical role in transforming raw, heterogeneous inputs into homogenized datasets that are ready for machine learning. The process begins with data ingestion through an Extract-Transform-Load (ETL) mechanism, which supports CSV inputs and connections to various database types including relational (Oracle) and non-relational (MongoDB, Elasticsearch) sources.

During transformation, data fields are mapped to corresponding FHIR resources using custom mappers and validation tools. Genomic mutations, for instance, are represented as FHIR 'Observation' resources using HGNC vocabularies and extended attributes such as number of mutations and tumor mutation load. Tools such as OHDSI’s Athena service support term harmonization and ensure consistent codification using standard vocabularies like LOINC and SNOMED CT.

**3. FHIR Implementation Guidelines**

Each clinical use case has its own FHIR implementation guideline. These define the exact mapping from source variables to FHIR resources. For example, 'gender' maps to the 'Patient' resource, while laboratory results map to 'Observation'. These mappings are documented using JSON schemas and are validated automatically during ingestion.

The original document illustrates the end-to-end data ingestion process, from CSV input to FHIR resource creation and storage. The process is modular and extensible, supporting the addition of new data types or institutional pipelines. The transformed data is stored using scalable NoSQL solutions such as Elasticsearch and MongoDB, enabling efficient access for subsequent extraction and model training tasks.

**4. Data Quality and Validation**

To ensure the reliability of the homogenized datasets, the platform generates both internal and external data quality reports. Using the Evidently AI framework, it computes key metrics such as the number of missing values, constant columns, duplicated rows, and column-wise correlation coefficients. These are evaluated against predefined thresholds—for example, any column with over 80% of identical values is flagged.

Two versions of the quality report are generated: a detailed HTML version for clinical validation and a lightweight JSON version shared with the FL platform. This dual-reporting structure balances the need for transparency with privacy considerations. The full document demonstrates how these reports highlight issues like constant values or highly correlated features.

**5. RESTful Services and User Interface**

To streamline usability and integration, a set of RESTful API endpoints and a web interface have been developed. Users can perform operations such as dataset ingestion, feature extraction, quality metric computation, and dataset registration directly from the interface. Each use case is provided its own module, and the system ensures that all transformations conform to the corresponding FHIR guidelines.

Endpoints include:

- /ingest: Ingests raw CSV into FHIR.

- /extract: Extracts features into a model-ready CSV.

- /metrics: Retrieves the data quality metrics.

- /register: Registers a custom dataset into the FL node.

These tools make it easy for hospitals to preprocess their data and contribute to federated training without exposing sensitive patient information. Additionally, datasets can be browsed, visualized, and verified through the web dashboard.

**6. Conclusion**

The GENOMED4ALL data homogenization framework provides the necessary infrastructure to ensure consistency, quality, and security of training data in FL settings. By leveraging standards like FHIR, tools like Evidently AI, and a modular architecture, it streamlines the preprocessing pipeline across multiple institutions. This lays a robust foundation for developing scalable, secure, and privacy-preserving AI models in healthcare.

### Supplementary Information 24| Deliverable 6.2 - Preliminary conclusions about federated learning applied to clinical data

**1. Introduction**

This deliverable collects the progress on FL achieved during the first year of the Genomed4All project, highlighting individual contributions from the involved partners. FL is introduced as a collaborative machine learning framework in which participating institutions train models locally on sensitive clinical data without sharing raw data. Instead, only essential model updates are communicated to a central server for aggregation, thereby ensuring privacy and regulatory compliance. The theoretical foundations of FL are laid out through an extended discussion on distributed convex optimization, with a focus on the Alternating Direction Method of Multipliers (ADMM), which serves as a basis for iterative learning across nodes. The report also introduces Partitioned Variational Inference as a distributed Bayesian estimation strategy, well-suited for non-convex models such as neural networks used in survival analysis, regression, and classification tasks.

UPM has led the development of a novel survival analysis method based on variational autoencoders, termed SAVAE. This approach combines generative modeling with predictive tasks and is currently being adapted for classification and clustering. Given the limitations of local data in FL settings, UPM is also experimenting with generative adversarial networks (GANs) to create synthetic, privacy-preserving "virtual patients" that emulate the statistical properties of real cohorts. This technique addresses challenges such as feature and distribution mismatching across centers.

UNIBO has focused on patient stratification through clustering, developing a federated implementation of Deep Embedded Clustering (DEC) using the Flower framework. Their model builds on parametric t-SNE and autoencoders to simultaneously learn compact feature representations and cluster assignments. The DEC model has been adapted for the MDS use case, and local model updates are aggregated through the FedAvg algorithm.

UNITO is advancing non-linear survival analysis by extending the traditional Cox proportional hazards model with neural networks. Recognizing the inadequacy of linear models for complex, real-world clinical data, they implemented three federated strategies (Sequential Federated, Weight Averaging, and Ensemble FL) on an MDS patient survival dataset. These models are evaluated under both balanced and unbalanced data distributions across centers, capturing practical deployment conditions.

UCPH is contributing by combining representation learning with transfer learning to reduce the dimensionality of clinical and omics data. They proposed a Deep Generative Decoder (DGD), a generative neural network that omits an encoder and instead relies on Maximum A Posteriori (MAP) estimation. This model is designed to handle missing data and imputation tasks within a federated framework. UCPH plans to integrate this model into the broader FL infrastructure and evaluate its performance on real-world datasets.

FORTH is exploring ensemble learning as a federated meta-learning approach, emphasizing the construction of ensemble models such as random forests. In their approach, individual nodes determine optimal decision tree splits, which are then aggregated by a central server. Despite the complexity of this distributed coordination, the method allows model updates to occur without disclosing class labels or raw data, maintaining privacy while enabling collaborative tree construction.

Together, these efforts illustrate a diverse and innovative application of FL across tasks including survival prediction, clustering, representation learning, and ensemble modeling, all designed to unlock the value of sensitive clinical data without compromising patient privacy.

**2. Federated Learning: An Overview**

Traditional deep learning methods rely on centralizing data for training, but this approach poses significant challenges in terms of privacy, scalability, and efficiency, especially in sensitive domains like healthcare. FL offers an alternative by enabling multiple institutions or devices to collaboratively train models without sharing raw data. Instead, each participant performs local training and only transmits model updates to a central server, which aggregates them to form a global model. This setup reduces privacy risks and accommodates the heterogeneous, non-independent and identically distributed (non-IID) nature of real-world data.

Initially proposed in 2016 to support distributed training across mobile devices, FL has since expanded to encompass both cross-device and cross-silo settings. Cross-device FL involves a large number of unreliable edge devices with limited connectivity and varying data quality. Cross-silo FL, more typical in healthcare, involves a smaller number of stable, institution-level participants such as hospitals, each managing siloed datasets. Compared to centralized approaches, FL systems are more decentralized, less synchronous, and must address issues related to client availability, communication efficiency, and data heterogeneity.

The architecture of FL includes three main components: local participants that train models on their data, a central coordinator that aggregates and redistributes model updates, and the communication-computation infrastructure that enables iterative training. The increasing demand for FL has spurred the development of several tools and frameworks designed to address its unique challenges. FATE offers a modular architecture with secure protocols and visualization tools, though it requires direct code customization for algorithm development. PaddleFL is designed around horizontal FL with a distributed training setup, still in early development stages with limited documentation. PySyft provides privacy-preserving deep learning capabilities using differential privacy and secure computation, supporting PyTorch and TensorFlow and following the FedAvg scheme for model aggregation. TensorFlow Federated offers easy-to-use federated computation primitives, but lacks built-in privacy mechanisms and is currently limited to single-device setups. IBM's FL framework focuses on enterprise settings, integrating well with Keras and TensorFlow while supporting secure multiparty computation. NVIDIA Clara, though not open-source, targets AI-powered healthcare applications and supports GPU acceleration and privacy-preserving techniques using a centralized architecture. To support evaluation and benchmarking in federated settings, LEAF provides federated datasets and example implementations, although it does not yet comprehensively address efficiency or privacy metrics. The development of robust benchmarks and tools remains a key area for future research as FL continues to mature and expand across domains.

As discussed, FL offers several advantages but also poses fundamental challenges, which is crucial to address to improve system performance.

One of the core challenges in FL is statistical heterogeneity. Data collected by clients are non-IID and originate from diverse contexts such as different locations, times, and user behaviors. This variability complicates modeling and increases the risk of stragglers during training. For example, features with the same label may differ across clients, or the distribution of labels itself may be highly skewed. To mitigate these issues, strategies such as personalized models, data augmentation, or limited data sharing (where privacy permits) can be employed. Algorithmic adaptations, including tuning hyperparameters or using modified versions of FedAvg like FedProx, also help. However, heuristic solutions that require data sharing may compromise privacy. An alternative approach is to shift from a single global model to personalized models tailored for each client, especially when client availability is inconsistent.

Privacy is another major challenge in FL. Although FL enhances privacy by keeping raw data local and only sharing model updates, these updates can still leak sensitive information through attacks such as gradient inversion. Beyond inference threats, other risks like data poisoning and model bias must be considered. Privacy must be maintained without significantly increasing computational costs or sacrificing model accuracy. Privacy-preserving techniques can be grouped into three categories. Secure computation approaches include Secure Multi-Party Computation (MPC) and Trusted Execution Environments (TEEs). MPC employs cryptographic methods such as homomorphic encryption to perform computations without exposing private inputs, while TEEs provide confidential and integrity-assured execution environments. Privacy-preserving disclosures involve Differential Privacy (DP), which injects noise into updates to obscure individual contributions, governed by privacy loss parameters. Lastly, verifiability ensures honest participation through methods like remote attestation and zero-knowledge proofs (ZKPs), although their practical application in FL remains challenging.

Systems heterogeneity also presents difficulties because clients differ widely in hardware capabilities, network connectivity, and power availability, resulting in inconsistent participation. Mitigation strategies include asynchronous communication, which helps address device stragglers but can suffer from unbounded delays. Active device sampling selects participants based on resource availability but typically assumes static system characteristics. Fault tolerance techniques, such as ignoring failures or using coded computation, improve resilience but may conflict with privacy constraints.

Communication overhead remains a significant bottleneck, especially in cross-device FL environments characterized by unreliable or slow connectivity. Three main strategies have been proposed to address this issue. Local updates reduce communication frequency by varying how often clients send updates, as exemplified by FedAvg. Compression schemes apply gradient or model compression techniques, including low-rank updates with Golomb encoding or lossy dropout, to reduce communication load. Decentralized training, which restricts communication to neighboring clients, can improve bandwidth efficiency and robustness in heterogeneous settings; however, this approach is often limited to linear models. Hierarchical models add scalability but depend on central aggregation points.

FL has attracted considerable interest as a method for privacy-preserving collaborative learning across distributed clients. While it holds significant promise, FL faces ongoing challenges related to data and system heterogeneity, privacy, communication constraints, and variability in client resources. Continued research is essential to adapt FL techniques effectively to specific data types, use cases, and system requirements.

**3. Centralized ADMM**

This chapter introduces convex optimization, focusing on supervised learning problems such as Ridge Regression, Lasso, Logistic Regression, and SVM, all unified under the Centralized ADMM algorithm. ADMM is well-suited for distributed implementations, which are described later.

A convex function $f_i$ satisfies:

$$f_i(\alpha x + \beta y) \leq \alpha f_i(x) + \beta f_i(y)$$

for all $x, y \in \mathbb{R}^n$ and all $\alpha, \beta \in \mathbb{R}$ where $\alpha, \beta \geq 0$ with $\alpha + \beta = 1$, ensuring at most one global minimum.

The general ML optimization problem is:

$$\min_{x\in\mathbb{R}^{d+1}} \frac{1}{N}\sum_{i=1}^{N} l(x^T u_i, y_i) + R(x)$$

where $l$ is a convex loss and $R$ a convex regularizer, often an $L_1$ or $L_2$ norm.

Key examples:

- Ridge Regression:

$$\min_x \frac{1}{N}\sum_{i=1}^{N}(x^T u_i - y_i)^2 + \frac{\lambda}{2}\|x\|_2^2$$

- Lasso:

$$\min_x(\frac{1}{N}\sum_{i=1}^{N}(x^T u_i - y_i)^2 + \lambda\|x\|_1)$$

- Logistic Regression:

$$\min_x(\frac{1}{N}\sum_{i=1}^{N}\ln\left(1 + e^{-y_i x^T u_i}\right) + \frac{\lambda}{2}\|x\|_2^2)$$

- SVM:

$$\min_x(\frac{1}{N}\sum_{i=1}^{N}\max\left(1 - y_i x^T u_i, 0\right) + \frac{\lambda}{2}\|x\|_2^2)$$

Non-differentiable functions like those in Lasso and SVM require proximal algorithms rather than gradient methods. ADMM is emphasized for its scalability and suitability in distributed convex optimization, starting from a centralized case and extending to distributed settings.

Starting from the convex problem:

$$\min f(x) \quad \text{s.t.} \quad Ax = b$$

Define the augmented Lagrangian to ensure convergence without assuming strict convexity:

$$\mathcal{L}_\rho(x, y) = f(x) + y^T(Ax - b) + \frac{\rho}{2}\|Ax - b\|_2^2$$

The method of multipliers updates are:

$$x_{k+1} = \arg\min_x \mathcal{L}_\rho(x, y_k)$$

$$y_{k+1} = y_k + \rho(Ax_{k+1} - b)$$

ADMM generalizes this to problems of the form:

$$\min_{x,z} \; g(x) + h(z) \quad \text{s.t.} \quad Ax + Bz = c$$

The augmented Lagrangian becomes:

$$\mathcal{L}_\rho(x, z, y) = g(x) + h(z) + y^T(Ax + Bz - c) + \frac{\rho}{2}\|Ax + Bz - c\|_2^2$$

With iterative updates:

$$x_{k+1} = \arg\min_x \mathcal{L}_\rho(x, z_k, y_k)$$

$$z_{k+1} = \arg\min_z \mathcal{L}_\rho(x_{k+1}, z, y_k)$$

$$y_{k+1} = y_k + \rho(Ax_{k+1} + Bz_{k+1} - c)$$

Using the scaled dual variable $w = \frac{1}{\rho}y$, we can rewrite the updates as:

$$x_{k+1} = \arg\min_x \left(g(x) + \frac{\rho}{2}\|Ax + Bz_k - c + w_k\|_2^2\right)$$

$$z_{k+1} = \arg\min_z \left(h(z) + \frac{\rho}{2}\|Ax_{k+1} + Bz - c + w_k\|_2^2\right)$$

$$w_{k+1} = w_k + Ax_{k+1} + Bz_{k+1} - c$$

To solve a problem of the form:

$$\min_x f(x) = \min_x (g(x) + h(x))$$

We rewrite it as:

$$\min_{x,z} (g(x) + h(z)) \quad \text{s.t.} \quad x - z = 0$$

Which gives us the ADMM updates:

$$x_{k+1} = \arg\min_x \left(g(x) + \frac{\rho}{2}\|x - z_k + w_k\|_2^2\right)$$

$$z_{k+1} = \arg\min_z \left(h(z) + \frac{\rho}{2}\|x_{k+1} - z + w_k\|_2^2\right)$$

$$w_{k+1} = w_k + x_{k+1} - z_{k+1}$$

ADMM for Lasso (Example)

We aim to solve:

$$\min_{x,z} \left(\frac{1}{N}\|Ax - b\|_2^2 + \lambda\|z\|_1\right) \quad \text{s.t.} \quad x - z = 0$$

Where:

$$g(x) = \frac{1}{N}\|Ax - b\|_2^2$$

$$h(z) = \lambda\|z\|_1$$

The augmented Lagrangian is:

$$\mathcal{L}_\rho(x, z, y) = \frac{1}{N}\|Ax - b\|_2^2 + \lambda\|z\|_1 + y^T(x - z) + \frac{\rho}{2}\|x - z\|_2^2$$

Using the scaled dual variable, the updates become:

$$x_{k+1} = \left(\frac{2}{N}A^T A + \rho I\right)^{-1}\left(\frac{2}{N}A^T b - \rho(z_k - w_k)\right)$$

$$z_{k+1} = \text{soft}(x_{k+1} + w_k, \frac{\lambda}{\rho})$$

$$w_{k+1} = w_k + x_{k+1} - z_{k+1}$$

With soft thresholding defined as:

$$\text{soft}(a, b) = (a - b)_+ - (-a - b)_+$$

Where $(x)_+ = \max(0, x)$.

We consider the following consensus optimization problem:

$$\min_x f(x) = \sum_{i=1}^{N_b} g_i(x)$$

This is reformulated with local variables and a global consensus constraint:

$$\min_{x_i} \sum_{i=1}^{N_b} g_i(x_i) \quad \text{s.t. } x_i - z = 0 \quad \forall i$$

**Augmented Lagrangian**:

$$\mathcal{L}_\rho(x_1, \ldots, x_{N_b}, z, y_1, \ldots, y_{N_b}) = \sum_{i=1}^{N_b}\left(g_i(x_i) + y_i^\top(x_i - z) + \frac{\rho}{2}\|x_i - z\|_2^2\right)$$

**ADMM Updates**:

$$x_{i,k+1} = \arg\min_{x_i}\left(g_i(x_i) + y^\top{}_{i,k}(x_i - z_k) + \frac{\rho}{2}\|x_i - z_k\|_2^2\right)$$

$$z_{k+1} = \frac{1}{N_b} \sum_{i=1}^{N_b} \left( x_{i,k+1} + \frac{1}{\rho} y_{i,k} \right)$$

$$y_{i,k+1} = y_{i,k} + \rho(x_{i,k+1} - z_{k+1})$$

**Scaled ADMM**

With the scaled dual variable $w_{i,k} = \frac{1}{\rho} y_{i,k}$, the update steps become:

$$x_{i,k+1} = \arg\min_{x_i} \left( g_i(x_i) + \frac{\rho}{2} \|x_i - z_k + w_{i,k}\|_2^2 \right)$$

$$z_{k+1} = \arg\min_{z} \left( h(z) + \frac{N_b \rho}{2} \|z - \bar{x}_{k+1} - \bar{w}_k\|_2^2 \right)$$

$$w_{i,k+1} = w_{i,k} + x_{i,k+1} - z_{k+1}$$

Where:

$$\bar{x}_k = \frac{1}{N_b} \sum_{i=1}^{N_b} x_{i,k}, \quad \bar{w}_k = \frac{1}{N_b} \sum_{i=1}^{N_b} w_{i,k}$$

*Federated Learning Interpretation (via ADMM)*

*Centralized ADMM*

In the centralized formulation, ADMM iteratively updates the variables $x$, $z$, and $w$ through the following sequence:

$$x_{k+1} = \arg\min_{x} \left( g(x) + \frac{\rho}{2} \left\| x - z_k + w_k \right\|_2^2 \right)$$

$$z_{k+1} = \arg\min_{z} \left( h(z) + \frac{\rho}{2} \left\| x_{k+1} - z + w_k \right\|_2^2 \right)$$

$$w_{k+1} = w_k + x_{k+1} - z_{k+1}$$

Where $g(x)$ and $h(z)$ may admit analytical or gradient-based solutions.

The update process can be compactly represented using a stacked variable vector:

$$\theta^k = \begin{bmatrix} x^k \\ z^k \\ w^k \end{bmatrix}$$

Under convexity assumptions, this iterative method guarantees convergence:

$$\theta^* = \lim_{k \to \infty} \theta_{i,k} \quad \forall i$$

*Federated ADMM*

In a distributed or FL scenario, the same optimization framework is applied locally at each client (node), with partial synchronization through a shared global variable $z$. The updates are as follows:

$$x_{i,k+1} = \arg\min_{x_i} \left( g_i(x_i) + \frac{\rho}{2} \|x_i - z_k + w_{i,k}\|_2^2 \right)$$

$$z_{k+1} = \arg\min_{z} \left( h(z) + \frac{N_b \rho}{2} \|z - \bar{x}_{k+1} - \bar{w}_k\|_2^2 \right)$$

$$w_{i,k+1} = w_{i,k} + x_{i,k+1} - z_{k+1}$$

Where:

$$\bar{x}_k = \frac{1}{N_b} \sum_{i=1}^{N_b} x_{i,k}, \quad \bar{w}_k = \frac{1}{N_b} \sum_{i=1}^{N_b} w_{i,k}$$

Each local node maintains its own variable vector:

$$\theta_{i,k} = \begin{bmatrix} x_{i,k} \\ z_k \\ w_{i,k} \end{bmatrix}$$

Despite distributed computation, all local iterations are guaranteed to converge to the global optimum:

$$\theta^* = \lim_{k \to \infty} \theta_i^k, \quad \forall i$$

In this federated setting, data remains private at each local node. Only partial updates $(x_{i,k}, w_{i,k})$ are communicated, while the central server coordinates through the shared $z_k$, ensuring convergence consistency across nodes.

**4. Non-Convex Optimization. Distributed Implementation**

In the context of FL, while convex optimization allows efficient and fast convergence (particularly with methods like ADMM) it lacks the expressive power needed for complex data modeling, such as in medical applications. To better capture such complexities, neural networks are introduced due to their universal approximation capabilities. However, their associated optimization problems are inherently non-convex, and naive approaches like directly averaging weights across distributed clients fail due to data heterogeneity.

To address this, the weights of neural networks are treated as random variables within a Bayesian framework, specifically through Bayesian Neural Networks (BNNs). Rather than producing a single prediction, this approach yields a predictive distribution, offering not only point estimates but also uncertainty quantification. This is especially useful in sensitive applications like healthcare.

The Bayesian formulation focuses on computing the posterior predictive distribution, which is generally intractable. To approximate it, techniques such as Maximum A Posteriori (MAP) estimation or, more robustly,

Variational Inference (VI) are used. VI replaces the intractable posterior with a tractable, parameterized distribution that is optimized to be close (in terms of Kullback-Leibler divergence) to the true posterior.

In practical implementations, especially in high-dimensional neural networks, VI is simplified using the mean-field assumption, often modeling the distribution as independent Gaussians. Optimization of the variational parameters is then carried out using techniques such as Bayesian Backpropagation or black-box stochastic optimization.

The result is a tractable and distributed approach to non-convex optimization, where each client contributes to a global variational model. The final prediction is obtained via Monte Carlo sampling from the optimized variational distribution, enabling decentralized learning while maintaining probabilistic rigor.

*Posterior Factorization in Federated Learning*

In a distributed setting, the global posterior over model parameters $w$ given data $D$ can be factorized as:

$$p(w \mid D) = \frac{p(w) \prod_n p(y_n \mid w, x_n)}{p(y \mid x)} \propto p(w) \prod_n p(y_n \mid w, x_n)$$

This decomposition allows local nodes (clients) to work on their own data, approximating their contribution to the global posterior, while a central server handles the prior and overall aggregation.

*Partitioned Variational Inference (PVI): Basic Idea*

To approximate the posterior distribution, PVI proposes:

$$p(w \mid D) \approx g(w) \propto g_0(w) \prod_{n=1}^{N} g_n(w)$$

Where:

- $g_0(w) \approx p(w)$ (handled by the server),
- $g_n(w) \approx p(y_n \mid w, x_n)$ (handled by each node).

Each node aims to optimize a local approximation term $g_n(w)$, and uses the cavity distribution:

$$g_{-i}(w) = \frac{g(w)}{g_i(w)} = \prod_{n \neq i} g_n(w)$$

*Main Definitions and Optimization*

Global Variational Inference (GVI) Objective:

$$F_{\text{GVI}}(q(w)) = -\text{KL}(q(w) \| p(w)) + \sum_{n=1}^{N} \mathbb{E}_{q(w)}[\log p(y_n \mid w, x_n)]$$

Local PVI Objective at Node $n$:

$$F^n_{\mathrm{PVI}}(q(w)) = -\mathrm{KL}(q(w) \| q^n_{-n}(w)) + \mathbb{E}_{q(w)}[\log p(y_n \mid w, x_n)]$$

Where:

- $q^n_{-n}(w)$ is the cavity distribution, used as a local prior at node $n$.
- PVI proves that maximizing each local objective leads to maximizing the global ELBO.

*PVI Algorithm Steps*

At node $i$, at time $t$:

1. Compute Cavity Distribution:
$$q^{i,t}_{-i}(w) = \frac{q^{i,t}(w)}{q^{i,t}_i(w)}$$
2. Local Update (solve PVI problem):
$$q^{i,t+1}(w) = \arg\min_{q(w)} F^i_{PVI}(q(w))$$
3. Send Update to Server:
$$\tilde{q}^{i,t+1}_i(w) = \frac{q^{i,t+1}(w)}{q^{i,t}_i(w)}$$
4. Server Update & Broadcast:
$$q^{t+1}(w) = q^t(w) \cdot \frac{\tilde{q}^{i,t+1}_i(w)}{\tilde{q}^{i,t}_i(w)}$$

This structure is conceptually related to Expectation Propagation (EP), but PVI offers a convergent version via distributed updates.

**5. Federated Data Augmentation (FDA)**

FL with neural networks offers a powerful framework for distributed modeling, but it faces practical challenges when dealing with complex, high-dimensional medical data. Calculating product and quotient distributions is often infeasible, and solving variational inference problems across multiple nodes can be computationally intensive. To overcome the limitations caused by scarce and heterogeneous local data, an alternative approach is proposed: instead of sharing real patient data (which is prohibited), nodes can share latent representations to generate synthetic, privacy-preserving virtual patients.

This idea is implemented using RadialGAN, a GAN architecture specifically designed to address two main issues in federated medical datasets: feature mismatch and distribution mismatch. Feature mismatch occurs when different hospitals collect different sets of variables, while distribution mismatch arises from differences in patient populations across hospitals.

RadialGAN introduces a common latent space into which all datasets are encoded. Each hospital uses an encoder to map its data into this shared latent space and a decoder to reconstruct data from it. This setup enables translation of patient data between domains while maintaining semantic consistency and privacy. By operating in a latent space, the model overcomes the inconsistencies in feature availability and distribution among datasets.

Formally, for each hospital $i$, the encoder $F_i : X^{(i)} \times Y \to Z$ maps input features and labels to the latent space, and the decoder $G_i : Z \to X^{(i)} \times Y$ reconstructs them. The augmented dataset for hospital $i$ is constructed by combining its original data $D_i$ with translated data from other hospitals:

$$D_i^e = D_i \cup \bigcup_{j \neq i} G_i(F_j(D_j))$$

Training RadialGAN involves optimizing two objectives. The adversarial loss ensures the generated data is indistinguishable from real data:

$$L_{adv}^i = \mathbb{E}[\log D_i(X^{(i)}, Y)] + \sum_{j \neq i} \alpha_{ij} \mathbb{E}[\log(1 - D_i(G_i(F_j(X^{(j)}, Y))))]$$

Here, $\alpha_{ij}$ weights contributions from other hospitals, often based on dataset sizes. The cycle-consistency loss guarantees that translating data to the latent space and back yields results close to the original:

$$L_{cyc}^i = \mathbb{E}\left[\|(X^{(i)}, Y) - G_i(F_i(X^{(i)}, Y))\|^2\right] + \sum_{j \neq i} \alpha_{ij} \mathbb{E}\left[\|F_j(X^{(j)}, Y) - F_i(G_i(F_j(X^{(j)}, Y)))\|^2\right]$$

The total objective is a minimax optimization problem across all hospitals:

$$\min_{G,F} \max_{D} \sum_{i=1}^{M} L_{adv}^i + \lambda \sum_{i=1}^{M} L_{cyc}^i$$

where $\lambda$ balances adversarial and consistency terms.

Once the encoders and decoders are trained, each hospital augments its local dataset using synthetic data generated via the shared latent space. Final prediction models $f_i$ are then trained independently on these expanded datasets using any standard machine learning method.

**6. UPM Contribution Summary**

UPM introduces a unified generative model framework based on VAEs, optionally combined with adversarial training, to address both supervised tasks,such as classification, stratification, and survival analysis, and unsupervised tasks, including clustering and representation learning. It is designed for FL environments, enabling multiple decentralized nodes to collaboratively learn a shared latent space while preserving data privacy.

In a single-node setup, UPM uses a standard VAE with a learnable prior to increase modeling capacity. The model learns a latent variable $z$ that explains observed data $x$ via the generative process $p_\theta(x|z)$, while an inference network $q_\phi(z|x)$ approximates the posterior. Training maximizes the Evidence Lower Bound (ELBO):

$$\mathcal{L}(\theta, \phi; x) = \mathbb{E}_{q_\phi(z|x)}[\log p_\theta(x|z)] - \mathrm{KL}(q_\phi(z|x) \| p(z))$$

In centralized multi-node setups, the same encoder-decoder architecture is shared across institutions. In the federated version, each site has private encoder-decoder pairs $(\phi_i, \theta_i)$, and local latent variables $z_i$, but all sites aim to approximate a shared latent prior $p(z)$ via communication-efficient aggregation.

Generative models such as VAEs offer a principled framework to jointly learn data generation and low-dimensional representations. The latent space can be structured for downstream tasks: clustering (via mixture priors), imputation (by integrating out missing values), and semi-supervised learning (by conditioning on partial labels). This unified latent space allows supervised losses to be easily combined with the generative ELBO, enabling robust multitask learning. UPM extends the VAE to time-to-event modeling through the

Survival Analysis VAE (SAVAE). For an input $x$, a latent representation $z \sim q_\phi(z|x)$ is inferred, and the time-to-event $t$ is modeled as drawn from a Weibull distribution conditioned on $z$:

$$t \sim \text{Weibull}(k(z), \lambda(z))$$

where $k(z)$ and $\lambda(z)$ are learned functions of $z$. The joint likelihood becomes:

$$p_\theta(x, t|z) = p_\theta(x|z) \cdot p_\theta(t|z)$$

and the modified ELBO objective incorporates both data reconstruction and survival likelihood:

$$\mathcal{L}(\theta, \phi; x, t) = \mathbb{E}_{q_\phi(z|x)}[\log p_\theta(x|z) + \log p_\theta(t|z)] - \text{KL}(q_\phi(z|x)\|p(z))$$

This formulation enables the model to predict survival distributions for new patients by marginalizing over the latent space:

$$p(t|x) = \int p(t|z) q_\phi(z|x)\, dz$$

Through this generative approach, it is possible to learn meaningful patient representations that support a broad range of clinical inference tasks within both centralized and federated data regimes.

*Coupling Learning Across Nodes in VAEs*

Two main strategies were explored to couple learning across federated VAE nodes: jointly shaping the latent prior and using virtual patient generation. Initial attempts to jointly define the prior across nodes, by aligning each node's aggregated posterior $q_\phi(z)$ with a shared prior $p_\lambda(z)$, did not yield strong performance. This strategy relies on the optimal condition

$$p_\lambda^*(z) = \frac{1}{N} \sum_{n=1}^{N} q_\phi(z|x_n)$$

which can be approximated using learnable pseudopoints $\alpha^{(k)}$, allowing a parameterized prior that doesn't directly depend on private data:

$$p_\lambda(z) \approx \frac{1}{K} \sum_{k=1}^{K} q_\phi(z|\alpha^{(k)})$$

In the federated setting, each node maintains its own encoder–decoder while sharing a global prior constructed as a product of local components:

$$p_\lambda(z) \approx \prod_{m=1}^{M} p_{\lambda_m}(z)$$

This product-of-experts approach allows nodes to synchronize via shared pseudopoints or by exchanging fitted parameters of a global prior model. More promising results were obtained with virtual patient generation, where high-performing nodes synthesize data to augment low-resource sites. Models such as RadialGAN enable mappings between feature distributions, enforcing a common latent space via adversarial and cycle-consistency losses. The process augments each local dataset as

$$D_i' = D_i \cup \{G_i(F_j(D_j))\}_{j \neq i}$$

which allows weak nodes to infer missing features and improve performance in both supervised and unsupervised tasks.

*Early Results and Evaluation*

FL in healthcare faces the challenge of data heterogeneity because nodes may have few samples, missing features, or both. Two strategies were explored: copying models from good nodes, which fails when feature sets differ significantly, and using synthetic data (virtual patients) from strong nodes to augment weak nodes. The latter enables feature imputation and more robust training, effectively acting as a denoising process. Evaluation must be sensitive to imbalance. Performance should be compared across three cases: good nodes trained in isolation (upper bound), bad nodes trained in isolation (lower bound), and bad nodes using federated augmentation (expected to approach the upper bound). This setup helps assess whether FL yields real improvements, especially in difficult conditions.

**7. UniBO Contribution Summary**

The UniBO contribution discusses the use of clustering techniques for patient stratification based on multi-omics data, such as genomic and imaging measurements, to predict clinical outcomes, particularly in hematological diseases. A notable example is the application of hierarchical Dirichlet process (HDP) clustering in MDS, which is favored due to its Bayesian non-parametric approach that automatically determines the number of clusters and fits well with the data format combining genomic, cytogenetic, and clinical variables. These variables are simplified into mutation presence/absence indicators, making patient data suitable for multinomial modeling. Despite HDP's success, no federated implementation currently exists, making federated approaches like Latent Dirichlet Allocation (LDA) (a related model) important benchmarks, with HDP considered the natural non-parametric extension of LDA. Clustering methods broadly fall into partitioning, hierarchical, fuzzy, and density-based categories, and can involve soft or hard membership. Dimensionality reduction techniques like t-SNE and UMAP are valuable for high-dimensional data visualization but lack federated versions, presenting an area for future work. The MDS dataset from GenoMed4All includes demographic, clinical, prognostic, cytogenetic, genomic, and outcome variables, organized by patient rows and feature columns, and includes specific scores and survival data. The HDP analysis identifies patient-specific latent components that reflect underlying biological heterogeneity.

Federated clustering is crucial for advancing personalized medicine by grouping patients based on clinical, phenotypical, demographic, and omics data to better predict disease outcomes and treatment responses. Large-scale, multi-center studies are necessary to achieve this, but privacy concerns and data sharing restrictions pose significant challenges. FL offers a solution by enabling collaborative analysis across centers without exposing private data, though most traditional clustering methods need significant adaptation to work in this setting. The authors focused on deep learning models, particularly the DEC approach, adapted for federated use via the Flower framework. DEC combines deep autoencoders to reduce data dimensionality and learn meaningful feature representations with clustering performed in this reduced space. Training DEC involves two stages: first, pretraining a stacked autoencoder to capture useful features, and second, optimizing cluster assignments by iteratively refining cluster centroids and the neural network parameters through minimizing the Kullback-Leibler divergence between soft cluster assignments and an auxiliary target distribution. This process leverages a Student's t-distribution kernel to measure similarity in the embedded space and relies on self-training, where confident predictions improve the model iteratively. Gradients from this optimization are backpropagated through the network, and training concludes when cluster assignments stabilize.
Training the DEC model on MDS data in a federated setting requires adaptations from the centralized approach. The initial autoencoder training uses the FedAvg algorithm to aggregate weights across clients, while the cluster centroid initialization is more complex because different clients may find different centroids when running k-means locally. To address this, each client runs k-means with multiple seeds, then sends the centroids to a central server that aggregates them by selecting a diverse set covering the feature space, which are then shared back for clustering initialization. The clustering layer is trained for 10,000 epochs with periodic updates of the auxiliary distribution using stochastic gradient descent. Adjustments were made to the network architecture,

including replacing dropout with a random flipping layer to better handle binary MDS data and reducing layer sizes to fit the dataset. The full dataset of 2043 samples was split equally among varying numbers of clients, with local train-test splits. Performance was evaluated using metrics on reconstruction accuracy for the autoencoder and cluster assignment stability for clustering. Different model configurations were tested by varying dropout, random flip rates, normalization, and latent space dimension. Results showed that normalization had the most significant impact on training stability, and federated training with multiple clients did not substantially degrade clustering quality. Finally, the federated DEC clustering was compared to the HDP, the current standard for MDS data clustering, and while DEC improved silhouette scores due to its optimization objectives, its cluster assignments remained consistent across different federation setups, demonstrating robustness and comparable performance to centralized methods.

**8. UniTO Contribution Summary**

The UniTO contribution focuses on survival analysis, which predicts the timing of events such as adverse outcomes in clinical settings. The widely used Cox proportional hazards model estimates the hazard function, representing the risk of an event occurring at a specific time given patient features. This model expresses the hazard as $h(t|x_i) = h_0(t) \exp(x_i^\top \beta)$, where $h_0(t)$ is a baseline hazard independent of individual features, and $\beta$ are coefficients estimated by maximizing the partial likelihood function. This likelihood considers the relative risk among patients who are event-free up to a given time and is defined as

$$\hat{\beta} = \arg\max_{\beta} \sum_{i=1}^{n} \delta_i \left( x_i^\top \beta - \log \sum_{j \in R_i} \exp(x_j^\top \beta) \right),$$

where $R_i$ is the risk set at time $t_i$, and $\delta_i$ indicates if the event was observed (uncensored). The baseline hazard cancels out in this formulation, simplifying parameter estimation.

Since the linearity assumption of Cox may be restrictive for complex clinical data, a nonlinear extension called DeepCox replaces the linear predictor with a neural network output, using the same partial likelihood as a loss function to train the network. This allows modeling nonlinear effects of covariates and can be adapted to different data types, such as images or time series.

Evaluations of survival models use the CI, which measures the agreement between predicted risk scores and observed survival times, accounting for censored data. CI calculates the proportion of concordant pairs where the subject with the higher predicted risk indeed has a shorter survival time, generalizing the ROC AUC to censored survival data.

The development and evaluation of privacy-preserving FL strategies was assessed for survival analysis, particularly in contexts where patient data are distributed across multiple hospitals and cannot be centralized. Three FL architectures were proposed and tested using data from MDS patients.

A key challenge in applying FL to the Cox proportional hazards model is that its loss function is non-separable; it depends on risk sets that span multiple patients, making it difficult to compute when data are siloed in separate institutions. To address this, the authors approximate the risk sets locally within each centre, effectively masking the full risk set matrix by only retaining the diagonal blocks corresponding to local patients. This leads to different behavior depending on whether centers are balanced (equal size) or unbalanced.

The first federated strategy, Sequential Federated, adapts the FedAvg method by training a single model sequentially across all centers. The model starts with an initial parameter set and is trained at one centre for a fixed number of epochs before being passed to the next, repeating this process for several federated epochs with randomized centre orders.

The second approach, Weighted Averaging, trains the same model in parallel across all centers. Each centre updates the model locally, and the parameter updates are then aggregated via a weighted average, favoring centers with more data, to form a new global model for the next round.

The third, Ensemble Federated, does not aggregate models but instead combines the predictions from independently trained local models. Each of the $K$ centers trains its own model, and their outputs are averaged to generate final predictions. Unlike the other two, this method avoids any shared model and does not use federated epochs, but it still respects data privacy.

For model validation, a centre-based K-Fold Cross-Validation was used. In each fold, one centre's data is held out as the test set, while the others are used for training. Since the CI is a relative measure that depends on comparisons across the full dataset, predictions from each fold's test set are aggregated to compute a reliable CI score over the entire population. This approach supports hyperparameter tuning while maintaining strict data isolation.

The results of applying FL to survival analysis were collected. The baseline model is a feedforward neural network with two hidden layers composed of 8 and 4 neurons respectively. The model uses both L1 and L2 regularization on the first hidden layer to mitigate overfitting. An adaptive learning rate strategy is adopted, decreasing the learning rate at each iteration where the loss does not improve, following the rule:

$$\mathrm{lr}_{i+1} = \max\left(\frac{\mathrm{lr}_i}{\sqrt{i+1}}, 0.001\right)$$

Hyperparameters, including the number of federated epochs, internal epochs, internal batches, and the initial learning rate, were selected through a cross-validation procedure applied only to the training data.

The evaluation was conducted on data from the Bersanelli et al. study on MDS(20), which includes 2,043 patients with 70 covariates spanning demographic, clinical, genetic, and cytogenetic information. The target variable was Overall Survival (OS), defined as the time to death, with 28.9% of patients uncensored. All features were standardized to the [0, 1] range. The dataset was randomly split into a training set (75%) and a test set (25%). To simulate FL, the training data were distributed across eight synthetic centers. Though these centers were simulated within the same physical machine, the goal was to replicate the conditions of decentralized data storage and processing.

Four scenarios were considered to evaluate the impact of data distribution on federated model performance. In the balanced IID scenario, all centers had an equal number of patients and identical distributions of covariates and survival times. In the unbalanced IID scenario, centre sizes varied, with one centre holding 40% of the patients, but the distributions of features remained IID. In the balanced Non-IID survival times scenario, centers had equal numbers of patients, but each was assigned a non-overlapping segment of the survival time distribution. Similarly, in the balanced non-IID BMB scenario, all centers had the same number of patients, but the distribution of one key feature (Bone Marrow Blasts percentage) was split across centers in non-overlapping intervals. This feature was chosen through feature importance analysis as the most predictive for the neural network. In both non-IID scenarios, hard thresholds were applied to simulate worst-case heterogeneity.

Centre-based K-Fold cross-validation was used for model selection only on training data, and performance was evaluated on the blind test set, which was treated as an unbiased, standalone centre. The evaluation was repeated 20 times using different train-test splits to ensure robustness.

The results were measured using the CI. In the centralized case, which serves as the upper bound, the model achieved a CI of 0.75 with a standard deviation of 0.02. When models were trained independently at each centre without federated coordination (isolated case), the CI dropped to 0.62 for both balanced and unbalanced IID scenarios. Applying federated strategies in these IID settings (Sequential, Weights Averaging, and Ensemble) all yielded performance close to the centralized upper bound, with CIs around 0.74 to 0.75 and very low standard deviations, indicating strong consistency.

Under non-IID conditions, model performance decreased. For non-IID survival times, isolated training yielded a CI of 0.55. The federated approaches improved on this baseline, with Weights Averaging achieving the highest CI of 0.68. In the non-IID BMB scenario, isolated training gave a CI of 0.59, whereas federated methods again improved performance, with Weights Averaging reaching 0.73. These results show that FL can still yield significant gains over isolated learning, even under challenging distribution shifts. Among the three federated strategies, Weights Averaging consistently demonstrated the most robustness across all settings.

The study concludes that it is feasible to apply FL to clinical survival analysis with encouraging results, even under non-IID data distributions. Furthermore, the study will be expanded by experimenting with more complex neural architectures capable of handling different data modalities such as medical images or temporal sequences. In these results, a plan is also discussed to incorporate more realistic FL constraints, such as centers being intermittently unavailable during training, and to explore survival prediction tasks using imputed data to expand the dataset. Additional survival endpoints like Leukemia-Free Survival (LFS) and Event-Free Survival (EFS) will be included in future analyses. Finally, the need for further investigation into how heterogeneous local data distributions influence performance, particularly due to the weight divergence that can arise in non-IID settings, is acknowledged.

### 9. UCPH Contribution Summary

UCPH is developing and refining the DGD, a probabilistic machine learning model designed to handle the challenges of medical data, such as small size, noise, heterogeneity, and missing values. Instead of relying on a traditional encoder, the DGD learns data representations through MAP estimation, making it simpler and more stable than comparable models like the VAE. It performs well in image reconstruction and generation tasks, and is particularly suited for representation learning, imputation of missing values, and FL. Within the GenoMed4ALL project, UCPH aims to test and extend the DGD, including evaluating how its learned representations support downstream tasks like classification or survival analysis. The model's design also supports privacy, as only model parameters, not raw data, need to be shared across nodes in a federated setting.

### 10. FORTH Contribution Summary

The FORTH contribution focuses on adapting the XGBoost gradient tree boosting algorithm for FL. Gradient boosting builds powerful models by combining many weak learners, typically decision trees, trained iteratively to minimize errors using gradient information from a loss function. In the federated setting, however, directly sharing the data needed for optimal splits (like residuals and prediction statistics) can compromise privacy.

To address this, FORTH proposes a privacy-preserving approach: each local node computes the top S best candidate splits based on a gain metric and sends just these, along with their scores and sample counts, to a central server. The server then selects the global best split using a weighted score that balances performance across nodes. This way, sensitive details are kept local, and communication overhead is reduced.

Once the trees are constructed, leaf weights are computed using aggregated statistics from each node, again minimizing exposure of raw data. The entire federated training process ensures all nodes work on building a shared, consistent model while maintaining privacy and efficiency.

### 11. Conclusion

This report represents a significant step forward in the project's development, as it has helped all partners gain deeper experience with FL architectures. Through a series of regular meetings, the consortium clarified the scope and limitations of various federated approaches, considering a wide range of ML problems from supervised to unsupervised scenarios. As a result, each partner now has a clearer understanding of the directions they wish to pursue. Several important needs for the next phase have also been identified. A unified codebase with standardized procedures for data generation and result analysis is essential to allow fair comparison between different approaches. There is also a recognized need for better coordination with other WP leaders to ensure that AI initiatives address all the key challenges posed by the case studies. Enhanced collaboration between partners could lead to productive synergies, especially given that many of the approaches share similar architectural foundations. Another major insight is the necessity of bringing AI work closer to clinical practice by focusing on tools that enhance interpretability and explainability of what are often black-box models. Finally, the project needs an objective testing framework to assess whether the federated implementations are genuinely effective. Such a framework should clearly differentiate performance between nodes with complete data and strong individual models, and nodes with limited, sparse data, where FL is expected to provide significant improvement. This contrast will serve as a concrete measure of the added value of federation.

### Supplementary Information 25| Deliverable 6.3 - HMs staging and output prediction

#### 1. Use case Myelodisplastic Syndrome

In the context of MDS, the implementation of data-driven methodologies is met with a number of persistent challenges, particularly in the acquisition and use of health data. Despite the transformative potential of AI and Data Science (DS) in healthcare (owing to their adaptability and capacity to manage complex data structures) their integration is often hindered by constraints in data access, standardization, and privacy regulation compliance.

Access to patient-level data remains one of the foremost barriers. Medical datasets typically include highly sensitive and personally identifiable information such as medical histories, genetic profiles, and ongoing treatment records. Regulatory and ethical obligations surrounding patient privacy inhibit data sharing, thereby limiting collaborative research and innovation. Anonymization is the prevailing strategy to mitigate these concerns, yet it is often manual, inconsistent, and insufficient to ensure irreversibility of re-identification.

A second critical limitation is the lack of harmonization across healthcare datasets. The absence of standardized data formats and shared anonymization protocols makes the integration of heterogeneous data sources particularly challenging. This is further complicated by the scarcity of comprehensive datasets for rare conditions such as MDS, which leads to issues of data sparsity and class imbalance, conditions that degrade the predictive reliability of machine learning models.

To address these obstacles, synthetic data has emerged as a compelling alternative. Synthetic datasets are artificially generated by models trained to replicate the statistical properties of real data without reproducing actual instances. Consequently, these datasets are exempt from many of the regulatory restrictions governing real patient data and can be shared more freely. They also enable the augmentation of existing datasets to address class imbalance and can support hypothesis generation and validation in settings where real data are limited.

The process of synthetic data generation can be generalized into three stages: model training on real data, synthetic sample generation, and validation. The generative models can be applied across multiple modalities, including structured data, imaging, and sequencing outputs. Once trained, they provide a scalable, low-cost avenue for producing high-dimensional synthetic datasets that preserve the key characteristics of the original distribution.

*HDP on real and synthetic datasets*

To validate the fidelity of synthetic data in the MDS domain, we applied a one-level HDP clustering algorithm to both real and synthetic patient datasets. The HDP is a Bayesian non-parametric model suitable for unsupervised clustering, particularly when the number of clusters is not known a priori. It relies on the Chinese Restaurant Process (CRP) metaphor, where the probability of joining an existing cluster increases with cluster size and similarity.

Our dataset consisted of 2043 patients described by 58 molecular features (46 genomic mutations and 12 karyotypic abnormalities). After excluding patients without alterations, we performed 5,000 burn-in iterations followed by 10,000 sampling iterations for the HDP model. The model identified five clusters in the real data and six in the synthetic data. Notably, the clusters extracted from synthetic data mirrored those from the real dataset in terms of dominant genomic features. The emergence of a sixth cluster in the synthetic set (driven by BCOR, which was part of the fifth cluster in the real data) suggests the generative model may have captured subtle substructures underrepresented in the original data. This finding opens three interpretative scenarios: (i) the model successfully identified a novel but real biological subgroup; (ii) the model amplified noise present in the training data; or (iii) the synthetic data require further tuning to match real-world complexity. Among these, the first hypothesis presents the most exciting implications for the use of synthetic data in rare disease research.

*UMAP and SCID for MDS patients stratification*

To further elucidate the stratification potential of our data, we applied a multi-step pipeline involving preprocessing, dimensionality reduction, and clustering to a refined subset of 1811 MDS patients. The selection excluded individuals with missing cytogenetic data and rare mutations with high missingness (>20%) or low occurrence (<10%). The final dataset included 12 cytogenetic features and 29 genetic mutations, all boolean and complete.

Dimensionality reduction was performed using UMAP, which constructs a k-nearest neighbor graph of patients based on similarity and embeds it into a lower-dimensional space while preserving the local and global structure of the data. Given the binary nature of our features, we employed the Jaccard distance:

$$J(X,Y) = \frac{C_{01} + C_{10}}{C_{00} + C_{01} + C_{10} + C_{11}}$$

where $C_{ij}$ represents the count of mutations for which patient X has value $i$ and patient Y has value $j$.

On the reduced-dimensional embedding, clustering was conducted using a modified Stable Co-clustering by Iterating DBSCAN (SCID) algorithm. Unlike traditional DBSCAN, SCID accommodates clusters of varying densities by iteratively adjusting the ε-parameter (representing maximum neighborhood radius) and excluding previously clustered points in subsequent iterations. Specifically, the set of ε-values was derived from quantiles of all unique pairwise Euclidean distances in the embedding space. The minimum sample size parameter, $min_s = 47$, was selected to preserve smaller, clinically relevant clusters.

This methodology enabled the identification of 13 distinct and reproducible clusters within the patient population, thereby underscoring the utility of synthetic and real data harmonization combined with robust unsupervised learning strategies for advancing MDS subtyping and precision diagnostics.

*Survival Analysis with Cox Model, Linear and Non-Linear approaches*

To evaluate whether the synthetic MDS dataset successfully replicates survival outcomes observed in the real data, we analyzed patient survival in relation to the occurrence of death or progression to AML. Patients were stratified according to WHO classification, and four key analyses were conducted: Kaplan-Meier survival curves were generated separately for each WHO class in the real and synthetic datasets; median survival times were computed per WHO group; a univariate Cox proportional hazards model was employed to test whether the data source (real or synthetic) was associated with differential survival; and, finally, mixed-effects Cox models with ridge penalties were fitted, grouping covariates into Clinical, CNA, Demographic, and Genetic categories in order to assess the relative importance of each group.

The Kaplan-Meier survival curves demonstrate that the synthetic dataset reproduces the principal survival trends evident in the real cohort. In both datasets, patients classified as MDS-EB2 exhibit the poorest survival, followed by MDS-EB1, with all other classes showing more favorable outcomes. Some divergence between real and synthetic data becomes apparent in the tails of the survival curves, particularly at longer survival durations. This discrepancy is likely due to the scarcity of long-surviving patients in the real dataset, which limits the generative model's ability to faithfully replicate those patterns. A more informative comparison might be obtained by restricting the analysis to patients with survival times under 100 months.

Median survival times are largely comparable between the two datasets across WHO classes. The real dataset includes 2,043 patients, of whom 703 experienced an event, while the synthetic dataset contains the same number of patients, with 748 events. While there are slight differences in specific subgroups (for instance, MDS-EB1 patients show a median survival of 46.1 months in the real data compared to 39.7 months in the synthetic) overall estimates are consistent. In some classes, such as MDS-MLD, median survival could not be estimated due to survival probabilities not dropping below 50% within the observation window. A univariate Cox model using a binary covariate to indicate dataset origin (real vs. synthetic) yields a non-significant p-value of 0.742, suggesting no substantial survival difference between the two datasets. This lack of statistical significance persists within each WHO class when considered separately, with all p-values exceeding a 0.01 threshold.

Further assessment was performed using mixed-effects Cox models incorporating ridge penalties, with covariates grouped into Clinical, Demographic, and Genetic categories. Partial concordance indices were similar between the real and synthetic datasets across most variable groups. For example, concordance for clinical variables was 0.683 in the real dataset and 0.708 in the synthetic, while demographic variables yielded concordance values of 0.581 and 0.601, respectively. The main difference emerged in the genetic category, where concordance was 0.654 in the real data and 0.722 in the synthetic. Global concordance was also comparable, measured at 0.736 (standard error 0.012) in the real dataset and 0.796 (standard error 0.012) in the synthetic. Overall, the patterns of variable importance were consistent across the two datasets, with one notable exception: the Genetic group exhibited a larger contribution in the synthetic data. This finding likely reflects the tendency of the generative model to introduce rare mutations more frequently, thereby reducing the sparsity that characterizes real-world genomic data.
In conclusion, the synthetic dataset replicates the survival trends, class-based differences, and variable importance profiles observed in the real data. While the synthetic model introduces slightly more genetic variability, particularly by mitigating sparsity in rare mutations, these differences do not compromise the overall consistency in survival modeling between the two datasets.

Survival analysis is a fundamental statistical approach widely used in clinical research to predict the time until a particular event occurs, such as death, disease onset, or progression. However, datasets emerging from clinical trials and genomic screenings, especially those related to hematological disorders, pose substantial challenges due to their high dimensionality, heterogeneity, censoring, and missing data. These characteristics often limit the efficacy of traditional statistical methods. In particular, censored data, where the outcome is not observed within the study period, requires specialized modeling approaches.

The Cox proportional hazards (CPH) model has long been the standard method for survival analysis. It is a semi-parametric model that evaluates the impact of various covariates on the hazard rate, assuming a linear relationship and proportionality of hazards over time. While it is computationally efficient and interpretable, its assumptions do not hold well in high-dimensional or complex datasets, and it struggles to model non-linear interactions or correlated features.

Recent advances in ML have led to the development of more flexible methods capable of handling these complexities. These approaches can model non-linear relationships and interactions without relying on restrictive assumptions, and they scale better to large, multidimensional datasets. Studies have shown that ML-based survival models often perform as well as or better than the Cox model, especially when integrating multiple data types, such as clinical, genomic, and cytogenetic variables.

To investigate this further, a systematic comparison was carried out between traditional statistical models and ML-based methods on a large, high-dimensional dataset from the GenoMed4All consortium. The dataset comprises anonymized clinical and genomic data for 2,043 patients diagnosed with MDS, a group of clonal hematopoietic disorders prone to progress into acute myeloid leukemia (AML). The dataset includes demographic information, hematological and bone marrow test results, mutational status across 47 genes, chromosomal alterations, and risk scores like IPSSR. Outcomes considered were OS and event-free survival

(EFS), with preprocessing steps involving data homogenization, exclusion of patients with missing or zero time-to-event data, and imputation of missing values.

The survival methods compared include both traditional and modern approaches. The classical Cox model was used as a baseline, acknowledging its limitations in high-dimensional settings. To overcome these, penalized variants like elastic net regression, combining L1 (LASSO) and L2 (ridge) penalties, were evaluated for their ability to handle feature selection and multicollinearity in complex datasets.

Among ML methods, random survival forests (RSFs) extended Breiman's random forest technique to censored data, enabling the modeling of non-linearities and variable interactions. These models use log-rank statistics to determine optimal data splits, offering robustness in heterogeneous settings. Similarly, gradient boosting machines, particularly the XGBoost implementation, were adapted to survival analysis, optimizing a differentiable loss function using an ensemble of decision trees.

In conclusion, the study highlights how ML-based methods provide a powerful alternative to traditional models in survival analysis, particularly when dealing with large-scale, heterogeneous, and censored clinical-genomic data. These findings underscore the growing relevance of non-linear models in precision medicine, offering enhanced prognostic accuracy for complex diseases such as MDS.

The evaluation of various survival analysis focused on two clinical outcomes: OS and EFS. The study explores multiple modeling strategies (including Elastic Net, Random Survival Forests (RSF), Gradient Boosting (GB), XGBoost, and DeepCox) across different input configurations: a complete feature set, IPSSR-based summarization, HDBSCAN clustering, and a baseline using only clinical and demographic features. For each method and event type parameter search spaces were defined. A robust evaluation was conducted through a 10-times repeated 3-fold cross-validation strategy, and the resulting concordance indices were calculated for both OS and EFS. Statistical significance between models and configurations was assessed using the Wilcoxon Rank Sum test.

The findings highlight a consistent pattern: using only demographic and clinical variables yields the poorest performance across all models. This underscores the importance of including genomic data to enhance predictive accuracy. No single model consistently outperformed the others; rather, each model exhibited strengths depending on how the input data were structured. Notably, the Cox model performed best when genetic data were pre-processed through clustering, which effectively codified key patterns. Conversely, more complex, non-linear models like RSF, GB, and XGBoost showed improved performance when raw, unprocessed genomic data were included, leveraging their ability to capture intricate relationships.

The "IPSSR" configuration, which aggregates cytogenetic information, proved especially effective in regularized models like Elastic Net and remained competitive in non-linear models. This could be due to the stronger internal correlation structure of cytogenetic features compared to the more heterogeneous mutational data, which helps mitigate issues like multicollinearity.

The study concludes that when constructing predictive models for clinical applications involving heterogeneous data types, trade-offs exist between performance and interpretability. While Cox regression remains popular among clinicians due to its simplicity and transparency, its performance relies heavily on informed pre-processing such as clustering. In contrast, non-linear models offer higher flexibility and generally better out-of-the-box performance, though at the cost of reduced explainability.

Future work will focus on enhancing the interpretability of these complex models. Specifically, the implementation of SHAP (SHapley Additive exPlanations) and LIME (Local Interpretable Model-agnostic Explanations) are currently being explored. SHAP uses game theory to assess the contribution of each input feature by evaluating all possible input combinations, ensuring consistency and local accuracy. LIME, on the other hand, builds interpretable local models around individual predictions to explain the decision logic of black-box models. These tools promise to bridge the gap between model performance and clinical usability by making machine learning-based survival predictions more transparent and trustworthy.

*Preliminary Results on Federated Learning*

In this preliminary study, we tested our Federated Deep Embedded Clustering (DEC) model on the EUROMDS dataset to evaluate its performance across multiple dimensions, including training quality, clustering fidelity, and generative capabilities. The DEC model combines a tied stacked autoencoder (TSAE) for feature space initialization and a federated k-means clustering stage, with both trained in a federated setting using variants of the FedAvg algorithm, namely FedAvg, FedAdam, and FedYog, depending on the local optimizer.

Due to the absence of ground truth labels in the EUROMDS dataset, clustering assignments from the HDP method were treated as a reference for performance evaluation. The training pipeline required specific adjustments to the federated setting, such as a non-trivial approach to initializing centroids. To address inconsistencies across clients, multiple centroid aggregation strategies were tested, with the most promising involving iterative central server aggregation based on similarity until a fixed number of centroids was reached. The missing values in the data were imputed using the empirical probability of observing a 1 in a given column, computed from patients with valid entries. The model optimization used two main loss functions: reconstruction loss (mean squared error between original and reconstructed data) and the Kullback-Leibler divergence (KLD) between predicted and target cluster distributions. To assess clustering quality, the Silhouette Coefficient (Sil) was adopted using cosine similarity, and the Calinski-Harabasz score (CH) as our evaluation metrics. Additionally, to test the DEC model's generative capabilities, we measured reconstruction loss and a novel metric called "cycle accuracy," which captures the consistency between cluster labels assigned to real and reconstructed data. This allows us to gauge how much the model preserves label-relevant features when reconstructing data.

Consistency of training under various federated settings was then investigated. Using the HDP-based labels as a benchmark, we distributed the 2043 EUROMDS patients across multiple clients under both uniform and skewed distributions (the latter using a skewed Gaussian). We tested a range of configurations by varying the optimizer, centroid aggregation strategy, and client data distribution. Results showed that the federated DEC model, even under heterogeneous data distributions, could replicate clustering structures consistent with HDP assignments.

The evaluation metrics from various model configurations highlighted the robustness and versatility of the federated DEC framework. Notably, models using FedAdam and FedYogi often outperformed those using FedAvg, especially in settings with skewed data distributions. Furthermore, centroid initialization strategies significantly impacted model performance, with "max-min" and "double k-means" approaches providing more stable results across settings.

Overall, these preliminary results validate the feasibility and reliability of our federated DEC model in producing high-quality, interpretable clusters from distributed medical data without requiring centralization, while also demonstrating its potential to generate consistent and useful synthetic data.

*Survival curves predictions*

In this study, survival curve predictions were used to evaluate and compare the outcomes of patients clustered using the HDP method and a new configuration developed by the project team. Survival was assessed through three key metrics: LFS, OS, and AML-adjusted overall survival. Each of these metrics considered whether an event occurred and the time to event, allowing for the application of the Kaplan–Meier estimator. The survival curves were generated for both the new clustering configuration and the HDP-based labels, providing a visual and quantitative comparison. Notably, the results highlighted that the survival curves corresponding to the new model's labels exhibited clear separation between classes, indicating a strong capacity to distinguish prognostic groups based solely on mutational profiles, even on data unseen during training.

*Synthetic Data Generation*

In addition to survival analysis, the project demonstrated the generation of synthetic patient data through a decoder operating in the final feature space. Unlike binary data, these synthetic profiles are probabilistic due to the imputation scheme used. Each synthetic sample represents a probabilistic reconstruction from well-characterized cluster distributions, enabling virtual patient generation for data augmentation and further analysis.

*Survival Analysis with Variational Autoencoders*

A broader goal of this work was the development of a unified architecture capable of addressing multiple challenges in hematological malignancy studies, such as prediction, stratification, clustering, missing data imputation, and synthetic data generation. This architecture is based on VAEs, particularly through an approach termed SAVAE. This model leverages a shared latent space to jointly model all covariates, eliminating the need for linear assumptions and enhancing performance over classical models. Early results, obtained under a centralized scheme using the MDS database, underscore the potential of this deep learning method to handle complex survival prediction tasks effectively. The long-term objective remains to transition this architecture into a FL framework, enabling collaborative model training across multiple data nodes while preserving data privacy.

In the Bayesian VAE, a latent variable is sampled from a prior distribution and then used to generate data via a conditional distribution. This process is framed as an optimization problem to approximate the true posterior using a variational distribution. The implementation employs Deep Neural Networks (DNNs), leveraging the reparameterization trick to calculate gradients of the ELBO and enabling analytical computation of the KL divergence. SAVAE extends this framework to the survival analysis context. It models the conditional distribution of the time-to-event variable, assumed to follow a Weibull distribution, alongside observed covariates. The model assumes conditional independence between the covariates and the event time, and estimates the true posterior with a single variational distribution. The latent variable, derived via reparameterization, captures the underlying structure required for predicting survival outcomes. A crucial feature of the SAVAE architecture is its capacity for data imputation. Given the high rate of missing data in real-world datasets like MDS, the study explores traditional imputation techniques (zero imputation, statistical imputation, and MICE), as well as VAE-based imputation, which leverages the model's latent space to infer missing values. This approach has shown strong performance in managing high-dimensional, heterogeneous data with missing features.

Model performance was assessed by comparing SAVAE against two standard models in survival analysis: CPH and DeepSurv. The results, evaluated using the CI, demonstrate that SAVAE consistently outperforms both baseline models across different survival outcomes (leukemia-free survival, overall survival, and AML-adjusted survival) and imputation methods. Notably, SAVAE shows robustness to missing data, performing comparably regardless of the imputation strategy used.

Finally, the document presents an FL approach to enable distributed training across multiple clinical data nodes while maintaining data privacy. Three scenarios are evaluated: (A) independent local training without data sharing, (B) sequential training using shared samples from the most data-rich node, and (C) a federated averaging scheme where model weights are periodically exchanged and aggregated centrally. The federated implementations aim to approximate centralized performance while accommodating variations in data quality and quantity across nodes. These setups lay the groundwork for privacy-preserving, scalable training in real-world clinical environments.

In this preliminary phase, experiments were conducted using the MDS dataset divided into only two nodes to simplify analysis and interpretation. Given that imputation methods showed minimal variation in centralized experiments with this dataset, a statistical imputation technique was adopted for the FL evaluations. The selected time-event pair for analysis was overall survival, commonly used in comparative studies of this kind.

To assess the impact of data imbalance between nodes in both quantity and quality, four configurations were examined. Each setup involved one node receiving 80% of the data and the other 20%. The baseline configuration maintained full data in both nodes, while the other three introduced data degradation in the smaller node: either by randomly removing 50% of each feature's values, or by removing features based on their importance as determined by Shapley Additive Global Importance (SAGE), a model interpretation tool that quantifies the overall contribution of each feature across all samples. Results demonstrated that FL, particularly in Scenarios B and C (where some level of data sharing occurs), consistently improved the performance of the less resourced node. This improvement was especially significant compared to Scenario A, where no data sharing occurred. Notably, configurations in which the most important features were removed performed worse than those where only less important features were excluded, reinforcing the relevance of SAGE-based feature assessment. These findings underscore the necessity of implementing collaborative strategies in FL to mitigate disparities in data distribution and quality.

*Imputation using Deep Generative Models*

Deep generative imputation models were developed and evaluated for addressing the widespread issue of missing values in large-scale datasets, particularly within the MDS dataset. Initial analysis revealed that missing data is prevalent across all variable types, cytogenetic (11%), clinical-biological (93%), and genomic (75%), highlighting the necessity for robust imputation strategies.

The proposed solution is a deep learning-based generative model that estimates missing values by learning from the observed data. Formally, a data sample $x = (x_1, x_2, ..., x_n)$ is modeled using a latent representation $z$, and the model is trained to optimize the conditional distribution $P(x|z, \theta)$, with $P(z|\phi)$ as a prior over the latent space. The joint model is defined as:

$$P(x, z, \phi, \theta) = P(x|z, \theta) \cdot P(z|\phi) \cdot P(\theta) \cdot P(\phi)$$

To accommodate the heterogeneous variable types in the MDS dataset, a composite loss function was employed. It integrates binary cross-entropy for cytogenetic data (modeled as Bernoulli), scaled Poisson negative log-likelihood for clinical-biological variables, and a Gaussian mixture model for the latent space distribution.

Imputation is performed by fixing the trained decoder and estimating the latent representation that maximizes the likelihood of the observed data $x_o$, then generating the missing values $x_m$ via:

$$P(x_m|z, \phi, x_o) = P(x_m|z) \cdot P(z|\phi, x_o) \cdot P(\phi)$$

The model architecture involves a two-layer decoder that transforms a 2-dimensional latent space into a 65-dimensional output, and it was trained over 200 epochs. For evaluation, 10% of the dataset was artificially masked, and the imputation accuracy was assessed. Results showed high accuracy for neutrophils, platelets, and monocytes, with predicted values falling within normal physiological ranges. However, the model underperformed on haemoglobin levels, where predictions often deviated from healthy standards.

In conclusion, the deep generative decoder demonstrated effective imputation for several clinical variables, but further refinement is required to improve performance across all data types.

*XGBOOST*

A federated XGBOOST model was also applied and evaluated on the MDS dataset, focusing on a classification task based on the target variable "Dead/Alive adjusted censored at transplantation," relevant for survival analysis. The baseline comparisons use the original XGBoost implementation, known for its strong performance in classification and regression on tabular data due to features like pruning, automatic feature selection, and distributed execution capabilities.

Data preprocessing was essential due to constraints in the federated implementation, which only accepts 2D integer arrays and does not support missing values. Continuous features were scaled (e.g., multiplied by 10 or 1000) and converted to integers. Missing data were imputed using the mean strategy, and features with over 90% missing values were discarded, resulting in 113 features. To address class imbalance, the SMOTE technique was applied in some configurations.

Six algorithmic configurations were tested: the original XGBoost on raw, imputed, and imputed+balanced data, and the federated XGBoost with 2 and 5 clients on imputed and imputed+balanced data. Each configuration was executed over 10 runs using a 70%,/30% train/test split, and performance was evaluated by accuracy, sensitivity, and specificity.

Results show that applying SMOTE significantly increases sensitivity but drastically reduces specificity, indicating a shift in bias toward the positive class (“Alive”). For example, in the configuration "XGBoost Yes Yes", sensitivity reached 0.93, but specificity dropped to 0.22. Federated configurations demonstrated similar performance to the original XGBoost under identical preprocessing conditions, suggesting that federated distribution does not degrade classification quality. For instance, "FED 2 Clients Yes No" achieved an accuracy of 0.74 with a sensitivity of 0.39 and specificity of 0.88, comparable to "XGBoost Yes No".

In conclusion, the federated XGBoost implementation retains comparable performance to its centralized counterpart under equivalent data preparation strategies. The number of federated clients had limited influence in this phase, though future exploration is required, particularly in scenarios with more clients and smaller local datasets.

**2. Use case Multiple Myeloma**

MM presents as a genetically complex and dynamically evolving disease, where tumor heterogeneity within the same patient complicates prognosis and therapeutic strategies. Although modern therapies have extended survival, relapse remains common, and early identification of patients likely to experience rapid progression remains an unmet clinical need. This study aims to address this gap by applying ML techniques to integrate baseline clinical, genomic, and imaging data with treatment response indicators, specifically minimal residual disease (MRD), to accurately identify high-risk patients early, particularly those relapsing within one year of initiating treatment.

A cohort of 300 newly diagnosed MM patients will be enrolled, with comprehensive data collected within four months of diagnosis, including immunophenotypic, genomic, and imaging assessments. MRD will be evaluated both by next-generation sequencing (NGS) and PET/CT imaging at four months to capture the dynamic nature of disease response.

Preliminary analysis on a subset of 80 patients examined copy number variations (CNVs) at diagnosis and relapse, using 2409 CNV measurements per patient. Data dimensionality was reduced via UMAP and clustered using DBSCAN, resulting in seven clusters at diagnosis and four at relapse. Patients showing similar CNV profiles at both stages were grouped into 14 evolutionary trajectories, offering insights into disease progression patterns.

A separate cohort of 88 patients underwent 18F-FDG PET/CT imaging and molecular profiling via Ultra Low Pass Whole Genome Sequencing (ULP-WGS) of both bone marrow-derived gDNA and peripheral blood cfDNA. The cfDNA tumor fraction (TF) at diagnosis was significantly lower than gDNA TF (median 4.4% vs. 59.7%), but strongly correlated with bone marrow tumor burden ($r=0.47$, $p<0.0001$). High cfDNA TF levels were associated with extramedullary disease, increased number of focal lesions, and higher metabolic activity on imaging, all markers of aggressive disease. Notably, high cfDNA TF predicted poorer progression-free and overall survival, and revealed spatial genomic heterogeneity in 87.5% of patients with divergent cfDNA and gDNA profiles.

Concordance between cfDNA TF and PET/CT findings post-induction therapy suggests cfDNA's potential as a non-invasive biomarker for disease monitoring. Collectively, these findings underscore cfDNA's prognostic value and its utility in capturing the spatial and temporal heterogeneity of MM, which may enhance personalized treatment strategies in future clinical practice.

### Supplementary Information 26| Deliverable 6.6 - Multiple Myeloma Features Extraction

#### 1. Multiple Myeloma: Use Case Introduction

MM is a relatively rare cancer but ranks as the second most common blood malignancy after leukemia. It arises from abnormal monoclonal plasma cells that accumulate in the bone marrow, impairing normal blood cell production. These plasma cells, responsible for producing antibodies, contribute to an immunosuppressed microenvironment and disrupt bone metabolism, leading to osteolytic lesions. Common symptoms include bone pain, fatigue, and increased risk of infection, and the disease frequently affects older adults, particularly men. Though currently incurable, multiple treatment options exist to manage the condition and enhance quality of life.

Whole-body low-dose CT (WBLDCT) is the gold standard for identifying bone lesions, staging the disease, and monitoring treatment response in MM. However, the complex structure of bones makes manual segmentation time-consuming and error-prone, prompting interest in automated, deep learning–based solutions. While recent efforts using U-Net variants and detection models such as Faster R-CNN and YOLO show promise, their generalizability remains limited due to small, proprietary datasets and a lack of focus on specific bones.

To address these challenges, a two-phase pipeline was developed in this work. First, the femur bone is detected using Faster R-CNN variants. Then, bone marrow within the femur is segmented using U-Net-based models enhanced with attention mechanisms. Complementing this segmentation task, radiomics and radiocytogenetics models were constructed to evaluate patient survival and relapse. These models incorporate both early and late fusion strategies for integrating multi-omics data, offering a comprehensive framework for clinical assessment and personalized management of MM.

#### 2. Imaging Data

This study analyzed a dataset of 200 consecutive patients newly diagnosed with MM, treated at Alexandra General Hospital, National and Kapodistrian University of Athens. Each patient underwent whole-body low-dose CT scans prior to therapy initiation, with imaging reviewed independently by two radiologists specializing in myeloma-related bone disease. Pixel-based bone marrow annotations were provided by experienced radiologists. The imaging data, acquired with varying slice spacing and consistent 512×512 pixel resolution, included between 500 and 700 slices per patient. Interpolation was applied before radiomics analysis to ensure consistent texture scaling. The cohort had a mean age of 66.45 years (±11), comprising 82 females and 118 males. This dataset supports outcome prediction, including therapy response and survival, through the integration of multi-omics data and machine learning. Clinical data encompassed demographic variables, treatment details, international staging system (ISS) classification, and survival outcomes such as progression-free and OS. Laboratory parameters included hemoglobin, creatinine, albumin, monoclonal protein levels, bone marrow infiltration, and other serum and urine tests. A total of 32 clinical and laboratory features were evaluated.

Cytogenetic analysis via FISH provided binary indicators for key genetic abnormalities, including del(13q), t(4;14), del(17p), amp(1q21), t(14;16), and t(11;14). These data informed calculations of R-ISS, R2-ISS, and cytogenetic risk classification. Due to limited availability, R2-ISS was excluded from early fusion modeling, and cytogenetic data were represented as a two-value vector comprising R-ISS and cytogenetic status.

#### 3. Methods for Deep Learning Based Segmentation

The deep learning-based segmentation approach for this study involved a two-step pipeline: initial bone detection followed by bone marrow segmentation. For the detection phase, the Faster R-CNN architecture was adapted, which integrates a Region Proposal Network (RPN) and a Region-based Convolutional Neural Network (RCNN) to generate and refine bounding box proposals. Non-Maximum Suppression was used to eliminate redundant predictions, and to improve computational efficiency, the default VGG-16 backbone was replaced with MobileNetV3 pre-trained on ImageNet.

For segmentation, several custom architectures inspired by U-Net were developed. These models incorporated residual and attention blocks to enhance feature extraction and spatial understanding, essential for precise semantic segmentation. The architecture adhered to the encoder-decoder paradigm, allowing for the effective down-sampling of input data and reconstruction of high-resolution output, particularly beneficial for tasks requiring fine-grained detail.

The models were trained on pixel-based annotations converted into COCO-format bounding boxes. CT images were pre-processed using gray-level mapping and normalized to a 0–1 floating-point range. To improve generalization and reduce overfitting, extensive on-the-fly data augmentation, including random rotations, flips, and deformations, was applied using the Albumentations framework. The segmentation model operated on 80×80 pixel input patches, using a learning rate of 0.001, binary cross-entropy as the loss function, the ADAM optimizer, and up to 100 training epochs. Early stopping and learning rate reduction strategies were employed to optimize model convergence.

**4. Methods for the proposed ML analysis**

The proposed machine learning analysis involved a comprehensive multi-step approach integrating radiomics, radiocytogenetics, data stratification, and oversampling strategies. Radiomics analysis extracted 2,996 features from CT scans using PyRadiomics, including first-order statistics, shape descriptors, and texture-based features derived from various matrices such as GLCM, GLRLM, and NGTDM. To enhance feature richness, images underwent isotropic resampling and were filtered using multiple techniques including wavelet transforms with various mother wavelets. All features were standardized prior to analysis.

Radiocytogenetic analysis aimed to enhance predictive accuracy in relapse and survival assessments by combining multi-omic data. Two integration strategies were applied: early fusion, where preprocessed data from different sources were concatenated into a unified representation; and late fusion, where separate models were trained for each modality and then combined using a novel heterogeneous multi-modal ensemble that leverages both hard and soft voting. This ensemble uniquely accommodates diverse input types such as radiomics, clinical, and cytogenetic data.

Patient-level four-fold cross-validation was used for model training and evaluation, ensuring class balance and avoiding data leakage by reserving the test set exclusively for final performance assessment. Each pipeline execution was repeated 100 times on varying data splits, with results reported as mean ± standard deviation to ensure robustness. To address class imbalance, particularly relevant given the skewed relapse probabilities in the cohort (79% low, 21% high), the synthetic minority oversampling technique (SMOTE) was applied. SMOTE generated new synthetic instances near the minority class decision boundary using k-nearest neighbors and was applied separately to training and testing sets to preserve data integrity.

To identify the most informative features for predicting clinical outcomes, a combination of univariate and multivariate selection methods was applied. Zero-variance filtering was first used to eliminate non-informative deep features and radiomics. ANOVA was then employed separately on clinical and imaging features to assess their individual significance, followed by L1-penalized linear regression to further reduce feature dimensionality and create a more compact and efficient representation.

For outcome prediction, various machine learning classifiers were tested for relapse assessment, including k-nearest neighbors, decision trees, Gaussian process classifiers with radial basis function kernels, and support vector machines using multiple kernel types (radial basis, linear, polynomial, and sigmoid). Survival outcomes were modeled using classifiers such as random forest, extra trees, survival trees, Cox regression, CPH models, and specialized SVM variants.

Model optimization involved testing different combinations of key hyperparameters, such as the number of neighbors in k-NN, L1 regularization strength, and SVM C-values. While some parameters like the GPC factor and kernel-specific scales were kept at their default settings due to their high computational cost, twelve parameter sets were systematically evaluated. This ensured that model selection was both methodologically sound and computationally efficient, with consistent use of SMOTE to address class imbalance across configurations.

**5. Results**

The study evaluated a deep learning-based pipeline for detecting, segmenting, and predicting relapse in bone marrow images of MM patients. For bone detection, Faster R-CNN models with different backbones were compared over 100 training epochs with 5-fold cross-validation. The model using a pretrained MobileNet v3 backbone achieved superior performance, with a mean average precision (mAP) of 0.878 and an F2 score of 0.998, and was thus selected for the detection step of the pipeline.

For the segmentation task, model performance was assessed using ground-truth annotations from radiologists. Metrics such as the Dice Similarity Coefficient (DSC), Intersection over Union (IoU), F1 score, and precision were used for evaluation. The residual attention U-Net architecture outperformed standard and attention-based U-Nets, demonstrating the highest accuracy across all metrics, with a DSC of 0.855. Training employed early stopping and typically converged in about 30–40 epochs, with a one-day training time per model. Inference took approximately 20 seconds for detection and 10 seconds for segmentation on an NVIDIA RTX 2080 GPU.

For relapse classification, radiomic features were extracted from bone marrow regions using PyRadiomics. Redundant and low-variance features were filtered using analysis of variance and L1-regularized logistic regression. SMOTE was used to balance classes, and multiple data fusion techniques were explored. Various classifiers were tested, including k-NN, decision trees, SVMs, and Gaussian Process Classifiers (GPC). The GPC with a radial basis function kernel performed best, with an AUC of 0.711 and a sensitivity of 0.634, outperforming all other models.

Finally, a comparison of models based on the full radiomic signature versus a condensed radiomic score revealed that while radiomic scores achieved marginally higher specificity, they consistently underperformed in sensitivity, indicating that the full feature set offers better predictive power for relapse within 12 months.

The clinical model based on a Gaussian process classifier with a radial basis function kernel demonstrated the highest performance among single-source models, achieving an AUC of $0.732 \pm 0.03$ on an unseen testing set. This model also offered improved sensitivity compared to others. Key clinical predictors included age at diagnosis, bone marrow infiltration, and various serum markers such as $\beta 2$-microglobulin, creatinine, IgG, urea, albumin, and monoclonal peak levels. In the multi-omics analysis, early fusion approaches combining radiomics and clinical data with cytogenetic features, including R-ISS, were tested. While adding R-ISS improved specificity, it did not yield statistically significant gains in overall performance, with the best early fusion model reaching an AUC of 0.729.

In survival analysis, early fusion models achieved a CI of 0.76, outperforming both radiomics-only (0.70) and clinical-only (0.72) models. To overcome the limitation of requiring complete datasets in early fusion, a late fusion (meta-model) strategy was employed. This approach combined independently trained models on heterogeneous data sources through ensemble methods, allowing broader patient inclusion. The best-performing late fusion model, integrating radiomics, clinical, and cytogenetic data, achieved the highest AUC of 0.754 and notable specificity of 0.837.

In terms of survival prediction, the radiocytogenetics model not only achieved the highest CI (0.76), but also qualitatively aligned better with actual progression-free survival outcomes compared to the clinical and radiomics models. Although the clinical model had a slightly higher concordance than radiomics, it yielded more false positives in survival curve predictions. These findings underscore the value of multi-modal integration and decision-level fusion for improving both predictive accuracy and interpretability in MM prognosis.

The analysis evaluated how different first-line treatment regimens affect progression-free survival (PFS) in patients with newly diagnosed MM. Treatments were classified as proteasome inhibitor (PI)-based, immunomodulatory drug (IMiD)-based, combinations of both (PI and IMiD), and anti-CD38-based therapies (daratumumab or isatuximab). Results suggest that combining PI and IMiD therapies leads to improved PFS, with further benefits observed when anti-CD38 monoclonal antibodies are included. These findings are consistent with clinical trial data. However, the study's statistical power was limited due to small subgroup sizes, which ranged from 11 to 97 patients. Notably, the daratumumab group included only 11 patients, with just one early relapse, leading to wide confidence intervals and limited significance. Kaplan-Meier curves and statistical analyses (Tables 11 and 12, Figures 11 and 12) illustrate these trends, but no comparisons reached statistical significance, underscoring the need for larger studies to confirm these observations.

### 6. Discussion

This study presents an integrated deep learning and multi-omics pipeline for the segmentation, analysis, and prediction of outcomes in MM patients. Accurate segmentation of bone marrow is essential for assessing myeloma bone disease, a key factor in personalizing treatment. Traditional methods often struggle with artifacts and anatomical variability; hence, deep learning approaches, particularly a Faster R-CNN for femoral shaft detection and a residual attention U-Net for segmentation, were employed to improve precision and efficiency, achieving a DSC of 0.855 on test data.

Radiomic analysis complemented this pipeline by quantifying lesion characteristics, focusing on shape and textural features associated with early relapse. Statistically significant features such as elongation, maximal correlation coefficient, and contrast provided insights into lesion complexity and tissue heterogeneity. Clinically, machine learning identified critical prognostic indicators including age, marrow infiltration, beta-2 microglobulin, creatinine, urea, albumin, IgG, and monoclonal peak levels, all of which are pivotal in assessing disease burden, organ involvement, and treatment response.

The study further explored radiocytogenetic modeling through early and late data fusion strategies, integrating radiomic and cytogenetic information. A late-fusion meta-model using transfer learning outperformed single-source approaches, achieving an AUC of 0.754. This demonstrates the value of multi-omics integration in predicting patient survival and guiding therapeutic strategies. The meta-model also generated survival curves closely aligned with real outcomes, reinforcing its clinical utility in managing care pathways and informing end-of-life decisions.

Despite these advances, limitations remain. The segmentation model focused exclusively on femoral CT scans, limiting generalizability to other common myeloma sites like the spine. However, this narrow focus also provides a solid foundation for future expansion. Radiomic feature extraction across the entire bone marrow rather than lesion-specific regions may have diluted model precision. Furthermore, imbalances in class distribution and missing cytogenetic data constrained the robustness of the early fusion models, especially under k-fold validation.

Nonetheless, the integration of advanced imaging analysis, clinical data, and genomics sets a promising precedent for precision medicine in MM, underscoring the importance of comprehensive, data-driven approaches in disease monitoring and personalized care.

**7. Conclusion**

The integration of radiocytogenetics in assessing recurrence and survival in MM has shown superior performance compared to models based solely on radiomics or laboratory data. Advances in diagnostic markers have significantly improved disease characterization, enabling more accurate prognostic assessments and informed therapeutic decisions. By combining radiological imaging with cytogenetic information, this multifaceted approach provides a deeper understanding of the disease's underlying biology. Consequently, radiocytogenetics holds considerable promise for enhancing patient care and treatment strategies while advancing research in MM. However, further studies and validation are needed to fully establish its clinical value.

### Supplementary Information 27| Deliverable 6.7 - Federated Learning Intermediate Results

**1. Validation Protocol**

This section outlines the validation protocol designed to assess model performance and limitations across two disease-specific use cases: MDS and MM. The approach begins with synthetic patient data generation to simulate real datasets in a computationally efficient yet representative manner. These datasets will include realistic patterns of missing data, introduced via synthetic loss masks, to reflect the challenges encountered in actual clinical environments.

To evaluate the behavior and reliability of these synthetic datasets, their statistical properties and performance will be validated against real data before model development begins. Simulations will involve different node types, each representing institutions with varying capacities, particularly in data quantity and quality, ranging from well-equipped centers to those lacking certain technologies like genomic sequencing.

Three node types will be defined for each use case, striking a balance between realistic deployment and manageable computational cost. Various deployment scenarios will be modeled by adjusting the proportion of each node type, allowing the evaluation of FL strategies under diverse real-world conditions. This setup enables a comprehensive understanding of how federated contributions perform across heterogeneous data sources and institutional capacities.

*MDS Use Case*

In the MDS use case, three types of nodes are defined to reflect varying data completeness and quality. Type 1 nodes contain fully complete data with no missing features, Type 2 nodes simulate datasets with randomly missing values based on real-world patterns, and Type 3 nodes represent partial genomic data loss in 40% of samples, modeled on real institutions such as the Institute for Biomedical Research of Salamanca.

The study evaluates model performance across four scenarios using combinations of these node types, each typically with 2000 samples, except in one scenario where the sample size in Type 3 nodes is varied to test robustness under reduced data availability. The primary analytical task is survival analysis, capitalizing on the availability of survival time and event data within the MDS dataset. Specifically, the analysis will focus on EFS censored at transplantation. To assess model performance and validate the synthetic data, the real MDS database will be used as a benchmark, ensuring that the synthetic data are suitable for accurate and meaningful survival modeling.

*MM Use Case*

In the MM use case, two types of nodes are defined: Type 1 nodes contain complete clinical and genomic information, while Type 2 nodes reflect missing-at-random data based on real-world patterns. Due to the limited dataset of only 100 patients, each scenario involves fewer samples compared to the MDS case. Patients are categorized according to three therapies (Ab-based, IMiD-based, and PI-based) which are evenly distributed across nodes to enable the analysis of therapeutic impact on patient outcomes.

Three scenarios are considered: one with fully complete data, one with data containing missing values, and one mixed scenario with varying sample sizes to simulate data imbalance. The study's main objective is to assess the probability of early relapse, defined as relapse or death within the first 12 months following treatment. This is determined using the "PFS_I_MONTHS" and "PFS_I_EVENT" variables. The analysis is split into two parts: one predicting early relapse based solely on pre-therapy features, and another incorporating post-therapy response data ("INDUCTION_RESPONSE") collected four months after treatment begins. This dual analysis aims to estimate, for each patient and therapy, the likelihood of early relapse both before and after considering treatment response. Given the limited availability of real-world data, validation will rely on synthetic datasets containing 100 samples per therapy.

**2. Synthetic Data Generation**

Numerous approaches have been developed to generate synthetic data, particularly to address issues such as data scarcity or privacy concerns. Deep learning models, including DNNs, transfer learning, and GANs, have been widely used for synthetic data generation in various domains, including high-dimensional image data and categorical data. However, in the medical field, these models are typically designed for specific diseases.

This section presents a methodology for generating and validating synthetic patient data using a model developed by UPM. The approach combines data imputation and missing data simulation through a two-module architecture. The first module imputes missing data and creates a complete synthetic dataset, while the second generates a synthetic missing data mask based on the original data's loss distribution. The final synthetic dataset is produced by applying a Hadamard product between these components.

UPM's model, described previously in D6.4, integrates VAEs with a GMM. The VAE learns the latent space of the original data, and the GMM facilitates sampling new data points by estimating the posterior distribution. This hybrid model supports both continuous and discrete variables, including censored survival times, and is designed to be versatile across domains while maintaining a focus on clinical applications.

The validation process for this synthetic data generation emphasizes both resemblance and utility. Resemblance is assessed by comparing joint distributions using a Random Forest classifier to determine how well synthetic data can be distinguished from real data. Utility is evaluated by assessing whether synthetic data yield comparable statistical conclusions to real data. For instance, in the MDS use case, survival predictions are assessed using the CPH model and the CI, which quantifies the correlation between predicted risk and observed outcomes. For MM data, alternative forms of validation are applied due to the absence of synthetically generated PFS information.

The following sections will detail the validation results for the MDS and MM datasets, using the criteria and methods outlined above.

*MDS Use Case*

The synthetic data generation process for MDS builds on the validation approach previously outlined in D6.4, with updated results due to minor preprocessing and model adjustments. Using a VAE-based generative model, 15 different random seeds were applied, and results from the top 3 were highlighted. An RF classifier achieved accuracy below 70% when distinguishing synthetic from real data, and confidence intervals consistently overlapped, confirming the robustness of the generative process. While an ideal discriminator would reach around 50% accuracy, results in the 65–70% range are deemed acceptable, given the small dataset size and inherent quality issues in the real data. Survival analysis using a CPH model further demonstrated that training on synthetic data yields CI values comparable to those from real data, with synthetic training slightly outperforming real data (0.68 vs. 0.64), albeit not significantly.

*MM Use Case*

In the case of MM, the provided dataset included 99 patients and 19 features, many of which contained missing values. Detailed preprocessing involved the removal of non-informative or sparsely populated features and the numerical encoding of categorical variables. Special attention was given to the "Induction Response" (IR) variable, as it significantly correlates with early relapse (ER), showing a correlation coefficient of -0.54. This relationship serves as a benchmark for validating synthetic data consistency. Dimensionality reduction was guided by clinical expertise to improve model training, and correlations between ER and clinical features were assessed across therapy types. Overall, the IR feature demonstrated the strongest and most reliable negative correlation with ER, while most other features lacked statistically significant associations. These findings validate the model's ability to extract and replicate meaningful clinical patterns in synthetic MM data.

The validation of synthetic data for the MM use case followed the methodology previously applied to MDS, with a focus on ensuring the statistical fidelity of generated data. A RF model was used to assess the distinguishability between real and synthetic data, with the ideal outcome being an accuracy close to 50%, indicating that the model cannot differentiate between the two. Given the limited sample size, the RF achieved an average accuracy of around 60% for covariates alone and 70% when mask variables were included, acceptable given the data constraints, despite broad confidence intervals.

Expert clinical validation was also conducted, emphasizing the comparison of cytogenetic alteration incidences between synthetic data and both a general MM population and the original 100-sample dataset. While the synthetic data broadly mirrored the original dataset, discrepancies were noted, particularly in features with a high number of missing values.

Additionally, the correlation between ER and IR in the synthetic data was examined. The real dataset showed a correlation of -0.54, while the synthetic version yielded -0.47, suggesting that the relationship was effectively preserved. Further analysis across different therapy types revealed that the model captured key correlations and, in some cases, introduced minor spurious associations, particularly in therapy groups with larger sample sizes.

In conclusion, the synthetic MM dataset demonstrates a strong resemblance to the real data in terms of feature distributions, cytogenetic alteration incidence, and inter-variable correlations, supporting its use for downstream FL applications.

### 3. Federated Learning

This section outlines the preliminary goals and technical implementation details of FL algorithms developed within the GENOMED4ALL project. The main objective is to evaluate the performance and deployment readiness of these algorithms in federated environments, with applications across two clinical use cases: MDS and MM. Each partner is expected to mathematically define their algorithm, emphasizing its federated structure, data-sharing procedures, and synchronization frequency. Synthetic datasets provided by UPM will be used for training across all scenarios to ensure comparability, with real data reserved for the MDS use case validation due to limited availability in the GM scenario.

The document identifies two secondary aims: first, to quantify the performance gains of FL over isolated training, particularly in cases of incomplete data, and second, to assess how FL performance evolves with varying sample sizes. These objectives help determine the necessity and benefit of federation, aiming for results closer to a centralized upper-bound performance.

To achieve these goals, the project leverages Flower, a FL framework in Python, for deploying models in a realistic distributed environment. Each partner must integrate their algorithm into the platform, using an object-oriented approach to encapsulate both local learners and aggregation strategies. The local models are implemented in TensorFlow or PyTorch, while the global model uses Flower’s built-in strategies such as

FedAvg. Technical considerations include importing modules within method scopes, managing dependencies, and using mlflow.pyfunc.PythonModel for model registration and instantiation.

The deliverable thus establishes a unified methodological and technical foundation to test, compare, and validate FL contributions across partners, ensuring practical deployment and robust performance evaluation.

**4. UPM Contribution**

The deliverable thus establishes a unified methodological and technical foundation to test, compare, and validate FL contributions across partners, ensuring practical deployment and robust performance evaluation.
The UPM contribution centers on implementing and validating a FL framework for two distinct medical use cases. FL, as described in D6.2, is a decentralized ML approach that enables model training across multiple devices or servers without aggregating raw data, preserving privacy and compliance with data protection standards. Among the various FL strategies, UPM has adopted the FedAvg algorithm due to its simplicity, scalability, and compatibility with large-scale systems. FedAvg works by iteratively distributing a model to local nodes, which train independently and return updated parameters to a central server, where a weighted average based on local data volume is computed to update the global model. This process is repeated to converge on a robust federated model.

For this deliverable, FedAvg has been selected as a baseline to generate initial results, which will later be compared with a novel FL strategy under development by UPM. Specifically, in the MDS survival analysis task and MM prediction task, nodes train the local model for a fixed number of epochs or iterations, share parameters, and receive an aggregated global model to continue training. This exchange is repeated five times, providing sufficient data for performance assessment.

*MDS Use Case*

In the MDS use case, UPM integrates its previously developed survival analysis algorithm into the FL environment. The architecture, known as SAVAE, is based on VAEs, selected for their ability to learn a shared latent representation of distributed data without centralizing it. This design supports complex survival analysis by modeling censored data, covariate interactions, and time-dependent risks. SAVAE approximates the posterior distribution using variational inference and optimizes the ELBO via the reparameterization trick. The model jointly reconstructs covariates and predicts time-to-event from the learned latent space. This federated implementation of SAVAE allows multiple nodes to collaboratively learn a predictive model for survival analysis while maintaining data privacy, laying the foundation for scalable and privacy-preserving medical AI applications.

In the distributed training analysis across four scenarios, consistent trends emerge regarding the performance of the SAVAE model under isolated, federated, and centralized conditions. In Scenario 1, where all three nodes are homogeneous and contain complete data, isolated training yields a CI of approximately 0.70 across all nodes. FL does not offer any performance advantage here, as each node already possesses sufficient and equivalent information. The centralized training case, with a single node housing the combined 6000 samples, also achieves a similar CI, confirming the adequacy of isolated training for this scenario.

Scenario 2 introduces random missing data in otherwise similar nodes. Despite this loss, the CI remains close to 0.70, with negligible differences between isolated and federated results. This indicates that FL offers limited benefit when nodes are equally incomplete and lack additional complementary information. The stability of the model performance is attributed to the dominance of non-genomic features, particularly age, in predicting outcomes, as well as the sparsity and low variability within the genomic data.

Scenario 3 simulates heterogeneity across nodes, including one with significant missing data. Surprisingly, performance remains stable across all nodes, with CI values comparable to Scenarios 1 and 2, regardless of the training mode. Even in the presence of substantial missing genomic data, the model maintains robustness due to the dataset's inherent structure and the predictive strength of certain covariates.

Scenario 4 uniquely examines the effect of varying sample size at one node. Here, FL proves beneficial. While isolated training fails to produce reliable results for Node 3 with fewer than 500 samples, FL significantly enhances model performance by leveraging shared weights from larger nodes. This effect is most prominent when Node 3 contains very limited data (10–100 samples), where isolated training yields poor results but FL restores performance to match that of well-sampled nodes. Notably, Nodes 1 and 2 remain unaffected during this process due to the use of the FedAvg strategy.

Overall, the comparative analysis confirms two major conclusions. First, the model's performance is minimally impacted by random missing data, likely due to data imbalances and the dominance of a few key features. Second, FL is particularly valuable when addressing data scarcity, enabling nodes with few samples to achieve strong predictive performance through collaborative learning.

This section presents the results of FL simulations conducted using the Flower library, which is intended for integration into the final GENOMED4ALL platform. The experimental setup replicates previous configurations, including the use of three client nodes and a server, all employing the FedAvg strategy. The comparison between the original UPM simulation and the Flower-based implementation demonstrates strong consistency across all scenarios. The CI values obtained using Flower closely mirror those from earlier simulations, with only minor discrepancies attributable to variations in random seed usage. These results validate the Flower library as a suitable and reliable framework for the deployment of the final FL model, confirming its alignment with previously observed model behavior.

*MM Use Case*

The MM use case outlines the application and evaluation of a logistic regression (LR) model for ER prediction using a synthetic MM dataset. The LR model predicts binary outcomes (ER=0 or ER=1) based on independent variables, with class balancing techniques applied where needed, particularly for Ab-based therapies where the data was highly imbalanced. Model performance was primarily assessed using F1 scores and confusion matrices, with a focus on minimizing false negatives due to their clinical implications.

In Scenario 1, where all nodes had complete data and an equal distribution of therapy samples, FL generally improved or unified performance across nodes. This was especially evident in the Ab-based therapy, where FL mitigated the effects of class imbalance and led to significant increases in F1 scores. Comparisons with centralized training showed that while federated approaches occasionally outperformed centralized models in terms of loss curves, confidence intervals for F1 scores typically overlapped, indicating statistically similar outcomes.

Scenario 2 introduced missing data to simulate a more realistic environment. Despite the challenge, imputation techniques allowed the model to perform comparably to Scenario 1. FL again helped unify performance across nodes, with a consistent reduction in overfitting following model weight sharing.

Scenario 3 explored data heterogeneity by varying the completeness and quantity of samples across nodes. Node 3, with drastically fewer samples (as low as 10), benefited markedly from FL, particularly for Ab-based therapies, where isolated performance was poor but improved substantially through FL. As the number of samples increased (e.g., to 20), performance stabilized, and even surpassed that of fully complete nodes in some cases, likely due to overfitting in larger datasets. FL consistently aided performance in nodes with limited or incomplete data, reinforcing its value in heterogeneous, real-world conditions.

Overall, the study demonstrated that FL not only preserved model performance across distributed settings but also often improved it, especially under data imbalance, incompleteness, or scarcity. The integration with the

Flower framework and the empirical results across diverse scenarios confirm FL's robustness and practical advantage in predictive modeling for MM therapies.

#### 5. UNIBO Contribution

*MDS Use Case*

This section presents the application of two deep learning models, DeepSurv and AESurv, for survival analysis in the context of MDS, exploring both centralized and FL scenarios.

DeepSurv is a deep feed-forward neural network that estimates the log-risk function of the Cox model, optimized via the negative partial log-likelihood. AESurv, on the other hand, uses a dual-headed architecture combining an autoencoder and a CPH model, with latent representations feeding into the survival analysis. The encoder compresses patient data into a lower-dimensional latent space, which is decoded and simultaneously used for risk prediction.

Both models were implemented in FL settings using standard Python and the Flower framework, applying a FedAvg aggregation strategy. Each node independently trains on local data before synchronizing global weights across the network. The models were evaluated through different scenarios that simulated isolated, federated, and centralized training on synthetic and real-world EUROMDS datasets.

In Scenarios 1 to 3, where nodes share similar distributions or degrees of data completeness, FL offered performance comparable to isolated or centralized models, achieving a CI close to 0.73 with DeepSurv. Scenario 4 introduced data imbalance and missingness at node level, revealing that FL substantially benefits nodes with fewer samples, without degrading performance on more complete nodes. Centralized models slightly outperformed federated counterparts but at the cost of requiring all data in one place.

The AESurv model consistently yielded higher CI values than DeepSurv, often exceeding 0.82 in federated settings. It proved particularly robust to data incompleteness and imputation strategies, suggesting that autoencoder-based architectures can be effectively leveraged in distributed settings for survival analysis. Across all scenarios, federated AESurv models demonstrated generalization ability on real data while showing resilience to data heterogeneity.

Validation using the Flower framework confirmed that the federated implementations maintain performance consistency. Minor discrepancies were noted in Scenario 4 with small sample sizes, but overall, Flower-based results were aligned with custom-distributed implementations.

In conclusion, the federated versions of DeepSurv and AESurv effectively preserve predictive accuracy while offering a privacy-preserving alternative to centralized training, particularly benefiting sites with limited or incomplete data. The AESurv model, in particular, shows strong promise for real-world federated healthcare applications.

*MM Use Case*

This section details the application of a penalized logistic regression model within a FL framework for predicting outcomes in MM treatment scenarios. The logistic regression model, regularized through ridge penalization, was chosen for its interpretability and robustness, especially under conditions of class imbalance. Model performance was evaluated using the F1-score, a suitable metric for imbalanced datasets.

The federated implementation was carried out using the FedAvg algorithm, leveraging the scikit-learn library, with training conducted on a synthetic dataset and validation performed on real-world data. Due to limited availability of labeled data, training data for all therapies were aggregated, while evaluation was carried out per therapy.

Across three evaluated scenarios (centralized, isolated, and federated) the results indicate that FL significantly benefits nodes suffering from data scarcity and imbalance, particularly for therapy 1, which consistently showed lower performance in isolated settings. FL helped mitigate this, most notably in node 3 where sample sizes were as low as 10. Incremental improvements in F1-score were observed across increasing sample sizes (10, 20, and 50), confirming the scalability and value of FL in low-resource environments.

A final comparison between the standard distributed implementation and a version implemented using the Flower framework showed negligible differences in performance, thus validating the robustness and reproducibility of the federated setup. Overall, the findings support FL as a viable and effective strategy for predictive modeling in MM treatment, particularly when patient data is decentralized and imbalanced.

**6. UNITO Contribution**

*MDS Use Case*

The contribution of UNITO to the MDS use case focused on implementing and evaluating CPH survival models within a FL framework. Both the traditional linear CPH model and its non-linear variant, DeepSurv, were explored to assess their effectiveness in distributed scenarios. The linear version was implemented using a feedforward neural network with linear activations and L2 regularization, while DeepSurv utilized a neural network to capture potential non-linear relationships. Model federation was managed via the FedAvg strategy, employing weighted aggregation based on client data size, with additional tests on how varying the number of internal training epochs per client influenced outcomes. Median imputation was used to handle missing data in relevant scenarios.

Performance evaluation compared centralized and isolated training as upper and lower bounds respectively, using average CI scores across five repeated runs. Across all scenarios, federated models consistently matched the centralized performance, showing minimal degradation even with missing data or imbalanced node quality.

In Scenario 1, where nodes were homogeneous and data quality was high, the difference between isolated and federated performance was negligible. Scenario 2, despite introducing missing values, maintained similar results, confirming model robustness. Scenario 3 introduced heterogeneous nodes and more pronounced missing data. Here, node-specific improvements were observed, particularly for those underperforming in isolation, highlighting FL's compensatory potential. Scenario 4 analyzed performance under varying sample sizes for a weak node, confirming that nodes with very limited data benefitted significantly from federation. Importantly, the federated model's performance remained stable and unaffected by the inclusion of this weaker node, regardless of its size.

In conclusion, FL provided consistent benefits, particularly for data-poor nodes, and reliably achieved centralized-level performance. No significant advantage was observed in using non-linear models over linear ones in this context, and increasing internal epochs primarily influenced training convergence rather than final model quality.

*MM Use Case*

In this use case, the objective was to predict ER as a binary outcome in MM patients using a FL approach. To enable clinically relevant performance analysis across each category, patients were categorized into three therapy-based groups: antibody-based, IMiD-based, and PI-based. A single fully connected feed-forward neural network was trained across all patients, rather than developing separate models for each therapy group, under the assumption that cross-group information might enhance predictive power. To avoid bias, the therapy type was excluded as a feature due to its non-random assignment in the trial. The model was trained with hyperparameter tuning, class-weighted binary cross-entropy to manage class imbalance, and median imputation for handling missing values. Training was federated using the Flower framework and the FedAvg strategy, with evaluations based on F1-score, repeated across 10 runs.

Across three simulation scenarios, performance was assessed for centralized, isolated, and federated settings. In Scenario 1, where data was complete and evenly distributed, the federated model approached centralized performance for antibody-based therapy and exceeded or matched isolated nodes for IMiD and PI groups. Scenario 2 introduced uniform missing data; here, the federated model consistently outperformed isolated nodes and closely matched centralized scores, particularly excelling in the PI-based group. Scenario 3 tested robustness under variable node quality and data size. With node 3 containing increasing patient volumes (n = 10, 20, 50), FL continued to show strong performance, especially at smaller scales, although some upper bounds were not reached in larger-scale tests.

Ultimately, the federated approach demonstrated strong potential, often outperforming isolated nodes and closely matching centralized models, especially when node data quality was heterogeneous. While a few underperforming cases were noted (mainly in the PI group for Scenario 1 and antibody-based group for Scenario 3 at n = 50) it remains unclear whether these were model shortcomings or artifacts of synthetic data. More definitive conclusions are expected once real patient data becomes available.

### 7. UCPH Contribution

*MDS Use Case*

In the MDS use case, DGD model was employed across multiple scenarios to evaluate the effectiveness of FL in comparison to centralized and isolated training frameworks. In Scenarios 1 through 3, the DGD consistently learned a meaningful representation of the data by clustering alive patients centrally, indicating higher survival probability, while deceased patients were more dispersed. Importantly, in the federated setting, all nodes were able to learn comparable structures, and the loss curves remained within expected bounds (between the lower bound of centralized training and the upper bound of isolated learning) indicating stable and effective learning. Scenario 3 further demonstrated that missing data did not significantly impair the model's performance, affirming the robustness of the approach. In Scenario 4, FL again outperformed isolated training, which showed signs of overfitting, while both federated and centralized models maintained generalizability.

*MM Use Case*

In the MM use case, the DGD was adapted for classification by aggregating data from three treatment groups and implementing a neural network with two hidden layers. To classify ER, the network's output neuron was transformed into a logistic regression layer, using a sigmoid function to yield probabilistic outputs. Model training was conducted using maximum likelihood estimation and optimized iteratively with the Adam optimizer. To mitigate overfitting (given the limited size of the dataset) weight decay regularization was applied.

Performance metrics, demonstrate F1 scores across various scenarios and therapies, comparing isolated, federated, and centralized learning. The results indicate that FL generally enhances model performance, particularly in nodes and scenarios with limited data. For instance, scenarios with low-data environments show a more pronounced benefit from the federated approach. Despite the typical limitations of generative models and neural networks on small datasets, the DGD framework demonstrated robust performance when applied in a federated context.

### 8. FORTH Contribution

FORTH's contribution to the project centers around the development and implementation of a federated version of the XGBoost algorithm, tailored to the constraints and architecture of FL. XGBoost, a well-known ensemble method based on gradient tree boosting, constructs decision trees sequentially by minimizing a loss function and incorporating regularization to control model complexity. The algorithm employs a greedy approach for identifying optimal data splits, balancing computational efficiency and accuracy.

In the federated context, FORTH introduces a mechanism whereby each client locally computes a limited number of top-ranking splits based on gain values and shares summary coefficients with a central server. The server then aggregates this information to identify and broadcast the globally optimal split, allowing each client to continue building the same decision tree in sync. This approach maintains data privacy while enabling a coherent model to be built collaboratively across decentralized datasets.

The federated training process begins with the exchange of key parameters between the server and clients, such as sample counts, learning rates, and initial class probabilities. These are aggregated, typically via weighted averages, to harmonize the training process. As training progresses, clients iteratively compute and share local residuals, probabilities, and split candidates. The server coordinates these exchanges, ensuring all clients build identically structured trees despite differences in local data distribution.

One of the practical challenges addressed is the synchronization of tree paths across clients who may not have data points aligned with specific branches. The server resolves this by lexicographically ordering paths and selectively instructing clients to continue construction only on valid branches, thereby optimizing communication and computation. Once all branches of a tree are constructed, a normalization step aggregates the residuals from all clients into a unified tree, ensuring consistent predictive behavior across the network.

The federated XGBoost implementation is fully integrated with the Flower FL framework through a specialized strategy called FIN. This design allows complex ML workflows to operate seamlessly in a federated setting by abstracting the roles of server and client into modular components. The communication between clients and server is mediated through asynchronous queues, ensuring thread-safe, scalable, and efficient operation. Through this FIN strategy, any algorithm conforming to the defined client-server communication pattern can be deployed within Flower with minimal overhead.

Ultimately, FORTH's implementation achieves an FL model that mimics the behavior and predictive power of centralized XGBoost, while preserving data privacy and supporting scalable deployment across distributed environments.

*MDS Use Case*

The application of our XGBoost implementation to the MDS use case required modifications to extend support from classification to regression, enabling survival analysis through the CPH model. This adaptation involved adjusting the gain formula and removing log-odds conversions. Data inputs included regression values, event status, and event duration. However, this approach is suboptimal as it does not fully leverage XGBoost's tree-based capabilities, and the current implementation does not handle missing values, which were imputed using K-nearest neighbors. Hyperparameters were optimized through internal validation with synthetic data, and the algorithm was run until convergence based on minimizing RMSE, producing deterministic results without confidence intervals.

Performance was evaluated across four scenarios involving federated and isolated training on multiple nodes. In Scenario 1, without missing data, FL outperformed isolated training, achieving higher concordance indices due to more informed tree splits derived from aggregated data. Unexpectedly, centralized training on combined data performed worse, likely due to sensitivity to missing values in validation data. Scenario 2 introduced missing data imputed via KNN, resulting in FL stagnating earlier and failing to improve beyond initial performance, indicating the negative impact of missing data on tree split quality. Scenario 3 involved nodes with uneven genetic information, where FL showed poorer performance on the node with less data, and early convergence at local minima was observed. Scenario 4 tested varying sample sizes for one node, showing that very small datasets (10-100 samples) led to poor local node performance, but federation improved overall model accuracy by mitigating deficiencies in underperforming nodes; with larger sample sizes (500-1000), performance between federated and isolated models converged.

Overall, FL demonstrated advantages in data integration and model performance when data quality and completeness allowed, though challenges remain in handling missing values and uneven data distributions, with room for future optimization and investigation.

*MM Use Case*

The MM use case involved straightforward classification experiments using the existing algorithm without modifications, with the F1 score as the quality metric. Results across three scenarios demonstrated that FL generally matched or exceeded the average performance of isolated nodes, although it did not significantly improve F1 scores overall. In Scenario 1, where there was no missing data, federated models achieved higher or equal accuracy compared to isolated models, particularly for PI-based therapies, while also enabling classification in nodes that previously failed to distinguish classes, albeit with modest scores. Scenario 2 showed a slight decline in PI-based therapy performance due to missing data, but IMiD and Ab-based therapies remained stable. Scenario 3, which tested varying sample sizes in one node, highlighted the ability of FL to maintain performance even with very limited local data, enabling hospitals with small datasets to classify effectively by leveraging the federation. The key insight is that federation acts as a valuable mechanism to compensate for underperforming or data-scarce nodes, effectively transferring knowledge across nodes and improving their classification reliability, rather than dramatically enhancing overall model accuracy.

**9. Conclusions**

This document represents a significant milestone by presenting preliminary results from FL experiments conducted on two use cases, MDS and MM. The partners successfully implemented model simulations within a FL framework using the Flower library, which serves as the foundation for the WP4 platform that will ultimately deploy the federated training models. The teams have effectively designed, developed, and validated algorithms for key tasks such as clustering, survival analysis, and classification, selecting the most promising algorithms to federate. This approach has yielded improvements in learning performance, particularly in scenarios characterized by limited or low-quality data. The progress achieved thus far is expected to further advance the project by providing innovative, sustainable solutions in collaborative learning and data privacy protection. Looking ahead to the next deliverable in December 2024, there remain opportunities for enhancement and new directions to explore. The partners have extended cooperation within WP6 through a shared methodology and agreed protocols, and have collaborated with clinical WPs to deepen understanding of use cases and results. The forthcoming priority is to establish full integration with WP4 to embed the federated models into the developed platform. Additionally, creating a common software repository will be essential to streamline future deployment and maintenance of the models.

### Supplementary Information 28| Deliverable 7.1 - Use Case Design and Planning

**1. MDS Use Case**

*Rationale and Objectives of the Study*

MDS are a group of myeloid neoplasms predominantly affecting the elderly, characterized by peripheral cytopenias and a risk of progression to AML. The 2016 WHO classification defines various MDS subtypes based on morphological and cytogenetic features. While cytogenetic analysis remains critical for prognosis and treatment decisions, the disease's clinical heterogeneity and the limitations of current prognostic tools, such as the Revised International Prognostic Scoring System (IPSS-R), highlight a pressing need for more precise, individualized approaches. IPSS-R does not fully capture mutational complexity or predict disease progression reliably on an individual level, as many relevant mutations are not considered in current models.

This study aims to address these gaps through four objectives. The first is the prevention of MDS by identifying high-risk individuals through genomic screening. Age-related clonal hematopoiesis, driven by somatic mutations in genes such as DNMT3A, TET2, and ASXL1, is associated with increased risks of hematologic malignancies, cardiovascular disease, and mortality. Understanding the mutational landscape in aging hematopoietic stem cells could enable early identification and monitoring of individuals predisposed to MDS.

The second objective focuses on improving the classification and prognosis of MDS through integrative “omics” approaches, including genomic and transcriptomic analyses. Recent discoveries link specific mutations, such as those affecting RNA splicing and epigenetic regulators, to distinct MDS phenotypes and outcomes. These findings suggest that incorporating mutational profiles into classification systems could refine prognostic models beyond what morphology and cytogenetics alone can provide.

The third objective aims to enhance clinical decision-making in MDS using “omics” data. Allogeneic hematopoietic stem cell transplantation (HSCT) remains the only curative option for MDS but is limited by high rates of relapse and mortality. Emerging evidence indicates that specific genetic mutations, including TP53 and others, influence post-transplant outcomes. Furthermore, mechanisms such as HLA loss and immune evasion contribute to relapse, particularly in haploidentical or unrelated donor settings. Thus, comprehensive molecular profiling may improve patient selection for HSCT and inform strategies to mitigate relapse.

Finally, the study seeks to explore drug repurposing opportunities by leveraging network-based analyses of disease-related gene interactions. Shared molecular pathways across cancer types suggest that existing therapies could be repurposed for specific MDS subtypes based on their genetic and regulatory network profiles. This approach could accelerate the development of targeted therapies tailored to individual mutational landscapes.

Overall, the study emphasizes a paradigm shift from morphology-based assessment to integrated molecular profiling in MDS, with the goal of improving prevention, classification, prognosis, and treatment outcomes.

*Study Design*

The study design incorporates a comprehensive, data-driven approach to address the four major research aims related to MDS, leveraging large patient cohorts and advanced AI methodologies to generate clinically impactful insights.

For the first aim, focused on the prevention of MDS through genomic screening, the study analyzes data from 11,000 healthy elderly individuals over the age of 65, using next-generation sequencing (NGS) to detect mutations in 50 genes associated with clonal hematopoiesis. A diverse set of AI techniques, including Bayesian networks, clustering algorithms, Cox regression models, DL, and penalized regression methods, are applied to identify individuals at heightened risk of developing MDS.

The second aim targets the refinement of disease classification and prognosis. This involves 6,700 MDS patients with targeted mutational data, 1,200 with RNA sequencing, and 900 with whole-exome or genome sequencing. The GENOMED4ALL consortium, building on prior collaborative efforts within the EUROBLOODNET network, has assembled a training dataset of over 2,000 patients to develop predictive algorithms. These tools stratify patients into molecularly defined subgroups based solely on genetic data, facilitating reproducible, clinically meaningful classifications. Survival analyses and classification models, validated on an independent cohort of 1,000 additional patients, employ cross-validation techniques and performance metrics such as the CI to ensure robustness and reliability.

The third aim enhances clinical decision-making in transplant-eligible MDS patients. Data from 1,500 individuals who underwent HSCT are analyzed using a combination of clinical decision support systems, Cox modeling, random survival forests, and DL. Time series analysis and stochastic modeling help predict individual trajectories, particularly with regard to disease relapse, thereby informing post-transplant therapeutic strategies.

The fourth aim explores drug repurposing opportunities through a systems biology approach. Utilizing genomic and transcriptomic data from approximately 2,400 patients with hematologic malignancies, the study maps disease-associated genes onto protein-protein interaction networks. Network diffusion algorithms and pathway resilience models are then used to identify actionable therapeutic targets, supporting the strategic reapplication of existing drugs for genetically defined MDS subsets.

This integrative research framework, combining extensive clinical data with cutting-edge AI, aims to shift the current paradigm in MDS from reactive treatment to proactive, precision medicine, improving early detection, classification, prognosis, treatment response, and therapeutic innovation.

*Clinical Validation and Evaluation*

The clinical evaluation and validation of AI algorithms represent a critical component of the GENOMED4ALL project's success. This process is grounded in an ongoing, collaborative dialogue between clinicians and the AI development team to ensure that the models address diverse clinical objectives using appropriate datasets. Leveraging the substantial body of knowledge accumulated in recent years regarding gene mutations and their impact on survival and disease progression in MDS, clinicians can undertake an initial validation of AI-generated models by comparing algorithmic predictions against well-established clinical and molecular correlations.

Mutational profiles, such as SF3B1 being linked to favorable outcomes and mutations in ASXL1, SRSF2, RUNX1, TP53, and EZH2 correlating with poor prognosis, provide concrete benchmarks for validating AI-generated risk stratifications. Furthermore, clinical heterogeneity within phenotypically similar subgroups (such as MDS with ring sideroblasts) highlights the importance of integrating mutational data to refine prognostic accuracy, with SF3B1 mutations indicating prolonged leukemia-free survival compared to SRSF2-driven cases.

Importantly, mutation screening is not only prognostic but also predictive, influencing treatment decisions. For instance, TP53 mutations are associated with poor responses to lenalidomide in 5q- MDS and elevated relapse risk post-transplantation. Similarly, SF3B1 mutations have been linked to improved erythroid responses to luspatercept, and TET2 mutations may predict sensitivity to hypomethylating agents. These insights provide a valuable clinical framework to validate and interpret AI model outputs, ensuring that the integration of algorithmic tools into practice is both biologically sound and clinically actionable.

**2. SCD Use Case**

*Rationale and Objectives of the Study*

SCD is the most prevalent rare hereditary hematological disorder worldwide, caused by a single but universally shared mutation leading to the production of abnormal hemoglobin S. This results in the polymerization of deoxygenated hemoglobin and the sickling of red blood cells, contributing to a spectrum of life-threatening complications. Despite a uniform genetic origin, SCD manifests with substantial clinical heterogeneity, presenting an array of organ complications such as anemia, infections, vaso-occlusive crises, stroke, cardiopulmonary conditions, and organ damage, all of which vary widely in severity and presentation among individuals. While HSCT remains the only curative treatment, and gene therapy is still under clinical investigation, most patients rely on symptomatic therapies, which underscores the pressing need for a personalized approach to disease management.

Given the variability in disease expression and the multiplicity of available treatments, there is a significant unmet clinical need to better define individual disease severity profiles and predict specific complications. The GENOMED4ALL project aims to address this challenge through the development of advanced AI algorithms that integrate genomic, metabolomic, radiomic, and comprehensive clinical data. These tools are designed to enhance our understanding of the disease and support precision medicine approaches in SCD.

In collaboration with AI specialists, clinical objectives have been translated into research questions that guide model development. Key goals include stratifying patients based on their genomic and metabolomic profiles, identifying correlations with inflammatory markers and clinical phenotypes, and utilizing brain MRI data through radiomics to detect silent infarctions, both for early prediction and diagnosis. Additionally, the integration of diverse data types through multivariate analyses and ML models will enable the development of predictive risk scores for major SCD complications such as strokes, vaso-occlusive crises, acute chest syndrome, kidney disease, liver failure, and retinopathy. These predictive tools aim to support early intervention strategies and optimize care pathways tailored to individual patients' risk profiles.

*Study Design*

The study design for the GENOMED4ALL SCD use case is built on a prospective, non-interventional cohort model, aimed at investigating the correlation between genomic data and clinical phenotype to prevent both acute and long-term complications in patients with SCD. Primary data collection began in January 2022 and is expected to conclude in June 2023, with approximately 1,000 patients to be enrolled across clinical institutions in Spain, Italy, France, and the Netherlands. Eligible participants must be at least one year old, of any SCD genotype, not previously treated with HSCT, and enrolled at a steady clinical state. Ethical approval is obtained independently by each participating center, and informed consent is secured according to local regulations.

The study collects a wide array of standardized data through electronic case report forms (eCRFs), including demographic, clinical, laboratory, radiological, genomic, and treatment information. Acute complications requiring hospitalization within the past 24 months, major complications over the patient's lifetime, and chronic manifestations are all captured. Laboratory values are recorded during routine visits when patients are stable, and specialized tests (such as rheological assessments using Lorrca Oxygenscan, genetic variant analysis, and metabolomics profiling) are also incorporated. Brain imaging data, particularly FLAIR sequences from cerebral MRI, are included as part of the radiological dataset.

To ensure data harmonization and quality, the study leverages the Rare Anemia Disorders European Epidemiological Platform (RADeep), which enforces EU-aligned data standards and performs multi-layered quality control. These include real-time validation, minimized free-text entry, completeness checks via relational database constraints, and asynchronous error detection to enhance integration accuracy.

AI methods will play a central role in data analysis. Unsupervised learning techniques will initially be employed to address the phenotypic heterogeneity of SCD, using clustering algorithms and dimensionality reduction tools such as UMAP and Topological Data Analysis (TDA). These approaches will help identify hidden patterns and relationships within high-dimensional clinical and molecular data. For later-stage analyses, supervised machine learning models will be applied to transform raw data into clinically actionable insights, stratify patients by risk profiles, and inform personalized treatment strategies. Ultimately, the development and application of these AI algorithms are expected to support individualized care by enabling more precise prediction of complications and improving long-term outcomes, including for newly diagnosed patients.

*Clinical Evaluation and Validation*

The clinical evaluation and validation of AI models within the GENOMED4ALL SCD use case are critical to ensuring their accuracy, robustness, and practical utility. This process is fundamentally structured around an FL framework, where models are trained using datasets from multiple institutions without sharing sensitive data, and validated against entirely independent patient cohorts adhering to the same protocols and data standards. A continuous collaboration between clinical and AI experts is central to this approach, fostering iterative refinement of the models through joint interpretation of results, identification of weaknesses, and application of clinical insights to enhance predictive performance.

Validation is carried out in two distinct phases. The initial phase emphasizes research and preparation for AI development, including data integration, harmonization, and exploratory analysis. The subsequent phase involves the training of AI algorithms and the validation of their outcomes. Key internal milestones were established: the first data submission occurred in September/October 2022 for the initial training and evaluation of AI models, while a second submission in March 2023 provided additional data for model validation. Clinicians assessed the early results for accuracy, interpretability, and clinical relevance, guiding further refinement by AI experts. This collaborative cycle ensures that AI outputs evolve to become meaningful clinical tools.

In addition to these phases, further validation efforts are ongoing, including initiatives to secure additional funding aimed at testing AI models using synthetic datasets under the Synthema project. Overall, the design ensures a rigorous, clinician-guided validation process that enhances both the credibility and clinical applicability of the AI models developed within the project.

**3. MM Use Case**

*Rationale and Objectives of the Study*

MM is a complex and genetically heterogeneous plasma cell malignancy, marked by the proliferation of monoclonal plasma cells predominantly in the bone marrow. A significant proportion of patients present with high-risk features at diagnosis, such as adverse cytogenetic abnormalities and extramedullary disease. Among these, minimal residual disease (MRD) status (particularly when assessed both within and outside the bone marrow) is emerging as a critical prognostic factor, strongly influencing treatment strategies and long-term outcomes.

Recent advancements in imaging technologies, including whole-body MRI and PET/CT, have established their utility beyond clinical trials, providing valuable diagnostic and prognostic insights. In parallel, innovative approaches like liquid biopsy offer a promising, less invasive alternative to traditional bone marrow sampling, potentially overcoming current limitations by capturing spatial and clonal heterogeneity of the disease.

Despite these developments, the integration of imaging data and liquid biopsy findings with genomic information remains an unmet clinical need. This gap limits the potential for personalized treatment approaches in MM. The study aims to explore whether radiomic analyses can uncover meaningful correlations between imaging features and molecular characteristics derived from bone marrow or liquid biopsy. The overarching objective is to improve the prediction of disease progression, treatment response, and MRD status through a multi-modal, precision medicine approach, ultimately refining therapeutic planning and patient management in MM.

*Study Design*

To address the complexity and spatio-temporal dynamics of MM, a combination of stochastic modeling and advancedAI methods will be employed. Specifically, the study will utilize Physics-Informed Deep Learning, integrating discrete diffusion equations and Markov models to capture disease dynamics. The genomic profile of MM will be analyzed through mutation co-occurrence and clonality assessments using unsupervised, non-parametric Bayesian approaches (for instance, Gaussian and Dirichlet processes) embedded within neural networks. Additionally, a hybrid AI architecture combining unsupervised feature extraction and supervised learning will be implemented to enhance interpretability and statistical rigor of the results.

A core component of this research involves a prospective radiomic and radiogenomic study using PET-CT imaging in 300 MM patients who have achieved complete response following initial therapy. The objective is to correlate imaging features with genomic data to identify predictive signatures for residual disease and patient outcomes, including progression-free and OS. The AI framework will support extensive radiomics feature extraction, alongside state-of-the-art ML techniques such as support vector machines, bagged trees, and random forests for feature selection and predictive modeling.

Furthermore, the study aims to assess the ability of radiomics and radiogenomics to predict genetic alterations both at diagnosis and at the point of best therapeutic response, evaluating the sensitivity and specificity of these techniques in detecting residual disease. Survival outcomes will be modeled using both non-parametric methods like Kaplan-Meier and semi-parametric techniques such as CoxBoost, leveraging the most predictive radiomics and genomic markers.

The study applies a comprehensive AI-driven approach across various data types, ranging from next-generation sequencing and gene expression profiles to PET-CT imaging and liquid biopsy, to describe MM complexity, map its evolution, assess progression risk, and develop predictive radiogenomic models.

### Supplementary Information 29| Deliverable 7.3 - Report on First Wave of Trials

#### 1. Introduction

Haematological diseases (HDs), many of which are rare and life-threatening, present a significant public health challenge due to their chronic nature and the multidisciplinary care they require. Despite major advancements in understanding the complex genetic and physiological pathways of these conditions, progress in personalized medicine remains limited by the lack of reliable biomarkers to monitor disease progression, predict prognosis, and assess treatment response. Research efforts are further hindered by the fragmentation of clinical and -omics data, which prevents researchers from accessing critical patient numbers necessary for robust basic and clinical studies.

Patient registries are essential to overcome these limitations, as they facilitate the study of disease natural history, treatment effectiveness, safety, and quality of care. However, until recently, the absence of unified standards for data collection and structuring has resulted in a multitude of uncoordinated and non-interoperable databases across Europe, particularly in the field of rare haematological diseases (RHDs), which affect fewer than 5 in 10,000 individuals.

In response, the European Union implemented Directive 2011/24/EU, establishing European Reference Networks (ERNs) in 2017 to foster collaboration among Centers of Expertise across Member States. ERN-EuroBloodNet was created to focus specifically on RHDs, bringing together over 100 multidisciplinary teams from 24 EU countries and supported by patient advocacy groups. To tackle the fragmentation of patient data, the EU Rare Disease Platform and its European RD Registry Infrastructure (ERDRI) were developed, offering key components like the European Directory of Registries, Central Metadata Repository, and pseudonymization services to make RD data findable and accessible at the European level.

Under this framework, the European Rare Blood Disorders Platform (ENROL) was launched as the central registry platform of ERN-EuroBloodNet. ENROL aims to harmonize data from existing and new RHD registries, promote interoperability standards aligned with the EU RD Platform, and facilitate data sharing in compliance with GDPR. The platform seeks to generate high-value, EU-wide datasets that capture demographics, survival, diagnostics, genetics, clinical manifestations, and treatment data. Its Common Data Set (CDS), composed of 79 elements including EU core data, is designed to support epidemiological surveillance and research cohort generation.

ENROL also provides a legal and ethical framework for secure data sharing and reuse. Access requests are evaluated by a Data Access Committee composed of clinicians, patient representatives, and legal experts to ensure alignment with ENROL's mission.

Building upon this ecosystem, the GenoMed4All project seeks to harness trustworthy and explainable AI, using advanced ML techniques and FL approaches to improve research and clinical care in HDs. The project covers three primary use cases demonstrating the potential of AI to derive meaningful insights without compromising data privacy. GenoMed4All will leverage data from ERN-EuroBloodNet, harmonized through ENROL and its sub-registries, and will focus on legal and ethical coordination, data availability, standardization, and analysis of early trial outcomes across the three use cases.

**2. Legal and Ethics Issues**

The legal and ethical framework of GenoMed4All is designed to ensure compliance with GDPR and uphold the highest standards in the handling of personal health and -omics data. Recognizing the consortium's role as a group of multiple legal entities sharing mutual interests in processing such data, a JCA was established to clearly define the roles, responsibilities, and legal obligations of each partner. This agreement ensures all parties adhere to ethical, regulatory, and contractual standards, particularly as even anonymized data must be processed by Data Controllers to prepare it for machine learning applications. Based on European Commission templates and informed by a project-wide DPIA, the JCA was developed, refined, and signed by relevant partners for each of the three use cases.

In parallel, each use case adheres to strict ethical standards, requiring informed consent and ethics committee approvals at participating clinical sites. For the MDS use case, a large, multi-centric retrospective study involving nearly 25,000 patients aims to improve personalized medicine through AI by enhancing prognostic tools and treatment strategies. Although explicit consent for GenoMed4All was not initially required due to the retrospective nature of the data and prior consents in place, full ethics approval was secured.

The SCD study follows a prospective, non-interventional design and targets 1,000 patients across several European centers. It aims to integrate genomics, clinical data, and imaging to support AI-driven personalized care. While most centers rely on prior consents permitting secondary use of data, new consents are being obtained where necessary. Ethics approvals have been secured for most centers, with some pending.

The MM use case is a retrospective study focused on exploring the potential of radiomics and genetic data to refine therapeutic decisions. It includes two patient cohorts and additional data from public repositories. Although the collection of extensive clinical, imaging, and genomic data is underway, formal ethics approval and explicit consent for GenoMed4All are still pending.

Overall, GenoMed4All ensures data governance is legally robust and ethically sound, supporting its mission to advance precision medicine through responsible use of patient data across Europe.

**3. Use Cases - 1st Trials Execution**

*MDS Use Case*

The first wave of trials in the MDS use case involved analyzing diverse study populations and data types aligned with four clinical aims: prevention through genomic screening, omics-based classification and prognosis, clinical decision-making, and drug repurposing. The data sets included thousands of patients and healthy elderly subjects, with extensive genomic, clinical, and transcriptomic information. Efforts began to standardize data collection through collaboration with the ENROL initiative, which aims to harmonize common data elements across rare haematological diseases, ensuring interoperability between registries.

Preliminary results focused primarily on aims two and three. For aim two, the consortium validated the Molecular International Prognostic Scoring System (IPSS-M) for MDS prognosis on a large independent cohort, confirming its clinical utility in improving risk stratification and treatment decision-making. Additionally, ML and DL models were developed and validated using multi-modal genomic and clinical data, demonstrating superior predictive accuracy compared to traditional methods. FL approaches enabled multi-center collaboration while preserving data privacy. These models produced explainable, robust patient classification and survival predictions, providing a foundation for personalized medicine in myeloid neoplasms.

Regarding aim three, work progressed on a clinical decision support system designed to optimize the timing of allogeneic stem cell transplantation in MDS patients. Using semi-Markov multistate models, the system simulates patient outcomes under different transplant timing scenarios, assisting clinicians in making personalized treatment decisions.

Collaborations have been established to expand the use case and validate AI models, including partnerships with the Harmony Alliance for data integration and external validation, and the Moffitt Cancer Center to harmonize conflicting classification systems for hematologic malignancies through data-driven approaches. Engagement with ERN-EuroBloodNet's network of European clinical centers is planned to further refine and validate models using standardized data. A forthcoming document will specify minimal data requirements for new clinical sites wishing to contribute to the MDS use case.

*SCD Use Case*

The SCD component of the GenoMed4All initiative is a comprehensive research program aimed at developing predictive models for critical clinical outcomes using integrated clinical and multi-omics data. The project encompasses five primary objectives: to stratify SCD patients based on genomic and metabolomic profiles and assess their correlation with inflammation markers and clinical phenotypes; to develop AI-based models to detect and predict silent cerebral infarcts using brain MRI (radiomics); and, ultimately, to create risk scores for the most severe and prevalent clinical complications such as stroke, vaso-occlusive crises, and acute chest syndrome.

Since January 2022, over 800 SCD patients across five European centers in France, Italy, Spain, and the Netherlands have been enrolled, with a target of 1,000. A rich dataset has been collected for each participant, including clinical records, laboratory tests, GWAS data, metabolomics, radiomics, and functional assessments of red blood cells. All data are standardized through RADeep, using harmonized formats and internationally recognized ontologies to ensure consistency and quality across participating sites.

AI methodologies span both unsupervised and supervised approaches. Initial clustering of patients is performed using genomic and metabolomic data to uncover molecular subtypes and their associations with disease severity. For radiomics, DL models such as UNet ensembles have been trained to detect silent infarcts, followed by logistic regression to distinguish pathological from benign findings. An active learning loop with manual expert validation supports continuous model refinement, leveraging FL and high-performance computing resources like the Marconi100 supercomputer.

Subsequent aims focus on building predictive models for key SCD complications. Supervised learning techniques, (Cox regression, random survival models, and neural networks) are applied to predict outcomes over time. These models are trained and validated using federated data sources, maintaining patient privacy while enhancing model generalizability.

Clinical validation is a critical component, involving continuous collaboration between AI developers and clinicians to ensure models are accurate, interpretable, and clinically actionable. The validation process is structured in two phases: an initial research phase to develop algorithms and a second phase dedicated to validating model performance using new, independently collected data. This iterative cycle ensures that AI outputs are robust and can be translated into meaningful clinical tools to support decision-making and improve patient care in rare haematological disorders such as SCD.

*MM Use Case*

The current study addresses a significant challenge in MM research: the inability of existing risk scores, such as R-ISS and IMWG, to adequately capture patient heterogeneity and accurately identify high-risk individuals. These traditional methods often exclude critical genomic and disease response dynamics, limiting their prognostic power. Given the toxicity of treatments, there is a pressing need for early, non-invasive prediction tools that can inform clinical decisions and improve patient outcomes.

To meet this need, the proposed research hypothesizes that integrating multimodal data can enhance risk stratification and reveal actionable clinical endpoints for precision therapy. AI approaches will be employed to identify key markers that distinguish patients likely to relapse within one year of treatment initiation, regardless of therapy type.

The study will analyze two cohorts of newly diagnosed MM patients. The first, comprising 110 individuals, will include comprehensive datasets such as PET-CT imaging, cfDNA-based liquid biopsy, and 11,817 measures of copy number alterations (CNAs) derived from both tumor and cfDNA samples. The second cohort includes 279 patients, with available baseline clinical data, imaging, and CNAs from SNP arrays. All datasets have been standardized, with genomic data normalized for statistical modeling and PET/CT images adjusted for patient-specific variables where possible.

AI models will incorporate physics-informed DL techniques, including Laplacian dynamics and Markov modeling, to interpret MM's spatial and temporal complexity. Genomic profiling will utilize both unsupervised methods (such as Gaussian and Dirichlet process-based neural networks) and supervised learning layers to ensure interpretable, statistically robust results. Radio-genomic analyses will aim to discover predictive signatures that correlate PET-CT imaging with genomic markers, focusing on outcomes like PFS and OS.

Initial analyses of a subset of 89 MM patients have utilized semi-automated PET segmentation techniques to extract imaging features from skeletal regions, which are now being used to predict clinical outcomes using penalized survival models. Depending on data availability, both non-parametric (Kaplan-Meier) and semi-parametric (CoxBoost) methods will be applied to model EFS.

Ultimately, this study aims to define MM complexity and progression risk through a powerful fusion of AI and multi-omic data integration, thereby improving early risk stratification and supporting individualized therapeutic strategies. The project also outlines the potential for expanding collaborations and onboarding new clinical sites to scale and validate its findings.

**4. Genomed4All Platform Implementation**

Genomed4All aims to harmonize and standardize diverse datasets by aligning data types, formats, sources, and concept codifications into a unified structure. The central goal is to offer an implementation guide that enables any data provider to convert their data into a common model compatible with the Genomed4All Platform. As outlined in D3.3, the project adopts the FHIR (Fast Healthcare Interoperability Resources) standard, making the identification and definition of appropriate FHIR resources fundamental to platform development.

The standardization process begins by analyzing CSV files for each use case to identify data items required for ML training. Each item is then mapped to a suitable FHIR resource, for example, gender is mapped to the 'Patient' resource, while measurements like blood pressure and cholesterol levels are captured through the 'Observation' resource.

For clinical data, standard codes have been assigned to each item using Athena, which maps terms to OHDSI's standardized vocabulary, ensuring consistency across datasets. Genomics data, whether processed by Genomed4All's pipelines or external providers, follows a similar mapping approach. However, codification varies across use cases to preserve clinical meaning; for instance, MDS data may be consolidated into a single Observation resource, while SCD may require both Molecular Sequence and Observation resources, in line with FHIR's genomics implementation guidance.

Radiomics data, unlike genomics data, is not processed by Genomed4All but is expected to be pre-processed and incorporated using Observation resources.

D5.4 provides a detailed account of the procedures implemented within the Data Homogenization Platform, including data standardization, ingestion into the FHIR-based common model, and feature extraction to prepare clinical and genomics information for machine learning applications.

#### 5. Conclusions

This deliverable outlines the initial steps undertaken to coordinate and implement the first wave of clinical trials focused on developing and validating the Genomed4all platform and its associated AI models across three selected clinical use cases. It highlights the identification of key barriers to platform deployment, including legal, ethical, and technical challenges encountered at the clinical site level. Additionally, it references the detailed reassessment and refinement of the platform's design, as documented in D4.3.

### Supplementary Information 30| Deliverable 7.4 - Report on First Trials Validation and Evaluation

#### 1. Genomed4All Platform Evaluation

The GenoMed4All federated platform is designed with a dual-component architecture, comprising a central server (Manager-Api) and multiple distributed edge servers (Worker-Api). The central Manager-Api handles tasks such as model registration, training supervision, and aggregation, while the Worker-Api nodes, typically hosted within individual hospitals, manage localized data and model training in a secure environment. These components function in tandem, supported by a data homogenization pipeline that ensures standardized input across institutions.

The development of the platform followed a multi-phase approach. The initial phase focused on deploying a centralized version using a subset of pseudo-anonymized data to facilitate model exploration and refinement. The subsequent phase enabled full federated training, integrating additional hospital nodes to enhance model generalization and performance by leveraging broader datasets.

Usability and reliability tests were conducted at the Humanitas Research Hospital (ICH) to assess the platform's capabilities and ease of use. Both Manager-Api and Worker-Api components were installed and evaluated. Through the user interface and REST-API endpoints, key functions were validated, including user management via Keycloak, node connectivity, dataset registration, model upload (using .pkl files), and federated task execution.

ICH successfully demonstrated that hospitals could securely upload datasets to local Worker-Api nodes and connect them to the central Manager-Api for federated model training. Tests confirmed the ability to run AI models, monitor training tasks in real-time, and access detailed performance metrics. While the platform proved functional and reliable, certain interface elements, particularly dataset and node registration, could benefit from further usability enhancements. Overall, the platform shows strong promise in enabling secure, decentralized AI training across clinical institutions.

#### 2. Integration and Testing of Federated Algorithms

The integration of federated algorithms into the platform has progressed significantly, with deployments completed at both the ICH hospital and on CINECA servers. Initial tests included training a simple classification model using the MNIST dataset, confirming the platform's core capabilities. The infrastructure is now prepared to host the WP6 models, which must be adapted for platform compatibility. As a first step, UPM has integrated its SAVAE model using synthetic MDS data. This serves both to validate the platform and provide a reference guide for other partners, streamlining future integrations.

Platform functionality was tested in two environments: ICH focused on usability and deployment procedures, while UPM evaluated technical aspects of federated training using varied node types and data completeness levels. These nodes simulate realistic data availability conditions, and the tests explored four scenarios with different data distributions across nodes. All experiments were conducted with synthetic data generated by a VAE developed by UPM, designed to resemble real datasets in both structure and utility.

The federated platform is built on FLOWER and supports multiple training nodes with varying data completeness. The SAVAE model, implemented in PyTorch, was adapted for federated training through a modular script that packages the model as a Pickle object, compatible with the platform. This object includes key methods for strategy definition, model architecture, and training logic. SAVAE employs a FedAvg strategy for weight aggregation and supports necessary functionalities such as training, prediction, evaluation, and model weight management. Supporting scripts also ensure consistent data loading and preprocessing across nodes.

The full codebase is available publicly to facilitate replication by other WP6 partners. Key technical requirements include Python 3.8 and Cloudpickle 2.1.0, and it is recommended to consult the FLOWER documentation for deeper insights into customizing server-client communication or defining new strategies. Through this structured approach, future model integrations into the federated platform are expected to be more efficient and standardized.

To verify the consistency of results obtained through the FL platform, a local simulation was conducted. Although optional, this simulation provides valuable insights into client-server interactions and enables performance analysis through graphical outputs. The process involved setting up a local server and three client nodes as defined in the Validation Protocol, following the guidelines provided in the Flower framework documentation.

The server is initiated using fl.server.start_server(), loading the model strategy from a pre-generated Pickle file. Each client implements the FlowerClient class, extending fl.client.NumPyClient, and defines the required methods get_parameters(), fit(), and evaluate(). These methods enable data loading, model training, and performance evaluation at each round. Logs generated during the simulation provide visibility into the number of active clients and the status of communication during each training and evaluation round.

While the current local implementation does not display detailed metrics such as the CI during training, manual extraction of this metric was included to monitor model performance. The results across multiple scenarios showed a general improvement in CI values over five aggregation steps, confirming the benefits of federated training. In particular, clients with limited or lower-quality data showed performance gains through aggregation with other clients, as seen in scenarios with asymmetric data distribution. Comparisons with centralized training revealed that FL helped stabilize performance in underperforming nodes.

Following local validation, the same model and associated data files were uploaded to the FL platform. The platform facilitates task execution across clients by allowing the user to designate data paths and monitor training in real-time. Each round consists of local training, aggregation of model weights at the server, and redistribution of the updated model to clients for further training. Once training concludes, performance metrics are available for download and analysis.

The results observed on the platform aligned closely with those from the local simulation, particularly in Scenarios 1 through 3. In Scenario 4, where data volume and quality varied significantly between clients, FL again demonstrated clear benefits, especially in clients with initially poor performance. These results validate the effectiveness of the FL approach and the robustness of the implementation. Future improvements may focus on scaling up data volume to reduce variability and enhance the reliability of the conclusions drawn.

The integration of the UPM model, SAVAE, into the FL platform has been successfully completed, producing consistent results across various scenarios. Looking ahead, the next objective is to establish a comprehensive validation protocol that does not rely on real data. Currently, a small portion of the training dataset is used for validation, which may not provide sufficiently robust conclusions. To address this, the plan is to generate a new set of synthetic data samples and apply them uniformly across all clients to ensure comparability. This will require adjustments to the client data loading process within the platform. Once these modifications are implemented and validated, other WP6 partners will be expected to follow the same integration and testing procedures to ensure the consistent and correct operation of their models within the platform.

#### 3. Pilot Case using MOSAIC framework

This section presents a pilot case study demonstrating the application of an FL framework in a simulated environment, using the MOSAIC project as a representative example. MOSAIC is an AI-based framework designed for multi-modal classification and personalized prognostic assessment in rare cancers, with a specific focus on MDS. The goal is to assess the potential of federated survival analysis within a realistic clinical scenario by enabling multiple institutions to collaboratively train a model while maintaining data privacy.

To simulate the federated setup, a single machine is configured to represent three nodes using the FedAvg approach. This method, known for its robustness to non-IID and unbalanced data distributions, combines local stochastic gradient descent (SGD) updates from each node with centralized model averaging. The CPH model, a standard in survival analysis, is employed and trained using SGD to ensure consistency in the federated context.

Each node is preprocessed identically and works with event-time combinations for LFS, OS, and EFS. The training process spans 40 epochs, with weight sharing every five epochs, and performance is evaluated using the CI metric under a 3-fold cross-validation protocol. Three experimental configurations are used for comparison: centralized training with the full dataset (Setting A), isolated training on local data without sharing (Setting B), and federated training using FedAvg (Setting C).

In the simulation, node one possesses 60% of the total data, node two holds 30%, and node three contains 10%, with considerable missing covariates. Results reveal that centralized training yields the highest performance, setting an upper bound. Node one, due to its large and complete dataset, achieves nearly equivalent results even when trained in isolation. Node three, impacted by data scarcity and missing features, shows lower performance in isolation but benefits significantly under FedAvg, as model aggregation allows it to leverage insights from nodes with more comprehensive data. The federated setup overall demonstrates improved generalization, especially for less-informed nodes, without compromising data privacy.

Graphical analysis of the training progress indicates that performance, measured by the CI, notably improves following each communication round in the federated environment, particularly benefiting nodes with limited data. These results validate the use of FL for collaborative prognostic modeling in healthcare, offering both performance gains and data confidentiality.

Ultimately, the MOSAIC framework exemplifies how FL can be applied effectively in real-world clinical contexts, enabling high-performing, privacy-preserving predictive models for rare diseases like MDS.

#### 5. Homogeneization Platform and Associated Tests

The Genomed4All data homogenization platform is a foundational element supporting FL by ensuring uniformity and compatibility across diverse data sources such as EHRs, medical imaging, and genomic data. It achieves this through a comprehensive process of data transformation, preprocessing, validation, and cleansing, ultimately converting data into the standardized FHIR format. Key preprocessing techniques address discrepancies in structure, volume, and quality through normalization, missing data management, and feature engineering.

The platform architecture is built around two core processes. First, an ETL pipeline reads the raw data from providers, applies necessary transformations to align it with the common data model defined by FHIR resources, and then loads it into an integrated data repository such as Elasticsearch or MongoDB. Given the variability of use cases and data types, the platform allows for tailored executor modules to support custom ETL workflows.

Second, a feature extraction service retrieves and filters relevant diagnostic data from the FHIR resources based on predefined codes. The extracted features are saved in CSV format to facilitate interoperability with ML workflows. To enhance usability, a proof-of-concept user interface has been developed, providing a more intuitive interaction with the platform.

Testing of the platform is conducted individually for each use case. Once data is submitted in the agreed-upon format, the ETL process is executed to transform and load the data into FHIR resources. Success is measured by the accurate mapping of all specified features into their corresponding FHIR structures. Subsequently, the feature extraction process is validated by confirming that the relevant data stored in the FHIR server is correctly retrieved and exported into CSV files for further analysis.

**6. Conclusions**

As outlined in the draft, platform components have been tested independently by ICH and UPM, with the exception of the homogenization platform, for which DED is expected to provide installation and testing guidance to the participating nodes. The federated platform is currently operational, having been deployed at CINECA as the central node and at ICH as a local node. Initial usability and reliability tests prompted several enhancements, all of which have been successfully implemented. A revised user guide, incorporating solutions to previously encountered technical issues and documented bugs, has been distributed to the remaining nodes to facilitate installation.

The platform's deployment across other hospitals is progressing, with varying degrees of completion. Integration of federated algorithms has begun, starting with the OS algorithm from the MDS use case, which required code adaptations to align with central libraries on the CINECA server. A detailed guideline outlining the necessary steps for integrating algorithms from all three use cases will be released shortly to support this process.

The MOSAIC pilot case has successfully demonstrated the platform's capability to operate effectively within a simulated federated environment, confirming the validity of the approach across multiple algorithms. Future testing will focus on integrating at least one algorithm per use case across three hospital nodes and evaluating the homogenization platform's capacity to standardize data from one dataset per use case.

### Supplementary Information 31| Deliverable 7.5 - Report on Second Wave of Trials

**1. Introduction**

HDs, many of which are chronic and life-threatening, represent a significant public health challenge due to the complexity of their diagnosis, treatment, and management. Despite considerable progress in understanding their genetic and pathophysiological underpinnings, efforts toward personalized medicine are hindered by the absence of robust clinical markers and fragmented research data, especially in the context of rare haematological diseases (RHDs). Patient registries are critical for tracking disease progression, treatment efficacy, and care quality, yet the lack of standardized, interoperable data collection methods has historically limited research impact.

To address this fragmentation, the European Union has established the ERNs, including ERN-EuroBloodNet, launched in 2017 to unify efforts across 24 Member States, encompassing 97 multidisciplinary teams and more than 450 RHDs. ERN-EuroBloodNet, supported by the European Haematology Association (EHA) and patient organizations, promotes cross-border collaboration and data sharing through its umbrella registry platform, ENROL. ENROL ensures interoperability, data quality, and GDPR-compliant data reuse, facilitating EU-wide epidemiological mapping and cohort identification for clinical research. It incorporates a standardized Common Data Set (CDS) with 79 elements to support comprehensive surveillance and research engagement.

Aligned with the EU Rare Disease Platform and supported by ERICA, ENROL integrates new and existing registries, improving access and utility of RHD data for medical, research, and industry stakeholders. The legal framework managed by a dedicated Data Access Committee ensures ethical and secure data usage.

Furthermore, the GenoMed4All project builds upon this infrastructure to demonstrate the benefits of trustworthy AI in personalized medicine. Through FL and advanced machine learning models, GenoMed4All facilitates secure, non-disclosive data utilization across multiple patient cohorts. Its efforts focus on clinical use cases including MM, MDS and SCD, with results from trial phases highlighting advancements in data standardization, ethical governance, and research outcomes.

**2. Legal and Ethics Issues**

*Joint Controller Agreement (JCA)*

The JCA within the GenoMed4All project was established to ensure that all partners comply with the legal, ethical, and regulatory standards required for the processing and reuse of personal health and -omics data. Under the GDPR, it is essential to define the roles of all parties involved in data processing and formalize these roles through a binding agreement. Although the processed data is ultimately anonymized for ML purposes, the original data providers, acting as Data Controllers, must handle personal data during its preparation, thereby necessitating compliance with data protection regulations.

To meet these obligations, GenoMed4All implemented a JCA, aligning with guidance from the European Data Protection Board. The process included defining the responsibilities of each party, selecting a suitable JCA template based on European Commission standards, and using the project-wide DPIA to outline the specifics of processing activities. Feedback was collected from all parties to refine the agreement, ensuring consensus and legal preparedness.

The agreement distinguished two parties with Processor roles: DEDALUS as the Data Processor and CINECA as a Sub-Processor. All other involved entities, given their role in determining the purposes and means of processing, were identified as Joint Controllers. Separate JCAs were prepared and signed for each clinical use case to involve only the relevant parties in each case. The finalized agreements were signed in February 2023, with further documentation detailed in deliverables D2.2 and D7.3.

*Ethics Board and Informed Consent*

*MDS*

The MDS study under GenoMed4All is a multi-centric, retrospective, observational, and non-interventional research initiative designed to advance personalized medicine through artificial intelligence. By integrating diverse data sources and developing ML tools, the study aims to deepen understanding of MDS biology, improve the prognostic and predictive accuracy of existing clinical tools, and enable more precise, targeted treatment strategies.

The study population is substantial and diverse, including 11,000 healthy elderly individuals over 65 years of age, alongside 6,700 MDS patients with clinical and targeted mutation data, 1,200 with RNA-sequencing, 900 with whole exome/genome sequencing, 1,500 transplant recipients, and 2,400 patients with various hematological malignancies possessing genomic and gene expression data. The cohort shows an unbalanced gender distribution, with a predominance of male participants.

Eligibility criteria include patients aged 18 or older with a confirmed MDS diagnosis based on the 2016 WHO classification and the ability to provide informed consent per ICH/EU GCP and national regulations. Patients without documented informed consent or lacking clinical or omics data are excluded.

Ethical clearance has been secured, with approvals dating back to January 27, 2021. A total of 4,780 retrospective patients have been included under secondary data use agreements, ensuring compliance with ethical and regulatory standards.

*SCD*

The SCD study conducted under GenoMed4All is a multi-centric, prospective, non-interventional study that adheres to standard care procedures for SCD patients. Its primary objective is to advance personalized medicine by developing and validating AI algorithms that integrate genomic, multi-omics, laboratory, imaging, and clinical data.

The study targets a cohort of 1,000 patients diagnosed with SCD across all genotypes, as classified by ORPHANET. Given the disease's equal prevalence among males and females, the study population is expected to be gender-balanced. Eligible participants include both pediatric patients older than one year and adults, provided they are able and willing to give informed consent, either personally or through a legally designated representative. Informed consent can be obtained at any point during follow-up, prior to data entry into the electronic case report form. Patients who have undergone stem cell transplantation or gene therapy, or those aged one year or younger, are excluded from the study.

Several clinical sites across Europe are participating, with varying degrees of ethics clearance and consent protocols. While some centers operate under previous consent that allows for data reuse, others have obtained explicit consent and ethics committee approval specific to GenoMed4All. Ethics approvals have been documented between March 2022 and July 2022, with full details provided in D7.3.

*MM*

The MM use case within the GenoMed4All project is structured as a retrospective study focused on assessing the potential of radiomics to uncover correlations between imaging features and molecular genetic characteristics. These include biomarkers derived from both bone marrow clones and liquid biopsies, with the ultimate goal of enhancing precision in therapy planning for MM patients.

The study encompasses two primary patient cohorts. The first includes 110 newly diagnosed MM patients, for whom comprehensive data were collected, including baseline clinical information, PET-CT imaging, liquid biopsy data (such as circulating tumor DNA fraction and measures of spatial heterogeneity), genomic alterations in the CD138+ tumor clone, treatment responses, and clinical outcomes. These patients were treated with combination regimens that featured novel agents (proteasome inhibitors, immunomodulators, and monoclonal antibodies) with treatment strategies adapted to patient age (autologous stem cell transplantation for those under 65, and continuous therapy for older individuals).

The second cohort includes 279 newly diagnosed MM patients with similar data sets, excluding liquid biopsy details but incorporating copy number alteration data from SNP array analysis. Additionally, publicly available clinical and genomic data from 750 MM patients in the CoMMpass database (release 1A12) supplement the study.

Across both cohorts, disease staging followed the revised International Staging System (R-ISS), incorporating ISS criteria, cytogenetic risk status, and LDH levels to classify patients into three distinct prognostic categories. All data were obtained retrospectively from the University of Bologna's Institute of Hematology, with no additional explicit consent or separate ethics committee approvals required, as the data were already available for research purposes.

**3. Use Cases - 2nd Trials Execution**

*MDS*

During the second wave of trials, the MDS team, comprising both clinical and technical experts, concentrated efforts on three primary objectives, producing significant outcomes that advance the understanding and classification of myeloid neoplasms.

To address the first objective (identifying genomic factors contributing to the development of MDS and enabling early detection of at-risk individuals) the team utilized the MOSAIC framework, an AI-based system for unsupervised multimodal data analysis. In a retrospective study involving over 20,000 MN patients, MOSAIC enabled the identification of 34 clinically distinct clusters by integrating morphological and genomic data. These clusters were linked to different risks of disease progression and survival, culminating in the development of a new pan-MN prognostic score. This integrative approach offers a refined, biologically consistent classification model that addresses limitations in current diagnostic systems and will be presented at the 66th American Society of Hematology (ASH) Annual Meeting in 2024.

Additionally, a focused study on Chronic Myelomonocytic Leukemia (CMML), another form of myeloid neoplasms, analyzed genomic data from 2,471 patients using the same AI framework. Eleven unique clusters were discovered, revealing substantial molecular heterogeneity and areas of diagnostic overlap with other neoplasms. The findings support a molecular-based classification for CMML that may enhance prognosis and treatment strategies. This study was presented at the 65th ASH Annual Meeting in 2023.

Furthermore, MOSAIC has been validated in a broader cohort of 4,427 patients with rare cancers, including MDS, demonstrating superior performance in classification and survival prediction compared to conventional methods. Advanced clustering algorithms, such as UMAP combined with HDBSCAN, provided more precise patient stratification, while AI-driven survival models outperformed traditional statistical tools. The use of explainable AI (SHAP) and FL has improved both model transparency and performance while maintaining data privacy. Collectively, these efforts underscore the transformative role of AI in advancing diagnostic precision, prognostic accuracy, and individualized care for patients with rare hematological malignancies.

As part of its second core objective, the GENOMED4ALL consortium focused on developing predictive models for patients with MDS by integrating comprehensive genomic and clinical information. MDS is characterized by heterogeneous clinical presentations and outcomes, making accurate prognosis and treatment planning a challenge. To address this, the consortium demonstrated that combining multi-layered data (including clinical, genomic, cytogenetic, transcriptomic, and demographic information) significantly enhances the ability to predict overall survival. In a cohort of 389 MDS patients, transcriptomic data emerged as the most influential predictor, accounting for 40% of the explained variance, followed by genomic and cytogenetic features (24%), demographics (20%), and clinical data (15%). This integrative modeling marks a major step toward more refined and individualized prognostication in MDS.

In parallel, the consortium leveraged natural language processing (NLP) to extract valuable clinical insights from unstructured text within patient reports, an often-overlooked source of early phenotypic and biological information. To this end, they developed a domain-specific AI language model named BERTholdo, trained on clinical reports from over 1,300 patients with MDS, myeloproliferative neoplasms (MPN), and AML. This model enabled unsupervised clustering of patient text embeddings, resulting in seven clinically coherent subgroups that aligned with known disease categories and genotypic-phenotypic correlations. Survival analyses validated the clinical relevance of these clusters.

BERTholdo outperformed generic models in key NLP benchmarks, underscoring the value of domain adaptation in medical AI applications. These findings confirm that clinical narratives, when processed through advanced NLP, offer a meaningful data layer that can enhance multimodal risk stratification and support next-generation personalized medicine models. The work has been presented at leading hematology conferences, further supporting the clinical relevance and potential of these innovations. The consortium is now advancing this line of research by exploring the application of large language models to further enhance precision in hematological care.

The GENOMED4ALL consortium addressed the challenge of limited availability of multimodal data in hematological malignancies by exploring the use of synthetic data generation through AI. These diseases are rare and complex, often requiring comprehensive clinical, genomic, and histopathological information to guide diagnosis, prognosis, and treatment. In particular, collecting cytological and histological bone marrow images that accurately reflect disease morphology is both resource-intensive and logistically difficult. To overcome this, the consortium developed a pipeline that uses generative models to produce synthetic images conditioned on textual pathology reports, which are more readily accessible and rich in clinical detail.

A Stable Diffusion model was fine-tuned on hematological datasets to generate realistic Hematoxylin and Eosin (H&E) images for 200 patients with myeloid neoplasms, including MDS, AML, and MPN. Textual conditioning was enabled through the BERTholdo language model, ensuring the generated images corresponded to specific pathological descriptions. To validate the authenticity and utility of these synthetic images, a Synthetic Images Validation Framework (SIVF) was created, which compared key morphological, color, and texture features between real and synthetic datasets. Results demonstrated a high degree of similarity in feature distributions and correlations, both across the overall dataset and at the individual patient level.

The consortium then assessed the impact of synthetic data augmentation on disease classification and survival prediction. Using morphological features extracted from both real and synthetic images, machine learning models, including XGBOOST classifiers, showed a 10% improvement in F1 Score for disease classification tasks when trained on combined datasets. Furthermore, when synthetic data were incorporated into prognostic models alongside real patient data, there was a greater than 10% improvement in the CI for predicting overall survival. Notably, synthetic image-derived morphological features such as nuclear dimensions emerged as significant predictors.

These findings confirm that AI-generated images faithfully replicate the diagnostic features of real histopathological data and can substantially enhance clinical models through data augmentation. This approach not only improves predictive accuracy but also facilitates data sharing and accelerates research, particularly in resource-constrained or privacy-sensitive settings. The study was presented at major hematology conferences, reinforcing the promise of synthetic data generation in advancing precision medicine in hematological oncology.

The GENOMED4ALL consortium conducted an extensive study to improve classification and personalized prognostic assessment in myeloid neoplasms (MN), with a particular focus on MDS, using a multi-modal and FL approach. MDS is a complex and heterogeneous group of hematopoietic disorders with potential progression to AML. Leveraging next-generation sequencing data, along with demographic, clinical, genetic, and cytogenetic information, the consortium implemented advanced ML and DL techniques to develop predictive and explainable models suited for clinical application.

Using two large patient cohorts, the team applied dimensionality reduction with UMAP followed by clustering techniques such as HDBSCAN and HDP. Classification models trained on these clusters, particularly those based on UMAP+HDBSCAN, demonstrated high accuracy and interpretability, with explainability enhanced through SHAP analysis. In predicting OS, non-linear ML/DL models consistently outperformed traditional CPH, particularly when integrating diverse data types beyond conventional scoring systems like IPSSR. The use of SHAP again confirmed the stability of key predictive features across cohorts.

A key innovation in the project was the use of FL, which enabled decentralized model training across multiple clinical nodes without compromising data privacy. Performance in FL settings was comparable to centralized training, and even small nodes benefited from participation, validating the approach's scalability and inclusiveness. To address missing data, a deep decoder model was introduced, showing strong imputation accuracy for both clinical and genomic features, further enhancing the robustness of the predictive framework.

Looking forward, the consortium is expanding these models to stratify individual responses to specific treatments. Central to this effort is the development of Clinical Decision Support Systems (CDSS), designed to guide clinicians in determining the optimal timing for allogeneic HSCT. Through a Semi-Markov multistate model, the system enables dynamic simulations of patient life expectancy based on various transplantation timelines, facilitating data-driven, personalized treatment planning and shared decision-making.

Overall, the GENOMED4ALL project demonstrates how integrating multi-modal data with advanced AI methodologies can deliver accurate, interpretable, and clinically applicable tools for prognosis and treatment in myeloid neoplasms, laying the foundation for precision medicine at scale.

*SCD*

The GenoMed4All consortium has completed enrollment of approximately SCD patients across multiple European clinical centers in Spain, Italy, France, and the Netherlands, achieving a comprehensive dataset that integrates clinical, genomic, metabolomic, and radiologic information. The primary objectives focus on stratifying SCD patients by their genomic and metabolomic profiles, assessing correlations with inflammation markers and disease phenotypes, and developing predictive risk scores for severe clinical outcomes such as recurrent vaso-occlusive crises, acute chest syndrome, and silent cerebral infarcts. To support these aims, extensive data collection was performed using a standardized electronic case report form (eCRF) capturing 271 variables including demographic data, medical history, laboratory tests, treatment information, and brain MRI scans specifically for radiomics analysis. Data standardization and quality control have been ensured through the Rare Anaemia Disorders European Epidemiological Platform (RADeep), which applies EU guidelines and international ontologies to maintain consistency across centers and facilitate high-quality, interoperable data integration. This robust and harmonized dataset forms the basis for the development of advanced AI-driven models to improve risk prediction and personalized management in SCD.

The clinical challenge addressed in this project stems from the significant phenotypic variability observed among patients with SCD despite sharing the same genetic mutation. The goal is to develop predictive algorithms that integrate multi-omics data alongside demographic, clinical, and laboratory features to forecast individual patient outcomes. The strategy involves several AI-driven approaches: unsupervised clustering of genomic and metabolomic profiles to identify patient subgroups correlated with inflammation markers and disease phenotypes; development of automated radiomics-based algorithms to detect and predict silent cerebral infarcts from brain MRI scans; and creation of predictive risk scores for major clinical outcomes such as recurrent vaso-occlusive crises, acute chest syndrome, and silent cerebral infarcts using multivariate and machine learning models. Genomic analyses rely on high-quality GWAS data processed with stringent quality controls and imputation techniques, while metabolomics data derive from high-resolution mass spectrometry of blood samples analyzed through advanced clustering and dimensionality reduction methods. Radiomics models utilize DL ensembles, supplemented by expert manual refinements and post-processing to ensure accurate segmentation of white matter lesions. Clinical endpoints for model development were carefully selected based on data availability and clinical relevance, with silent infarcts, acute chest syndrome, and recurrent pain crises prioritized, while outcomes like stroke and organ failures are considered for future analyses supported by synthetic data augmentation. Overall, this integrated AI framework aims to improve patient stratification and personalized risk prediction in SCD, addressing the complexity and heterogeneity of the disease through cutting-edge computational methods and validated clinical data.

The AI methodology for this project employs both unsupervised and supervised techniques to analyze complex and heterogeneous SCD data. Initially, unsupervised approaches are used to stratify patients based on genomic and metabolomic profiles, integrating biomarker knowledge and clinical measures such as CRP levels and rheological data. Two complementary unsupervised strategies guide this stratification: one data-driven, identifying unbiased patient subgroups, and one hypothesis-driven, selecting features based on established relevance. Techniques such as HDBSCAN clustering, UMAP, and Topological Data Analysis are applied to uncover both local and global data structures. For predictive modeling of clinical outcomes, supervised methods, including various regression and ML models, are explored. Radiomic analysis incorporates advanced techniques like Manyfold Learning for synthetic MRI data generation and Approximated Bayesian UNet for improved silent infarct detection. The integration of clinical, molecular, and imaging data enables the development of robust prognostic models, which will be validated across multiple centers using FL frameworks to ensure generalizability and privacy. Continuous collaboration between AI specialists and clinicians ensures iterative refinement, with clinical feedback driving model interpretation, error correction, and validation. The validation process occurs in two phases: an initial research-focused phase to build models using early datasets, followed by a training and validation phase where models are tested and improved with additional data. This rigorous evaluation framework, supported by predefined performance metrics and cross-validation, aims to produce clinically actionable tools that enhance patient stratification and support decision-making in SCD care.

*MM*

The current study addresses the unmet need to integrate diverse clinical, genomic, and imaging data in MM patients to improve risk stratification and early prediction of disease progression. Existing risk scores, which rely on limited baseline information and often exclude genomic data or disease dynamics, fall short in reliably identifying high-risk patients who may relapse within one year of treatment. Given the significant side effects associated with therapies, early and precise outcome prediction using non-invasive methods is critical. This study aims to leverage AI-driven approaches to integrate baseline genomic, clinical, and imaging data with longitudinal disease markers such as minimal residual disease (MRD) to identify high-risk patients for tailored treatment strategies.

Two cohorts of newly diagnosed MM patients are included: the first with 110 patients providing baseline clinical, PET-CT imaging, liquid biopsy, and detailed copy number alteration (CNA) data derived from ultra-low pass whole genome sequencing (ULP-WGS), alongside therapy response and follow-up outcomes; the second cohort includes 279 patients with similar clinical, imaging, and CNA data obtained through SNP arrays. Data standardization protocols have been applied to ensure comparability, including normalization of CNAs and PET/CT imaging parameters.

The analytical framework combines stochastic and AI methods, particularly physics-informed DL utilizing diffusion equations and Markov models to capture MM's spatial-temporal complexity. Genomic analyses focus on the CNA landscape, employing unsupervised Bayesian methods embedded in neural networks for patient stratification, complemented by hybrid supervised-unsupervised approaches to maximize interpretability. The radio-genomic study integrates PET-CT imaging with genomic profiles, aiming to identify signatures predictive of minimal residual disease negativity and clinical outcomes such as PFS and OS. Semi-automated segmentation techniques applied to PET images enable extraction of metabolic features for outcome prediction using penalized time-to-event statistical models, including Kaplan-Meier and CoxBoost analyses.

This integrative approach is designed to elucidate MM heterogeneity, improve risk stratification, and support personalized clinical decision-making through advanced AI models validated on well-characterized patient cohorts with comprehensive molecular and imaging datasets.

**4. Use Cases - Onboarding**

An Associate Members Onboarding Handbook has been developed in collaboration with WP9 to facilitate the integration of new members from all three use cases into the GenoMed4All platform. This handbook serves as a comprehensive user guide, providing an overview of the project's context, vision, mission, and team, as well as detailing the AI-driven approach to diagnosis, prognosis, and treatment of hematological diseases. It emphasizes GenoMed4All's dedication to advancing rare disease research through FL, which is presented as a pivotal technology for achieving this goal. The handbook clearly outlines the onboarding process, including the platform's architecture, data dictionaries, timeline, and specific collaboration areas.

In partnership with the ERN-EuroBloodNet Scientific Coordination team, an onsite meeting was held on June 13, 2024, during the EHA annual congress in Madrid. This event brought together 29 specialists in MDS, SCD, and MM, providing a valuable opportunity for direct dialogue about ongoing work within GenoMed4All and Synthema. The meeting facilitated the exchange of insights and tools to support hematological disease research within the ERN-EuroBloodNet framework. Further details regarding the onboarding process for each use case are provided in the subsequent sections.

*Myeloid Disorders*

The ERN-EuroBloodNet Myeloid disorders subnetwork, comprising 47 clinical centers across 18 European Union member states, is positioned to contribute to the validation and refinement of AI models developed within the GenoMed4All project. A key partnership with the Harmony Alliance, a European public-private initiative focused on big data in hematology, aims to integrate Harmony's central data repository with GenoMed4All's federated platform, enhancing the collection and sharing of data on rare hematological cancers with significant unmet clinical needs. Currently, this collaboration centers on developing a Decision Support System for transplantation in MDS, where GenoMed4All provides the learning cohort and Harmony contributes an independent external validation cohort.

Additionally, a new collaboration with the Moffitt Cancer Center seeks to harmonize two recently introduced classifications of hematological neoplasms: the World Health Organization (WHO) 2022 and the International Consensus Classification (ICC) 2022. Given the clinical challenges posed by divergent classification systems, this partnership will leverage datasets from GenoMed and Moffitt, comprising over 7,000 patients characterized by clinical and genomic data, to create a data-driven classification approach. Methodologically, hierarchical Dirichlet models will identify key components, while SHAP values will be applied to ensure interpretability of the classification process.

The ERN-EuroBloodNet Myeloid subnetwork also engaged in an onsite meeting held on June 13, 2024, during the European Hematology Association annual congress in Madrid, where representatives from 11 centers, totaling 16 attendees, participated. This meeting provided an important opportunity for onboarding new centers and fostering collaboration within the network.

*Sickle Cell Disease*

The ERN-EuroBloodNet network for red blood cell disorders comprises 53 clinical centers across 19 European Union member states, poised to contribute to the validation and refinement of AI models developed under GenoMed4All. A survey conducted in April 2021 identified clinical sites willing to participate in the SCD use case, gathering information on available demographic, laboratory, clinical, radiological, genomic, and rheological data, as well as legal and ethical considerations such as patient consent and ethical approvals. Although no center possessed a complete dataset meeting all clinical aims, nine centers could share neuroradiologists' reports from MRI FLAIR sequences, and seven were prepared to share anonymized imaging data. Currently, images from five European centers have been shared with teams at Padova and Bologna Universities to develop models for detecting silent infarctions, with additional images from other institutions planned for future validation. Three clinical sites, including Oxford and Brussels, have expressed willingness to contribute datasets for the second phase of trials, with collaboration agreements forthcoming.

Furthermore, 23 representatives from 16 centers within the Red Blood Cell subnetwork attended an onsite meeting held on June 13, 2024, during the European Hematology Association annual congress in Madrid. This meeting provided a valuable opportunity to onboard new centers and strengthen collaboration within the network.

**5. Genomed4All Platform Implementation**

A core objective of GenoMed4All is to homogenize and standardize diverse datasets across data types, formats, sources, and coding systems to enable seamless integration into a common data model that serves as the entry point to the platform. The project has adopted the FHIR (Fast Healthcare Interoperability Resources) standard as the foundational data model. This involved analyzing the specific data elements required for model training within each use case and mapping them to appropriate FHIR resources, for example, using the 'patient' resource for demographic data like gender, and the 'observation' resource for clinical measurements such as blood pressure. Clinical data items have been assigned standardized codes through the Athena tool, aligning with OHDSI standards to ensure consistent terminology across the platform.

For genomics data, preprocessing is performed either via GenoMed4All pipelines or by data providers, with mappings tailored to each use case to preserve clinical meaning. For instance, genomics information in MDS can be represented within a single Observation resource, while SCD requires a combination of Molecular Sequence resources for allele identification and Observations for variant details, following established FHIR genomics reporting guidelines. Radiomics data, unlike genomics, is ingested as preprocessed information and encoded using Observation resources.

D5.4 provides a comprehensive overview of the processes involved in data homogenization, detailing the ingestion of source data into the FHIR-aligned common data model according to GenoMed4All implementation guidelines, and describing feature extraction services that prepare clinical and genomics data for machine learning training.

**6. Conclusions**

This deliverable presents a summary of the initial efforts undertaken to coordinate and conduct the first wave of trials focused on developing and validating the GenoMed4All platform components and AI models across the three selected clinical use cases. It identifies key barriers to platform deployment, including legal, ethical, and technical challenges at the clinical site level. A detailed reassessment of the platform design addressing these issues is provided in D4.3.

### Supplementary Information 32| Deliverable 8.1 - Genomics Data Standardization Plan

**1. Recommendations for Genomics data standardization**

This document reviews existing standards and best practices for genomics data management, focusing on guidelines from the Global Alliance for Genomics and Health (GA4GH) and the European Genomics Archive (EGA). It outlines the typical file formats used throughout different stages of genomics data processing, from raw sequencing reads in FASTQ format to aligned sequences stored in SAM/BAM or CRAM files, and ultimately to variant data represented in Variant Call Format (VCF). The recommended standards emphasize the use of SAM/BAM formats for alignment data, highlighting their flexibility, efficiency, and widespread acceptance in next-generation sequencing workflows. The VCF format is described in detail, including its structure, meta-information, genotype representation, and handling of structural variants, all aligned with GA4GH guidelines to ensure interoperability. Additional recommendations address file normalization, metadata inclusion, encryption protocols, and data transfer standards, underscoring the importance of standardized, secure, and well-documented genomics data for effective sharing and analysis.

**2. Phenotypes, Metadata and Data Discovery**

The exchange of human genomics data alongside clinical phenotypes and metadata such as medical history, age, sex, and diagnosis is crucial for studying the relationship between genomic features and their effects. However, these data are sensitive and demand robust privacy and anonymization protocols, a focus of GenoMed4All's WPs 2 and 5. Beyond privacy, the lack of standardized, interoperable data protocols and adherence to FAIR principles remains a significant barrier in the biological data community. To address this, organizations like the Global Alliance for Genomics and Health (GA4GH), European Genome-phenome Archive (EGA), and dbGaP collaborate to promote awareness and implementation of FAIR data sharing practices.
A common data model is essential for the data discovery, the development and deployment of AI algorithms in genomics research, facilitating data interoperability and compatibility across platforms. Establishing a standardized "data model contract" prior to AI model development ensures alignment between datasets and analytical tools, streamlining algorithm training and evaluation.

A significant challenge in human genomics research is enabling federated discovery of identifiable genomic data while maintaining stringent privacy controls. The GA4GH addressed this issue through Beacon, which facilitates secure, federated querying of genomic variants across worldwide datasets. Beacon allows hospitals and research institutions to share their data under controlled access policies without compromising privacy or ownership, enabling different levels of data access depending on the requester's affiliation. The tool is agnostic to data standards and supports querying and exporting data in both formats via its API.

Beacon Version 2 expands capabilities beyond variant identification to include queries involving phenotypes and other clinical data, supporting both predefined studies and user-defined cohort searches. Key features beneficial for GenoMed4All include advanced query filters by attributes such as age or gender, streamlined data permission processes, links to other systems for data access and variant annotations incorporating expert assessments on pathogenicity and phenotype correlations.

### Supplementary Information 33| Deliverable 8.2 - Training Programme Definition

**1. General Overview**

This deliverable, D8.2 is part of the GenoMed4All WP8, which focuses on genomics standardization, value-based healthcare, open science data, and training/education, specifically under Task T8.4 related to the training and education programme. It outlines the initial design and development of the training programme aimed at providing clear concepts and guidance on the technologies used in the project and their clinical outcomes to the Hematological Diseases community, including researchers, healthcare practitioners, patients, relatives, and other stakeholders. The document is structured into three key sections: an executive summary highlighting its main points, a general overview stating the deliverable's purpose, and a detailed description of the training programme. This includes the establishment of the GenoMed4All Educational Scientific/Technical Committee, the methodology for programme development, the five core content areas forming the programme's structure, a preliminary outline of courses and webinars, the communication and dissemination strategy, and the evaluation process for the programme. It is intended as a living document, to be updated as the programme evolves, with the final version and related materials to be presented in the project's concluding report, D8.6.

**2. Genomed4All Training Programme**

The GenoMed4All training programme has been developed in alignment with the ERN-EuroBloodNet webinars strategy to raise awareness and provide education on the various aspects of the GenoMed4All project. The programme's foundation is structured around five key thematic areas, as identified by the GenoMed4All Educational Scientific/Technical Committee (ES/TC), which consists of 17 members representing 13 partner institutions alongside two patient representatives. The committee's collaborative work and regular meetings ensure comprehensive coverage of the project's topics, content accuracy, and overall programme quality. The programme's development was informed by insights gathered during the Patient Engagement e-Workshop, a collaborative event with ERN-EuroBloodNet that involved patient associations across hematological conditions of the Genomed4All three use cases. This workshop evaluated patient communities' awareness of AI applications in these rare diseases and provided an opportunity to discuss their needs and expectations for clinical AI tools. The GenoMed4All training initiative will also align with existing educational programmes from ERN-EuroBloodNet, ENROL, and European research infrastructures on rare diseases, fostering integration across educational resources while minimizing redundancy.

The GenoMed4All online training programme is structured into two editions, with the first launching in 2023 featuring one course aimed at a broad audience covering five key areas, and a second edition in 2024 offering two courses designed to deepen engagement. The initial course seeks to raise awareness by addressing each main topic through focused sessions that include introductions, current state-of-the-art overviews, and challenges within rare hematological diseases. The subsequent courses will elaborate on onboarding, project results, future exploitation, and sustainability, tailored to both general and expert audiences including health professionals, IT specialists, and patient advocates. The programme's five core areas encompass an introduction to GenoMed4All and its European context, an overview of AI applications in biomedicine including multi-omics data integration and advanced clinical decision support systems, secure data science with a focus on data protection under GDPR and tools for ethical data sharing like the Beacon project, FL principles emphasizing equity, autonomy, reliability, and regulatory compliance, and finally, data standardization and interoperability within rare hematological diseases, highlighting the role of the ENROL platform, common data models, and the importance of standardized terminologies and ontologies for consistent data use across European research infrastructures.

The objective of this section is to identify the key challenges in hematological disorders (HDs), particularly rare hematological disorders (RHDs), and demonstrate how AI-driven solutions, specifically through GenoMed4ALL, can address these issues. HDs encompass a broad range of approximately 450 disorders involving abnormalities in blood cells, lymphoid organs, and coagulation factors, many of which are rare but collectively affect a significant patient population. Hematological malignancies, for instance, constitute around 5% of all cancers and often require complex, multidisciplinary management due to their chronic and potentially life-threatening nature.

GenoMed4ALL aims to harness trustworthy and explainable AI technologies to leverage extensive omics data, enhancing research and personalized medicine through three distinct use cases that reflect the genetic heterogeneity and complexity of HDs. Each one of these three use cases presents unique challenges ranging from genomic diversity and mutation interactions to integrating clinical, metabolomic, and imaging data.

In the case of MDS, an acquired oncological disorder marked by somatic mutations, the challenges lie in explaining genomic heterogeneity and improving disease classification, prognosis, and treatment stratification through AI-supported analysis of large genomic and clinical datasets. The project emphasizes the importance of registries, AI-driven biomarker discovery, and personalized medicine approaches.

For SCD, a monogenic congenital disease with a variable clinical spectrum, the main challenge is to understand the determinants influencing clinical outcomes and complications, which remain poorly defined. AI methodologies are proposed to develop predictive models combining clinical, genetic, metabolomic, and imaging data to better classify risk and guide treatment decisions, including early detection of silent infarcts via MRI analysis.

MM, a genetically heterogeneous plasma cell cancer, presents challenges in integrating multi-modal imaging and liquid biopsy data with genomics to improve risk stratification and treatment response prediction. AI will be used to analyze these diverse datasets, characterize disease heterogeneity and evolution, and develop prognostic models that can inform personalized therapy.

Overall, GenoMed4ALL seeks to advance precision medicine in hematological disorders by leveraging AI to integrate complex, multi-dimensional data across these use cases, addressing unmet clinical needs and fostering collaborative research through training and educational initiatives.

The GenoMed4All training program is structured into two phases, with an initial wave in 2023 introducing the project and its core focus areas, followed by a second wave in 2024 that will provide onboarding courses tailored for both general and expert audiences. This program is modeled after the ERN-EuroBloodNet's established Topic on Focus educational series, which has successfully delivered accredited training for health professionals, patients, and patient organizations, enhancing awareness and education around rare hematological disorders. The webinars will be conducted via an interactive Learning Management Platform, such as Zoom, featuring a 60-minute format with a lecture followed by a Q&A session facilitated through the Slido platform to ensure dynamic engagement.

Communication and dissemination are central to the program's success, with a robust strategy designed to maximize reach and participation. The training program will align with GenoMed4All's visual identity across all materials to maintain consistent branding. The ERN-EuroBloodNet website will serve as the main hub for registration, content distribution, and participant feedback, supported by reciprocal links on the GenoMed4All website to drive traffic and maximize outreach. The program targets diverse stakeholder groups by offering content adapted to varying levels of expertise, ensuring relevance and accessibility.

Promotion will be led by AUSTRALO and ERN-EuroBloodNet, leveraging their established social media channels, newsletters, and institutional networks to engage a broad audience. Detailed communications will include webinar information and speaker profiles, with registrants receiving connection details and preparatory materials in advance. The dissemination plan includes making recorded webinars publicly available on YouTube channels for both GenoMed4All and ERN-EuroBloodNet, creating a repository of session slides and videos on dedicated website sections, and producing short “training pills” extracted from webinar recordings to sustain audience engagement. Additionally, a training catalogue based on speaker abstracts will be provided to participants, reinforcing the educational impact of the program.

The evaluation of the GenoMed4All training program will be based on a structured performance review, employing Key Performance Indicators (KPIs) to measure its effectiveness. The program intends to adopt the Kirkpatrick Model for comprehensive assessment. Key metrics include tracking the number of attendees by country through a registration system, aiming for a minimum of 40 participants per session. Attendee satisfaction will be assessed using rating questionnaires delivered via the ERN-EuroBloodNet education platform, with a target average rating of at least 5. Additionally, the reach of the program’s content will be monitored by the number of views on YouTube, with a goal of achieving a minimum of 50 views per video. These KPIs will provide a clear framework to gauge engagement, satisfaction, and dissemination success.

**3. Conclusions**

This deliverable represents the initial phase in the development of the GenoMed4All training program, concentrating on defining the core topics and methodologies. The subsequent phase, which will focus on the program’s development outcomes, will be detailed in the final report at the conclusion of the project. The progress achieved thus far results from the collaborative efforts of VHIR as the lead, ERN-EuroBloodNet, AUSTRALO, and the GenoMed4All ES/TC, with the establishment of a dedicated committee proving essential for guiding this phase. Five primary topics, aligned with GenoMed4All’s core areas, have been identified to form the foundation of the webinar lectures. The training program will comprise a series of webinars conducted across 2023 and 2024, tailored to educate and engage audiences with varying levels of expertise on relevant project themes. This comprehensive webinar series will utilize ERN-EuroBloodNet’s established e-learning platform and resources, providing accessible educational materials for health professionals, patients, and patient organizations. Recorded sessions, along with lecture slides, will be edited and published on dedicated sections of both the ERN-EuroBloodNet and GenoMed4All websites, as well as their YouTube channels. Each one-hour webinar will be moderated by ERN-EuroBloodNet personnel who will introduce speakers and facilitate the interactive Q&A sessions. The diverse educational content, including videos, slides, training snippets, and catalogs, will support the program’s outreach and engagement goals. Ultimately, the program’s success will be evaluated against predefined Key Performance Indicators to ensure its effectiveness.

### Supplementary Information 34| Deliverable 8.3 - Standardization Final Report

**1. Executive Summary**

The decreasing cost of genomic sequencing and its growing application in clinical practice are rapidly generating vast amounts of human sequencing data. If properly shared, these datasets could become an invaluable resource for understanding the genetic foundations of disease. However, the diverse and complex nature of genomic information presents significant challenges for data integration and bioinformatic analysis. Standardizing genotype representation requires conceptual clarity and semantic precision to ensure interoperability. The Global Alliance for Genomics and Health (GA4GH), established in 2013, plays a central role in advancing human health by developing genomic data standards, such as the widely used Variant Call Format (VCF).

Within this context, the Genomed4all initiative targets three hematological disorders and aims to develop a platform that supports the creation and validation of artificial intelligence models for personalized medicine. Genomic profiling is a cornerstone of this approach, enabling more precise disease management. Each research group involved in Genomed4all has implemented specific bioinformatic pipelines to process genomic data according to a well-structured workflow, ensuring high-quality and reproducible outputs. These pre-processed datasets serve as inputs for AI model training and prediction.

This report outlines the current landscape of genomic data standardization and examines the interoperability requirements across the various Genomed4all use cases. It highlights the need for harmonized formats to incorporate new data into model training pipelines and to process individual patient data for clinical predictions. The report also addresses key challenges and limitations in genomic standardization and offers general recommendations for improving data integration and usability.

**2. State of the Art**

Next-generation sequencing (NGS) has transformed human genome analysis, and its decreasing cost is driving widespread adoption in clinical genomics. The reuse of genomic datasets generated in healthcare offers immense potential to advance medical research. However, integrating complex and multidimensional data from various sources remains a major challenge in modern bioinformatics. To fully realize the benefits of genomic science, researchers and clinicians must align on common standards for data collection, storage, analysis, and sharing.

A key obstacle to integration lies in the heterogeneity of data formats across healthcare domains. Ensuring genuine biological insights rather than artifacts demands rigorous standardization of procedures, terminologies, data structures, and quality assurance practices. NGS workflows rely on various file types at different stages of analysis. The initial sequencing step produces FASTQ files containing reads and quality scores, which are then aligned to a reference genome and stored in BAM or SAM files. Subsequent analysis yields data on quality, coverage, structural and epigenetic variations, and notably, genomic variants stored in the widely adopted Variant Call Format (VCF). Although VCF was designed with flexibility to support diverse research contexts, this variability can hinder clinical interpretation due to inconsistencies in how variants are described.

Several initiatives support genomic data standardization. Metadata standards such as MIGS-MIMS, MIAME, and MINSEQE aim to ensure consistent contextual information across experiments. Projects like MAQC promote quality metrics and comparability of results across platforms. File format standardization efforts led by organizations like GA4GH include defining core standards such as VCF and Beacon. Data integration initiatives, such as Genome in a Bottle and NIST's efforts, further aim to align genomic procedures with clinical needs.

Genomed4all seeks to create a platform that enables the collaborative development of AI models for the diagnosis and treatment of hematological diseases in the EU. The integration of genomic data with clinical information from electronic health records has the potential to significantly enhance diagnostic precision. To make this possible in real-world clinical practice, standardization is essential. Specifically, it is necessary to structure input data in a way that ensures interoperability across health systems and to define robust quality metrics for model validation.

**3. Genomic Data & Analysis Overview**

Hematological diseases encompass a broad range of disorders affecting the blood and blood-forming organs, including blood cell cancers, rare genetic conditions, anemia, and sickle cell disease. Many of these diseases have a genetic origin, either inherited or acquired, and are associated with chronic, severe, and potentially life-threatening health issues. Genomed4All concentrates on three specific hematological conditions: MDS, SCD, and MM. In these cases, genomic profiling plays a crucial role in enhancing disease classification, refining diagnosis, and informing treatment strategies. The clinical objectives related to these efforts are further detailed in report D7.1.
In MDS, a genetically complex myeloid neoplasm primarily affecting the elderly, the project aims to identify individuals at risk, refine prognostic classifications, and support transplant-related decision-making through

large-scale sequencing initiatives. In SCD, despite a single shared mutation causing the disease, clinical outcomes vary widely; Genomed4All leverages genomic data to stratify patients based on severity and predict phenotypic outcomes. For MM, a plasma cell malignancy marked by chromosomal alterations, the project focuses on integrating genomic, clinical, and imaging data to identify patients at high risk of early relapse.

**4. Genomic Data Standardization**

In GenoMed4All, each use case focuses on addressing critical unmet clinical needs within specific hematological diseases, supported by the collection and analysis of relevant samples as detailed in deliverables D5.2 and D5.4. Research groups have independently developed dedicated bioinformatic pipelines tailored to their respective use cases, all of which must follow a well-defined workflow to ensure the generation of high-quality, reproducible results. These pipelines, fully documented in deliverable D5.2, produce output files that serve as the input for AI model development. File standardization plays a central role in GenoMed4All, enabling the integration of both prospective and retrospective patient data into continuously improving AI models, ensuring compatibility with the platform for individual-level predictions, advancing data FAIRification (particularly interoperability and reusability), and facilitating data sharing. This section outlines the essential aspects of genomic data standardization across use cases.

*Myelodysplastic Syndrome*

The MDS pipeline in GenoMed4All is specifically designed for a custom panel of approximately 50 genes, with 30 of them being critical for the classification of MDS. The output of the pipeline is a standardized CSV file, where each row corresponds to a patient and each set of three columns corresponds to a gene, detailing mutation presence (binary), the number of mutations, and their variant allele frequencies (VAFs). If multiple mutations are detected in a gene, multiple VAFs are listed. For preprocessing, only FASTQ or BAM files aligned to the hg38 reference genome are accepted, VCFs are not supported. The gene list must adhere to HGNC nomenclature, and while not all genes in the panel must be sequenced, any missing genes will be noted as incomplete data. For the AI model to function, this standardized CSV format is required as input. The pipeline uses both public databases and an internal "whitelist" developed by the consortium to annotate genetic variants, analyzing only those with a coverage above 500x and VAF above 1%. Public databases used include 1000Genomes, Exome Sequencing Project, ExAC, GnomAD, and COSMIC v91. Pathogenicity is assessed through a multi-step algorithm that incorporates known oncogenic profiles, COSMIC reporting frequency in hematologic malignancies, and in silico predictive tools such as SIFT, FATHMM, PolyPhen-2, and MutationTaster. The full pipeline, along with the variant classification logic, is detailed in deliverables D5.2 and D5.4. The MDS pipeline is deployed on CINECA Galileo 100 (G100) servers, enabling the automated processing of raw data from both internal and external partners.

*Sickle Cell Disease*

The SCD pipeline in GenoMed4All employs genome-wide association studies (GWAS) to identify disease-associated single nucleotide polymorphisms (SNPs) across the entire genome using a commercial SNP array. The output is a standardized CSV file where the first column contains a unique sample identifier, and subsequent columns correspond to specific SNPs labeled by their rsIDs. Each SNP column holds dosage values ranging from 0 to 2, representing the number of alternate alleles present: 0 for homozygous reference, 1 for heterozygous, and 2 for homozygous alternate. Intermediate values reflect imputed probabilities. For preprocessing, accepted input formats include PLINK or GEN, with VCF also usable after conversion. Input data must originate from whole genome sequencing (WGS), not whole exome sequencing (WES), and imputation panels such as TopMed or 1000Genomes are supported. The AI model requires the standardized CSV output as its input. Variants must be annotated using rsIDs rather than genomic coordinates, with additional pipeline details available in deliverable D5.2.

*Multiple Myeloma*

The MM pipeline in GenoMed4All analyzes copy number alterations (CNAs) using data derived from SNP arrays or ultra-low-pass WGS of newly diagnosed MM patients. The standardized output is a CSV file, where each row corresponds to a patient and includes the sequencing method used (coded as 1 for SNP array or 2 for WGS), as well as binary indicators for amplifications and deletions across the p and q arms of each chromosome. Amplifications and deletions are recorded separately for each arm, with values indicating presence (1) or absence (0) of alteration. For preprocessing, the pipeline accepts input in FASTQ, BAM, or BED formats, generated through WGS but not WES, and it supports both hg19 and hg38 reference genomes. The AI model requires the CSV file as described for input. While the SNP array-based workflow is now considered legacy and no longer maintained, the WGS-based pipeline is active and documented in deliverable D5.2, with the implementation available on GitHub. CNAs are quantified as weighted averages based on segment length, and only alterations affecting at least 50% of cells are considered: amplifications with values ≥2.50 and deletions ≤1.50. The MM pipeline is also deployed on the CINECA Galileo 100 (G100) infrastructure for automated processing of raw data from consortium and external partners.

### 5. Standardization Limitations

Genomic data standardization plays a critical role in ensuring consistency, interoperability, and scalability of genomic analyses, particularly as NGS becomes more integrated into clinical practice. However, widespread adoption of reference standards has been slow due to the complexity of genomic data, the rapid evolution of sequencing technologies, and the diverse needs of research projects. Within GenoMed4All, the three distinct use cases presented varying genomic requirements and analytical approaches, including targeted gene panels, SNP arrays, and WGS. This diversity necessitated support for different input formats such as FASTQ, BAM, PLINK, GEN, and BED, which, while justified by clinical objectives, complicates efforts toward harmonized workflows. A significant challenge in standardization lies in the lack of consensus around oncogenic variant classification. For instance, the MDS pipeline relies on both public databases and an internal registry informed by clinical experience. While this internal database adds valuable domain-specific insights, it limits external reproducibility and interpretability. Furthermore, the pace at which technologies advance introduces a dual burden: integrating new standards while managing the obsolescence of legacy systems. The MM pipeline illustrates this tension, supporting both SNP array and WGS technologies, even as WGS increasingly replaces SNP arrays due to lower costs and broader clinical adoption. This coexistence is especially relevant when incorporating retrospective data. Regulatory constraints also pose practical limitations. For example, although the TOPMed reference panel offers high-quality genotype imputation, data protection regulations within Europe prevented the use of U.S.-based servers at the University of Michigan, restricting access to this resource for GWAS data. Synthetic alternatives were not viable due to privacy concerns, underscoring broader challenges in cross-border genomic collaboration. Additional issues include nomenclature inconsistencies, as seen in the SCD use case, where SNPs are referenced using rsIDs. While convenient, rsIDs do not capture all variants and lack information on specific nucleotide changes, potentially affecting biological interpretation. Altogether, these limitations highlight the complexity of achieving true genomic standardization. Continued progress will depend on cross-disciplinary collaboration, increased awareness of the importance of standards, and investment in training and education to ensure widespread and informed adoption across the research and clinical communities.

### 6. Conclusions

GenoMed4All addressed three distinct genomic use cases, each with specific standardization requirements, and identified several key factors essential for successful data integration and sharing. A clear definition of accepted input file formats is crucial for enabling external data processing. The MDS use case prefers FASTQ or BAM files but can also accept finalized CSV files containing mutation data and variant allele frequencies. SCD accepts PLINK or GEN formats, while MM can process FASTQ, BAM, or BED files. It is important to note that SCD and MM pipelines support data generated from WGS technologies but not from WES. Specifying the reference genome used is critical for ensuring reproducibility. MDS relies on hg38, MM supports both hg19 and hg38 (with hg19 used in the project), and SCD refers to reference panels such as TOPMed and 1000Genomes for imputation.

To support FAIR principles, the project emphasizes the importance of standardized metadata, which provides context for data interpretation and increases dataset findability and reusability. GenoMed4All metadata will be made accessible through the EOSC catalogue. Additionally, adherence to standardized gene nomenclature, such as HGNC guidelines, is vital for consistent referencing, currently applied only in the MDS use case. It is also recommended to reference SNPs using genomic coordinates instead of rsIDs to avoid ambiguity and ensure clarity on nucleotide changes. Finally, the project highlights the importance of depositing genomic data in trusted public repositories like EGA, dbGaP, or JGA, which ensure secure access and long-term availability. Equally important is the open sharing of bioinformatic pipelines and workflows through platforms such as GitHub, bio.tools, SciCrunch.org, or WorkflowHub to foster reproducibility and transparency across the research community.

### Supplementary Information 35| Deliverable 8.4 - Genomics for Value-based Healthcare

#### 1. Introduction

Rising healthcare costs represent a major challenge for European healthcare systems, largely driven by expenses related to disease treatment. Value-Based Healthcare (VBHC) offers a promising approach by focusing on maximizing patient outcomes relative to costs. Central to delivering high-value care is a comprehensive understanding of each patient's individual needs, where genomics plays a crucial role. By generating extensive patient data and uncovering genetic risk factors, genomics enables the customization of diagnostics and therapies, improving patient outcomes while potentially reducing unnecessary procedures and associated costs. This approach is particularly pertinent to hematological diseases, which impose an estimated annual cost of €23 billion across Europe. Despite significant genetic classification within these diseases, many patients remain undiagnosed or misdiagnosed, with conditions like MDS frequently subject to initial misclassification. The diagnostic journey for rare diseases is often prolonged and complex, leading to substantial additional costs from delayed diagnosis. Genomic profiling, enhanced by technologies such as next-generation sequencing, has advanced understanding of disease mechanisms and supported the development of prognostic models, allowing for more precise diagnosis and treatment selection. Given the genetic diversity among hematological disease patients, care must be highly individualized, emphasizing accurate disease characterization, refined risk stratification, and personalized therapeutic strategies that deliver meaningful value to both patients and healthcare systems.

#### 2. Artificial Intelligence and value-based healthcare

The evolving healthcare landscape necessitates a shift towards VBHC, which prioritizes enhanced patient outcomes alongside efficient resource use. Achieving this requires balancing the needs of multiple stakeholders and analyzing vast amounts of data, where AI plays a pivotal role, especially in the management of hematological diseases (HRDs). Challenges in this field include geographically dispersed patient populations and fragmented data, which can be overcome by securely pooling resources from multiple institutions. AI's ability to analyze large datasets facilitates the identification of hidden patterns, enabling early disease detection, personalized treatment plans, and proactive care. NGS generates extensive genomic data, but interpreting this volume exceeds the capacity of individual clinicians, making AI essential for refining disease understanding, risk stratification, and treatment personalization. Integrating data from genomics, transcriptomics, proteomics, and metabolomics offers a comprehensive view of patient biology, with metabolomics providing real-time insights into physiological states. Such complex, multi-layered data necessitates AI to develop predictive models that are accessible to healthcare providers and patients alike. AI excels at analyzing individual patient data to forecast risks, recommend tailored therapies, and monitor treatment responses, thereby improving patient experience and healthcare value. Its predictive capabilities also support proactive management, resource optimization, and a shift from reactive to preventive care-core VBHC objectives. Early AI-enabled detection in HRDs can reduce the significant clinical, economic, and temporal burdens on patients and healthcare systems. Diseases such as MM, MDS and SCD require advanced diagnostics and complex treatments, often involving multiple specialists, which can delay timely care. Leveraging genomics and AI within personalized medicine empowers European healthcare systems to transition to value-driven models, enhancing patient outcomes while optimizing costs. Beyond resource efficiency, this approach can improve patient satisfaction through streamlined diagnostic and treatment pathways, which are linked to better adherence and improved clinical outcomes.

*Myelodysplastic Syndrome*

MDS is a complex group of hematological malignancies marked by defective stem cells that impair normal blood cell formation in the bone marrow. Notably, around 40% of high-risk MDS patients progress to AML, a severe condition with increased mortality and more intensive treatment requirements, underscoring the critical need for early identification of those at risk of transformation. Emerging research indicates that combining AI with genomic sequencing significantly enhances the accuracy and efficiency of MDS diagnosis, patient classification, and prognosis. Given the importance of early detection in MDS, AI and ML offer substantial potential to improve outcomes through timely interventions that may prevent progression to AML. By rapidly analyzing extensive datasets, AI algorithms can uncover subtle patterns and correlations beyond human capability, facilitating the identification of high-risk patients and enabling targeted preventive strategies. These advancements in early diagnosis, risk stratification, and personalized care form a strong foundation for implementing VBH approaches for MDS patients in the near future.

*Multiple Myeloma*

MM is a hematological malignancy characterized by malignant plasma cells in the bone marrow, causing multiple tumor sites and complications such as bone damage and infections. Despite advancements improving survival, early morbidity from treatment and high mortality in high-risk patients remain significant challenges. The disease's clinical course is highly heterogeneous due to underlying molecular variations, with over 25% of patients presenting high-risk factors at diagnosis. Applying AI to genomic data can enhance early risk stratification, enabling personalized treatment strategies that target specific lesions, thereby improving efficacy and minimizing unnecessary toxicity. Although novel therapies have advanced MM treatment, relapse remains common due to variable patient responses driven by complex alterations within cancer cells and their microenvironment, including gene expressions related to chemoresistance. AI algorithms can improve predictions of individual treatment responses, potentially reducing relapse rates and guiding optimal treatment sequencing amid numerous drug options. DL approaches, already used in tumor detection and drug response prediction in other cancers, hold promise for improving MM management. These AI-driven strategies can significantly enhance patient outcomes and healthcare efficiency by facilitating better treatment selection, optimizing therapeutic pathways, reducing toxicity, and promoting faster recovery with lower resource utilization.

*Sickle Cell Disease*

SCD is a globally prevalent inherited blood disorder caused by a mutation in the hemoglobin gene, resulting in sickled red blood cells that obstruct blood flow and cause severe complications, including recurrent vaso-occlusive crises, acute chest syndrome, stroke risk, and progressive organ damage. Although SCD originates from a single genetic mutation, significant variability exists in patient symptoms and disease progression, likely influenced by mutations and polymorphisms in additional genes. These genetic differences affect individual responses to standard treatments, often resulting in suboptimal therapeutic outcomes. AI is increasingly applied to improve SCD diagnosis, predict disease severity, stratify risk, and enhance complication detection, aiming to accelerate appropriate interventions and optimize resource allocation for high-risk patients. NGS offers a powerful tool to screen multiple gene loci simultaneously, making SCD an ideal candidate for integrating NGS with personalized medicine approaches to advance value-based healthcare and deliver more tailored, effective treatments.

**3. Impact of AI on VBHC outcomes: Clinicians' perspective**

An online survey was conducted to capture clinicians' perspectives on the role of AI in implementing VBHC for haematological rare diseases, targeting both Genomed4All project participants and external experts to mitigate potential bias. The survey, conducted in multiple phases between November 2023 and May 2024, engaged clinicians working on MDS, MM, and SCD. Results from 19 Genomed4All clinicians demonstrated a broadly positive attitude towards AI, with respondents acknowledging AI's potential to enhance clinical decision-making by integrating diverse data sources for more accurate diagnoses and personalized treatments. In particular, AI was seen as a tool to improve early diagnosis, risk stratification, complication prediction, treatment response forecasting, and patient prognosis across all three diseases. SCD clinicians noted AI's capacity to increase patient involvement and optimize screening schedules, while MDS professionals highlighted its utility in refining ambiguous diagnoses. The survey also identified the need to improve AI implementation, including the analysis of larger, diverse datasets, harmonization and accessibility of clinical data, and educational resources to build clinician confidence in AI tools.

A subsequent cross-validation survey of ten external clinicians echoed these positive views, albeit with slightly more caution, especially among MDS specialists. External respondents confirmed AI's value in integrating complex data and enhancing diagnostic accuracy, though some impacts reported by Genomed4All participants, such as increasing patient involvement and rescheduling screenings, were not noted by the external group. Both groups agreed that AI could support treatment personalization and risk stratification, with external clinicians emphasizing the importance of incorporating new prognostic markers into international guidelines, strict regulation of data access, expert support for AI interpretation, and the necessity of large-scale real-world trials. Overall, while minor bias towards AI enthusiasm was observed among Genomed4All clinicians due to their project involvement, the alignment between internal and external perspectives supports the broader applicability of AI models post-project. Both groups underscored the importance of comprehensive education and support to facilitate AI adoption in clinical practice and highlighted the need to prioritize external clinicians' input moving forward, given their majority representation in the field.

**4. VBHC model framework**

The core principle of VBHC focuses on enhancing health outcomes while optimizing healthcare resource use. This framework proposes a theoretical health economic model designed to capture VBHC outcomes and cost indicators relevant to hematologic rare diseases from the perspective of European healthcare systems, encompassing both direct and societal burdens. The model centers on the patient care pathway, illustrating the impact of introducing AI as patients progress through diagnostic and treatment stages. Developed following the European Institute of Innovation and Technology's VBHC handbook and standards set by the International Society for Pharmacoeconomic and Outcomes Research, the model emphasizes AI's effect on diagnosis, classification, and management of three HRDs compared to current standard care.

The patient journey begins with symptom reporting and diagnostic testing, progressing through diagnosis confirmation, treatment planning, and subsequent treatment phases. Outcomes vary from disease remission or progression to treatment failure, with transitions influenced by survival rates and treatment efficacy. Mortality risk is accounted for at all stages. To accurately model patient transitions over time, a semi-Markov approach is recommended, capturing changing probabilities influenced by patient age, treatment type, and diagnostic accuracy.

The model simulates a hypothetical cohort with average patient characteristics, which can be adjusted as needed. Diagnostic results may be true or false positives/negatives, influencing subsequent care decisions. Treatment cycles continue until success, death, or time horizon completion, with adjustments made based on response or adverse events.

To address uncertainty, probabilistic and one-way sensitivity analyses are applied, varying input parameters across distributions to assess the robustness of outcomes. Key inputs include AI efficacy metrics, diagnostic and clinical parameters, clinical outcome assessments such as quality of life measures, physician and patient satisfaction scores, healthcare resource utilization, and associated costs.

Model outputs reflect the total costs required to achieve relevant outcomes, presenting value as a ratio of health outcomes to costs or visualized on a multidimensional value plane that incorporates clinical, patient-centered, and cost measures. This approach acknowledges the complexity and variability of stakeholder perspectives, emphasizing a comprehensive assessment of value in healthcare delivery.

#### 5. VBHC model adaptation for MDS

The adapted VBHC model for high-risk MDS integrates both diagnostic and treatment pathways. The diagnostic process typically begins with patients presenting symptoms such as anemia-related fatigue, bleeding, or frequent infections. Initial assessment includes a complete blood count (CBC), with normal results usually ruling out MDS and prompting further evaluation for other conditions. Abnormal CBC findings lead to a bone marrow aspirate to identify dysplastic cells and increased blasts, alongside cytogenetic analysis and molecular testing using NGS to detect genetic mutations linked to MDS. However, 10-20% of cases may lack identifiable mutations, reflecting the disease's multifactorial nature. The incorporation of AI enhances this pathway by improving diagnostic precision through advanced interpretation of genomic data, clinical inputs, and bone marrow image analysis, facilitating earlier and more accurate risk profiling and treatment selection.

Treatment for high-risk MDS follows a hierarchical approach informed by the International Prognostic Scoring System (IPSS) and its refinements, which classify disease severity based on clinical, cytogenetic, and genetic factors. First-line therapy typically involves hypomethylating agents such as azacitidine or decitabine, followed by second-line treatments like lenalidomide for non-del(5q) patients, which has a limited response rate. HSCT offers a potential cure for younger, fit patients but carries significant risks, including transplant-related mortality and relapse. When curative options are not viable, palliative care focuses on symptom management and quality of life, supported by blood transfusions, antibiotics, and growth factors, with experimental treatments available through clinical trials.

The VBHC model emphasizes improved patient outcomes through enhanced diagnostic accuracy, reduced time to diagnosis and treatment, extended life expectancy, longer remission durations, and better quality of life. Additionally, the model supports cost-effectiveness by optimizing resource use through fewer diagnostic repetitions and more targeted treatments.

### Supplementary Information 36| Deliverable 8.5 - Open Science data collaboration in Genomed4All plan and report

#### 1. European Open Science

The European Open Science Cloud (EOSC) is an initiative designed to establish a federated computational ecosystem that supports the sharing, discovery, and reuse of research data, tools, and services across disciplines within the European Union. EOSC aims to ensure seamless access to resources while adhering to principles of trust, transparency, and public interest protection. Central to its mission is the implementation of FAIR principles, ensuring data and associated research outputs are findable, accessible, interoperable, and reusable, thereby creating a robust Web of FAIR Data and services that enables diverse value-added scientific activities. This approach has evolved from a centralized portal to a federated network of EOSC Nodes, enhancing accessibility and collaboration.

The FAIR principles underpin EOSC's data management strategy, focusing heavily on metadata to improve machine-readability and automate data handling processes. Data and metadata are assigned unique, persistent identifiers, enriched with detailed descriptions, and registered in searchable repositories, ensuring discoverability even if the original data become unavailable. Accessibility is maintained through trusted repositories such as Zenodo, which support standardized protocols for data retrieval and clearly defined access conditions. Interoperability is achieved by adopting international standards and controlled vocabularies, like SNOMED CT and LOINC, along with data models such as FHIR, enabling seamless data exchange across systems. Reusability is supported by clear licensing, thorough provenance documentation, and rich metadata annotations, facilitating responsible and informed data reuse.

The transition to the EOSC Node model represents a distributed, federated infrastructure that provides researchers with easy access to a wide range of open science resources while promoting collaboration and data management aligned with FAIR principles. This network eliminates the need for centralized user certification, allowing interoperability among autonomous nodes that share content through common standards and policies. Users can select from a variety of community-endorsed repositories, with Zenodo serving as a primary platform for depositing research data, software, and preprints, ensuring broad dissemination and long-term preservation.

Zenodo, developed by CERN in partnership with OpenAIRE, functions as an open-access repository designed to support the principles of Open Science. It offers infrastructure and tools for managing large-scale data and making research outputs widely accessible, especially for projects funded by the European Commission.

The integration of the GENOMED4ALL project within the EOSC framework exemplifies the benefits of this ecosystem, enabling the extensive sharing and valorization of biomedical data generated during the project. This collaboration not only preserves and extends the impact of GENOMED4ALL's datasets but also encourages future partnerships and joint research initiatives. By facilitating open access to high-quality, peer-reviewed information, the synergy between EOSC and GENOMED4ALL accelerates scientific discovery, fosters transparency, and supports a collaborative research environment aimed at transformative advancements in biomedicine and beyond.

**2. Genomed4All Partner's Role**

The role of T8.3 partners encompasses several key responsibilities that have evolved throughout the project as the requirements for data and metadata sharing became clearer. Initially, CINECA developed a robust local infrastructure to securely host, process, and manage project data. Partners coordinated internally across WPs and institutions to determine which data should be shared among project members and which could be made publicly accessible, informed by a questionnaire distributed to use case leaders. Given the sensitivity of genomic, clinical, and demographic data, which cannot be openly shared, the consortium agreed to publicly release only metadata for the three use cases on the EOSC Node platform, while full dataset access would be granted on a controlled, case-by-case basis through a formal request process.

Metadata was standardized and prepared collaboratively by partners to align with the EOSC guidelines and FAIR data principles, then uploaded to the Zenodo repository for open discoverability.

CINECA's infrastructure adheres to EOSC and FAIR standards, offering a secure, scalable, and sustainable environment capable of processing raw data, anonymizing sensitive information, and extracting metadata. The infrastructure emphasizes security and compliance, including ISO 27001 certification and GDPR adherence, with high-performance computing resources integrated into a cloud platform (ADA Cloud) built on OpenStack. This environment supports user access through virtual machines equipped with popular analytical tools, accompanied by documentation and example scripts to facilitate ease of use.

The data sharing strategy began with a comprehensive assessment of data sources through questionnaires, revealing that most data are sensitive and unsuitable for public sharing. Consequently, metadata was selected for open sharing to provide transparency and insight without compromising privacy, while full datasets remain accessible only through controlled procedures. This approach balances openness with ethical and legal responsibilities.

Publicly shared metadata focuses on three disease cohorts and includes aggregated data where appropriate to maintain confidentiality. Metadata and aggregated information are hosted on Zenodo and indexed via the EOSC EU Node. Researchers seeking access to complete datasets must submit requests through a cohort-specific form, with approvals managed by designated use case referents to ensure compliance with GDPR and alignment with the consortium's research objectives. This structured framework supports FAIR principles, promotes responsible data sharing, and facilitates impactful collaborative research while safeguarding participant privacy.

**3. Metadata made available on the EOSC EU Node**

This section presents key extracts from the metadata sources of databases identified for sharing, focusing on the project's three primary diseases: MDS,MM and SCD.

MDS are a group of bone marrow failure disorders primarily affecting the elderly, characterized by insufficient production of healthy blood cells and potential progression to AML. The metadata relates to an international retrospective cohort of 2,043 patients diagnosed according to the 2016 WHO criteria. The dataset includes comprehensive genomic and transcriptomic information, such as targeted mutation screening of 47 genes linked to clonal haematopoiesis, whole-exome and whole-genome sequencing data, and clinical parameters including blood counts, cytogenetic abnormalities, treatment details, and patient outcomes. This rich dataset supports an understanding of disease mechanisms, progression, and therapeutic resistance.

MM is a bone marrow cancer involving abnormal plasma cells that disrupt normal blood cell production and cause organ damage. Metadata was collected from two distinct cohorts totaling over 250 newly diagnosed patients, complemented by publicly available data from 750 patients in the CoMMpass database. Data includes baseline clinical characteristics, imaging studies, tumor fraction measurements from liquid biopsies, chromosomal copy number alterations, treatment regimens, and outcomes. These datasets facilitate multimodal disease characterization, evaluation of treatment response, and study of genomic instability patterns associated with progression and relapse.

SCD is a hereditary disorder characterized by misshapen red blood cells that impair blood flow and cause chronic complications. The metadata comprises 1,000 standardized samples collected under the Rare Anemia Disorders European Epidemiological Platform, ensuring compliance with EU guidelines and data quality standards. It includes pseudonymized patient demographics, clinical histories, laboratory and imaging results, genetic diagnostics, treatment records, and involvement in clinical trials. The dataset supports investigation into genetic and clinical factors influencing disease severity, employing genomic analyses, AI-based risk stratification models, and radiomic assessment of brain MRI scans to predict complications such as silent infarctions.

All metadata have been uploaded to the Zenodo portal and indexed through the EOSC EU Node distributed architecture, selected for its alignment with open science principles and FAIR data standards. This platform enhances the visibility and accessibility of the consortium's data within the European research community, maximizing its scientific impact. The GenoMed4All project community on Zenodo serves as a central hub for accessing these datasets.

**4. Final Remarks**

This deliverable outlines significant progress made in advancing the EOSC initiative. The necessary infrastructure was successfully established to support the execution of project tasks, including technical environments, tools, and workflows for effective data management, monitoring, and analysis. A comprehensive understanding of the evolving EOSC ecosystem was achieved, adapting to the transition from the EOSC Portal to the new EOSC EU Node distributed strategy, which enabled the successful uploading of relevant metadata on the Zenodo portal. Key datasets of high relevance to the scientific community were identified and made publicly available within the EOSC framework, accompanied by a formal procedure for requesting full access to the complete datasets. A tailored data life cycle framework was developed to guide data management from collection through to storage, sharing, and archiving, aligned with EOSC principles to ensure data stewardship, longevity, and compliance. All GenoMed4All materials have been uploaded and are accessible via Zenodo, integrated within the GenoMed4All H2020 project community, and the project's commitment to open science is prominently featured on its website. These accomplishments represent critical advancements in enhancing

EOSC's capacity to support open science and foster collaborative research, with plans to continue improving systems, expanding datasets, and refining infrastructure to solidify EOSC's role as a leading platform for global scientific collaboration.

### Supplementary Information 37| Deliverable 8.6 - Final Report on the Genomed4All Training

#### 1. Introduction

This deliverable builds on the foundational work established in the previous report, D8.2, GenoMed4All Training Program Definition, which set the strategic framework and educational objectives for the GenoMed4All training initiative. Developed collaboratively by the Educational Scientific/Technical Committee, the earlier deliverable defined key components including the program's objectives to raise awareness and provide education on GenoMed4All's core themes, the identification of five main topic areas covering both foundational concepts and advanced applications related to rare hematological diseases, and the adoption of webinars as the primary training format. It also emphasized content customization to address the needs of a broad audience, ranging from the general public to researchers, healthcare professionals, AI experts, and patient advocates, supported by a comprehensive communication strategy involving key partners to maximize outreach and assess effectiveness. The five pedagogical areas focus on an introduction to GenoMed4All, secure data science, FL and regulatory issues, data standardization in rare hematological diseases, and practical use cases including SCD, MM, and MDS. The training program's phased implementation began with a 2021 edition aimed at patient organizations through an introductory e-workshop shaped by stakeholder input, highlighting the importance of AI in disease management. The subsequent edition, planned for 2023-2024, includes two tailored courses for both general and expert audiences. This deliverable advances the training program by building on these established structures and insights to further develop and refine educational efforts that address the diverse needs of its audience.

#### 2. Methods for Educational Programmes' Development

The design and implementation of the GenoMed4All and ERN-EuroBloodNet Educational Programs were guided by a collaborative, iterative methodology consistent with the objectives of WP 8, Task T8.4. Development was driven by the GenoMed4All Educational Scientific/Technical Committee, composed of experts in hematology, artificial intelligence, education, and key stakeholders including researchers, healthcare professionals, and patient advocates. The pedagogical strategy focused on two main targets: a public-oriented program aimed at foundational topics to raise general awareness through accessible, interactive formats, and an expert-focused program delivering specialized content tailored for health professionals, researchers, and AI developers, emphasizing advanced methodologies, regulatory issues, and clinical AI integration. The program's design aligned closely with GenoMed4All's thematic priorities and addressed the varying expertise levels of its audiences. Methodological consensus was achieved through a series of semi-structured meetings incorporating consensus-building, brainstorming, and iterative feedback, ensuring inclusivity and comprehensive content. Speaker selection followed a consensus-driven process, choosing internationally recognized experts in precision medicine, AI, and rare hematological diseases, with an emphasis on effective communication and alignment with program goals. The programs were delivered via Zoom to facilitate real-time engagement and global participation, with sessions recorded and made available on GenoMed4All and ERN-EuroBloodNet e-learning platforms and YouTube channels to ensure sustainability and wider accessibility. This structured, participatory approach ensured scientific rigor while maintaining accessibility, supporting the diverse needs of the audience and the long-term objectives of GenoMed4All.

#### 3. Genomed4All & ERN-Eurobloodnet Educational Program for the public at large

The GenoMed4All & ERN-EuroBloodNet Educational Program represents a pioneering initiative aimed at deepening the understanding and raising awareness of artificial intelligence (AI) applications in hematology, with a strong focus on personalized medicine. Developed by the GenoMed4All Educational Scientific/Technical

Committee, the program was tailored to healthcare professionals, researchers, and technical experts, aligning with GenoMed4All's broader mission to integrate AI-driven solutions into the diagnosis, treatment, and management of rare hematological diseases.

The educational program unfolded in two series, delivered in 2023 and 2024 via Zoom, and featured leading experts in AI, precision medicine, and multi-omics. The sessions were designed to be highly interactive, offering live discussions and Q&A forums to promote knowledge exchange and practical engagement. Recordings were made available on both the GenoMed4All and ERN-EuroBloodNet YouTube channels, ensuring continued access to the material.

The program's core objectives included raising awareness of GenoMed4All's contributions to AI-based precision medicine, fostering cross-border collaboration, and standardizing AI tools and data for improved patient care. By addressing both introductory and advanced themes, it successfully bridged the gap between research and clinical practice, encouraged public engagement with emerging AI technologies, and facilitated collaboration across professional and patient communities.

Each webinar focused on a specific aspect of AI in hematology, covering GenoMed4All's mission, disease-specific AI applications, data standardization, and multi-modal data analysis. These sessions emphasized the transformative potential of integrating AI with genomics, proteomics, and radiomics to enhance diagnostic accuracy and treatment personalization.

With 190 participants, the program attracted a diverse and interdisciplinary audience, including hematologists, computational engineers, bioinformaticians, laboratory specialists, and representatives from patient advocacy and healthcare organizations. Most attendees fell within the 30–50 age range, and Italy, Spain, and Cyprus were among the most represented countries.

A post-event survey, completed by 39 participants, reflected high levels of satisfaction. Respondents rated the webinars positively for content relevance, applicability to professional practice, and overall educational value. The average ratings ranged from 4.5 to 4.7 out of 5, underscoring the program's effectiveness in meeting educational goals and generating strong interest in continued learning.

Overall, the program not only advanced GenoMed4All's educational strategy but also significantly contributed to building a collaborative, knowledgeable community ready to embrace AI innovations in hematology.

**4. Genomed4All & ERN-Eurobloodnet Educational Program for Expert Audiences**

The second edition of the GenoMed4All & ERN-EuroBloodNet Educational Program for expert audiences was designed to deepen specialist knowledge in AI applications for rare hematological diseases, with a strong emphasis on multi-omics approaches and the integration of high-quality data within regulatory frameworks. Its objectives centered on advancing understanding of AI's latest innovations in hematology, addressing the complexities of data standardization and model bias, and fostering critical discussions on compliance and data governance.

This program further solidified GenoMed4All and ERN-EuroBloodNet's commitment to leveraging AI to transform the diagnosis, management, and research of rare blood disorders. It provided in-depth technical insight into real-world AI use cases for MDS, SCD and MM. By bringing together clinicians, researchers, and data scientists, the program fostered a multidisciplinary environment conducive to collaboration and the development of clinically relevant AI tools.

Structured as a series of six webinars conducted in late 2024, the program explored themes ranging from foundational overviews to technical case studies. Topics included the evolution of AI in hematology, data reliability and fairness, regulatory requirements, and disease-specific onboarding protocols. These sessions, delivered by leading experts, highlighted the role of data quality and integration in enhancing AI accuracy and clinical utility. Recordings remain accessible on the ERN-EuroBloodNet EDU and GenoMed4All YouTube channels.

The program engaged 111 participants, representing a diverse professional landscape. Hematologists were the most represented group, followed by laboratory specialists, computational engineers, students, and a range of other roles spanning bioinformatics, project management, clinical research, and patient advocacy. Geographically, the audience was primarily drawn from Portugal, Cyprus, and Italy, with broader participation from various countries across Europe, the Middle East, and North Africa. Age demographics were concentrated in the 30 to 50 range, reflecting the program's appeal to mid-career professionals.

Feedback from participants was overwhelmingly positive. Out of 35 responses, the webinar series received an average rating of 4.7 for both knowledge enhancement and applicability in professional settings, and a 4.5 rating for likelihood to recommend. The same high score of 4.7 was recorded for interest in exploring related topics further, indicating sustained engagement and educational impact.

In summary, this second installment of the educational program significantly advanced AI literacy within the hematology community. It supported the broader GenoMed4All mission by equipping professionals with the tools, frameworks, and interdisciplinary knowledge necessary to integrate AI responsibly and effectively in the care of patients with rare hematological conditions.

**5. Dissemination Plan**

The dissemination strategy for the GenoMed4All & ERN-EuroBloodNet joint educational program was designed to ensure maximum visibility, accessibility, and alignment with the overarching identity of the GenoMed4All initiative. A cohesive visual identity was developed to clearly reflect the program's association with GenoMed4All, using established branding elements such as logos, typography, and color schemes. These visual standards were consistently applied across all materials (from presentation templates to promotional graphics and video thumbnails) reinforcing the collaborative nature of the program with ERN-EuroBloodNet, which also contributed visuals in its own distinctive style for dissemination purposes.

The ERN-EuroBloodNet website served as the central platform for webinar registration, participant interaction, and content distribution. With over 360,000 page visits, it provided a robust infrastructure to manage registrations, post-event surveys, and public access to educational materials. A dedicated section was created for the training program, which attracted more than 2,300 visits. In parallel, GenoMed4All hosted a comprehensive training hub on its own website, offering landing pages for both waves of the program, webinar recordings, detailed session descriptions, and information on the educational committee overseeing the content.

Audience targeting played a crucial role in shaping the dissemination approach. The program was designed to address both general and expert audiences, accommodating varying levels of familiarity with the subject matter. This inclusivity ensured broader stakeholder engagement and aligned with the project's overarching impact strategy.

Promotion of the webinars was actively supported through digital communication channels. Social media platforms across both ERN-EuroBloodNet and GenoMed4All were utilized to share updates, event announcements, and visual content. Metrics from these platforms ,such as over 1,400 followers on LinkedIn and more than 1,500 on X (formerly Twitter), indicate a solid outreach capability. Additionally, regular newsletters, including education-specific issues in July and September, served to inform and engage stakeholders directly.

Crucially, the program's educational materials have been made freely accessible. All recorded webinars are publicly available on both organizations' YouTube channels, accompanied by downloadable PDF slide decks. This open-access model ensures that knowledge dissemination extends well beyond the live events, supporting continuous professional development and the wider adoption of AI in hematology.

Overall, the dissemination plan has effectively supported the visibility, reach, and impact of the educational program, fostering a strong digital presence and promoting enduring access to high-quality, specialized knowledge across the hematology and AI communities.

**6. Conclusion**

The GenoMed4All & ERN-EuroBloodNet Educational Program, developed through collaborative input from GenoMed4All's Educational and Scientific/Technical Committee, marks a significant milestone in advancing the understanding and application of artificial intelligence in hematology. Built on a foundation of structured dialogue and consensus, the program delivered a comprehensive, dual-track educational initiative tailored to both the general public and expert audiences.

For the public, the program succeeded in raising awareness through accessible and engaging webinar content that introduced the fundamentals of precision medicine and the transformative role of AI in clinical research. It addressed key issues such as data standardization and multi-omics integration in a format designed to inform and inspire a broader audience.

In parallel, the expert-focused sessions offered a deep dive into advanced AI methodologies, including data federation and multi-modal applications specific to hematology. Featuring insights from leading specialists, this component provided a high-level exchange of knowledge aimed at enhancing the integration of AI into personalized medicine and clinical practice.

The program's reach was significantly extended through interactive elements such as live Q&A sessions, and by making recorded content publicly available on GenoMed4All and ERN-EuroBloodNet's e-learning platforms. This ensured that educational value would persist beyond the live events, offering ongoing access to high-quality content for a global audience.

Crucially, the entire initiative was guided by the strategic goals of GenoMed4All, ensuring all content aligned with the overarching mission to embed AI in hematological research and clinical application. By fostering knowledge dissemination, professional development, and interdisciplinary collaboration, the program not only delivered immediate educational benefits but also established a strong foundation for future innovation.

With over 360,000 online visits and more than 200 participants, the program has demonstrated strong engagement and impact. It stands as a powerful example of how targeted educational efforts can accelerate the adoption of AI in medicine, enhance patient care, and drive the future of personalized hematology through sustained learning and collaboration.

### Supplementary Information 38| Deliverable 9.1 - Impact Master Plan

**1. Introduction**

This deliverable, D9.1 – *GenoMed4All Impact Master Plan*, has been developed within the framework of WP9 on Impact, Outreach, and Collaboration, specifically under task T9.1 dedicated to dissemination and communication planning. Its overarching purpose is to establish a foundational strategy to accelerate and enhance the transfer of knowledge from the GenoMed4All project to its relevant stakeholders, thereby maximizing the project's value and reach.

The document defines the initial structure of a multi-faceted approach to stakeholder engagement, public outreach, and exploitation of results. It sets out a collaboration framework to build synergies across target groups, proposes communication strategies aimed at raising awareness, and presents a plan to evaluate the project's visibility and outcomes through defined performance indicators. In addition, it addresses frameworks for intellectual property management, technology transfer, and participation in standardization efforts at both European and international levels.

As the first in a series of deliverables under WP9, D9.1 serves as a strategic roadmap for future outputs, including interim and final reports on dissemination (D9.2 and D9.4) and exploitation (D9.3 and D9.5). Notably, this document is intended as a living strategy, adaptable to evolving market dynamics and emerging standardization opportunities, ensuring GenoMed4All remains responsive and aligned with broader innovation and policy landscapes over the course of the project.

The structure of this document includes an introductory section outlining its purpose, followed by a presentation of the stakeholder collaboration framework, a detailed overview of the communication and dissemination strategy, a discussion on the exploitation of key results, and concluding remarks summarizing the strategic vision at this stage of the project.

**2. Stakeholder Collaboration Framework**

As part of its mission under the H2020 framework, GenoMed4All is committed to building a dynamic and responsive ecosystem of stakeholders to maximise the impact of its work. While inherently decentralised, the project recognises the critical importance of fostering structured collaboration with a diverse network of organisations and initiatives that influence or are influenced by its objectives. To achieve this, GenoMed4All is deploying an Agile Stakeholder Engagement Framework, a methodology designed to ensure continuous interaction and feedback through iterative 6-month sprints across three phases: scouting, interacting, and learning. This approach enables the project to progressively identify, engage, and adapt to the evolving landscape of stakeholders throughout its lifecycle.

The initial scouting phase focuses on mapping and classifying target groups based on their relevance and influence, forming a stakeholders map that guides strategic outreach. In the interaction phase, GenoMed4All builds active relationships with these groups through participation in working groups, publications, events, and direct collaboration. Finally, the learning phase involves assessing these engagements through consultations and integrating the insights gained into future sprints, fostering a cycle of continuous improvement.

Stakeholder categories are carefully defined to encompass a wide array of actors, including clinical institutions, technology and software providers, European health and research networks, advocacy groups, policy makers, and the general public. The consortium's ten clinical partners serve as a foundation for engaging with healthcare professionals, while research institutions, SMEs, and industrial partners play a crucial role in advancing digital and AI-based healthcare solutions. GenoMed4All also actively collaborates with major EU initiatives and networks, such as ERN-EuroBloodNet, GA4GH, ICPerMed, and the Beyond 1M Genomes project, to align with broader goals in genomics and personalised medicine. Additionally, engagement with related H2020 projects and European platforms in AI, high-performance computing, and open science ensures alignment with leading-edge technological and regulatory developments.

Recognising the strategic value of digital innovation hubs and the influence of policymakers, GenoMed4All incorporates these groups into its engagement model, ensuring broad impact and policy alignment. Moreover, the project places a strong emphasis on societal communication, aiming to foster public trust in AI-driven healthcare and promote the benefits of personalized medicine across Europe.

The stakeholder map visually represents the initial segmentation of these target groups based on their mutual influence and engagement potential, offering a practical guide to prioritise collaboration and outreach efforts. This evolving map and framework underpin GenoMed4All's commitment to a sustainable, impactful, and inclusive approach to stakeholder engagement throughout the project's duration.

**3. Communication and Dissemination**

The GenoMed4All Horizon 2020 project places a strong emphasis on strategic communication and dissemination, recognizing these as central elements for maximizing the project's visibility, engagement, and impact. While these two functions are closely interrelated, they are addressed through distinct yet complementary approaches to ensure tailored outreach to diverse stakeholders.

The communication strategy adopts an agile, phased approach designed to generate awareness and foster interest among both technical and non-specialist audiences. Beginning with foundational planning and tool development, the plan evolves through a broad awareness campaign and culminates in targeted communications aligned with the project's maturity and results. A wide array of digital and print tools, ranging from the official website and social media channels to newsletters, press releases, and multimedia content, will support this effort. Special focus is given to creating clear, accessible messaging supported by visual branding to engage key audiences such as healthcare professionals, digital service providers, policy makers, and the general public.

Internally, effective communication within the consortium is maintained through weekly check-ins and structured updates, ensuring alignment and transparency throughout the project lifecycle.

In parallel, GenoMed4All's dissemination plan is focused on making project results broadly accessible and actionable. It is structured into three successive phases: identifying and studying the dissemination landscape, conducting outreach to key influencers, and fostering adoption of project outcomes. Dissemination tools include open access repositories (e.g., Zenodo), peer-reviewed publications, technical papers, stakeholder consultations, and participation in key conferences and exhibitions. Target audiences span the scientific community, digital innovation sectors, advocacy networks, EU policymakers, and the broader healthcare ecosystem.

Both communication and dissemination efforts are designed not only to inform but also to actively involve stakeholders, ultimately enhancing the uptake and long-term sustainability of GenoMed4All's innovations in genomics-driven personalized medicine.

**4. Exploitation Plan**

The GenoMed4All project outlines a comprehensive and strategic approach to the exploitation of its results, in line with Article 28.1 of the Grant Agreement, which mandates efforts to ensure the use of results for at least four years following the project's conclusion. Exploitation in this context refers to the utilization of the project's outcomes, both tangible and intangible, in research, product or service development, standardization, and broader societal impact. The primary objective is to turn scientific and technological advancements into practical applications with economic, clinical, and societal value.

From the outset, GenoMed4All has aimed to maximize its impact across the EU by enabling cost-effective, interoperable AI-based healthcare solutions, initially targeting haematological diseases. The project's exploitation plan is designed to support clinical application, innovation, and entrepreneurship, and is structured around two key development sprints. The first sprint culminates in an interim impact, exploitation, and sustainability report (D9.3), focusing on stakeholder identification, market analysis, and value proposition development. It maps the project's ecosystem, evaluates market readiness, and defines how KERs align with specific audience needs.

Building on Sprint #1, the second sprint, detailed in report D9.5, advances toward commercialization and innovation activation. This phase includes the development of business models for each commercially viable KER and an assessment of intellectual property and knowledge management strategies to ensure long-term sustainability and legal compliance.

GenoMed4All's exploitation strategy spans several domains including financial commercialization, research and development, education, community building, knowledge transfer, open-source contributions, and the ethical use of AI in healthcare. In addition to the consortium-wide strategy, individual partners are encouraged to pursue their own exploitation initiatives.

The core exploitable results are grouped into two primary categories: a robust technological platform and data infrastructure, and AI-based services for clinical support. These include data curation modules, secure FL systems, compliance tools, and clinical algorithms for diagnosis, prognosis, and treatment planning.

The commercialization strategy is phased to accommodate the evolving nature of both the project and the markets it addresses. Step one focuses on assessing market readiness through detailed research and stakeholder engagement to understand competitive landscapes, existing solutions, and sustainable models. Step two involves crafting a complete Business Model Canvas for each KER, supported by targeted workshops and thematic leadership from key project partners.

The consortium has identified three main market domains for GenoMed4All's outputs: genomics and personalized medicine, rare disease care, and FL technologies. Each of these domains is characterized by active innovation, substantial investment, and a rapidly evolving landscape. By monitoring developments in these areas and aligning GenoMed4All's outputs with current needs and standards, the project aims to position itself strategically within these critical sectors, paving the way for successful exploitation and long-term impact.

GenoMed4All has established a robust Intellectual Property Rights (IPR) management strategy aimed at safeguarding and leveraging the knowledge generated throughout the project. The Project Management Office (PMO) oversees this framework, supporting the Project Officer in monitoring knowledge management across the consortium. The project promotes individual and joint innovation, governed by the principles set in the Consortium Agreement. Sole contributors retain full rights to their discoveries, while jointly developed knowledge is managed through Joint Ownership Agreements. Access rights are structured to promote openness under fair and reasonable terms. The European Commission is granted royalty-free access to outcomes for public dissemination. GenoMed4All's IPR strategy encompasses IP protection, a technology transfer system involving professional knowledge transfer bodies, and legal mechanisms to resolve disputes and enforce rights, all documented in D9.5.

In parallel, GenoMed4All is contributing to European and global standardization efforts, particularly in genomics and clinical data. WPs 8 and 5 lead initiatives to implement standardized frameworks that support machine learning and data integration from bench to bedside. While the full scope of impacted standardization bodies is still evolving, GenoMed4All actively engages with entities such as GA4GH and European Reference Networks to influence emerging norms. The project is aligning with key data formats and protocols across bioinformatics, clinical use cases, and imaging data, and aims to make its frameworks widely accessible. D8.3 will summarize the impact and scope of these efforts.

To ensure the relevance and commercial viability of its outputs, GenoMed4All has adopted a segmented value proposition strategy. Through targeted workshops involving stakeholders from industry, SMEs, clinical sites, research institutions, and policy makers, the project gathers insights to fine-tune its value offering. These discussions will inform commercialization pathways and are critical to the development of the business model outlined in D9.5.

Each stakeholder group within the consortium has distinct exploitation plans. Industry partners aim to enhance their technological platforms and market offerings with GenoMed4All's AI services and FL infrastructure. SMEs seek to use the project's innovations to diversify their portfolios and enter new markets. Clinical sites plan to leverage the project's outputs to improve diagnostic services, promote patient-centric care, and accelerate research into hematological diseases. Research institutions will utilize the project's data, algorithms, and infrastructure to advance AI applications and foster continued innovation in rare disease research.

Overall, GenoMed4All's comprehensive approach to IPR, standardization, value proposition, and exploitation reflects its commitment to maximizing the impact of its work across the biomedical and AI ecosystems in Europe and beyond.

**5. Monitoring of Communication, Dissemination and Exploitation efforts**

Continuous monitoring and customization of the Communication, Dissemination, and Exploitation Plan are critical to the success of GenoMed4All. This ongoing oversight enables the consortium to identify and address potential deviations from project timelines or resource allocations early, ensuring timely corrective actions to meet objectives. The consortium prioritizes pre-assessment of information needs, appropriate monitoring frequency, and a structured methodology for evidence collection. Effective execution relies on responsive and adaptable mechanisms, supported by confidential impact KPIs shared with the European Commission and internally within the consortium. To facilitate transparency and coordination, internal reporting tools have been established, enabling partners to track and report their communication, dissemination, and exploitation efforts, including activities such as publications, standardization contributions, and platform demonstrations.

To support these efforts, GenoMed4All maintains a communication and dissemination database compiling available resources across partner networks, a collective tracker for documenting related activities to ensure traceability and compliance, and an editorial calendar to schedule and coordinate content publication across various channels. These tools foster a coordinated approach to sharing information both within the consortium and externally.

Externally, GenoMed4All leverages its official website and social media platforms like LinkedIn and Twitter to maintain an open and continuous dialogue with the broader European community. These channels facilitate the dissemination of exploitation-related information in an accessible and timely manner, enhancing visibility and engagement with the project's ongoing activities and achievements.

**6. Conclusions**

This document presents GenoMed4All's framework for stakeholder collaboration, communication, dissemination, and exploitation strategies, all managed under WP9, with WP8 leading standardization efforts. Throughout the project's four-year duration, a clearly defined approach is essential for engaging target audiences, promoting knowledge adoption, and scheduling WP9 activities. The document outlines the roles, objectives, and timelines for outreach strategies, emphasizing a thorough assessment of the stakeholder ecosystem to tailor value propositions for each group.

The communication and dissemination plan is designed to facilitate efficient internal coordination across WPs and ensure effective external engagement through platforms such as the project's website and social media channels. The exploitation strategy focuses on three key areas: business, standardization, and intellectual property rights, aiming to commercialize GenoMed4All's knowledge, establish sustainable business models, contribute to European genomics standardization efforts, and provide a robust legal framework to protect innovative knowledge.

As an initial roadmap, this document guides consortium members in executing their communication, dissemination, and exploitation tasks. Given its early release in the project lifecycle, certain strategy details may evolve, with updates to be incorporated in forthcoming deliverables to reflect ongoing progress and refinements.

### Supplementary Information 39| Deliverable 9.2 - Interim Communication & Dissemination report

**1. Introduction**

The long-term success of disruptive innovation depends heavily on the level of awareness it generates and the engagement it fosters. A well-designed dissemination and communication strategy is crucial to ensuring that a project's results receive broad attention and have lasting impact beyond its initial duration. Such a strategy should focus on actively involving key stakeholders to build a committed community that can leverage networking opportunities for future growth.

This deliverable, D9.2 – Communication & Dissemination Interim Report, falls under GenoMed4All's WP9 – Impact, Outreach and Collaboration, specifically within task T9.1 focused on dissemination and communication plans. It provides a comprehensive overview of communication and dissemination activities carried out during the project's first 18 months, including an analysis of digital channel performance, event participation, and publication outputs. Building on the initial strategy outlined in D9.1 – GenoMed4All Impact Master Plan, this report documents the progress of outreach and engagement efforts during the first review period and will be further expanded and updated in the final report, D9.4.

**2. Dissemination and Communication Strategy**

GenoMed4All's outreach strategy aims to increase overall awareness and interest in the project, thereby establishing a robust customer base and ecosystem for future dissemination and exploitation. It focuses on communicating technical and scientific results to specialized stakeholders while delivering accessible messages to non-technical audiences, ultimately broadening the project's reach to the widest possible community. The communication activities have been structured into three phases: Plan & Revisit, Early Bird Communication, and Targeted Communication, designed to build and grow a community around the project, maximize visibility, and engage end users for ongoing dialogue and feedback.

During the first six months, foundational work was completed, including creating the project's visual identity, establishing communication channels, and mapping stakeholders. The following phase focused on strengthening digital presence, initiating conversations with related initiatives, and producing promotional content to explain core concepts. In the most recent phase, efforts have intensified on increasing event participation, publications, and refining the value proposition and engagement strategy to onboard new clinical sites and research institutions. The project is now in the Targeted Communication phase, aiming to reinforce key messages and link them to upcoming results, such as the first iteration of its business plan.

Over the initial 18 months, GenoMed4All has consolidated its online presence, amassing approximately 1,300 social media followers, producing over 430 social media posts, 24 blog entries, and 12 publications on Zenodo. The project has increased participation in events, hosting workshops, webinars, and featuring at key conferences, supported by a consistent branding identity and comprehensive promotional materials in both print and digital formats. These efforts have contributed to steady progress against defined impact KPIs, including scientific publications, event participation, social media engagement, and outreach outputs, demonstrating significant momentum in communication and dissemination activities as the project advances.

**3. Communication and Dissemination Activities Implemented**

GenoMed4All established a comprehensive visual identity early in the project, including a distinctive logo, brand assets, typography, color schemes, and official templates to ensure consistency across all communications and documentation. The project's digital communication toolkit comprises a dedicated website, active social media accounts on Twitter and LinkedIn, an Open Access repository on Zenodo, and an internal newsletter managed via Sendinblue. The website serves as the primary platform to present GenoMed4All's core concepts, use cases, and updates, attracting a steady flow of visitors and returning users. Social media channels have shown significant growth and engagement, driven by a mix of broad awareness campaigns and targeted communications aligned with project milestones.

Promotional materials developed include slide decks, brochures, stickers, a factsheet, and multimedia content hosted on YouTube, supporting outreach and event participation. The consortium has maintained a steady publication pipeline, producing peer-reviewed scientific articles, clinical studies, posters, and abstracts, alongside non-scientific content such as blog articles and press releases, enhancing both scientific and public visibility. Despite pandemic-related challenges, GenoMed4All actively participated in numerous high-profile scientific events, webinars, and workshops, both virtual and in-person, further extending its outreach and fostering engagement within the hematology and rare disease communities. This integrated approach to visual identity, digital presence, publications, and event participation has strengthened the project's communication impact and positioned GenoMed4All as a recognized initiative in precision medicine.

##### 4. Engagement Strategy

During the first 18 months, GenoMed4All has primarily focused on developing its engagement and onboarding strategy, supported by targeted initiatives such as the Patient Engagement e-workshop and the implementation of online surveys designed to evaluate data needs and identify potential clinical sites. The Patient Engagement e-workshop was a key component, bringing together representatives from major European patient associations related to MDS, MM and SCD, to present GenoMed4All's mission, clinical pilots, and AI use cases, fostering ongoing dialogue around patient-centric development of clinical AI tools. Concurrently, a series of surveys were conducted to assess data availability, regulatory compliance, and ethical considerations, facilitating the onboarding of external organizations through a structured Associate Member form that collects detailed clinical and genomic data while offering academic recognition and access to GenoMed4All's tools. Networking has played a significant role, especially through collaboration with the ERN-EuroBloodNet community, with efforts underway to onboard new clinical sites. Partnerships have also been established with the Beyond 1M Genomes (B1MG) initiative, contributing to the development of standards for secure, cross-border genomic data sharing while addressing ethical, legal, and social implications. GenoMed4All has actively engaged with sister projects under the AI for Genomics and Personalized Medicine umbrella, fostering dialogue on data sharing and FL infrastructure. Additionally, exploratory discussions with the HARMONY Alliance have identified opportunities to integrate extensive hematological cancer datasets into GenoMed4All's platform and collaborate on workshops, strengthening the project's capacity to develop federated AI models for clinical applications.

##### 5. Conclusions

This deliverable provides a periodic update on GenoMed4All's Dissemination and Communication strategy, reflecting the project's progress over its first 18 months. During this time, key milestones were achieved, including the development of a cohesive visual identity and branding assets, the establishment and consolidation of main digital channels, and successful initial publication and promotional activities. These efforts have resulted in a strong, recognizable brand and a growing, engaged community focused on AI's impact in hematology and healthcare more broadly. The communication strategy remains dynamic and collaborative, evolving alongside the project to ensure all outreach materials and channels accurately represent GenoMed4All's ongoing developments and achievements. As the project matures, there will be an increased focus on targeted scientific dissemination, with intensified publishing and networking activities aimed at maximizing reach and visibility. These initiatives will also support the development of a tailored business plan, value proposition, and service portfolio, which will be detailed further in the upcoming D9.3 – Interim Impact Assessment, Exploitation and Sustainability Report.

### Supplementary Information 40| Deliverable 9.3 - Interim GenoMed4All Impact Assessment, Exploitation and Sustainability Report

##### 1. Introduction

This report provides the consortium with an initial overview of the planned strategies for the exploitation and sustainability of the GenoMed4All project. It addresses not only the platform itself but also various exploitable outcomes from partners' research and innovation efforts, including open research, publications, patents, products, and services. The deliverable begins with an outline of the general methodology for impact assessment, exploitation, and sustainability planning, followed by a preliminary analysis of the health information exchange market. It then details the KERs and proposed exploitation strategies as of month 23, alongside the sustainability strategy for the GenoMed4All platform, incorporating the engagement approach for external partners. The report concludes with a summary and an outlook on planned activities for the upcoming months.

### 2. Impact Assessment, Exploitation and Sustainability Planning Methodology

The impact assessment, exploitation, and sustainability planning for GenoMed4All has been structured through a series of interconnected and overlapping activities. Initially, a market analysis was conducted to explore the health information exchange market and its relevance to the project. This was followed by the preliminary identification of KERs, which were collectively reviewed and presented during the First Periodic Review. Currently, these KERs are being discussed, prioritized, and aligned with evolving project outcomes, while relevant exploitation pathways are being developed. These strategies are continuously refined through exploitation workshops planned for the third and fourth years of the project, supplemented by detailed questionnaires to address the challenges of coordinating large partner groups. In the final year, these strategies will be finalized and implemented. Parallel to this, sustainability planning for the GenoMed4All platform has progressed through the development of an onboarding strategy for external partners, the establishment of a value proposition, a legal framework, and the creation of an end-to-end business infrastructure through coordinated efforts across multiple WPs.

### 3. Market Analysis

The health information exchange (HIE) market, encompassing the lawful and standardized sharing of health-related data among stakeholders for clinical, research, and commercial purposes, is projected to grow substantially, reaching $2 billion by 2025. This growth is driven by factors such as cost containment, patient-centric care, widespread adoption of EHRs, and government initiatives. The COVID-19 pandemic further underscored the need for enhanced data interoperability and remote healthcare solutions. Despite these drivers, challenges remain, including data fragmentation, interoperability barriers, significant infrastructure investments, and privacy concerns heightened by regulations like GDPR, which complicate data sharing especially for rare diseases.

Within the HIE market, health data exchange platforms are the fastest-growing segment. These platforms facilitate value-based interactions between data producers and consumers by providing open, seamless connectivity, efficient data utilization, and user-centric convenience while maintaining robust security and privacy controls. Success depends on engaging diverse stakeholders through tailored value propositions and ensuring trust and data quality.

Research platforms, a subset of HIE platforms, vary from centralized national repositories to participant-driven initiatives, each with distinct governance models and levels of patient involvement in data sharing.

The global AI in healthcare market is rapidly expanding, expected to reach $67.4 billion by 2027 with a CAGR of 46.2%, propelled by the influx of complex healthcare data, growing partnerships across domains, cost pressures, and technological advances. AI adoption is particularly prominent in elderly care, multi-omics data analysis, diagnostics, and drug discovery. Despite this growth, barriers include data privacy challenges, regulatory constraints, and hesitance among medical professionals due to concerns about AI reliability, the potential disruption of physician-patient relationships, and the technology’s soft skill limitations.

ML, a critical subset of AI, supports a broad range of healthcare applications, including disease management, epidemic forecasting, robotic surgery, drug discovery, clinical trial optimization, and personalized treatment, all aimed at enhancing clinical decision-making.

FL emerges as a promising approach to address data privacy and fragmentation issues inherent in healthcare by enabling ML model training across multiple decentralized data sources without sharing sensitive data. FL enhances scalability, data protection, and model generalizability, allowing collaborative AI development that can reduce bias and improve predictive accuracy across diverse clinical settings.

Haematological diseases illustrate a key use case for AI applications, where ML enhances diagnostic accuracy through analysis of cytomorphology, cytogenetics, immunophenotyping, and molecular genetics data. Advanced molecular techniques and high-throughput sequencing combined with ML hold significant promise for personalized diagnosis, prognosis, and patient management, ultimately aiming to improve treatment outcomes and reduce diagnostic errors.

**4. Key exploitable results and envisaged exploitation strategies**

By month 18 (M18), GenoMed4All had identified a preliminary set of KERs aligned with the project's work plan, including a technological platform encompassing a cross-border data infrastructure for dataset management and AI model development, as well as a governance strategy addressing legal and compliance frameworks. The project also developed AI-based clinical support services targeting MDS, SCD, and MM, offering diagnostic, prognostic, and clinical decision support tools.

These KERs were further refined following a technical partners' workshop at month 30 (M30) and a subsequent exploitation questionnaire at month 36 (M36), with ongoing efforts to engage all partners and clinical researchers. By month 38 (M38), updated key innovations included advanced centralized and FL algorithms for medical data, novel validation protocols, a FL platform enabling data privacy, and clinical decision support systems such as one for optimizing HSCT timing in MDS patients. Additional innovations encompassed statistical methods for gene expression analysis, multimodal deep generative models, survival prediction tools outperforming traditional methods, MRI-based lesion segmentation for SCD, scalable validation frameworks, and data homogenization processes integrated with FL.

Most results currently range from technology readiness levels 3 to 6, leading partners to pursue primarily research-based exploitation through peer-reviewed publications and continued R&D, with some advanced technologies considered for patent applications.

Exploitation strategies under consideration include software as a service (SaaS), which offers cloud-based, remotely managed applications providing access to data infrastructure, AI model development, and legal compliance tools. Software licensing options are also being evaluated to protect intellectual property and define user rights, balancing proprietary and open-source approaches tailored to each partner's needs. Peer-reviewed publication remains the primary dissemination channel, especially for clinical AI tools. Patents may be pursued for select high-impact innovations, particularly AI algorithms. Additionally, open research models are contemplated to foster transparency and collaboration, although clinical data privacy requirements may limit full openness. Overall, GenoMed4All is actively adapting its exploitation pathways to maximize impact and ensure responsible use of its technological and clinical advances.

**5. GenoMed4All platform sustainability planning**

The long-term sustainability of GenoMed4All's platform is central to its exploitation and sustainability strategy. The consortium is exploring multiple exploitation pathways, including SaaS/PaaS and open research, and has developed a partners' engagement strategy as a preliminary step to attract and onboard clinical and research collaborators. This strategy is founded on three key pillars: a clear value proposition distinguishing clinical and research operational modes, a comprehensive end-to-end infrastructure integrating data engineering, AI development, and validation, and a robust legal framework that governs data sharing and onboarding processes.

The platform's value proposition supports clinicians with local decision support tools based on FL models and offers researchers an AI sandbox for model training and benchmarking using real-world data, addressing the challenges of clinical AI integration. GenoMed4All distinguishes two operational modes: clinical mode for real-time patient data input and prediction, and research mode for retrospective data analysis and model development, each with defined actors, functionalities, and benefits.

A comprehensive legal framework underpins data sharing both within the consortium and with external parties, including JCAs for internal partners and Associate Member Agreements for third parties, ensuring compliance with ethical and data protection standards. The end-to-end business infrastructure defines onboarding scenarios, operational models, data flows, and a future service portfolio to support platform lifecycle management. Currently, four onboarding scenarios are being considered to engage external research organizations, ranging from introducing new disease use cases to enhancing existing AI models or utilizing the platform's clinical decision support tools.

The operating model and data flows are being aligned with FL validation protocols to facilitate efficient data acquisition, model training, and deployment. Concurrently, work is underway to define a preliminary service portfolio tailored to different engagement options, encompassing core platform capabilities and governance functions. External partners are expected to contribute assets aligned with platform standards, ensuring integration and compliance within the FL ecosystem. This comprehensive approach aims to secure GenoMed4All's long-term impact, fostering collaboration and advancing personalized medicine for hematological diseases.

**6. Conclusions**

As of month 38, the consortium has identified several KERs emerging from the project's technical activities. These include the GenoMed4All technological platform and services as well as AI-based clinical support tools focused on diagnostic and prognostic algorithms. Various exploitation routes are being considered for these KERs, ranging from software as a service (SaaS), software licensing models (both proprietary and open-source), peer-reviewed publications, patents, and open research. Most results currently have technology readiness levels (TRLs) between 3 and 6 and are planned for research-focused exploitation through academic publication and further R&D, with potential patent filings for the most advanced innovations.

Looking ahead to months 39 to 48, exploitation and sustainability efforts will focus on consolidating and refining these results, developing tailored exploitation strategies, and ensuring the long-term sustainability of the GenoMed4All platform. This phase will include prioritizing key results, exploring suitable exploitation approaches, conducting further workshops and consultations with internal and external stakeholders, and finalizing the platform's sustainability plan and exploitation framework.

### Supplementary References

1. Goodfellow I, Pouget-Abadie J, Mirza M, et al. Generative adversarial networks. Commun ACM. 2020;63(11):139–44.

2. Kiran A, Kumar SS. A Methodology and an Empirical Analysis to Determine the Most Suitable Synthetic Data Generator. IEEE Access. 2024;12:12209–28.

3. Patki N, Wedge R, Veeramachaneni K. The Synthetic Data Vault. In: 2016 IEEE International Conference on Data Science and Advanced Analytics (DSAA). Montreal, QC, Canada: IEEE; 2016. p. 399–410.

4. Beutel DJ, Topal T, Mathur A, et al. Flower: A Friendly Federated Learning Research Framework . arXiv; 2022. Available at: http://arxiv.org/abs/2007.14390

5. Apellániz PA, Parras J, Zazo S. An improved tabular data generator with VAE-GMM integration. arXiv; 2024. Available at: http://arxiv.org/abs/2404.08434

6. Kingma DP, Welling M. Auto-Encoding Variational Bayes. arXiv; 2022. DAvailable at: http://arxiv.org/abs/1312.6114

7. McMahan B, Moore E, Ramage D, Hampson S, Arcas BA y. Communication-Efficient Learning of Deep Networks from Decentralized Data. In: Proceedings of the 20th International Conference on Artificial Intelligence and Statistics. PMLR; 2017. p. 1273–82.

8. Apellániz PA, Parras J, Zazo S. Improving Synthetic Data Generation Through Federated Learning in Scarce and Heterogeneous Data Scenarios. Big Data and Cognitive Computing. febbraio 2025;9(2):18.

9. Koller D, Friedman N. Probabilistic graphical models: principles and techniques. Nachdr. Cambridge, Mass.: MIT Press; 2010. 1231 p. (Adaptive computation and machine learning).

10. Kaur D, Sobiesk M, Patil S, et al. Application of Bayesian networks to generate synthetic health data. J Am Med Inform Assoc. 2020;28(4):801–11.

11. Zhang J, Cormode G, Procopiuc CM, Srivastava D, Xiao X. PrivBayes: Private Data Release via Bayesian Networks. ACM Trans Database Syst. 2017;42(4):25:1-25:41.

12. Hittmeir M, Mayer R, Ekelhart A. Efficient Bayesian Network Construction for Increased Privacy on Synthetic Data. In: 2022 IEEE International Conference on Big Data (Big Data). 2022. p. 5721–30.

13. Kissner L, Song D. Privacy-Preserving Set Operations. In: Shoup V, curatore. Advances in Cryptology – CRYPTO 2005. Berlin, Heidelberg: Springer; 2005. p. 241–57.

14. Zhang Y, Chen Q, Zhong S. Efficient and Privacy-Preserving Min and k th Min Computations in Mobile Sensing Systems. IEEE Transactions on Dependable and Secure Computing. 2017;14(1):9–21.

15. De Falco I, Della Cioppa A, Koutny T, et al. A Federated Learning-Inspired Evolutionary Algorithm: Application to Glucose Prediction. Sensors. 2023;23(6):2957.

16. Rastogi V, Nath S. Differentially private aggregation of distributed time-series with transformation and encryption. In: Proceedings of the 2010 ACM SIGMOD International Conference on Management of data. New York, NY, USA: Association for Computing Machinery; 2010 . p. 735–46. (SIGMOD '10).

17. D'Amico S, Dall'Olio D, Sala C, et al. Synthetic Data Generation by Artificial Intelligence to Accelerate Research and Precision Medicine in Hematology. JCO Clin Cancer Inform. 2023;7:e2300021.

18. Cox DR. Regression Models and Life-Tables. Journal of the Royal Statistical Society Series B (Methodological). 1972;34(2):187–220.

19. Harrell FE Jr, Califf RM, Pryor DB, Lee KL, Rosati RA. Evaluating the Yield of Medical Tests. JAMA. 1982;247(18):2543–6.

20. Bersanelli M, Travaglino E, Meggendorfer M, et al. Classification and Personalized Prognostic Assessment on the Basis of Clinical and Genomic Features in Myelodysplastic Syndromes. JCO. 2021;39(11):1223–33.
